# Survey: Accessibility of At-Home COVID-19 Tests for Users with Disabilities

**DOI:** 10.1101/2024.08.13.24311938

**Authors:** Sarah Farmer, Amanda Peagler, Rebecca Sheiner, Adina Martinez, Victoria Razin

## Abstract

Widespread access to over-the-counter at-home COVID-19 tests was a crucial component of the United States’ public health response to the COVID-19 pandemic. For at-home testing to be effective, it is essential for the tests to be not only available but also usable and accessible to people with disabilities. In the present study, a survey was conducted to collect feedback about people’s experiences using over-the-counter COVID-19 tests, with an aim to identify barriers to the accessibility of COVID-19 tests for people with disabilities. Survey respondents self-identified as having little to no vision, low vision, or limited dexterity. The survey covered ten at-home COVID-19 test brands. For each test that a respondent had used, the survey included questions about whether the user was able to complete the test independently or whether they needed assistance, the ease of use for performing each part of the testing procedure, and any difficulties that the user encountered while using the test. The survey also included questions for each test regarding the use of mobile applications, digital instructions, and customer service. This report presents the results of nearly 400 responses to the accessibility survey. The results offer an improved understanding of accessibility barriers for at-home COVID-19 tests; designers of at-home diagnostic tests can apply this knowledge in order to improve the accessibility of over-the-counter COVID-19 tests and other at-home diagnostic tests to people with disabilities.

## 3 Introduction

The ability to test for contagious viruses at home is crucial for public health, though home users must be able to correctly complete and understand the tests for them to be effective. Home diagnostics requires a deep understanding of users’ perspectives and abilities. Companies often overlook challenges faced by people with disabilities, such as low or no vision and dexterity limitations. Failing to design with these limitations in mind can exclude a large portion of the population from using these tests.

To control the COVID-19 pandemic, the FDA issued Emergency Use Authorizations that allowed many COVID-19 tests to enter the market, allowing the public to test for COVID-19 at home. Access to at-home COVID-19 tests to American households increased significantly with the free delivery of tests via the US Postal Service. The increase in test availability also increases the amount of user experiences to be studied.

The purpose of this survey was to collect feedback about people’s experiences with over-the-counter COVID-19 at-home test kits. This feedback was collected to identify barriers to the accessibility of COVID-19 tests for people with disabilities.

## 4 Methods

Ten at-home COVID-19 test brands were identified and selected based on availability and popularity. All ten COVID-19 tests included in the accessibility survey were lateral flow assays (LFAs). One test included a reader that presented results via a smartphone application and the others were read visually, all with varying protocol complexity. The tests selected for the survey are listed below. If participants had used a test not listed or could not remember the test used, “Other” and “I don’t know” responses were available.

- BinaxNow, Abbott
- iHealth, iHealth Labs, Inc.
- QuickVue, QuidelOrtho Corporation
- CareStart, Access Bio, Inc.
- Flowflex, ACON Laboratories, Inc.
- Roche, Roche Diagnostics
- InteliSwab, OraSure Technologies, Inc.
- On/Go, Access Bio, Inc.
- Ellume, Ellume Limited
- Siemens Clinitest, Siemens Healthineers

Representation across various disability groups was prioritized. In particular, the following groups were targeted: blind users, users with low vision, users with limited dexterity, and older adult users.

Participants were recruited through Georgia Tech’s HomeLab, advocacy organizations, email, and word of mouth. The survey was conducted online via the Qualtrics platform from August 16, 2022 to November 17, 2022, and received 840 total responses.

Participants provided their informed consent prior to beginning the survey. Participation for this study was voluntary and participants had the right to withdraw at any time without reason and without penalty. The survey took approximately 30 minutes to complete. This study was approved by the Georgia Tech Institutional Review Board (IRB). Before distribution, the survey was checked to ensure it was accessible to people with vision-related disabilities. To ensure high-quality data, duplicate and incomplete responses were removed. This resulted in a total of 394 responses included in the study.

### 4.1 Limitations

Limited demographic data was collected; participants were asked to provide only their age and which disability type (if any) they identified with. Categorization into disability types was self-identified and many participants identified with multiple disability types. Because this survey was distributed and taken online only, individuals who are less proficient with smartphones and computers were likely under-represented; this is an important user group to study in the future as users with fewer technological skills are less likely to benefit from accessibility aids such as mobile apps, reader technologies, and digital or video instructions.

Due to a conservative approach to data cleaning, legitimate responses may have been removed from the dataset.

### 4.2 Survey

The survey consisted of three parts: general questions, test-specific questions, and closing questions. The survey began with a series of general questions related to the participant’s demographics, self-identification with a disability, and comfort with technology. Participants also selected which at-home COVID-19 test brands they had used: BinaxNow, iHealth, QuickVue, CareStart, Flowflex, Roche, InteliSwab, On/Go, Ellume, Siemens Clinitest (or “Other” or “I don’t know”). The general questions and answer choices are given in Table 1.

**Table 1.** Overall Questions.

| Question | Answer Choices |
| --- | --- |
| **What is your age group?** | • 18-24<br>• 25-34<br>• 35-44<br>• 45-54<br>• 55-64<br>• 65-74<br>• 75-84<br>• 85+ |
| **Do you identify with any of the following? If so, indicate all that apply:** | • I have very little to no vision<br>• I have low vision<br>• I have limited hand dexterity or limited fine motor control<br>• None of the above |
| **Do you have a smartphone or tablet?** | • Yes<br>• No |
| **How would you rate your smartphone, tablet, and computer skills?** | • Extremely proficient<br>• Somewhat proficient<br>• Average<br>• Somewhat limited<br>• Extremely limited |
| **Are you associated with any of the following groups?** | • National Federation of the Blind<br>• American Council of the Blind<br>• Administration for Community Living<br>• Protection and Advocacy System<br>• Client Assistance Program<br>• World Institute on Disability (WID)<br>• Other: _____<br>• None of the above |
| **Have you taken any COVID-19 home tests in the last year?** | • Yes<br>• No |
| **Which test(s) have you used?** | • BinaxNow<br>• iHealth<br>• QuickVue |
|  | • CareStart<br>• Flowflex<br>• Roche<br>• InteliSwab<br>• On/Go<br>• Ellume<br>• Siemens Clinitest<br>• Other<br>• I don't know |

For each test that participants indicated they had used, they were prompted with a series of questions regarding the accessibility of that test. If participants selected ‘I don’t know’ in response to which test(s) they had used, they were prompted with the same questions about their test with the test name removed from the questions. The test-specific questions and answer choices are given in Table 2.

**Table 2:** Test-Specific Accessibility Questions.

| Question | Answer Choices |
| --- | --- |
| **Think of the first time you used the [test name] test. Did you have difficulty completing any of the following tasks? Select all that apply:** | • Opening the packaging<br>• Reading the instructions<br>• Identifying test components<br>• Collecting the test sample<br>• Manipulating test components<br>• Interpreting the test results<br>• Other<br>• I did not have difficulty completing the test |
| **What tools or assistance, if any, did you use to perform the test?** | • Timer<br>• Scissors<br>• Remote visual assistance (AIRA, Be My Eyes, Facetime, Video-chat, etc.)<br>• Screen Reader<br>• Magnification<br>• Other technology (OCR, Bar Code Readers, Light Detectors, etc.)<br>• In person human assistance<br>• Other<br>• I did not need any tools or assistance |
| **Were you able to conduct this test on your first try?** | • Yes<br>• No |
| **How long did it take to complete the steps required to perform** | • Under 5 minutes<br>• 5 - 10 minutes |
| **the test, not including the time you spent waiting for results.** | • 10 - 15 minutes<br>• Over 15 minutes |
| **What format of instructions did you use?** | • Paper<br>• Digital (webpages, downloaded, etc.)<br>• Associated Covid Test App (Smartphone)<br>• None of the above |
| **Were you able to access electronic information via a QR Code?** | • Yes<br>• No |
| **How easy was opening the package of the *[test name]* test for you? (without assistance)** | • Very difficult<br>• Difficult<br>• Neither easy nor difficult<br>• Easy<br>• Very easy |
| **How easy was completing the test protocol for you? (without assistance)** | • Very difficult<br>• Difficult<br>• Neither easy nor difficult<br>• Easy<br>• Very easy |
| **How easy was interpreting the test results for you? (without assistance)** | • Very difficult<br>• Difficult<br>• Neither easy nor difficult<br>• Easy<br>• Very easy |
| **Were you able to interpret the test results? (without assistance)** | • Yes<br>• No |
| **If there was an associated app for the test, how easy was it for you to use? (without assistance)** | • Very difficult<br>• Difficult<br>• Neither easy nor difficult<br>• Easy<br>• Very easy<br>• There was not an app<br>• There is an app, but I didn't use it |
| **How helpful was the app in assisting you with completing the test?** | • Very unhelpful<br>• Unhelpful<br>• Neither unhelpful nor helpful<br>• Helpful<br>• Very helpful |
| **Did you receive a valid test result with the [test name] test (positive or negative)?** | • Yes<br>• No |
| **Did you attempt to contact [test name] 's customer support?** | • Yes<br>• No |
| **Was customer support helpful in answering your question(s)?** | • Very unhelpful<br>• Unhelpful<br>• Neither helpful nor unhelpful<br>• Helpful<br>• Very helpful |
| **If you have performed the [test name] test multiple times, how did your experience change?** | • Much worse<br>• Worse<br>• Neither better nor worse<br>• Better<br>• Much better<br>• N/A |
| **What did you like about the [test name] test?** | *Free response* |
| **What would you like to see improved for the [test name] test?** | *Free response* |

After completing the test-specific questions for each of the tests selected from the test list, participants were asked two closing questions. The closing questions and answer choices are given in Table 3.

**Table 3:**
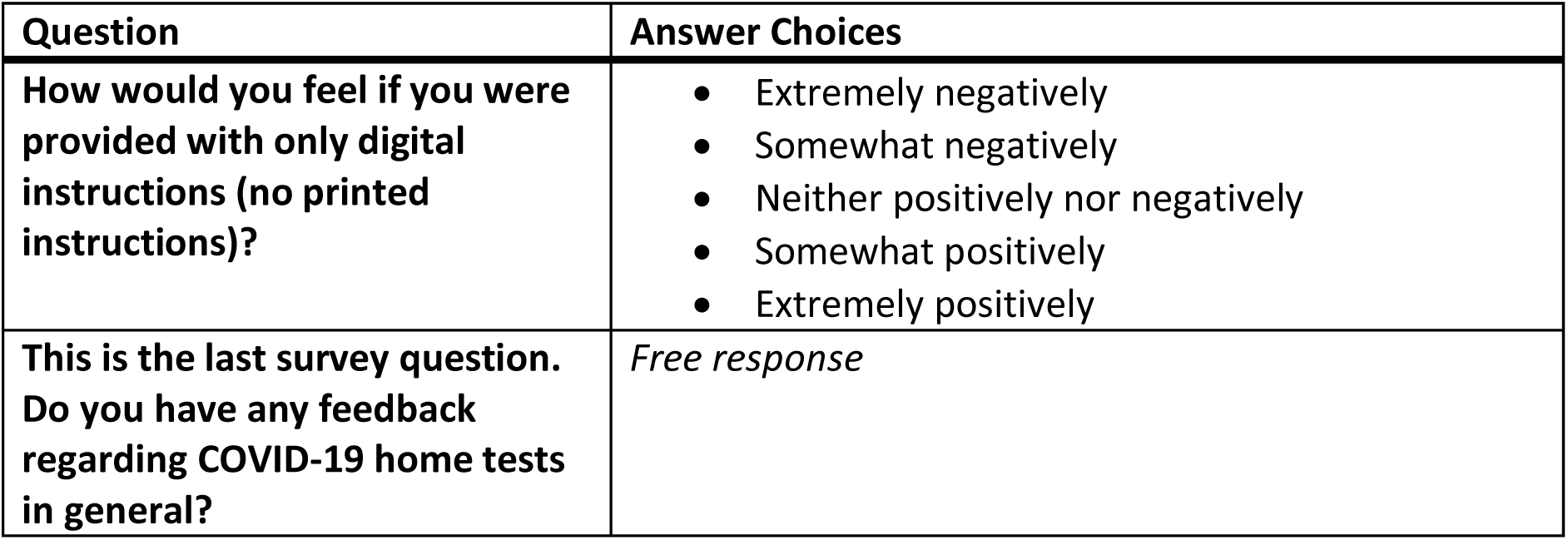
Closing Questions.

## 5 Results

Results are provided in table format below.

### 5.1 General Questions

Table 4 presents the number and percentage of participants in each age group provided as a survey response option.

**Table 4.**
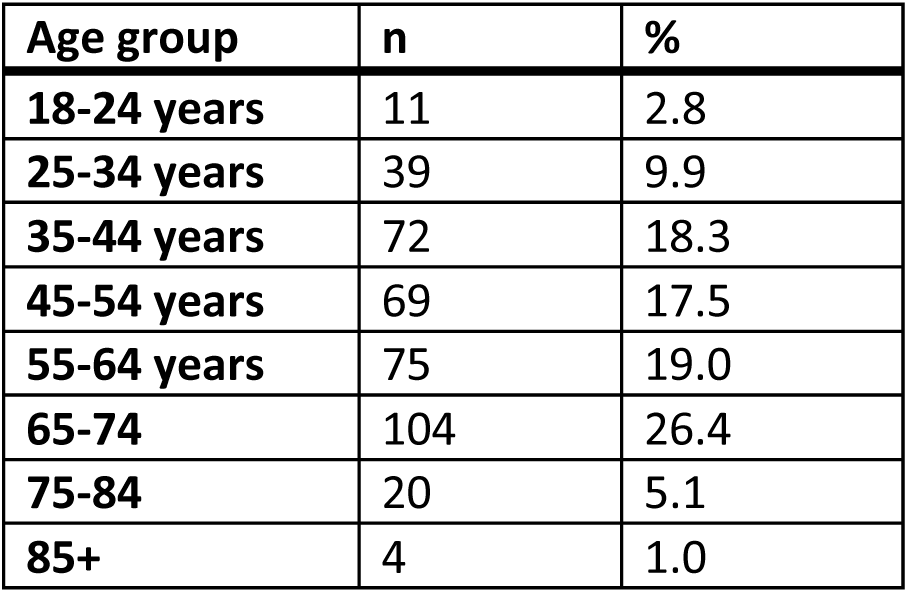
Age groups.

| Age group | n | % |
| --- | --- | --- |
| 18-24 years | 11 | 2.8 |
| 25-34 years | 39 | 9.9 |
| 35-44 years | 72 | 18.3 |
| 45-54 years | 69 | 17.5 |
| 55-64 years | 75 | 19.0 |
| 65-74 | 104 | 26.4 |
| 75-84 | 20 | 5.1 |
| 85+ | 4 | 1.0 |

Table 5 presents the number of participants who self-identified as belonging to one of the disability categories provided as a response option; participants were able to select multiple responses.

**Table 5.** Self-identified multiple selection disability type.

| Disability type | n |
| --- | --- |
| Very little to no vision | 207 |
| Low vision | 109 |
| Limited hand dexterity or limited fine motor control | 28 |

Table 6 presents the self-reported smartphone and computer skills proficiency ratings of participants. Seventeen responses (4.3%) were missing for this question.

**Table 6.** Self-reported smartphone and computer skills proficiency rating.

| Skills Rating | n | % |
| --- | --- | --- |
| Extremely proficient | 194 | 49.2 |
| Somewhat proficient | 104 | 26.4 |
| **Average** | 64 | 16.2 |
| **Somewhat limited** | 13 | 3.3 |
| **Extremely limited** | 2 | 0.5 |

### 5.2 At-home test groups

All ten COVID-19 tests included in the accessibility survey were lateral flow assays (LFAs). These tests were divided into groups depending on whether the results were read visually or with an app reader. The visually read LFAs split into two groups: those with fluid transfer required via a tube with a dropper tip (Group 1), and those without fluid transfer (Group 2). Group 3 consisted of an LFA that used an app reader. The groups are outlined in Table 7.

**Table 7:** Three groups of tests.

| **Group number** | **Description** | **Test in Group** | **n** |
| --- | --- | --- | --- |
| **Group 1** | Read visually, Fluid transfer required | BinaxNow, iHealth, CareStart, Flowflex, Roche, On/Go, Siemens Clinitest | 207 |
| **Group 2** | Read visually, No fluid transfer required | QuickVue, IntelliSwab | 31 |
| **Group 3** | App reader | Ellume | 103 |

The survey responses were aggregated for all tests within a group. Survey responses for each group of tests are given below.

### 5.3 Difficulty Completing Tasks

Table 8 provides the number and percent of users of each group of tests who had difficulty completing the tasks comprising the testing process.

**Table 8.** Responses to question: “Think of the first time you used the [test name] test. Did you have difficulty completing any of the following tasks? Select all that apply.”

| **Difficulty Completing Tasks** | **Group 1:**<br>**LFA +**<br>**fluid**<br>**transfer**<br>**(n)** | **Group 1:**<br>**LFA +**<br>**fluid**<br>**transfer**<br>**(%)** | **Group 2:**<br>**LFA**<br>**without**<br>**fluid**<br>**transfer**<br>**(n)** | **Group 2:**<br>**LFA**<br>**without**<br>**fluid**<br>**transfer**<br>**(%)** | **Group 3:**<br>**LFA +**<br>**reader**<br>**(n)** | **Group 3:**<br>**LFA +**<br>**reader**<br>**(%)** |
| --- | --- | --- | --- | --- | --- | --- |
| **Opening the packaging** | 25 | 12.08 | 3 | 9.68 | 16 | 15.53 |
| **Reading the instructions** | 95 | 45.89 | 14 | 45.16 | 44 | 42.72 |
| **Identifying test components** | 66 | 31.88 | 9 | 29.03 | 53 | 51.46 |
| **Collecting the test sample** | 31 | 14.98 | 3 | 9.68 | 24 | 23.30 |
| **Manipulating test components** | 87 | 42.03 | 12 | 38.71 | 50 | 48.54 |
| **Interpreting the test results** | 82 | 39.61 | 13 | 41.94 | 20 | 19.42 |
| **Other** | 11 | 5.31 | 1 | 3.23 | 22 | 21.36 |
| **I did not have difficulty completing the test** | 58 | 28.02 | 10 | 32.26 | 17 | 16.50 |

### 5.4 Tools or Assistance to Perform Test

Table 9 provides the number and percent of users of each group of tests who used tools or assistance to perform the test.

**Table 9.** Responses to question: “What tools or assistance, if any, did you use to perform the test?”

| **Used Tools or Assistance to Perform Test** | **Group 1:**<br>**LFA +**<br>**fluid**<br>**transfer**<br>**(n)** | **Group 1:**<br>**LFA +**<br>**fluid**<br>**transfer**<br>**(%)** | **Group 2:**<br>**LFA**<br>**without**<br>**fluid**<br>**transfer**<br>**(n)** | **Group 2:**<br>**LFA**<br>**without**<br>**fluid**<br>**transfer**<br>**(%)** | **Group 3:**<br>**LFA +**<br>**reader**<br>**(n)** | **Group 3:**<br>**LFA +**<br>**reader**<br>**(%)** |
| --- | --- | --- | --- | --- | --- | --- |
| **Timer** | 114 | 55.07 | 14 | 45.16 | 28 | 27.18 |
| **Scissors** | 59 | 28.50 | 7 | 22.58 | 29 | 28.16 |
| **Remote visual assistance (AIRA, Be My Eyes, Facetime, Video-chat, etc.)** | 43 | 20.77 | 6 | 19.35 | 20 | 19.42 |
| **Screen Reader** | 14 | 6.76 | 2 | 6.45 | 62 | 60.19 |
| **Magnification** | 49 | 23.67 | 5 | 16.13 | 12 | 11.65 |
| **Other technology (OCR, Bar Code Readers, Light Detectors, etc.)** | 14 | 6.76 | 2 | 6.45 | 6 | 5.83 |
| **In person human assistance** | 64 | 30.92 | 10 | 32.26 | 30 | 29.13 |
| **Other** | 8 | 3.86 | 1 | 3.23 | 10 | 9.71 |
| **I did not need any tools or assistance** | 22 | 10.63 | 2 | 6.45 | 7 | 6.80 |

### 5.5 Conducting the Test on the First Try

Table 10 provides the number and percent of users of each group of tests who were or were not able to conduct the test on their first try.

**Table 10.** Responses to question: “Were you able to conduct this test on your first try?”

| **Conducted the Test on the First Try** | **Group 1:**<br>**LFA +**<br>**fluid**<br>**transfer**<br>**(n)** | **Group 1:**<br>**LFA +**<br>**fluid**<br>**transfer**<br>**(%)** | **Group 2:**<br>**LFA**<br>**without**<br>**fluid**<br>**transfer**<br>**(n)** | **Group 2:**<br>**LFA**<br>**without**<br>**fluid**<br>**transfer**<br>**(%)** | **Group 3:**<br>**LFA +**<br>**reader**<br>**(n)** | **Group 3:**<br>**LFA +**<br>**reader**<br>**(%)** |
| --- | --- | --- | --- | --- | --- | --- |
| **Yes** | 158 | 76.33 | 24 | 77.42 | 68 | 66.02 |
| **No** | 42 | 20.29 | 6 | 19.35 | 34 | 33.01 |

### 5.6 Length of Time to Perform Test

Table 11 indicates the length of time users spent performing the test, not including the time spent waiting for results.

**Table 11.**
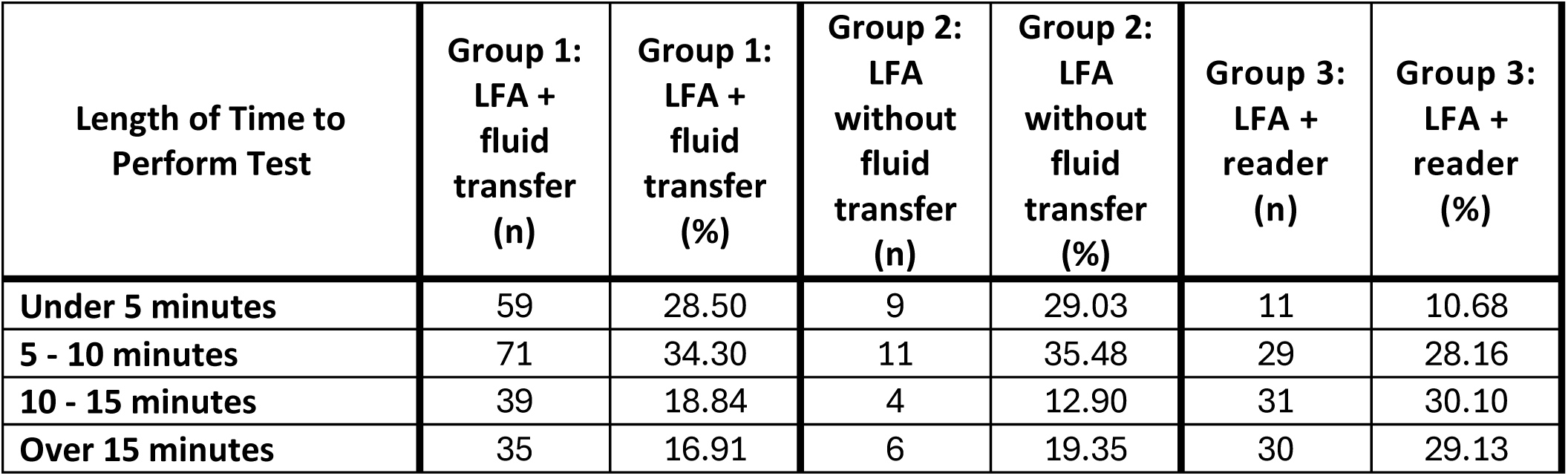
Responses to question: “How long did it take to complete the steps required to perform the test, not including the time you spent waiting for results?”

### 5.7 Instruction Format and Access to Electronic Information

Table 12 indicates the format of instructions used with the test.

**Table 12.** Responses to question: “What format of instructions did you use?”

| **Instruction Format** | **Group 1:**<br>**LFA +**<br>**fluid**<br>**transfer**<br>**(n)** | **Group 1:**<br>**LFA +**<br>**fluid**<br>**transfer**<br>**(%)** | **Group 2:**<br>**LFA**<br>**without**<br>**fluid**<br>**transfer**<br>**(n)** | **Group 2:**<br>**LFA**<br>**without**<br>**fluid**<br>**transfer**<br>**(%)** | **Group 3:**<br>**LFA +**<br>**reader**<br>**(n)** | **Group 3:**<br>**LFA +**<br>**reader**<br>**(%)** |
| --- | --- | --- | --- | --- | --- | --- |
| **Paper** | 149 | 71.98 | 23 | 74.19 | 10 | 9.71 |
| **Digital (webpages,**<br>**downloaded, etc.)** | 23 | 11.11 | 4 | 12.90 | 24 | 23.30 |
| **Associated Covid Test**<br>**App (Smartphone)** | 17 | 8.21 | 3 | 9.68 | 85 | 82.52 |
| **None of the above** | 22 | 10.63 | 3 | 9.68 | 4 | 3.88 |

Table 13 indicates the number and percent of users who were able to access electronic information via a QR Code.

**Table 13.** Responses to question: “Were you able to access electronic information via a QR Code?”

| **Accessed Electronic**<br>**Information via QR Code** | **Group 1:**<br>**LFA +**<br>**fluid**<br>**transfer**<br>**(n)** | **Group 1:**<br>**LFA +**<br>**fluid**<br>**transfer**<br>**(%)** | **Group 2:**<br>**LFA**<br>**without**<br>**fluid**<br>**transfer**<br>**(n)** | **Group 2:**<br>**LFA**<br>**without**<br>**fluid**<br>**transfer**<br>**(%)** | **Group 3:**<br>**LFA +**<br>**reader**<br>**(n)** | **Group 3:**<br>**LFA +**<br>**reader**<br>**(%)** |
| --- | --- | --- | --- | --- | --- | --- |
| **Yes** | 15 | 7.25 | 1 | 3.23 | 17 | 16.50 |
| **No** | 19 | 9.18 | 4 | 12.90 | 71 | 68.93 |

### 5.8 Ease of Use for Opening Packaging

Table 14 indicates the ease of use for opening the test packaging without assistance.

**Table 14.**
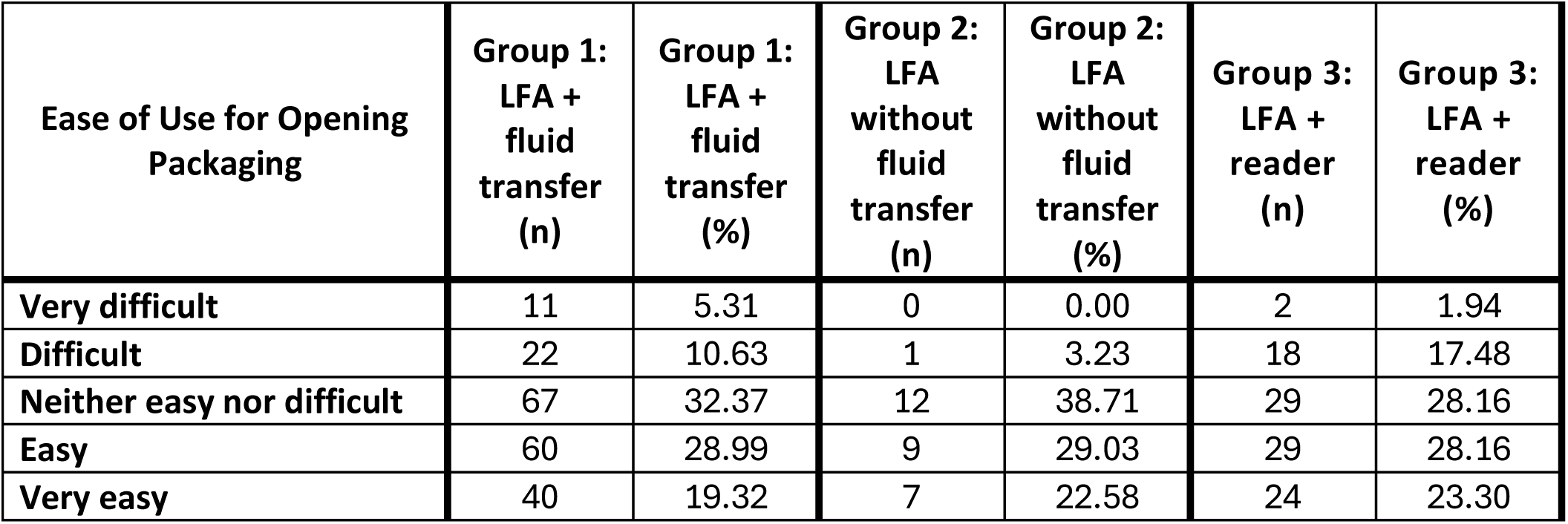
Responses to question: “How easy was opening the package of the [test name] test for you? (without assistance)”

### 5.9 Ease of Use for Completing Test Protocol

Table 15 provides the ease of use for completing the test protocol without assistance.

**Table 15.** Responses to question: “How easy was completing the test protocol for you? (without assistance)”

| Ease of Use for Completing Test Protocol | Group 1: LFA + fluid transfer (n) | Group 1: LFA + fluid transfer (%) | Group 2: LFA without fluid transfer (n) | Group 2: LFA without fluid transfer (%) | Group 3: LFA + reader (n) | Group 3: LFA + reader (%) |
| --- | --- | --- | --- | --- | --- | --- |
| Very difficult | 49 | 23.67 | 9 | 29.03 | 26 | 25.24 |
| Difficult | 41 | 19.81 | 3 | 9.68 | 29 | 28.16 |
| Neither easy nor difficult | 43 | 20.77 | 10 | 32.26 | 20 | 19.42 |
| Easy | 38 | 18.36 | 3 | 9.68 | 20 | 19.42 |
| Very easy | 27 | 13.04 | 4 | 12.90 | 6 | 5.83 |

### 5.10 Interpreting Test Results

Table 16 indicates the ease of use for interpreting test results without assistance.

**Table 16.** Responses to question: “How easy was interpreting the test results for you? (without assistance)”

| Ease of Use for Interpreting Test Results | Group 1: LFA + fluid transfer (n) | Group 1: LFA + fluid transfer (%) | Group 2: LFA without fluid transfer (n) | Group 2: LFA without fluid transfer (%) | Group 3: LFA + reader (n) | Group 3: LFA + reader (%) |
| --- | --- | --- | --- | --- | --- | --- |
| Very difficult | 63 | 30.43 | 11 | 35.48 | 15 | 14.56 |
| Difficult | 21 | 10.14 | 3 | 9.68 | 6 | 5.83 |
| Neither easy nor difficult | 25 | 12.08 | 5 | 16.13 | 12 | 11.65 |
| Easy | 46 | 22.22 | 2 | 6.45 | 25 | 24.27 |
| Very easy | 44 | 21.26 | 6 | 19.35 | 41 | 39.81 |

Table 17 indicates whether or not users were able to interpret test results without assistance.

**Table 17.** Responses to question: “Were you able to interpret the test results? (without assistance)”

| Able to Interpret Results? | Group 1: LFA + fluid | Group 1: LFA + fluid | Group 2: LFA without fluid | Group 2: LFA without fluid | Group 3: LFA + reader (n) | Group 3: LFA + reader (%) |
| --- | --- | --- | --- | --- | --- | --- |

|  | **transfer**<br>**(n)** | **transfer**<br>**(%)** | **transfer**<br>**(n)** | **transfer**<br>**(%)** |  |  |
| --- | --- | --- | --- | --- | --- | --- |
| **Yes** | 121 | 58.45 | 15 | 48.39 | 77 | 74.76 |
| **No** | 82 | 39.61 | 13 | 41.94 | 24 | 23.30 |

### 5.11 Associated App

Table 18 indicates the ease of use of an app associated with the test.

**Table 18.** Responses to question: “If there was an associated app for the test, how easy was it for you to use (without assistance)?”

| **App's Ease of Use** | **Group 1:**<br>**LFA +**<br>**fluid**<br>**transfer**<br>**(n)** | **Group 1:**<br>**LFA +**<br>**fluid**<br>**transfer**<br>**(%)** | **Group 2:**<br>**LFA**<br>**without**<br>**fluid**<br>**transfer**<br>**(n)** | **Group 2:**<br>**LFA**<br>**without**<br>**fluid**<br>**transfer**<br>**(%)** | **Group 3:**<br>**LFA +**<br>**reader**<br>**(n)** | **Group 3:**<br>**LFA +**<br>**reader**<br>**(%)** |
| --- | --- | --- | --- | --- | --- | --- |
| **Very difficult** | 4 | 1.93 | 2 | 6.45 | 9 | 8.74 |
| **Difficult** | 7 | 3.38 | 0 | 0.00 | 22 | 21.36 |
| **Neither easy nor difficult** | 9 | 4.35 | 2 | 6.45 | 17 | 16.50 |
| **Easy** | 9 | 4.35 | 1 | 3.23 | 31 | 30.10 |
| **Very easy** | 4 | 1.93 | 0 | 0.00 | 17 | 16.50 |
| **There was not an app** | 0 | 0.00 | 0 | 0.00 | 0 | 0.00 |
| **There is an app, but I**<br>**didn't use it** | 58 | 28.02 | 11 | 35.48 | 3 | 2.91 |

Table 19 indicates how helpful participants found the app for completing the test.

**Table 19.** Responses to question: “How helpful was the app in assisting you with completing the test?”

| **Helpfulness of App** | **Group 1:**<br>**LFA +**<br>**fluid**<br>**transfer**<br>**(n)** | **Group 1:**<br>**LFA +**<br>**fluid**<br>**transfer**<br>**(%)** | **Group 2:**<br>**LFA**<br>**without**<br>**fluid**<br>**transfer**<br>**(n)** | **Group 2:**<br>**LFA**<br>**without**<br>**fluid**<br>**transfer**<br>**(%)** | **Group 3:**<br>**LFA +**<br>**reader**<br>**(n)** | **Group 3:**<br>**LFA +**<br>**reader**<br>**(%)** |
| --- | --- | --- | --- | --- | --- | --- |
| **Very unhelpful** | 8 | 3.86 | 2 | 6.45 | 12 | 11.65 |
| **Unhelpful** | 5 | 2.42 | 1 | 3.23 | 17 | 16.50 |
| **Neither unhelpful nor**<br>**helpful** | 9 | 4.35 | 0 | 0.00 | 7 | 6.80 |
| **Helpful** | 5 | 2.42 | 2 | 6.45 | 39 | 37.86 |
| **Very helpful** | 6 | 2.90 | 0 | 0.00 | 20 | 19.42 |

### 5.12 Valid Test Results

Table 20 indicates the number and percent of users of each group of tests who received a valid test result.

**Table 20.** Responses to question: “Did you receive a valid test result with the [test name] test (positive or negative)?”

| **Received Valid Test Result?** | **Group 1:**<br>**LFA +**<br>**fluid**<br>**transfer**<br>**(n)** | **Group 1:**<br>**LFA +**<br>**fluid**<br>**transfer**<br>**(%)** | **Group 2:**<br>**LFA**<br>**without**<br>**fluid**<br>**transfer**<br>**(n)** | **Group 2:**<br>**LFA**<br>**without**<br>**fluid**<br>**transfer**<br>**(%)** | **Group 3:**<br>**LFA +**<br>**reader**<br>**(n)** | **Group 3:**<br>**LFA +**<br>**reader**<br>**(%)** |
| --- | --- | --- | --- | --- | --- | --- |
| **Yes** | 183 | 88.41 | 28 | 90.32 | 85 | 82.52 |
| **No** | 19 | 9.18 | 2 | 6.45 | 17 | 16.50 |

### 5.13 Customer Support

Table 21 indicates the number and percent of users of each group of tests who attempted to contact customer support.

**Table 21.** Responses to question: “Did you attempt to contact [test name]’s customer support?”

| **Tried to Contact Customer Support?** | **Group 1:**<br>**LFA +**<br>**fluid**<br>**transfer**<br>**(n)** | **Group 1:**<br>**LFA +**<br>**fluid**<br>**transfer**<br>**(%)** | **Group 2:**<br>**LFA**<br>**without**<br>**fluid**<br>**transfer**<br>**(n)** | **Group 2:**<br>**LFA**<br>**without**<br>**fluid**<br>**transfer**<br>**(%)** | **Group 3:**<br>**LFA +**<br>**reader**<br>**(n)** | **Group 3:**<br>**LFA +**<br>**reader**<br>**(%)** |
| --- | --- | --- | --- | --- | --- | --- |
| **Yes** | 4 | 1.93 | 1 | 3.23 | 6 | 5.83 |
| **No** | 201 | 97.10 | 28 | 90.32 | 95 | 92.23 |

Table 22 indicates the number and percent of users of each group of tests who were able to talk with customer support.

**Table 22.** Responses to question: “Were you able to talk with customer support?”

| **Talked to Customer Support?** | **Group 1:**<br>**LFA +**<br>**fluid**<br>**transfer**<br>**(n)** | **Group 1:**<br>**LFA +**<br>**fluid**<br>**transfer**<br>**(%)** | **Group 2:**<br>**LFA**<br>**without**<br>**fluid**<br>**transfer**<br>**(n)** | **Group 2:**<br>**LFA**<br>**without**<br>**fluid**<br>**transfer**<br>**(%)** | **Group 3:**<br>**LFA +**<br>**reader**<br>**(n)** | **Group 3:**<br>**LFA +**<br>**reader**<br>**(%)** |
| --- | --- | --- | --- | --- | --- | --- |
| **Yes** | 2 | 0.97 | 0 | 0.00 | 4 | 3.88 |
| **No** | 1 | 0.48 | 1 | 3.23 | 2 | 1.94 |

Table 23 indicates how helpful users found customer support in answering their question(s).

**Table 23.** Responses to question: “Was customer support helpful in answering your question(s)?”

| Helpfulness of Customer Support | Group 1:<br>LFA +<br>fluid<br>transfer<br>(n) | Group 1:<br>LFA +<br>fluid<br>transfer<br>(%) | Group 2:<br>LFA<br>without<br>fluid<br>transfer<br>(n) | Group 2:<br>LFA<br>without<br>fluid<br>transfer<br>(%) | Group 3:<br>LFA +<br>reader<br>(n) | Group 3:<br>LFA +<br>reader<br>(%) |
| --- | --- | --- | --- | --- | --- | --- |
| Very unhelpful | 0 | 0.00 | 0 | 0.00 | 1 | 0.97 |
| Unhelpful | 0 | 0.00 | 0 | 0.00 | 2 | 1.94 |
| Neither unhelpful nor helpful | 1 | 0.48 | 0 | 0.00 | 0 | 0.00 |
| Helpful | 1 | 0.48 | 0 | 0.00 | 1 | 0.97 |
| Very helpful | 0 | 0.00 | 0 | 0.00 | 0 | 0.00 |

### 5.14 Change in Experience over Time

Participants who performed a test multiple times were asked about the change in their experience with the test over time. Responses to this question are given in Table 24.

**Table 24.** Responses to question: “If you have performed the [test name] test multiple times, how did your experience change?”

| Change in Experience with Test Over Time | Group 1:<br>LFA +<br>fluid<br>transfer<br>(n) | Group 1:<br>LFA +<br>fluid<br>transfer<br>(%) | Group 2:<br>LFA<br>without<br>fluid<br>transfer<br>(n) | Group 2:<br>LFA<br>without<br>fluid<br>transfer<br>(%) | Group 3:<br>LFA +<br>reader<br>(n) | Group 3:<br>LFA +<br>reader<br>(%) |
| --- | --- | --- | --- | --- | --- | --- |
| Much worse | 3 | 1.45 | 0 | 0.00 | 0 | 0.00 |
| Worse | 7 | 3.38 | 1 | 3.23 | 2 | 1.94 |
| Neither better nor worse | 83 | 40.10 | 11 | 35.48 | 26 | 25.24 |
| Better | 36 | 17.39 | 4 | 12.90 | 22 | 21.36 |
| Much better | 17 | 8.21 | 0 | 0.00 | 11 | 10.68 |
| N/A | 0 | 0.00 | 0 | 0.00 | 0 | 0.00 |

### 5.15 Final questions

Table 25 presents participants’ feelings toward theoretically receiving only digital instructions in future tests (i.e., no printed instructions received with test kits).

**Table 25.** Feelings toward all digital instructions.

| Feeling | n | % |
| --- | --- | --- |
| **Extremely negatively** | 45 | 11.4 |
| **Somewhat negatively** | 57 | 14.5 |
| **Neither negatively nor positively** | 92 | 23.4 |
| **Somewhat positively** | 100 | 25.4 |
| **Extremely positively** | 98 | 24.9 |

This work is supported by NIH grants U54 EB027690 02S1 and U54 EB027690 05S1. The content is solely the responsibility of the authors and does not necessarily represent the official views of the National Institutes of Health.

## 6 Appendix

Raw unprocessed data exported from Qualtrics: this includes all responses received prior to data cleaning

COVID-19 Home Test Survey

**June 14th 2024, 12:26 pm MDT**

### Q1 - What is your age group?

**Table 26.** Stats for Q1: “What is your age group?”

| Field | Minimum | Maximum | Mean | Std Deviation | Variance | Count |
| --- | --- | --- | --- | --- | --- | --- |
| What is your age group? | 1.00 | 8.00 | 4.33 | 1.64 | 2.70 | 624 |

**Table 27.**
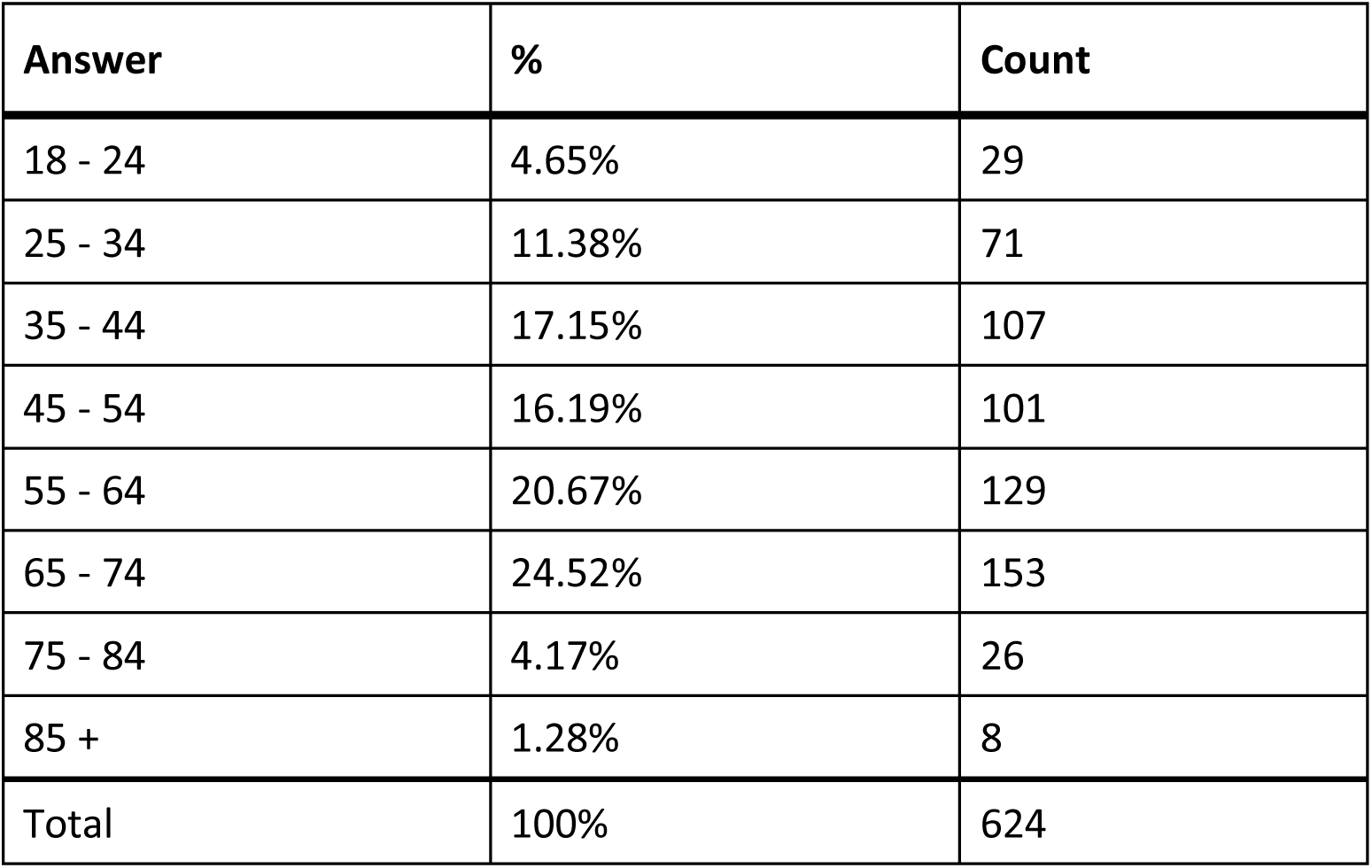
Responses to Q1: “What is your age group?”

### Q2 - Do you identify with any of the following? If so, indicate all that apply

**Table 28.**
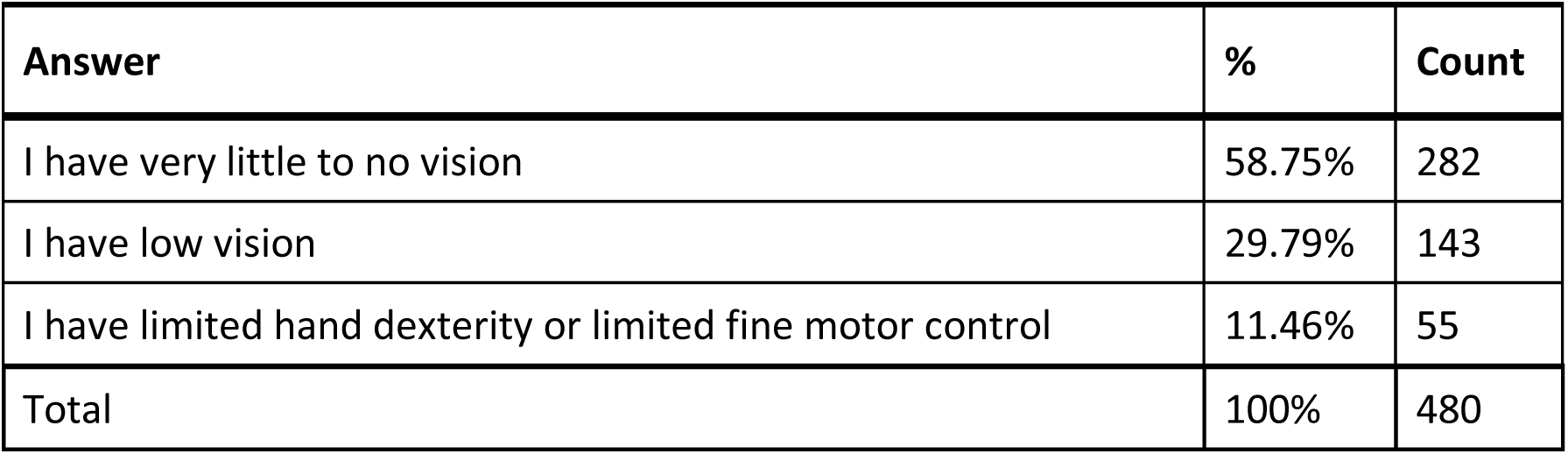
Disability identification. Q2: “Do you identify with any of the following?”

### Q211 - Do you have a smartphone or tablet?

**Table 29.** Stats for Q211: “Do you have a smartphone or tablet?”

| Field | Minimum | Maximum | Mean | Std Deviation | Variance | Count |
| --- | --- | --- | --- | --- | --- | --- |
| Do you have a smartphone or tablet? | 1.00 | 2.00 | 1.06 | 0.23 | 0.05 | 601 |

**Table 30.** Responses to Q211: “Do you have a smartphone or tablet?”

| Answer | % | Count |
| --- | --- | --- |
| Yes | 94.34% | 567 |
| No | 5.66% | 34 |
| Total | 100% | 601 |

### Q212 - How would you rate your smartphone, tablet, and computer skills?

**Table 31.** Stats for Q212: “How would you rate your smartphone, tablet, and computer skills?”

| Field | Minimum | Maximum | Mean | Std Deviation | Variance | Count |
| --- | --- | --- | --- | --- | --- | --- |
| How would you rate your smartphone, tablet, and computer skills? | 1.00 | 5.00 | 1.77 | 0.93 | 0.87 | 548 |

**Table 32.** Q212: Response to “How would you rate your smartphone, tablet, and computer skills?”

| Answer | % | Count |
| --- | --- | --- |
| Extremely proficient | 50.73% | 278 |
| Somewhat proficient | 26.82% | 147 |
| Average | 17.70% | 97 |
| Somewhat limited | 3.83% | 21 |
| Extremely limited | 0.91% | 5 |
| Total | 100% | 548 |

### Q207 - Are you associated with any of the following groups?

**Table 33.** Group affiliation. Responses to Q207: “Are you associated with any of the following groups?”

| Answer | % | Count |
| --- | --- | --- |
| National Federation of the Blind | 50.89% | 258 |
| American Council of the Blind | 27.61% | 140 |
| Administration for Community Living | 4.14% | 21 |
| Other | 8.68% | 44 |
| Protection and Advocacy System | 3.75% | 19 |
| Client Assistance Program | 2.17% | 11 |
| World Institute on Disability (WID) | 2.76% | 14 |
| Total | 100% | 507 |

#### Q207_4_TEXT – Other

**Table 34.** Open response to Other - Group Affiliations.

| Other – Text |
| --- |
| Licensed Home Care Agency |
| Independent Living Centers - currently Brooklyn Center for Independence of the Disabled, formerly National Council on Independent Living |
| ITEM Coalition |
| AARP |
| AARP |
| North Carolina Deafblind Associates, Division of Services for the Deaf and Hard of Hearing |
| Lighthouse for the Blind |
| Lighthouse for the Blind of the Palm Beaches |
| Guide dogs for the blind |
| TN Family Support Program Council, TN Disability Coalition employee |
| National council of independent living an associate progress for rule, independent living |
| National Multiple Sclerosis Society |
| Ncidd |
| Cal Voices |
| Commission For The Blind, American foundation for the blind. |
| Lions club |
| none of the above |
| American Association of People with Disabilities |
| Parent to a blind child |
| Center for Independent Living |
| Foundation Fighting Blindness (FFB) |
| Future in Sight |
| Inspirational three foundation Inc. I'm number three and support group call Inspirational Three Support On and JOICE Brooks. Louis Steve JOICE Brooks. Ranky crack and Lois BAYS |
| The Arc |
| I work with them but am not a "member" of them. (NFB ACB WID) and receive grant funding from ACL |
| M.A., Vision Rehabilitation Therapist Intern for Earle Baum Center of the Blind, Calif. |
| ADA Cleveland |
| UVM Center on Disability & Community Inclusion (VT UCEDD) |
| Disabilities Right's Advocate/ Advisory |
| Member of ACB, have interacted with others I've checked, but do not work at any of them. |
| National Council Independent Living |
| Testing CENA staff |
| Foundation Fighting Blindness |
| My nonprofit |
| National Disability Rights Network, American Civil Liberties Union, Mental Health America, Association of Blind Citizens, American Foundation for the Blind, American Printing House for the Blind, Blind LGBT Pride International |
| National Disability Rights Network |

### Q3 - Have you taken any COVID-19 home tests in the last year?

**Table 35.** Stats for Q3: “Have you taken any COVID-19 home tests in the last year?”

| Field | Minimum | Maximum | Mean | Std Deviation | Variance | Count |
| --- | --- | --- | --- | --- | --- | --- |
| Have you taken any COVID-19 home tests in the last year? | 1.00 | 2.00 | 1.12 | 0.33 | 0.11 | 615 |

**Table 36.**
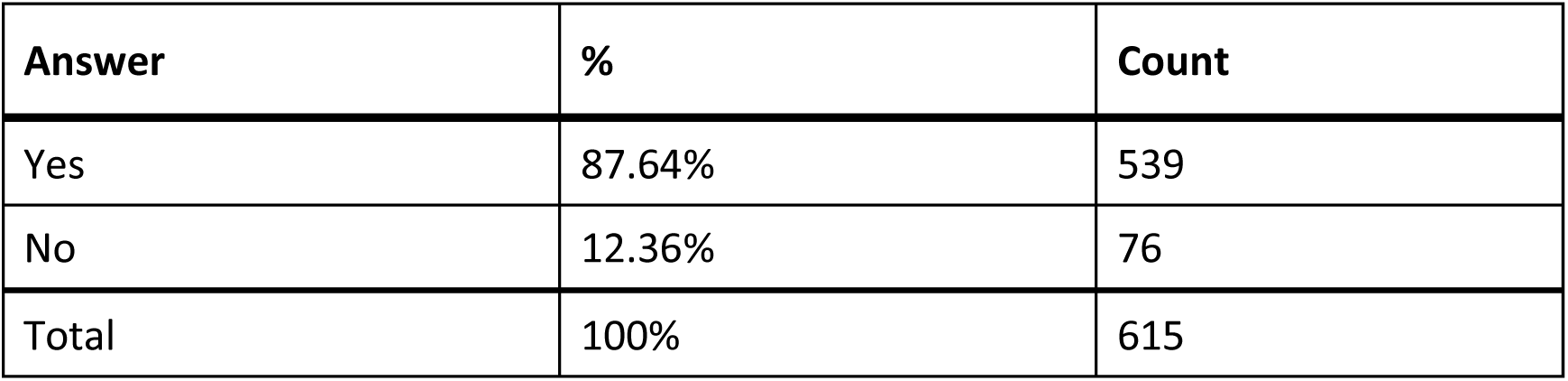
Responses to Q3: “Have you taken any COVID-19 home tests in the last year?”

### Q4 - Which test(s) have you used?

**Table 37.** Responses to Q4: “Which test(s) have you used?”

| Answer | % | Count |
| --- | --- | --- |
| BinaxNow | 17.93% | 144 |
| iHealth | 17.31% | 139 |
| QuickVue | 6.35% | 51 |
| CareStart | 1.25% | 10 |
| Flowflex | 4.73% | 38 |
| Roche | 2.74% | 22 |
| InteliSwab | 1.99% | 16 |
| On/Go | 2.24% | 18 |
| Ellume | 18.31% | 147 |
| Other | 6.10% | 49 |
| Siemens Clinitest | 2.37% | 19 |
| I don't know | 18.68% | 150 |
| Total | 100% | 803 |

#### Q4_10_TEXT – Other

**Table 38.** Open response to Q4: “Which test(s) have you used?”

| **Other - Text** |
| --- |
| Kevin's Covid Test |
| Cue Health |
| Kevin's Covid Test |
| Kevin's Covid Test |
| Cue Health |
| Kevin's Covid Test |
| ABC |
| Kevin's Covid Test |
| I don't know |
| Q Health |
| I don't know which it was. |
| Quidel |
| BTNX |
| Cue Health |
| The rapid tests that were provided by the government |
| They were the home tests that were sent through the US Postal Service and the ones that Walgreens was giving to Medicare people for free. I have no idea what brand they were. I did them with the full assistance of a cited friend. |
| Cue Health (Nucleic Acid Amplification Test) |
| Don't know. My home nurse administered it and read it off. |
| Only tested with the accessible kit |
| Non-electronic test kit. |
| Cue Health |
| SD Biosensor |
| Labcorp's Pixel On Demand where you send swab to lab for processing. |
| Cue health |
| Azure FasTep BD Veriyor Maxim ClearDetect |
| Lepu Medical SARS-CoV-2 Antigen Rapid Test (Colloidal Gold Immunochromatography) |
| Cue Health and tests that I purchased at my local pharmacy and got from the US federal government Covid testing program, before the Accessible tests were available, but I do not know the brands because I could not read the packaging. |
| Ellume |
| CueHealth |
| I was assisted by a sighted person to administer other kinds of Covid tests, so I'm not sure which kind/kinds they were. |
| the one issued by the Government. Not sure of the name of it. It just says Home Covid Test |
| testing |
| Don't know |
| Cue Health |

### Q208 - iHealth Think of the first time you used the iHealth test. Did you have difficulty completing any of the following tasks? Select all that apply

**Table 39.** Response to Q208: iHealth - Did you have any difficulty completing the following tasks?”

| Answer | % | Count |
| --- | --- | --- |
| Opening the packaging | 10.64% | 30 |
| Reading the instructions | 20.57% | 58 |
| Identifying test components | 12.06% | 34 |
| Collecting the test sample | 7.80% | 22 |
| Manipulating test components | 20.57% | 58 |
| Interpreting the test results | 17.02% | 48 |
| Other | 1.77% | 5 |
| I did not have difficulty completing the test | 9.57% | 27 |
| Total | 100% | 282 |

#### Q208_7_TEXT – Other

**Table 40.** Open response to Other, Q208: “iHealth - Did you have any difficulty completing the following tasks?”

| Other - Text |
| --- |
| Reading instructions on app. |
| Finally had ti get sighted help ti get solution improper vial an test tray even thru was test fir visually handicapped an blind. |
| did not get results due to improper techniques |
| I needed to have someone else administer the test. |
| It would have been very difficult for me to do this w/out sighted assistance. |

### Q209 - What tools or assistance, if any, did you use to perform the test?

**Table 41.** Response to Q209: “iHealth - What tools or assistance, if any, did you use to perform the test?”

| Answer | % | Count |
| --- | --- | --- |
| Timer | 28.82% | 66 |
| Scissors | 17.90% | 41 |
| Remote visual assistance (AIRA, Be My Eyes, Facetime, Video-chat, etc.) | 9.17% | 21 |
| Screen Reader | 4.37% | 10 |
| Magnification | 10.92% | 25 |
| Other technology (OCR, Bar Code Readers, Light Detectors, etc.) | 2.62% | 6 |
| In person human assistance | 18.34% | 42 |
| Other | 3.06% | 7 |
| I did not need any tools or assistance | 4.80% | 11 |
| Total | 100% | 229 |

#### Q209_8_TEXT – Other

**Table 42.** Open responses to Other Q209: “iHealth - What tools or assistance, if any, did you use to perform the test?”

| Other - Text |
| --- |
| Alexa |
| Application Glasses. It was super difficult. |
| Youtube. |
| Spot lighting |
| I didn't require any "extra" tools that I wouldn't normally use to open a package and do something that you would time. |

### Q40 - Were you able to conduct this test on your first try?

**Table 43.** Stats for Q40: “Were you able to conduct this [iHealth] test on your first try?”

| Field | Minimum | Maximum | Mean | Std Deviation | Variance | Count |
| --- | --- | --- | --- | --- | --- | --- |
| Were you able to conduct this test on your first try? | 1.00 | 2.00 | 1.19 | 0.39 | 0.15 | 122 |

**Table 44.**
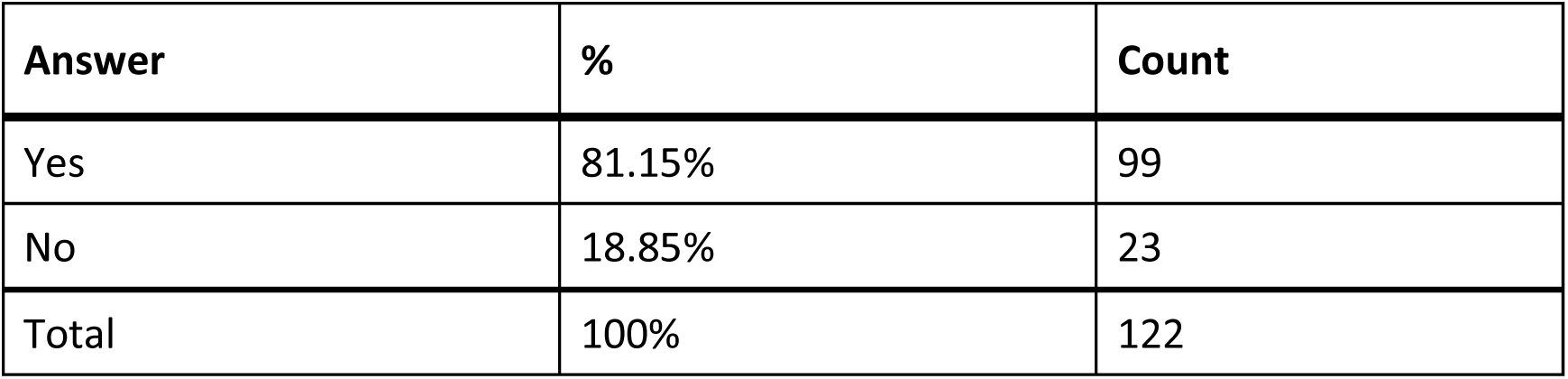
Response to Q40: “Were you able to conduct this [iHealth] test on your first try?”

| Answer | % | Count |
| --- | --- | --- |
| Yes | 81.15% | 99 |
| No | 18.85% | 23 |
| Total | 100% | 122 |

### Q41 - How long did it take to complete the steps required to perform the test, not including the time you spent waiting for results

**Table 45.** Stats for Q41: “How long did it take to complete the steps required to perform the [iHealth] test?”

| Field | Minimum | Maximum | Mean | Std Deviation | Variance | Count |
| --- | --- | --- | --- | --- | --- | --- |
| How long did it take to complete the steps required to perform the test, not including the time you spent waiting for results. | 1.00 | 4.00 | 2.40 | 1.03 | 1.06 | 122 |

**Table 46.** Response to Q41: “How long did it take to complete the steps required to perform the [iHealth] test?”

| Answer | % | Count |
| --- | --- | --- |
| Under 5 minutes | 20.49% | 25 |
| 5 - 10 minutes | 39.34% | 48 |
| 10 - 15 minutes | 19.67% | 24 |
| Over 15 minutes | 20.49% | 25 |
| Total | 100% | 122 |

### Q42 - What format of instructions did you use?

**Table 47.**
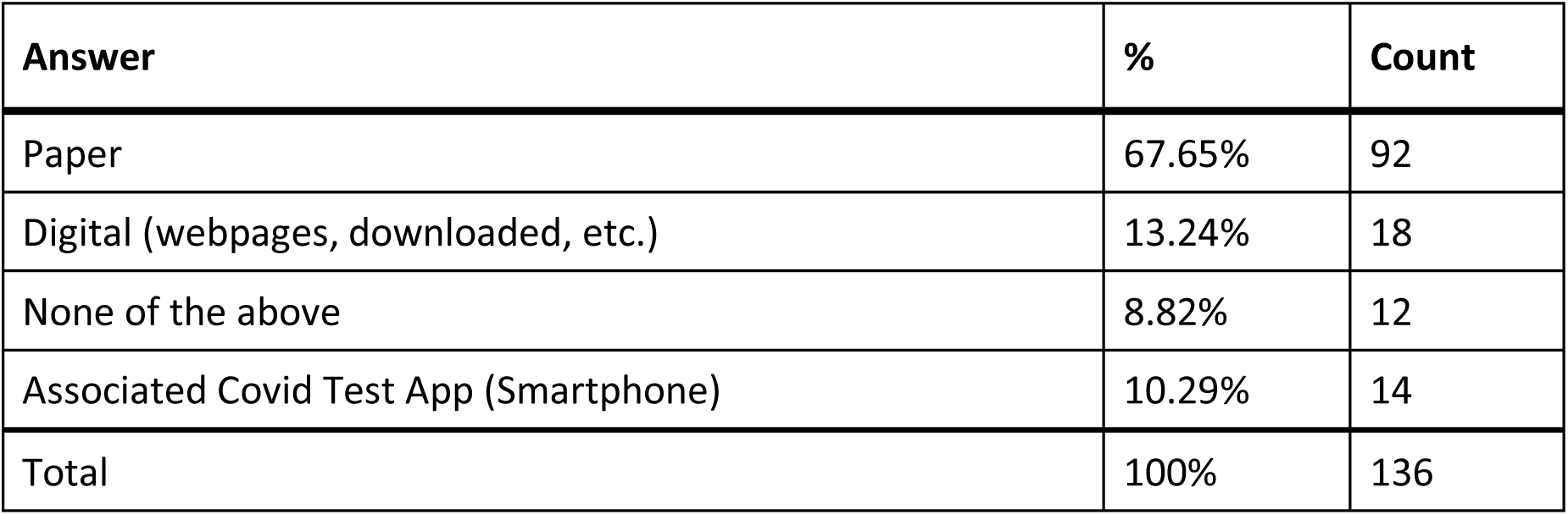
Responses to Q42: “What format of [iHealth] instructions did you use?”

| Answer | % | Count |
| --- | --- | --- |
| Paper | 67.65% | 92 |
| Digital (webpages, downloaded, etc.) | 13.24% | 18 |
| None of the above | 8.82% | 12 |
| Associated Covid Test App (Smartphone) | 10.29% | 14 |
| Total | 100% | 136 |

### Q44 - Were you able to access electronic information via a QR Code?

**Table 48.**
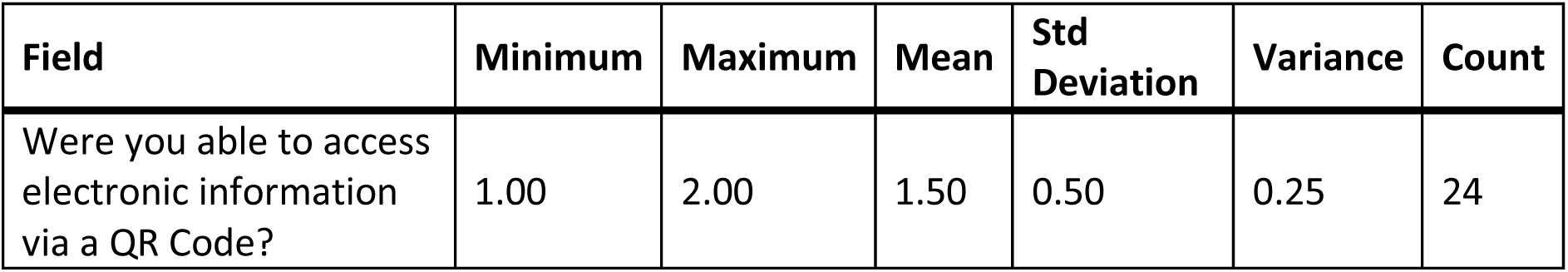
Stats for Q44: “Were you able to access electronic information via a [iHealth] QR code?”

| Field | Minimum | Maximum | Mean | Std Deviation | Variance | Count |
| --- | --- | --- | --- | --- | --- | --- |
| Were you able to access electronic information via a QR Code? | 1.00 | 2.00 | 1.50 | 0.50 | 0.25 | 24 |

**Table 49.** Responses to Q44: “Were you able to access electronic information via a [iHealth] QR code?”

| Answer | % | Count |
| --- | --- | --- |
| Yes | 50.00% | 12 |
| No | 50.00% | 12 |
| Total | 100% | 24 |

### Q611 - How easy was opening the package of the iHealth test for you? (without assistance)

**Table 50.** Stats for Q611: “How easy was completing the [iHealth] test protocol for you? (without assistance)”

| Field | Minimum | Maximum | Mean | Std Deviation | Variance | Count |
| --- | --- | --- | --- | --- | --- | --- |
| How easy was opening the package of the iHealth test for you? (without assistance) | 1.00 | 5.00 | 3.24 | 1.18 | 1.39 | 113 |

**Table 51.** Responses to Q45:“How easy was complete the [iHealth] test protocol for you? (without assistance)”

| Answer | % | Count |
| --- | --- | --- |
| Very difficult | 26.50% | 31 |
| Difficult | 23.08% | 27 |
| Neither easy nor difficult | 17.09% | 20 |
| Easy | 19.66% | 23 |
| Very easy | 13.68% | 16 |
| Total | 100% | 117 |

### Q46 - How easy was interpreting the test results for you? (without assistance)

**Table 52.** Stats for Q46: “How easy was interpreting the [iHealth] test results for you? (without assistance)”

| Field | Minimum | Maximum | Mean | Std Deviation | Variance | Count |
| --- | --- | --- | --- | --- | --- | --- |
| How easy was interpreting the test results for you? (without assistance) | 1.00 | 5.00 | 2.93 | 1.54 | 2.38 | 119 |

**Table 53.** Response to Q46: “How easy was interpreting the [iHealth] test results for you? (without assistance)”

| Answer | % | Count |
| --- | --- | --- |
| Very difficult | 30.25% | 36 |
| Difficult | 12.61% | 15 |
| Neither easy nor difficult | 10.08% | 12 |
| Easy | 27.73% | 33 |
| Very easy | 19.33% | 23 |
| Total | 100% | 119 |

### Q210 - Were you able to interpret the test results? (without assistance)

**Table 54.** Stats for Q210: “Were you able to interpret the [iHealth] test results? (without assistance)”

| Field | Minimum | Maximum | Mean | Std Deviation | Variance | Count |
| --- | --- | --- | --- | --- | --- | --- |
| Were you able to interpret the test results? (without assistance) | 1.00 | 2.00 | 1.38 | 0.48 | 0.23 | 114 |

**Table 55.** Response to Q210: “Were you able to interpret the [iHealth] test results? (without assistance)”

| Answer | % | Count |
| --- | --- | --- |
| Yes | 62.28% | 71 |
| No | 37.72% | 43 |
| Total | 100% | 114 |

### Q47 - If there was an associated app for the test, how easy was it for you to use? (without assistance)

**Table 56.**
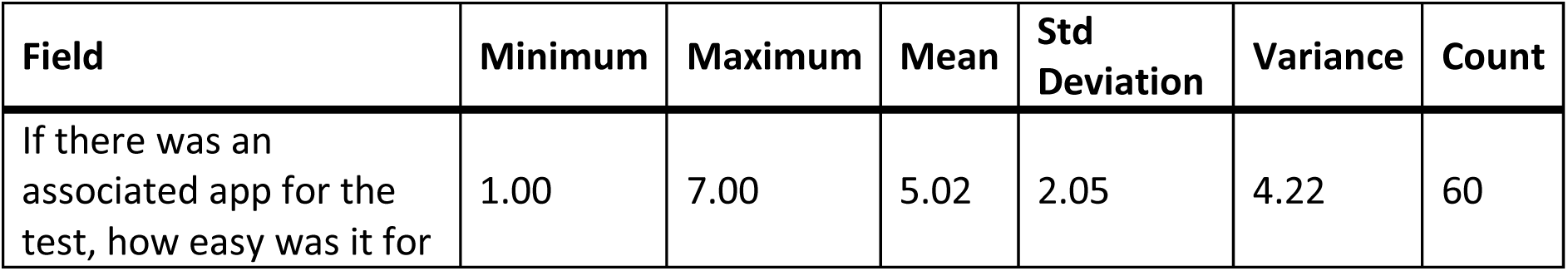
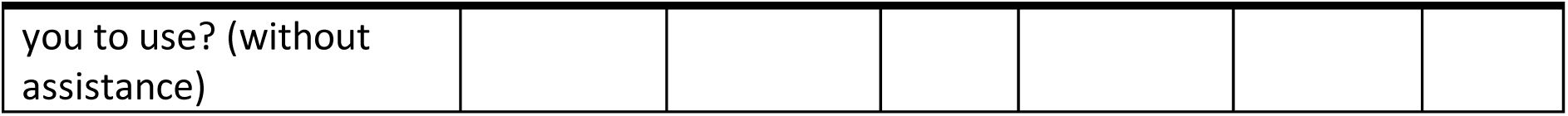
Stats for Q47: “If there was an associated app for the [iHealth] test, how easy was it for you to use? (without assistance)”

**Table 57.** Response to Q47: “If there was an associated app for the [iHealth] test, how easy was it for you to use? (without assistance)”

| Answer | % | Count |
| --- | --- | --- |
| Very difficult | 5.00% | 3 |
| Difficult | 8.33% | 5 |
| Neither easy nor difficult | 16.67% | 10 |
| Easy | 13.33% | 8 |
| Very easy | 10.00% | 6 |
| There is an app, but I didn't use it | 46.67% | 28 |
| Total | 100% | 60 |

### Q417 - How helpful was the app in assisting you with completing the test?

**Table 58.** Stats for Q417: “How helpful was the app in assisting you with completing the [iHealth] test?”

| Field | Minimum | Maximum | Mean | Std Deviation | Variance | Count |
| --- | --- | --- | --- | --- | --- | --- |
| How helpful was the app in assisting you with completing the test? | 1.00 | 5.00 | 3.07 | 1.36 | 1.85 | 27 |

**Table 59.** Response to Q417: “How helpful was the app in assisting you with completing the [iHealth] test?”

| Answer | % | Count |
| --- | --- | --- |
| Very unhelpful | 18.52% | 5 |
| Unhelpful | 14.81% | 4 |
| Neither unhelpful nor helpful | 25.93% | 7 |
| Helpful | 22.22% | 6 |
| Very helpful | 18.52% | 5 |
| Total | 100% | 27 |

### Q48 - Did you receive a valid test result with the iHealth test (positive or negative)?

**Table 60.** Stats for Q48: “Did you receive a valid test result with the iHealth test (positive or negative)?”

| Field | Minimum | Maximum | Mean | Std Deviation | Variance | Count |
| --- | --- | --- | --- | --- | --- | --- |
| Did you receive a valid test result with the iHealth test (positive or negative)? | 1.00 | 2.00 | 1.06 | 0.24 | 0.06 | 118 |

**Table 61.** Response to Q48: “Did you receive a valid test result with the iHealth test (positive or negative)?”

| Answer | % | Count |
| --- | --- | --- |
| Yes | 94.07% | 111 |
| No | 5.93% | 7 |
| Total | 100% | 118 |

### Q50 - Did you attempt to contact iHealth’s customer support?

**Table 62.** Stats for Q50: “Did you attempt to contact iHealth’s customer support?”

| Field | Minimum | Maximum | Mean | Std Deviation | Variance | Count |
| --- | --- | --- | --- | --- | --- | --- |
| Did you attempt to contact iHealth's customer support? | 1.00 | 2.00 | 1.94 | 0.24 | 0.06 | 119 |

**Table 63.** Response to Q50: “Did you attempt to contact iHealth’s customer support?”

| Answer | % | Count |
| --- | --- | --- |
| Yes | 5.88% | 7 |
| No | 94.12% | 112 |
| Total | 100% | 119 |

### Q213 - Were you able to talk with customer support?

**Table 64.** Stats for Q213: “Were you able to talk with [iHealth] customer support?”

| Field | Minimum | Maximum | Mean | Std Deviation | Variance | Count |
| --- | --- | --- | --- | --- | --- | --- |
| Were you able to talk with customer support? | 1.00 | 2.00 | 1.75 | 0.43 | 0.19 | 4 |

**Table 65.**
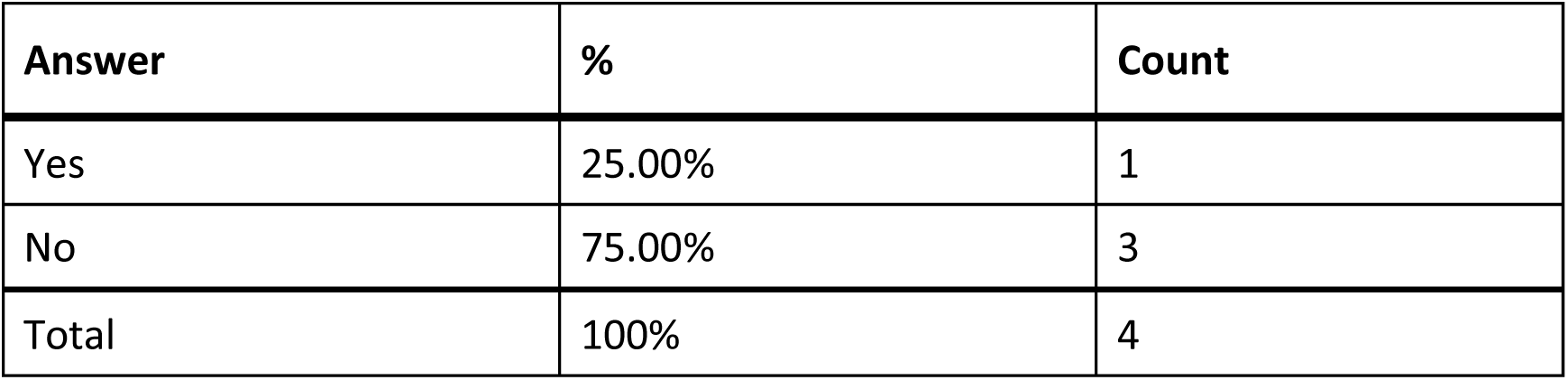
Responses to Q213: “Were you able to talk with [iHealth] customer support?”

| Answer | % | Count |
| --- | --- | --- |
| Yes | 25.00% | 1 |
| No | 75.00% | 3 |
| Total | 100% | 4 |

### Q214 - Was customer support helpful in answering your question(s)?

**Table 66.** Stats for Q214: “Was [iHealth] customer support helpful in answering your question(s)?”

| Field | Minimum | Maximum | Mean | Std Deviation | Variance | Count |
| --- | --- | --- | --- | --- | --- | --- |
| Was customer support helpful in answering your question(s)? | 5.00 | 5.00 | 5.00 | 0.00 | 0.00 | 1 |

**Table 67.** Responses to Q214: “Was [iHealth] customer support helpful in answering your question(s)?

| Answer | % | Count |
| --- | --- | --- |
| Very unhelpful | 0.00% | 0 |
| Unhelpful | 0.00% | 0 |
| Neither helpful nor unhelpful | 100.00% | 1 |
| Helpful | 0.00% | 0 |
| Very helpful | 0.00% | 0 |
| Total | 100% | 1 |

### Q54 - If you have performed the iHealth test multiple times, how did your experience change?

**Table 68.** Stats for Q54: “If you have performed the iHealth test multiple times, how did your experience change?”

| Field | Minimum | Maximum | Mean | Std Deviation | Variance | Count |
| --- | --- | --- | --- | --- | --- | --- |
| If you have performed the iHealth test multiple times, how did your experience change? | 1.00 | 5.00 | 3.48 | 0.90 | 0.80 | 94 |

**Table 69.**
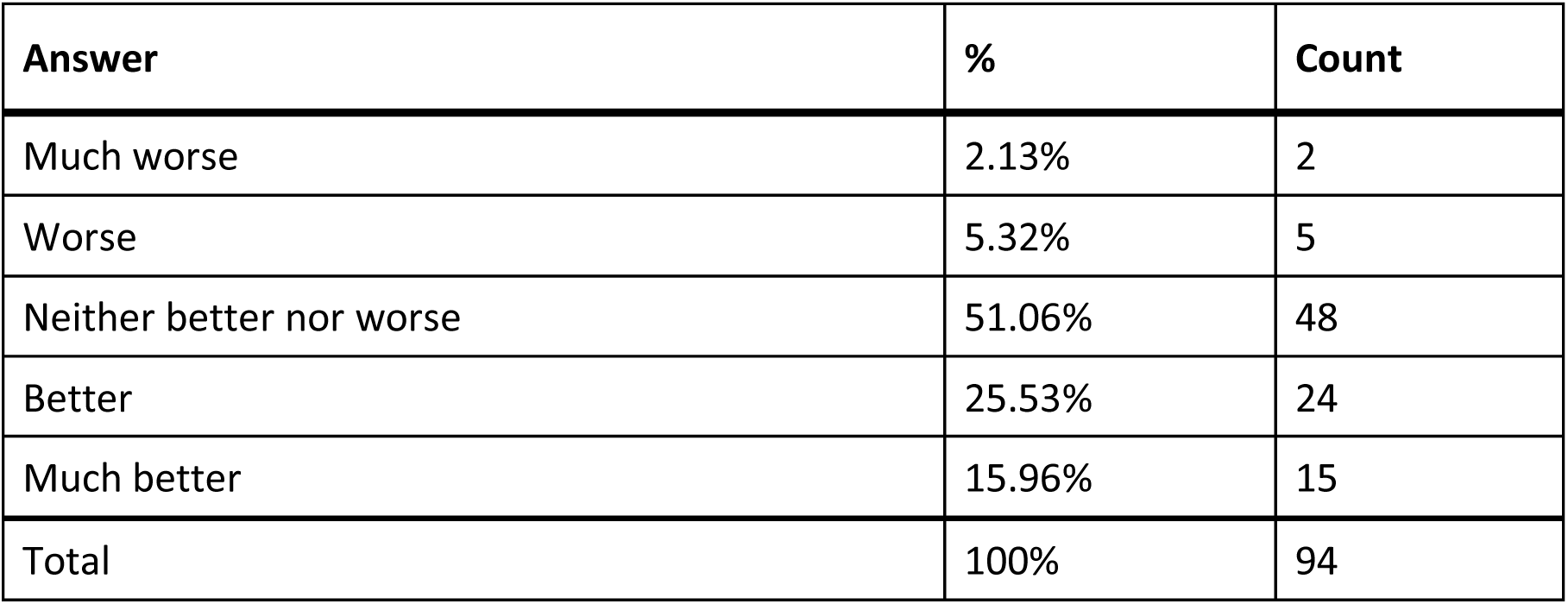
Responses to Q54: “If you have performed the iHealth test multiple times, how did your experience change?”

### Q51 - What did you like about the iHealth test?

**Table 70.** Open responses to Q51: “What did you like about the iHealth test?”

| What did you like about the iHealth test? |
| --- |
| It was easy to undersand and do it. |
| it's Simplicity |
| It was easy to use & free |
| results were clear |
| easy |
| The app |
| It was easy to order on line with my iHealth App. |
| Easy to use |
| Fairly easy |
| it was free |
| Fairly easy to perform and interpret. |
| Easy to use and interpret results. |
| N/A |
| N/A |
| N/A |
| N/A |
| Test |
| I went to ihealth youz/ you tube |
| Didn't have to leave home |
| Availability |
| Half of the supplied packages came premixed in vial. Other half I had to insert mix in vial myself. |
| I didn't have to leave my house to test. It was pretty easy, and very user friendly. |
| Easy to figure out what goes where |
| Easy to use, easy to read |
| Free! No travel to the test site. |
| Clear instructions. |
| easy to read test results |
| Sturdy components, and easy to manipulate. Good that the test solution was already in the container that would already function as a dropper. |
| Easy to use. Paper print needs to be larger font. Too small |
| the end result is easy to read |
| Availability, ease of use |
| a heads up if it showed positive |
| it was free. It was okay if you had some sight. |
| no opinion |
| There were step-by-step instructions that were easy to follow. |
| Easy to understand. |
| I could open the swab and the test thing you put the sample in. I didn't have to jam the swab into my brain. |
| I like that the app was included |
| Inexpensive |
| The iHealth test is pretty easy to do yourself once you know the steps. I am able to set everything up myself and time the test, but cannot read the result. |
| It was easy to use the app. |
| Easy to read |
| Nothing |
| quick |
| There were limited pieces/prep and the components were easy to distinguish. |
| It was free and sent to me in the mail |
| it was portable |
| Nothing |
| The quick results. |
| They tried to come up with one to help the handicapped |
| Compact |
| I thought it was very easy to use. We had to test twice a week at home for many months. For the first test, because we had to do it quickly, my supervisor gave me instructions but I did everything. The next time she watched. I downloaded instructions, tweaked them and was on my own after that. Eventually, we switched to the pre-filled vials because I and a number of other people occasionally had trouble with the other, although I don't know why. |
| Test solution was properly measured and stored for 1 test only. |
| Once I had some practice with taking the test, I found it easy to take the test, with the exception of reading the test results. |
| Everything was very easy, except for interpreting the results |
| It's thought to be the most accurate. Slender package and design. Got used to taking it so would just have to call Aira to read the results. |
| I could do this in easier to open packaging and easier to read result area privacy of my own home. |
| N/A |
| I had to have sided assistance to complete the test. There is nothing about it that I like, I had just ordered them because I did not know what else to order. |
| It worked |
| Rapid result |
| It worked, and I did it independently. |
| Easy to use besides the reactor stuff being hard to open. |
| Nothing |
| Easy to use, especially when the liquid was already in the test vial. |
| It was free |
| it was ffree. |
| Administered for me |
| Even though a blind person can't read the test results, the test is easy to self-administer once one has had a little practice. |
| I didn't like it because it was so difficult to do it independently. |
| Nothing to compare with |
| it was easy to use |
| The app allowed me to store my info in it so I didn't have to re-enter it each time. |
| The test had an app for the phone. |
| I liked being able to test from home instead of needing to drive to a testing center. |
| Testing at home is safer, faster and more convenient |
| I did not have to watch redundant videos |

### Q52 - What would you like to see improved for the iHealth test?

**Table 71.** Open response to Q52: “What would you like to see improved for the iHealth test?”

| What would you like to see improved for the iHealth test? |
| --- |
| More simple user friendly instructions available in various formats |
| cheaper cost |
| there are a few steps to take maybe having the serum already in the tube would help |
| Packaging was too difficult and there were too many small pieces. |
| NA |
| Larger test components, less precision needed to manipulate testing components |
| n/a |
| Nothing |
| less peices to be put together |
| it really hurts my hands to perform almost every step |
| Squeezing that small amount of liquid into the receptacle. |
| The first IHEALTH test I used was difficult to use and somewhat confusing. They dramatically improved usability with the next round. I am totally happy with the current IHealth test. |
| N/A |
| N/A |
| N/A |
| N/A |
| Test |
| I just handed it to someone else to do for me. The only part I handled was the swab. |
| all parts of test kit are very small, at list made them colored, and instruction is very, very small letters... |
| Dark ink /lesswording |
| Easy identifiable components in the text Kit |
| larger tools |
| More premixed vials. |
| The color of the strips test result strips. There is an extremely high number of people in the world who can't or have difficulty seeing red or green. I did almost miss a positive result for my daughter because of this, thankfully my husband was there to check it with me. |
| nothing |
| can't think of anything. Video on their website very helpful! |
| new separation of reagent from the reagent dispenser makes it much more difficult for people with low/no vision |
| The test requires you to only squeeze 4 drops onto the test. Anyone that has any dexterity issues, such as my older parent or any person disabled would have difficulty performing this function. I had to help both my mom and her friends that lived alone because they couldn't control the number of drops and too much would cause a false finding on the test. |
| Same as above |
| I don 't have low vision but the print on the included instruction sheet is way too small! |
| easier opening packaging and accessible simple instruction |
| Larger font on app. A way to read results better than a pregnancy it (using lines) Results in the app would be better. |
| no opinion |
| more accessibility features for blind users |
| It would be nice if the results were accessible without having to get assistance. I wish they would make putting the sample in the fluid a little bit easier as well as putting the fluid into the testing card. |
| An accessible way to interpret test results. |
| Na |
| Have the solution in the thing you swirl the swab in, instead of a separate thing to open and then squeeze into that tube. And label the kits differently for the prefilled ones, so you don't accidentally get the kind you have to fill the tube. And, if we need to squeeze the swab, put a doohickey in the kit that we can close around the tube so pulling out the swab would automatically squeeze it. And a better way to get three drops into the well. If you miss too many times, you have to do another test. |
| zoom in/out would be helpful |
| Explain just to place sample drops in only one hole on strips. |
| It would be ideal to have a way for blind people to read the test result independently. Also, pouring the liquid into the small container is sometimes a little tricky nonvisually. |
| N/A |
| No |
| Braille |
| It is really hard to get the drops on the oval. Is there an easier way? |
| Having the liquid solution in its own packaging, which was TINY and had to be opened and put in the tube was hard. Having the tube contain premeasured solution would be better. It was hard to get the solution out of its packaging and into the tube without spilling. |
| I would like to have been able to access results without assistance. |
| I would like a test that is accessible so that I can do it myself |
| Not needing to transfer liquid |
| more blue tooth type connectivity with results and set up |
| The iHealth test was completely inaccessible to my blind son. Even when using the app he needed my (sighted) assistance for the entire thing. |
| fully accessible for totally blind people without smart phones or other high tech support |
| Accuracy |
| An accessible app or a QR code that will bring up instructions also, it was hard to tell which end of the swab to use while in the packaging and you don't want to touch the swab with your fingers |
| To have the test solution already in the bottle you put sick in |
| Directions |
| Is you to read. Easier to perform. Take the visually challenged into account |
| Just having pre-filled vials as the only option would work. Other than that, I think it works really well. The only other issue I encounter is not being able to get someone from Aira or having a long wait with Be My Eyes. Without either eof those apps, it would not have worked. |
| Easier way to efficiently pour solution into test tube i.e. pointed lid that would fit into the opening of the test chamber. |
| I would like the test to have accessible instructions, and I'd like the test to have an accessible way to indicate the results of the test. |
| I am not sure what could be improved. I did not realize until just now that there was an app that I could've used. I think that would've made it much easier because I would not have been struggling to read the print for instructions on how to complete the test. |
| A way to nonvisually and independently read the results without having to rely on personal or remote sighted assistance. More accessible instructions with nonvisual descriptions. |
| Easier to open packaging and Easier to read test results |
| Use Ellum brand as a blueprint for accessible Covid testing. |
| I would like to be able to take the test independently without cited assistance. At this point, there is no way that would happen. |
| Correct instructions for accessible kit which is different from instructions included which are for non accessible kit |
| Larger font on instructions; less pressure required to squeeze out reagent |
| I'd like for it to bee more tolerant of touch since inserting the swab exactly is difficult. Should it come with a glove that would not contaminate the sample? |
| Make the reactor stuff easier to open please. |
| This test needs to be made completely accessible |
| Always have the liquid in the vial so you don't have to twist off the liquid's top & squeeze. |
| That I did not have to interrupt the results like a pregnancy test. |
| Nonvisual results. No need to count out drops into a tiny hole. No need to pour fluid into tube. |
| accessible instructions, easier administration of the test, accessible results |
| I have no idea. |
| A way for a blind person to read the test results independently. |
| If there was an app, I wasn't aware of its availability. |
| Maybe instruction material |
| once I understood how to use it, nothing |
| It is hard to locate the small hole for the sample and to get the drops in there when you can't see much. |
| It is difficult for me to say b/c I had total sighted help to actually perform the test. I would not have wanted to do it alone. |
| I would like to see the test results improve with accuracy. I know people who have tested negative for COVID-19 with at-home tests yet test positive with the PCR tests the same week. |
| More access to affordable tests. |
| The instructions were convoluted and decanting the liquid from the storage capsule to the test thingie was hard. |
| Large print instructions |

### Q656 - BinaxNow Think of the first time you used the BinaxNow test. Did you have difficulty completing any of the following tasks? Select all that apply

**Table 72.**
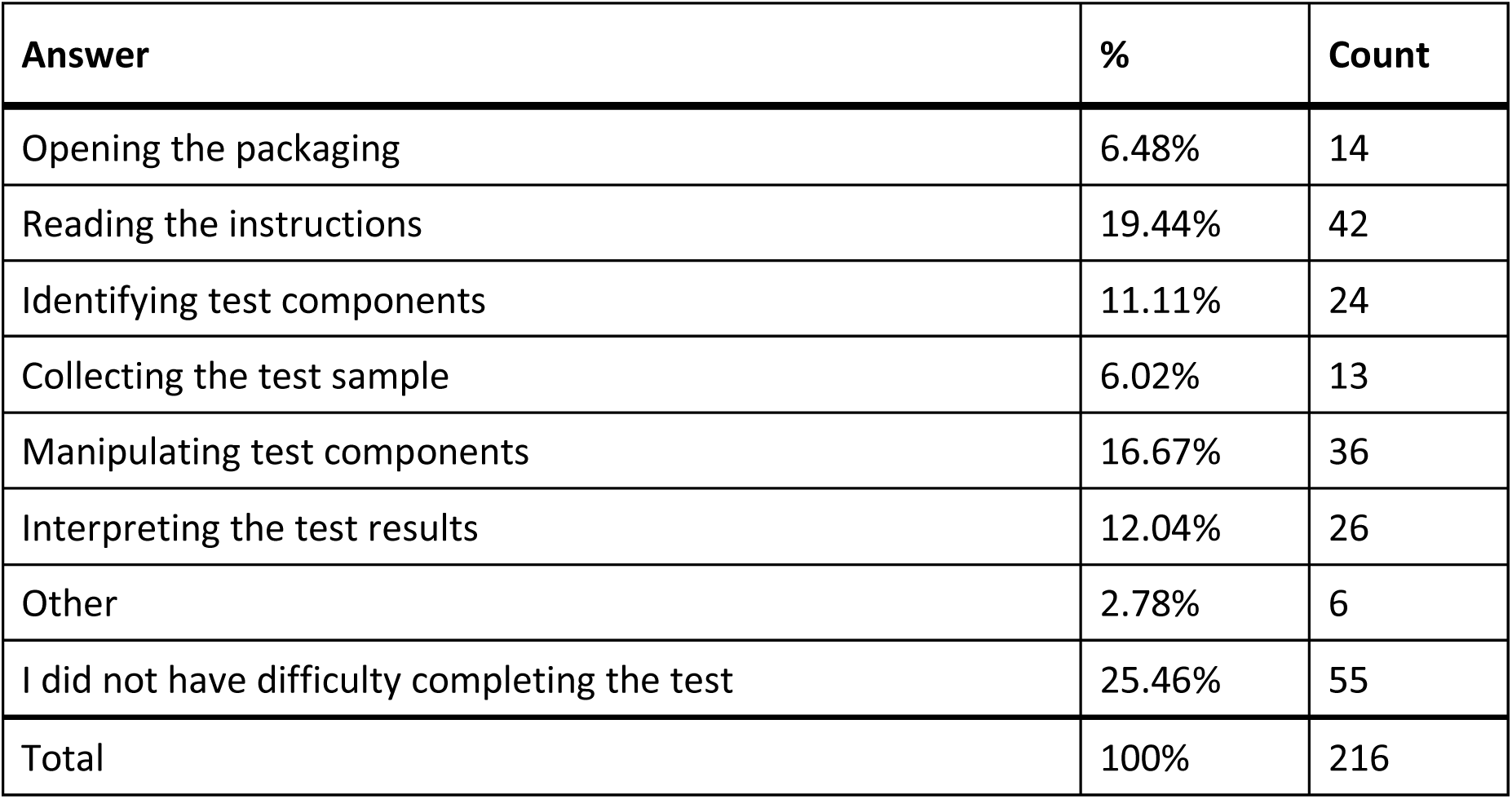
Response to Q656: “Binax Now - Did you have difficulty completing any of the following tasks?”

| Answer | % | Count |
| --- | --- | --- |
| Opening the packaging | 6.48% | 14 |
| Reading the instructions | 19.44% | 42 |
| Identifying test components | 11.11% | 24 |
| Collecting the test sample | 6.02% | 13 |
| Manipulating test components | 16.67% | 36 |
| Interpreting the test results | 12.04% | 26 |
| Other | 2.78% | 6 |
| I did not have difficulty completing the test | 25.46% | 55 |
| Total | 100% | 216 |

#### Q656_7_TEXT – Other

**Table 73.** Open response to Other - Q656: “Binax Now - Did you have difficulty completing any of the following tasks?”

|  |
| --- |
| **Other - Text** |
| I used the AIRA program sponsored by NFB to complete this test. Could not have done it by myself. |
| I don't remember. I had help from another person |
| Counting the drops as they fell on the test page. This was a skill I had to newly master nonvisually. Feeling or hearing a single drop is very tricky! |
| My daughter had to take care of all the steps to do the test except for the swabbing. |
| I needed someone with sight to do this test |
| Understanding the instructions and following them |

### Q657 - What tools or assistance, if any, did you use to perform the test?

**Table 74.**
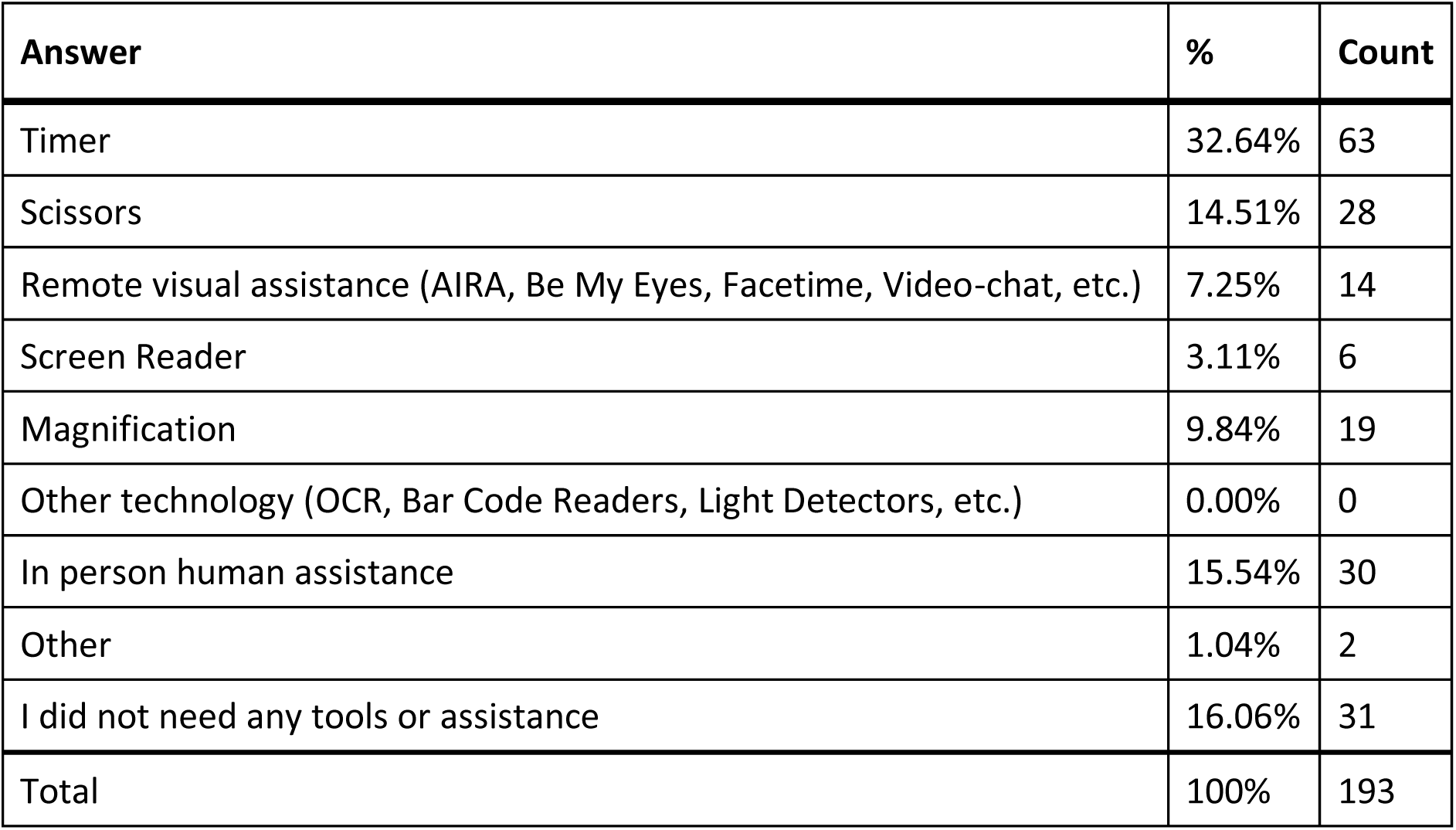
Responses to Q657: “What tools or assistance, if any, did you use to perform the [BinaxNow] test?”

#### Q657_8_TEXT – Other

**Table 75.**
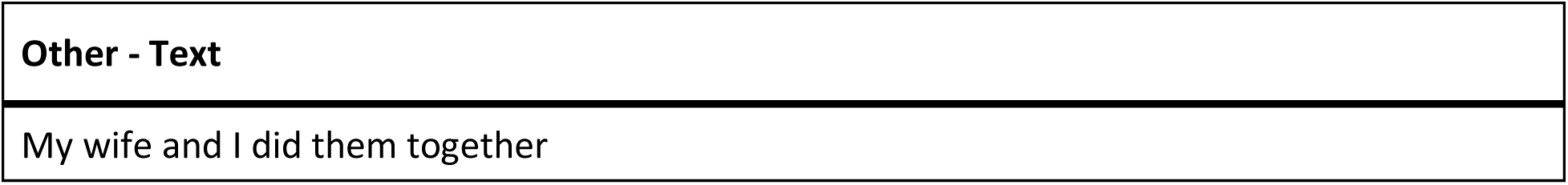
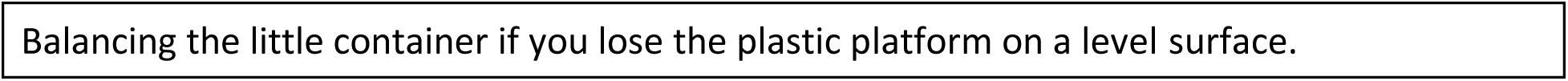
Open response to Other - Q657: “What tools or assistance, if any, did you use to perform the [BinaxNow] test?”

### Q658 - Were you able to conduct this test on your first try?

**Table 76.** Q658: “Were you able to conduct this [BinaxNow] test on your first try?”

| Field | Minimum | Maximum | Mean | Std Deviation | Variance | Count |
| --- | --- | --- | --- | --- | --- | --- |
| Were you able to conduct this test on your first try? | 1.00 | 2.00 | 1.16 | 0.37 | 0.13 | 119 |

**Table 77.** Responses to Q658: “Were you able to conduct this [BinaxNow] test on the first try?”

| Answer | % | Count |
| --- | --- | --- |
| Yes | 84.03% | 100 |
| No | 15.97% | 19 |
| Total | 100% | 119 |

### Q659 - How long did it take to complete the steps required to perform the test, not including the time you spent waiting for results

**Table 78.** Q659: “How long did it take to complete the steps required to perform the [BinaxNow] test, not including the time you spent waiting for results?”

| Field | Minimum | Maximum | Mean | Std Deviation | Variance | Count |
| --- | --- | --- | --- | --- | --- | --- |
| How long did it take to complete the steps required to perform the test, not including the time you spent waiting for results. | 1.00 | 4.00 | 2.07 | 1.06 | 1.13 | 120 |

**Table 79.** Responses to Q659: “How long did it take complete the steps required to perform the [BinaxNow] test?”

| Answer | % | Count |
| --- | --- | --- |
| Under 5 minutes | 39.17% | 47 |
| 5 - 10 minutes | 29.17% | 35 |
| 10 - 15 minutes | 17.50% | 21 |
| Over 15 minutes | 14.17% | 17 |
| Total | 100% | 120 |

### Q660 - What format of instructions did you use?

**Table 80.** Response to Q660: “What format of [BinaxNow] instructions did you use?”

| Answer | % | Count |
| --- | --- | --- |
| Paper | 83.33% | 105 |
| Digital (webpages, downloaded, etc.) | 6.35% | 8 |
| None of the above | 7.14% | 9 |
| Associated Covid Test App (Smartphone) | 3.17% | 4 |
| Total | 100% | 126 |

### Q661 - Were you able to access electronic information via a QR Code?

**Table 81.**
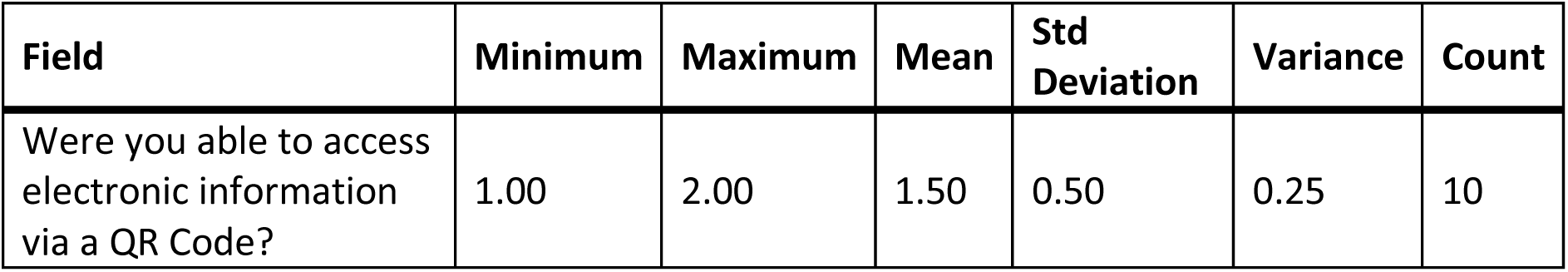
Stats for Q661: “Were you able to access electronic information via a [BinaxNow] QR Code?”

| Field | Minimum | Maximum | Mean | Std Deviation | Variance | Count |
| --- | --- | --- | --- | --- | --- | --- |
| Were you able to access electronic information via a QR Code? | 1.00 | 2.00 | 1.50 | 0.50 | 0.25 | 10 |

**Table 82.**
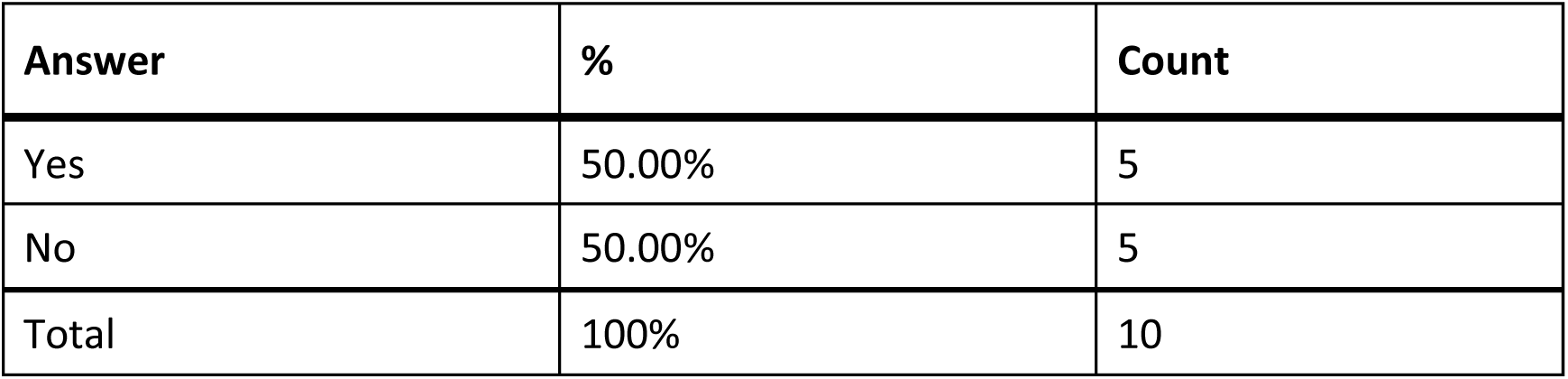
Responses to Q661: “Were you able to access electronic information via [BinaxNow] QR Code?”

| Answer | % | Count |
| --- | --- | --- |
| Yes | 50.00% | 5 |
| No | 50.00% | 5 |
| Total | 100% | 10 |

### Q662 - How easy was opening the package of the BinaxNow test for you? (without assistance)

**Table 83.** Stats for Q662: “How easy was opening the package of the BinaxNow test for you? (without assistance)”

| Field | Minimum | Maximum | Mean | Std Deviation | Variance | Count |
| --- | --- | --- | --- | --- | --- | --- |
| How easy was opening the package of the BinaxNow test for you? (without assistance) | 1.00 | 5.00 | 3.58 | 0.94 | 0.89 | 115 |

**Table 84.** Responses to Q662: “How easy was opening the package of the BinaxNow test for you? (without assistance)”

| Answer | % | Count |
| --- | --- | --- |
| Very difficult | 2.61% | 3 |
| Difficult | 6.09% | 7 |
| Neither easy nor difficult | 40.00% | 46 |
| Easy | 33.04% | 38 |
| Very easy | 18.26% | 21 |
| Total | 100% | 115 |

### Q663 - How easy was completing the test protocol for you? (without assistance)

**Table 85.**
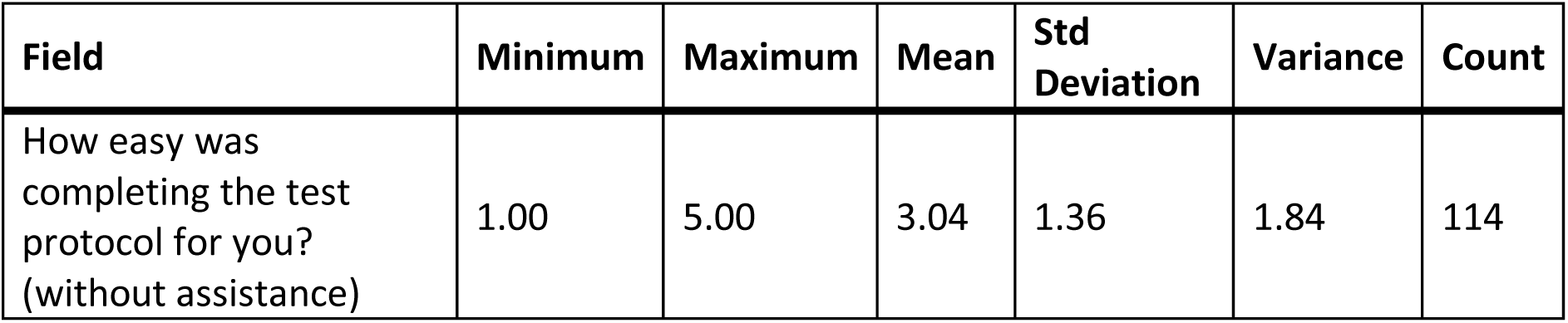
Stats for Q663: “How easy was completing the [BinaxNow] test protocol for you? (without assistance)”

**Table 86.** Response to Q663: “How easy was completing the [BinaxNow] test protocol for you? (without assistance)”

| Answer | % | Count |
| --- | --- | --- |
| Very difficult | 20.18% | 23 |
| Difficult | 14.91% | 17 |
| Neither easy nor difficult | 21.05% | 24 |
| Easy | 28.95% | 33 |
| Very easy | 14.91% | 17 |
| Total | 100% | 114 |

### Q664 - How easy was interpreting the test results for you? (without assistance)

**Table 87.** Stats for Q664: “How easy was interpreting the [BinaxNow] test results for you? (without assistance)”

| Field | Minimum | Maximum | Mean | Std Deviation | Variance | Count |
| --- | --- | --- | --- | --- | --- | --- |
| How easy was interpreting the test results for you? (without assistance) | 1.00 | 5.00 | 3.39 | 1.42 | 2.01 | 114 |

**Table 88.** Responses to Q664:“How easy was interpreting the [BinaxNow] test results for you? (without assistance)”

| Answer | % | Count |
| --- | --- | --- |
| Very difficult | 18.42% | 21 |
| Difficult | 7.89% | 9 |
| Neither easy nor difficult | 15.79% | 18 |
| Easy | 32.46% | 37 |
| Very easy | 25.44% | 29 |
| Total | 100% | 114 |

### Q665 - Were you able to interpret the test results? (without assistance)

**Table 89.**
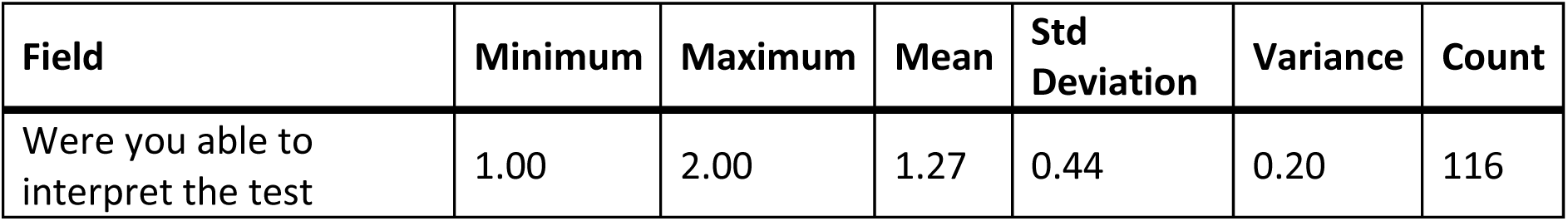
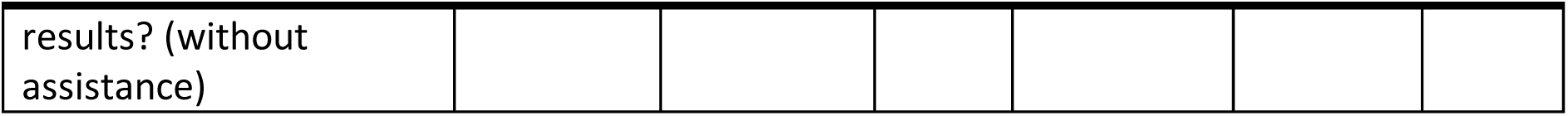
Q665: “Were you able to interpret the test results? (without assistance)”

### Q666 - If there was an associated app for the test, how easy was it for you to use? (without assistance)

**Table 90.** Stats for Q666: “If there was an associated app for the [BinaxNow] test, how easy was it for you to use? (without assistance)”

| Field | Minimum | Maximum | Mean | Std Deviation | Variance | Count |
| --- | --- | --- | --- | --- | --- | --- |
| If there was an associated app for the test, how easy was it for you to use? (without assistance) | 1.00 | 7.00 | 5.63 | 1.95 | 3.81 | 52 |

**Table 91.** Responses to Q666: “If there was an associated app for the [BinaxNow] test, how easy was it for you to use? (without assistance)”

| Answer | % | Count |
| --- | --- | --- |
| Very difficult | 5.77% | 3 |
| Difficult | 1.92% | 1 |
| Neither easy nor difficult | 11.54% | 6 |
| Easy | 11.54% | 6 |
| Very easy | 5.77% | 3 |
| There is an app, but I didn't use it | 63.46% | 33 |
| Total | 100% | 52 |

### Q667 - How helpful was the app in assisting you with completing the test?

**Table 92.**
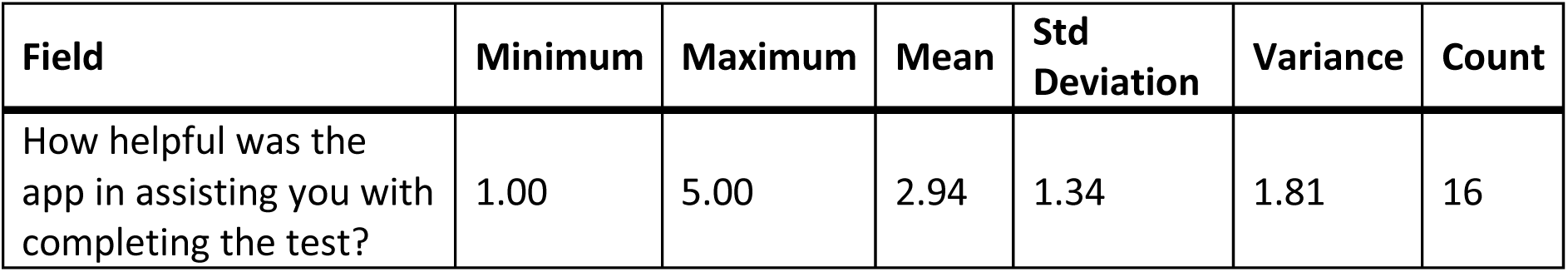
Stats for Q667: “How helpful was the app in assisting you with completing the [BinaxNow] test?”

| Field | Minimum | Maximum | Mean | Std Deviation | Variance | Count |
| --- | --- | --- | --- | --- | --- | --- |
| How helpful was the app in assisting you with completing the test? | 1.00 | 5.00 | 2.94 | 1.34 | 1.81 | 16 |

**Table 93.** Responses to Q668: “How helpful was the app in assisting you with completing the [BinaxNow] test?”

| Answer | % | Count |
| --- | --- | --- |
| Very unhelpful | 25.00% | 4 |
| Unhelpful | 6.25% | 1 |
| Neither unhelpful nor helpful | 31.25% | 5 |
| Helpful | 25.00% | 4 |
| Very helpful | 12.50% | 2 |
| Total | 100% | 16 |

### Q668 - Did you receive a valid test result with the BinaxNow test (positive or negative)?

**Table 94.** Stats for Q668: “Did you receive a valid test result with the BinaxNow test (positive or negative)?”

| Field | Minimum | Maximum | Mean | Std Deviation | Variance | Count |
| --- | --- | --- | --- | --- | --- | --- |
| Did you receive a valid test result with the BinaxNow test (positive or negative)? | 1.00 | 2.00 | 1.06 | 0.24 | 0.06 | 113 |

**Table 95.** Responses to Q668: “Did you receive a valid test result with the BinaxNow test (positive or negative)?”

| Answer | % | Count |
| --- | --- | --- |
| Yes | 93.81% | 106 |
| No | 6.19% | 7 |
| Total | 100% | 113 |

### Q669 - Did you attempt to contact BinaxNow’s customer support?

**Table 96.**
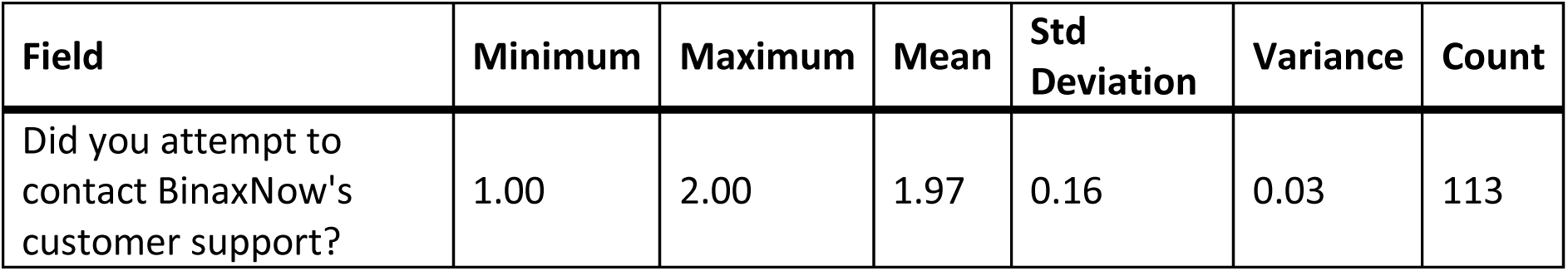
Stats for Q669: “Did you attempt to contact BinaxNow’s customer support?”

| Field | Minimum | Maximum | Mean | Std Deviation | Variance | Count |
| --- | --- | --- | --- | --- | --- | --- |
| Did you attempt to contact BinaxNow's customer support? | 1.00 | 2.00 | 1.97 | 0.16 | 0.03 | 113 |

**Table 97.**
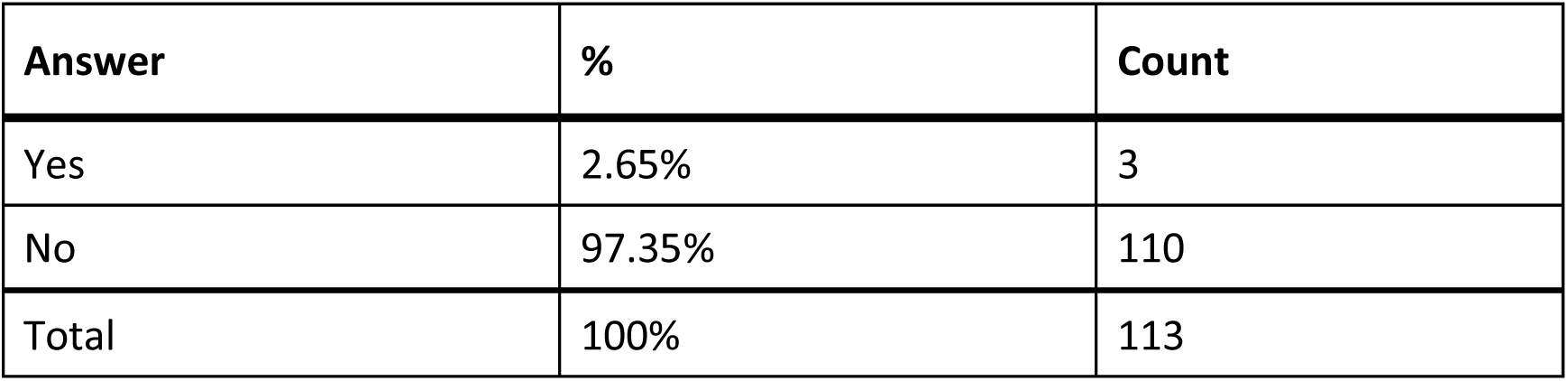
Responses to Q669: “Did you attempt to contact BinaxNow’s customer support?”

| Answer | % | Count |
| --- | --- | --- |
| Yes | 2.65% | 3 |
| No | 97.35% | 110 |
| Total | 100% | 113 |

### Q670 - Were you able to talk with customer support?

**Table 98.** Stats for Q670: “Were you able to talk with [BinaxNow] customer support?”

| Field | Minimum | Maximum | Mean | Std Deviation | Variance | Count |
| --- | --- | --- | --- | --- | --- | --- |
| Were you able to talk with customer support? | 1.00 | 1.00 | 1.00 | 0.00 | 0.00 | 3 |

**Table 99.**
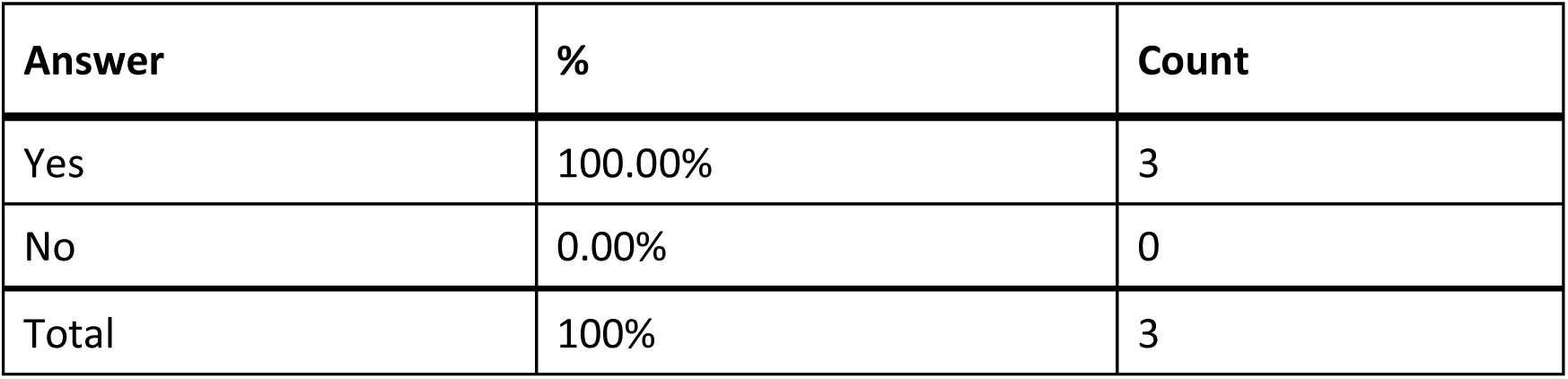
Responses to Q670: “Were you able to talk with [BinaxNow] customer support?”

### Q671 - Was customer support helpful in answering your question(s)?

**Table 100.**
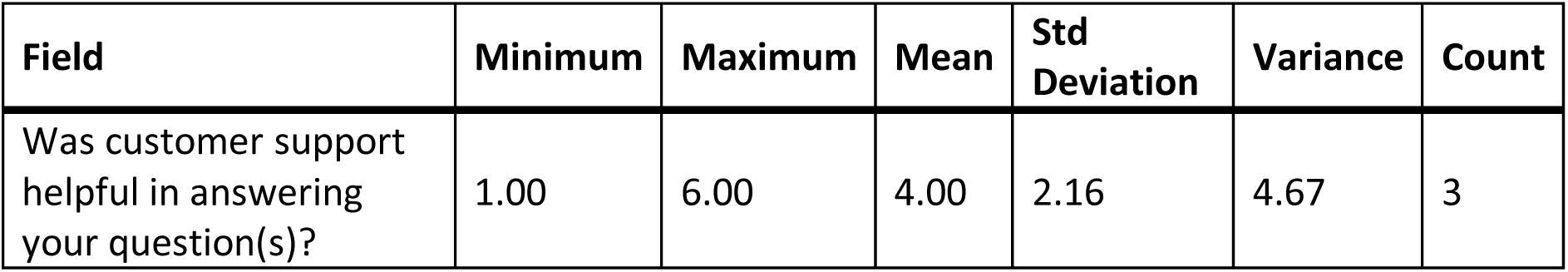
Stats for Q671: “Was [BinaxNow] customer support helpful in answering your question(s)?”

| Field | Minimum | Maximum | Mean | Std Deviation | Variance | Count |
| --- | --- | --- | --- | --- | --- | --- |
| Was customer support helpful in answering your question(s)? | 1.00 | 6.00 | 4.00 | 2.16 | 4.67 | 3 |

**Table 101.**
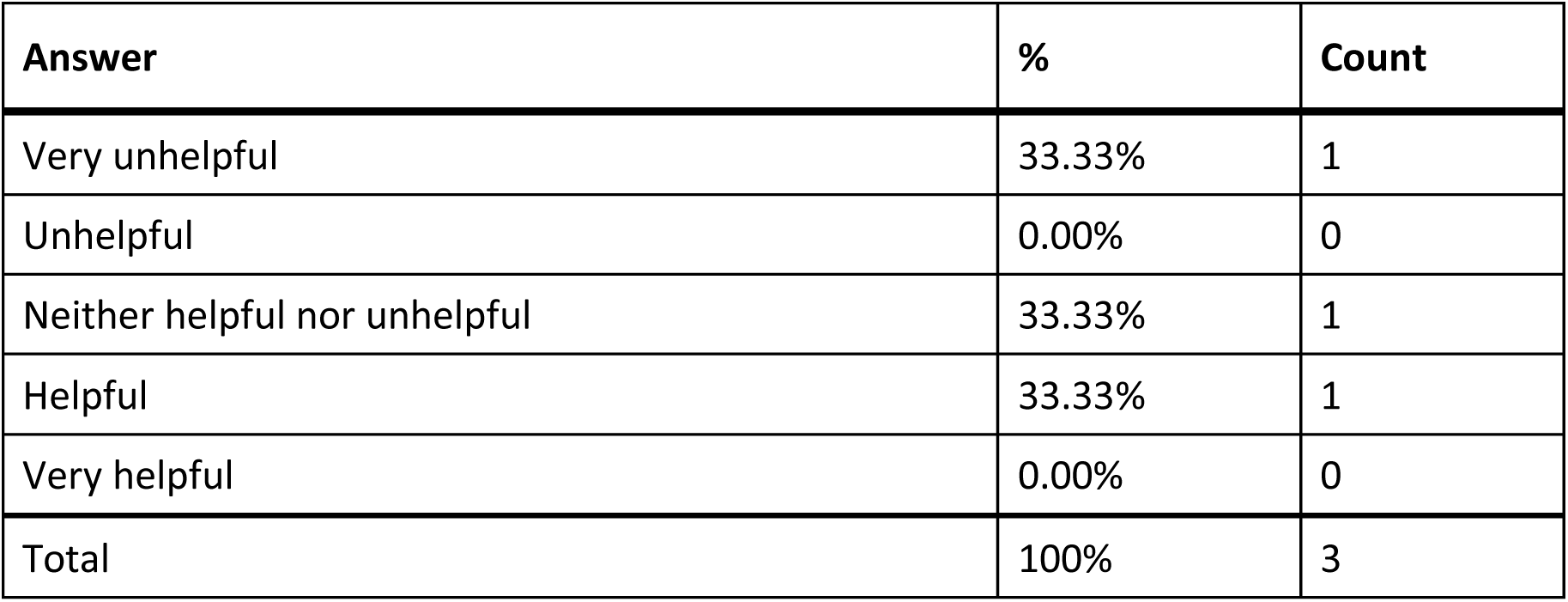
Responses to Q671: “Was [BinaxNow] customer support helpful in answering your question(s)?”

### Q672 - If you have performed the BinaxNow test multiple times, how did your experience change?

**Table 102.** Stats for Q672: “If you have performed the BinaxNow test multiple times, how did your experience change?”

| Field | Minimum | Maximum | Mean | Std Deviation | Variance | Count |
| --- | --- | --- | --- | --- | --- | --- |
| If you have performed the BinaxNow test multiple times, how did your experience change? | 1.00 | 5.00 | 3.41 | 0.75 | 0.56 | 75 |

**Table 103.** Responses to Q672: “If you have performed the BinaxNow test multiple times, how did your experience change?”

| Answer | % | Count |
| --- | --- | --- |
| Much worse | 1.33% | 1 |
| Worse | 1.33% | 1 |
| Neither better nor worse | 62.67% | 47 |
| Better | 24.00% | 18 |
| Much better | 10.67% | 8 |
| Total | 100% | 75 |

### Q673 - What did you like about the BinaxNow test?

**Table 104.**
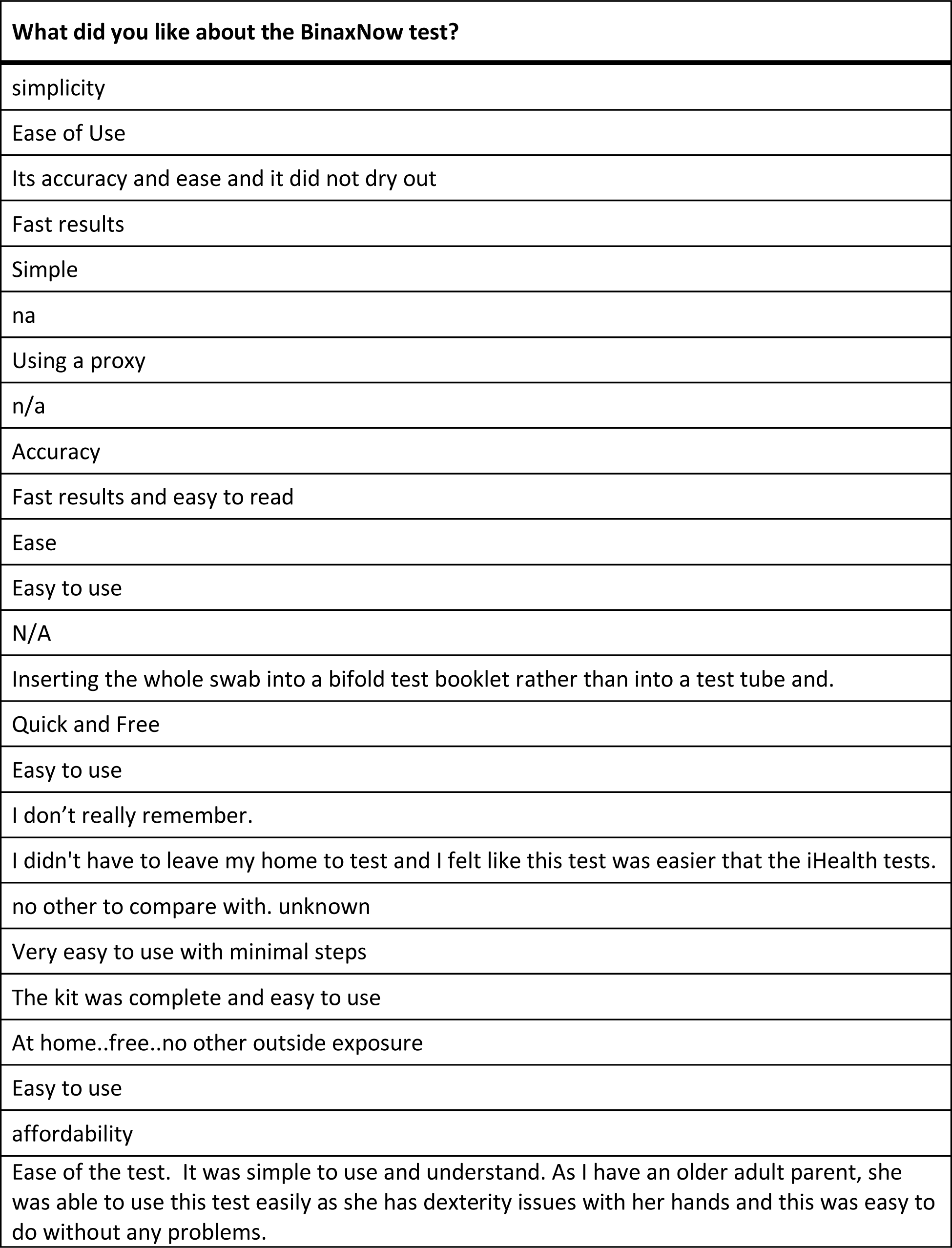
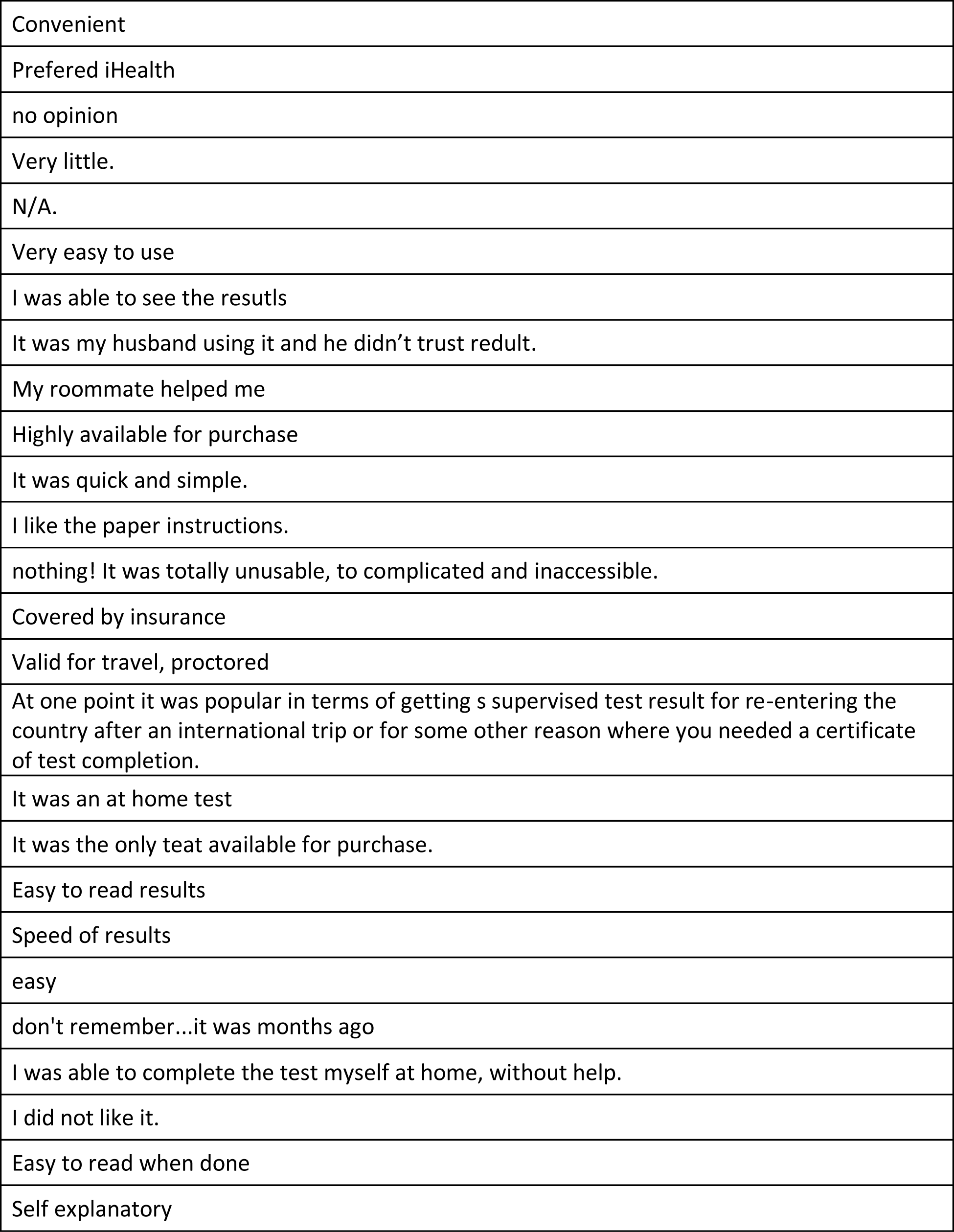
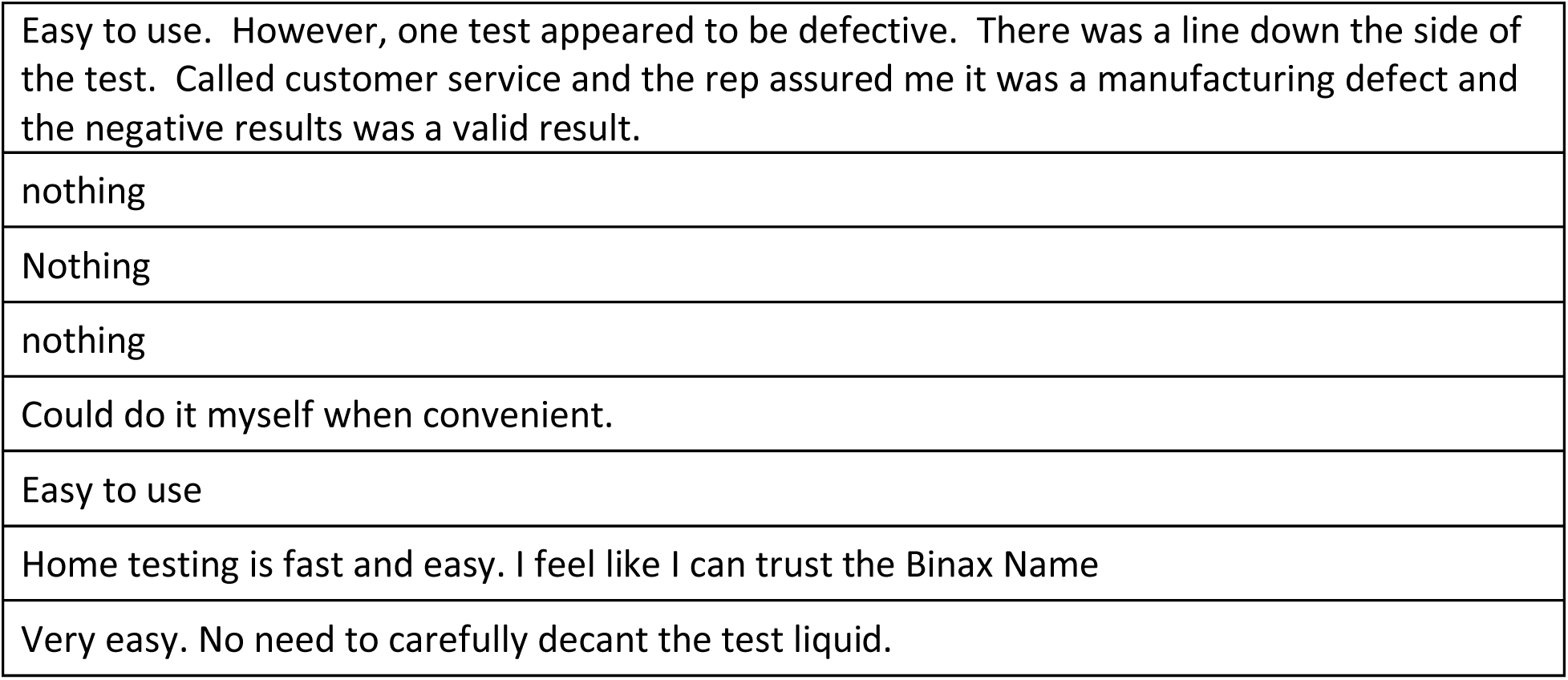
Open response to Q673: “What did you like about the BinaxNow test?”

### Q674 - What would you like to see improved for the BinaxNow test?

**Table 105.** Open responses to Q674: “What would you like to see improved for the BinaxNow test?”

| **What would you like to see improved for the BinaxNow test?** |
| --- |
| I prefer the other tests with the dropper feature instead of pushing the swab in the card hole. |
| DOH reporting / several were positive but didn't report results |
| Instructions should be user friendly for individuals with various disabilities |
| lower price |
| Bigger font or magnifying |
| Wings on the top of the reagent container for easier twist off. |
| na |
| less precision needed to collect test sample and put sample on testing strip |
| n/a |
| Less steps to complete |
| Almost every step hurt my hands |
| Nothing |
| N/A |
| Provide accessible instructions and a way to receive results. |
| A more rapid test would be nice, and easier to understand/use instructions, components |
| I just handed it to someone else to do most of it. The only part I handled was the swab. |
| An app that makes it easy for those who are blind or low vision |
| Nothing |
| Instructions need to be a little easier to understand and the font needs to be bigger. |
| The color of the test result strips. A large number of people can't or have trouble seeing red or green. |
| If I am thinking of the right one, apparently there were two ways that the test could have come. The directions were for both ways, so I had to figure out which one before I moved forward with the directions |
| better illustration |
| Instructions were compls...lots of warnings of failure if the user was not exact in each stem. Worried about a false indication. |
| Perhaps more eco-friendly materials to reduce waste |
| Easier to use, and read |
| Reliability- two test sets gave no results |
| the test protocol is a bit complex, mainly the insertion of the test swab into the hold properly |
| This could be difficult for someone with mobility/dexterity limitations- the small vials of fluid require removal of a seal that could be challenging for someone with limited feeling in their hands or arthritis. |
| no opinion |
| Packaging easier to open; accurate way to know how many drops of liquid I have put into the hole |
| Gett an app. |
| N/a |
| More detailed instructions, including a description of the various parts. Also, the ability to independently measure the solution and interpret the test results. |
| instructions need to be in larger print. Parts should be labelled better. Anb app with audio instructions would be helpful. |
| Nothing. |
| Reduce price |
| Easier to interpret- positive or negative not just a line that could be very faint and hard to see |
| I think instead of self administering the drops, there should be some type of internal capsule that can be punctured by applying pressure, that will release the proper amount of drops directly into the strip. It would leave little human error in measuring for those who have difficulty remembering how many drops entered or visually impaired who may have difficulty seeing where to apply the drops, or even those who suffer mobile control such as shaking that would cause overage in the amount of drops onto the strip. |
| I wish the app was easier to use/see. |
| Everything, otherwise take it off the market! |
| That card was REALLY hard to use and it was hard to get the drops in the right spot. |
| the process and result to be accessible. |
| Why can't the dropper just have the right amount of fluid?? (Ok evaporation, I guess it makes sense but really counting drops is hard nonvisually) |
| Use descriptive words as test results instead of a line segment or two. For example, if the test was positive for covid, then the test result reads "positive." Similarly, a negative result would show up as "netative." This way, screen reading software & apps could give audible feedback. Note that the screen contrast must be very high (like black text on white background) for screen-reading tools to be able to read the text. |
| Info passed between proctors of condition |
| Able to nonvisually read documentation, results, better design and description of test components. For instance, which hole do you insert the swab in, there are 2 holes on the card. |
| The use of the word positive or negative rather than a test line. |
| Instructions were not exactly clear on where and how to place the swab on the test pad. |
| Larger font on instructions |
| none |
| Everything |
| No suggestions. |
| same as above, dropping seven drops of liquid into a tiny hole, really? The test should have the necessary amount of liquid in a container into which the swab is placed, very poor design! |
| Clearer instructions on which way to fold over the card sensor to seal it. |
| Nothing is recyclable. The instructions were too small. I have trouble reading small print. I have Parkinson's and fine motor coordination is difficult. |
| N/A |
| The dropper with the reagent could be larger. |
| The person I spoke to could not help me. |
| Preparing the test card, sample processing, and how results are displayed. |
| made completely accessible to blind users |
| Easier to follow instructions and examples of test results. |
| Not much to improve upon. |
| Make everything larger. The vials were tiny. In fact,I did drop the first one. |
| More access to affordable testing. |

### Q675 - QuickVue Think of the first time you used the QuickVue test. Did you have difficulty completing any of the following tasks? Select all that apply

**Table 106.** Responses to Q675: “QuickVue - Did you have difficulty completing any of the following tasks?”

| Answer | % | Count |
| --- | --- | --- |
| Opening the packaging | 8.54% | 7 |
| Reading the instructions | 20.73% | 17 |
| Identifying test components | 12.20% | 10 |
| Collecting the test sample | 6.10% | 5 |
| Manipulating test components | 15.85% | 13 |
| Interpreting the test results | 17.07% | 14 |
| Other | 3.66% | 3 |
| I did not have difficulty completing the test | 15.85% | 13 |
| Total | 100% | 82 |

#### Q675_7_TEXT – Other

**Table 107.** Open responses to Other - Q675: “QuickVue - Did you have difficulty completing any of the following tasks?”

| Other - Text |
| --- |
| The instructions were written in Spanish!! |
| Requested assistance from a family member |
| seeing the test results |

### Q676 - What tools or assistance, if any, did you use to perform the test?

**Table 108.** Responses to Q676: “What tools or assistance, if any, did you use to perform the [QuickVue] test?”

| Answer | % | Count |
| --- | --- | --- |
| Timer | 25.40% | 16 |
| Scissors | 15.87% | 10 |
| Remote visual assistance (AIRA, Be My Eyes, Facetime, Video-chat, etc.) | 14.29% | 9 |
| Screen Reader | 6.35% | 4 |
| Magnification | 11.11% | 7 |
| Other technology (OCR, Bar Code Readers, Light Detectors, etc.) | 3.17% | 2 |
| In person human assistance | 14.29% | 9 |
| Other | 1.59% | 1 |
| I did not need any tools or assistance | 7.94% | 5 |
| Total | 100% | 63 |

#### Q676_8_TEXT – Other

**Table 109.** Open response to Other - Q676: “What tools or assistance, if any, did you use to perform the [QuickVue] test?”

| Other - Text |
| --- |
| timer on Amazon echo device |

### Q677 - Were you able to conduct this test on your first try?

**Table 110.** Stats for Q677: “Were you able to conduct this[QuickVue] test on your first try?”

| Field | Minimum | Maximum | Mean | Std Deviation | Variance | Count |
| --- | --- | --- | --- | --- | --- | --- |
| Were you able to conduct this test on your first try? | 1.00 | 2.00 | 1.21 | 0.40 | 0.16 | 39 |

**Table 111.** Response to Q677: “Were you able to conduct this [QuickVue] test on your first try?”

| Answer | % | Count |
| --- | --- | --- |
| Yes | 79.49% | 31 |
| No | 20.51% | 8 |
| Total | 100% | 39 |

### Q678 - How long did it take to complete the steps required to perform the test, not including the time you spent waiting for results

**Table 112.**
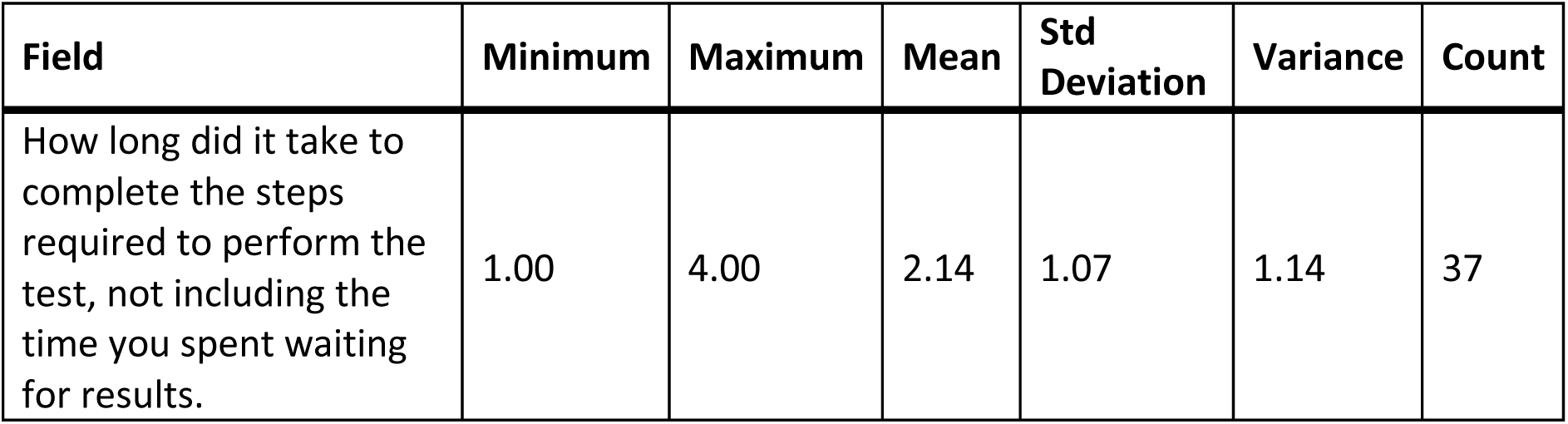
Stats for Q678: “How long did it take to complete the steps required to perform the [QuickVue] test?”

| Field | Minimum | Maximum | Mean | Std Deviation | Variance | Count |
| --- | --- | --- | --- | --- | --- | --- |
| How long did it take to complete the steps required to perform the test, not including the time you spent waiting for results. | 1.00 | 4.00 | 2.14 | 1.07 | 1.14 | 37 |

**Table 113.** Response to Q678: “How long did it take to complete the steps required to perform the [QuickVue] test?”

| Answer | % | Count |
| --- | --- | --- |
| Under 5 minutes | 32.43% | 12 |
| 5 - 10 minutes | 40.54% | 15 |
| 10 - 15 minutes | 8.11% | 3 |
| Over 15 minutes | 18.92% | 7 |
| Total | 100% | 37 |

### Q679 - What format of instructions did you use?

**Table 114.** Responses to Q679: “What format of [QuickVue] instructions did you use?”

| Answer | % | Count |
| --- | --- | --- |
| Paper | 65.91% | 29 |
| Digital (webpages, downloaded, etc.) | 15.91% | 7 |
| None of the above | 11.36% | 5 |
| Associated Covid Test App (Smartphone) | 6.82% | 3 |
| Total | 100% | 44 |

### Q680 - Were you able to access electronic information via a QR Code?

**Table 115.** Q680: “Were you able to access electronic information via QR Code?”

| Field | Minimum | Maximum | Mean | Std Deviation | Variance | Count |
| --- | --- | --- | --- | --- | --- | --- |
| Were you able to access electronic information via a QR Code? | 1.00 | 7.00 | 4.75 | 2.90 | 8.44 | 8 |

**Table 116.**
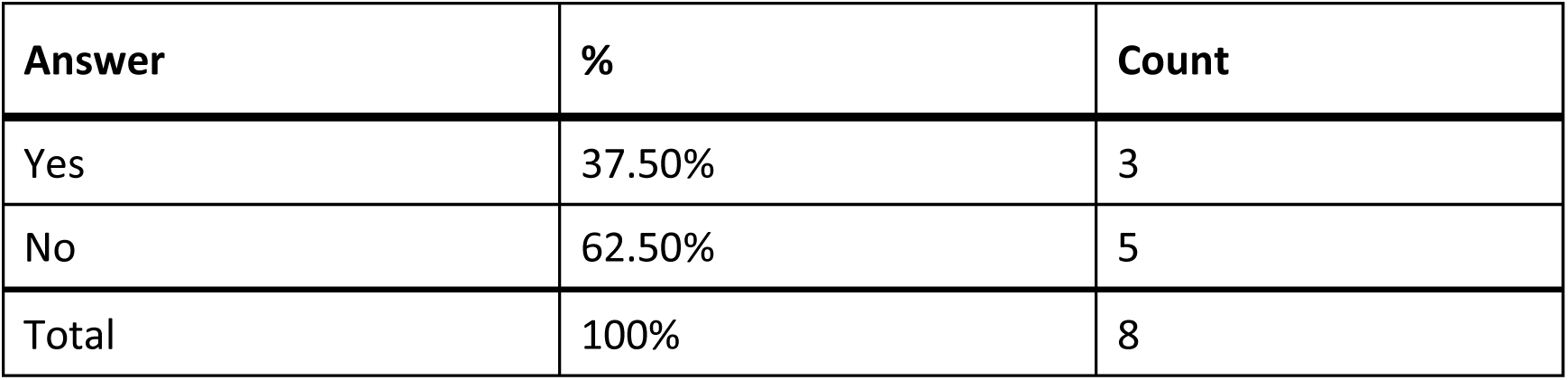
Responses to Q680: “Were you able to access electronic information via [QuickVue] QR Code?”

| Answer | % | Count |
| --- | --- | --- |
| Yes | 37.50% | 3 |
| No | 62.50% | 5 |
| Total | 100% | 8 |

### Q681 - How easy was opening the package of the QuickVue test for you? (without assistance)

**Table 117.** Stats for Q681: “How easy was opening the package of the QuickVue test for you? (without assistance)”

| Field | Minimum | Maximum | Mean | Std Deviation | Variance | Count |
| --- | --- | --- | --- | --- | --- | --- |
| How easy was opening the package of the QuickVue test for you? (without assistance) | 1.00 | 5.00 | 3.41 | 1.08 | 1.16 | 37 |

**Table 118.** Responses to Q681: “How easy was opening the package of the QuickVue test for you? (without assistance)”

| Answer | % | Count |
| --- | --- | --- |
| Very difficult | 5.41% | 2 |
| Difficult | 10.81% | 4 |
| Neither easy nor difficult | 40.54% | 15 |
| Easy | 24.32% | 9 |
| Very easy | 18.92% | 7 |
| Total | 100% | 37 |

### Q682 - How easy was completing the test protocol for you? (without assistance)

**Table 119.**
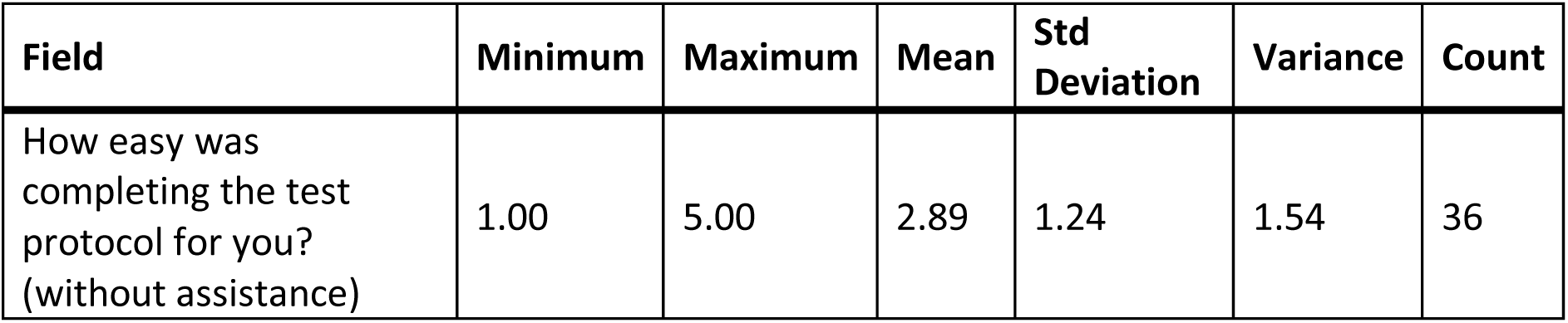
Stats for Q682: “How easy was completing the [QuickVue] test protocol for you? (without assistance)”

| Field | Minimum | Maximum | Mean | Std Deviation | Variance | Count |
| --- | --- | --- | --- | --- | --- | --- |
| How easy was completing the test protocol for you? (without assistance) | 1.00 | 5.00 | 2.89 | 1.24 | 1.54 | 36 |

**Table 120.** Responses to Q682: “How easy was completing the [QuickVue] test protocol for you? (without assistance)”

| Answer | % | Count |
| --- | --- | --- |
| Very difficult | 19.44% | 7 |
| Difficult | 13.89% | 5 |
| Neither easy nor difficult | 36.11% | 13 |
| Easy | 19.44% | 7 |
| Very easy | 11.11% | 4 |
| Total | 100% | 36 |

### Q683 - How easy was interpreting the test results for you? (without assistance)

**Table 121.** Stats for Q683: “How easy was interpreting the [QuickVue] test results for you? (without assistance)”

| Field | Minimum | Maximum | Mean | Std Deviation | Variance | Count |
| --- | --- | --- | --- | --- | --- | --- |
| How easy was interpreting the test results for you? (without assistance) | 1.00 | 5.00 | 2.88 | 1.47 | 2.16 | 34 |

**Table 122.** Responses to Q683: “How easy was interpreting the [QuickVue] test results for you? (without assistance)”

| Answer | % | Count |
| --- | --- | --- |
| Very difficult | 26.47% | 9 |
| Difficult | 14.71% | 5 |
| Neither easy nor difficult | 23.53% | 8 |
| Easy | 14.71% | 5 |
| Very easy | 20.59% | 7 |
| Total | 100% | 34 |

### Q684 - Were you able to interpret the test results? (without assistance)

**Table 123.** Stats for Q684: “Were you able to interpret the [QuickVue] test results? (without assistance)”

| Field | Minimum | Maximum | Mean | Std Deviation | Variance | Count |
| --- | --- | --- | --- | --- | --- | --- |
| Were you able to interpret the test results? (without assistance) | 1.00 | 2.00 | 1.34 | 0.47 | 0.23 | 35 |

**Table 124.** Responses to Q684: “Were you able to interpret the [QuickVue] test results? (without assistance)”

| Answer | % | Count |
| --- | --- | --- |
| Yes | 65.71% | 23 |
| No | 34.29% | 12 |
| Total | 100% | 35 |

### Q685 - If there was an associated app for the test, how easy was it for you to use? (without assistance)

**Table 125.**
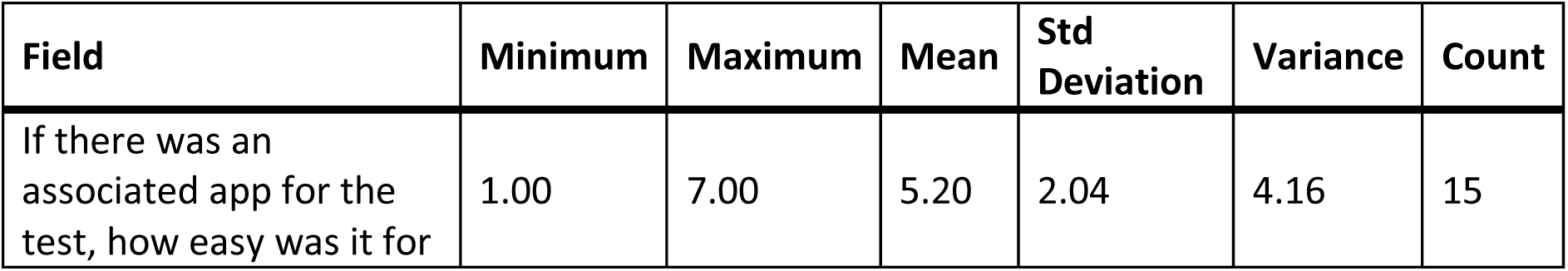
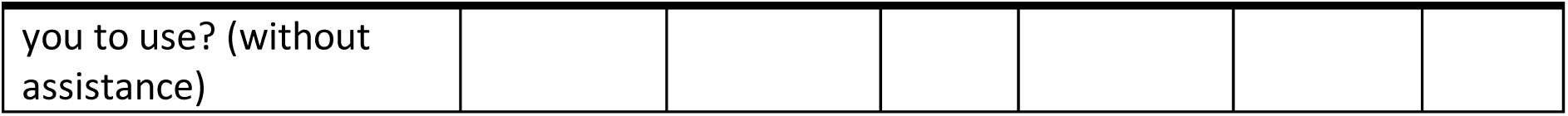
Stats for Q685: “If there was an associated app for the [QuickVue] test, how easy was it for you to use? (without assistance)”

**Table 126.** Responses to Q685: “If there was an associated app for the [QuickVue] test, how easy was it for you to use? (without assistance)”

| Answer | % | Count |
| --- | --- | --- |
| Very difficult | 6.67% | 1 |
| Difficult | 0.00% | 0 |
| Neither easy nor difficult | 20.00% | 3 |
| Easy | 20.00% | 3 |
| Very easy | 0.00% | 0 |
| There is an app, but I didn't use it | 53.33% | 8 |
| Total | 100% | 15 |

### Q686 - How helpful was the app in assisting you with completing the test?

**Table 127.** Stats for Q686: “How helpful was the app in assisting you with completing the [QuickVue] test?”

| Field | Minimum | Maximum | Mean | Std Deviation | Variance | Count |
| --- | --- | --- | --- | --- | --- | --- |
| How helpful was the app in assisting you with completing the test? | 1.00 | 4.00 | 3.00 | 1.41 | 2.00 | 6 |

**Table 128:** Responses to Q686: “How helpful was the app in assisting you with completing the [QuickVue] test?”

| Answer | % | Count |
| --- | --- | --- |
| Very unhelpful | 33.33% | 2 |
| Unhelpful | 0.00% | 0 |
| Neither unhelpful nor helpful | 0.00% | 0 |
| Helpful | 66.67% | 4 |
| Very helpful | 0.00% | 0 |
| Total | 100% | 6 |

### Q687 - Did you receive a valid test result with the QuickVue test (positive or negative)?

**Table 129.**
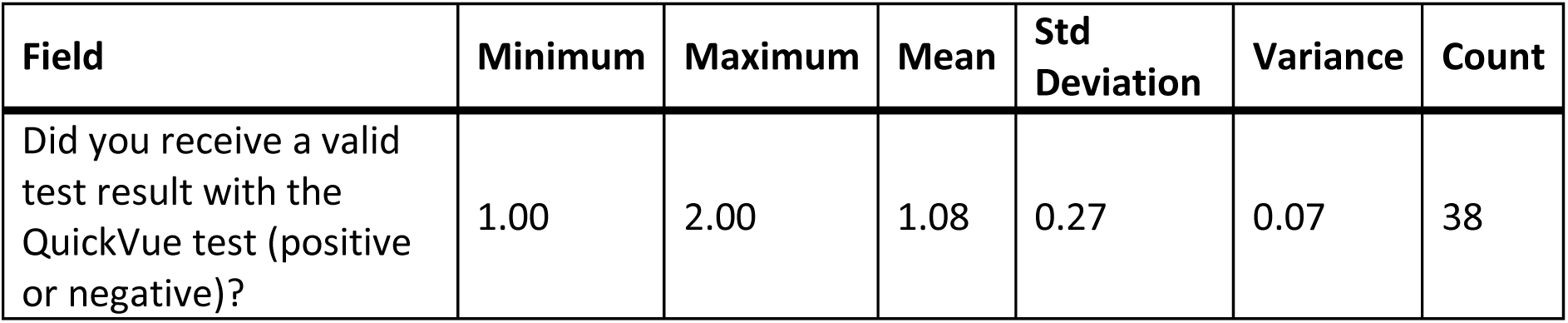
Stats for Q:687: “Did you receive a valid test result with the QuickVue test (positive or negative)?”

**Table 130.** Response to Q687: “Did you receive a valid test result with the QuickVue test (positive or negative)?”

| Answer | % | Count |
| --- | --- | --- |
| Yes | 92.11% | 35 |
| No | 7.89% | 3 |
| Total | 100% | 38 |

### Q688 - Did you attempt to contact QuickVue’s customer support?

**Table 131.** Stats forQ688: “Did you attempt to contact QuickVue’s customer support?”

| Field | Minimum | Maximum | Mean | Std Deviation | Variance | Count |
| --- | --- | --- | --- | --- | --- | --- |
| Did you attempt to contact QuickVue's customer support? | 1.00 | 2.00 | 1.92 | 0.27 | 0.07 | 37 |

**Table 132.** Response to Q688: “Did you attempt to contact QuickVue’s customer support?”

| Answer | % | Count |
| --- | --- | --- |
| Yes | 8.11% | 3 |
| No | 91.89% | 34 |
| Total | 100% | 37 |

### Q689 - Were you able to talk with customer support?

**Table 133.** Stats for Q689: “Were you able to talk with [QuickVue] customer support?”

| Field | Minimum | Maximum | Mean | Std Deviation | Variance | Count |
| --- | --- | --- | --- | --- | --- | --- |
| Were you able to talk with customer support? | 1.00 | 2.00 | 1.67 | 0.47 | 0.22 | 3 |

**Table 134.** Response to Q689: “Were you able to talk with [QuickVue] customer support?”

| Answer | % | Count |
| --- | --- | --- |
| Yes | 33.33% | 1 |
| No | 66.67% | 2 |
| Total | 100% | 3 |

### Q690 - Was customer support helpful in answering your question(s)?

**Table 135.** Stats for Q690: “Was [QuickVue] customer support helpful in answering your question(s)?”

| Field | Minimum | Maximum | Mean | Std Deviation | Variance | Count |
| --- | --- | --- | --- | --- | --- | --- |
| Was customer support helpful in answering your question(s)? | 1.00 | 1.00 | 1.00 | 0.00 | 0.00 | 1 |

**Table 136.** Responses to Q690: “Was [QuickVue] customer support helpful in answering your question(s)?”

| Answer | % | Count |
| --- | --- | --- |
| Very unhelpful | 100.00% | 1 |
| Unhelpful | 0.00% | 0 |
| Neither helpful nor unhelpful | 0.00% | 0 |
| Helpful | 0.00% | 0 |
| Very helpful | 0.00% | 0 |
| Total | 100% | 1 |

### Q691 - If you have performed the QuickVue test multiple times, how did your experience change?

**Table 137.** Stats for Q691: “If you have performed the QuickVue test multiple times, how did your experience change?”

| Field | Minimum | Maximum | Mean | Std Deviation | Variance | Count |
| --- | --- | --- | --- | --- | --- | --- |
| If you have performed the QuickVue test multiple times, how did your experience change? | 1.00 | 4.00 | 3.14 | 0.76 | 0.57 | 22 |

**Table 138.** Responses to Q691: “If you have performed the QuickVue test multiple times, how did your experience change?”

| Answer | % | Count |
| --- | --- | --- |
| Much worse | 4.55% | 1 |
| Worse | 9.09% | 2 |
| Neither better nor worse | 54.55% | 12 |
| Better | 31.82% | 7 |
| Much better | 0.00% | 0 |
| Total | 100% | 22 |

### Q692 - What did you like about the QuickVue test?

**Table 139.** Open response to Q692: “What did you like about the QuickVue test?”

| What did you like about the QuickVue test? |
| --- |
| easy for client to read |
| It was quick |
| all the good test test test stuffff |
| N/A |
| N/A |
| it was easy to find |
| It was provided at no charge |
| Easy to use |
| I loved the ease in use of the test. As a person who had an older parent with dexterity issues, this test was easy to use without needing assistance. |
| Nothing |
| it is inaccessible so I don't like anything about it |
| I didn't have to count drops or transfer any liquid. I like that it does not require a smart phone. The test is easy to perform and accurate. |
| I could perform most of the test procedure myself. |
| Nothing. I wouldn't buy it again. |
| It was free. |
| Detailed instructions and easy to perform and interpret |
| Simple |
| I liked that I didn't have to stick the swab way up my nose. |
| Liked having to drop reagent in a tube rather than into a small hole. |
| Easy to swab and put swab in tube |

### Q693 - What would you like to see improved for the QuickVue test?

**Table 140.** Open responses to Q693: “What would you like to see improved for the QuickVue test?”

| What would you like to see improved for the QuickVue test? |
| --- |
| cost |
| Less pieces and needed stands for both tests in one OTC kit...it only comes with one reagent stand, but we want 2 people to use them at the same time while being physically distanced. |
| Need to have one side of instructions in English |
| test test test yay |
| Every step was really painful for my hands |
| N/A |
| N/A |
| n/a |
| Easier packaging, more clear/obvious test results |
| Maybe a quick-start guide with pictures that quickly and easily guides you through the process |
| Easier to interpret results. Clearly positive or negative. Hard to see faint lines. |
| A ap |
| obviously, make it accessible |
| I'm not sure if it is possible or not but an audible announcement of whether the test is positive or negative would be nice, but I don't want it to require my smart phone. Many blind people cannot afford or learn to use a smart phone. |
| You're really asking this? Okay, I'll play the game. I would really like to not have to coordinate with somebody else to test myself, specifically find somebody to read the results. |
| Accessible online instructions and another way that does not involve vision to interpret the results |
| Everything regarding reading instructions, accessible labeling, and accessible test results. All of it. |
| N/A |
| It's very difficult to know when drops come out of the dropper...it's easier to count drops using an eye dropper with the soft plastic that you can just squeeze and feel it better. Also, screwing the liid on the collection is a bit cumbersome while juggling the dirty swab that you're supposed to put back in its wrapper. It is difficult to identify the correct end of the swab without touching it and it would be helpful to somehow clearly mark the outside of the swab package to open. |
| Nothing |
| Impossible to count drops and read result. |

### Q694 - CareStart Think of the first time you used the CareStart test. Did you have difficulty completing any of the following tasks? Select all that apply

**Table 141.** Responses to Q694: CareStart - Did you have any difficulty completing any of the following tasks?”

| Answer | % | Count |
| --- | --- | --- |
| Opening the packaging | 33.33% | 2 |
| Reading the instructions | 16.67% | 1 |
| Identifying test components | 0.00% | 0 |
| Collecting the test sample | 16.67% | 1 |
| Manipulating test components | 16.67% | 1 |
| Interpreting the test results | 0.00% | 0 |
| Other | 0.00% | 0 |
| I did not have difficulty completing the test | 16.67% | 1 |
| Total | 100% | 6 |

#### Q694_7_TEXT - Other

None

### Q695 - What tools or assistance, if any, did you use to perform the test?

**Table 142.** Responses to Q695: “What tools or assistance, if any, did you use to perform the [CareStart] test?”

| Answer | % | Count |
| --- | --- | --- |
| Timer | 25.00% | 2 |
| Scissors | 12.50% | 1 |
| Remote visual assistance (AIRA, Be My Eyes, Facetime, Video-chat, etc.) | 12.50% | 1 |
| Screen Reader | 0.00% | 0 |
| Magnification | 25.00% | 2 |
| Other technology (OCR, Bar Code Readers, Light Detectors, etc.) | 0.00% | 0 |
| In person human assistance | 12.50% | 1 |
| Other | 0.00% | 0 |
| I did not need any tools or assistance | 12.50% | 1 |
| Total | 100% | 8 |

#### Q695_8_TEXT - Other

None

### Q696 - Were you able to conduct this test on your first try?

**Table 143.** Stats for Q696: “Were you able to conduct this [CareStart] test on your first try?”

| Field | Minimum | Maximum | Mean | Std Deviation | Variance | Count |
| --- | --- | --- | --- | --- | --- | --- |
| Were you able to conduct this test on your first try? | 1.00 | 2.00 | 1.25 | 0.43 | 0.19 | 4 |

**Table 144.** Responses to Q696: “Were you able to conduct this [CareStart] test on your first try?”

| Answer | % | Count |
| --- | --- | --- |
| Yes | 75.00% | 3 |
| No | 25.00% | 1 |
| Total | 100% | 4 |

### Q697 - How long did it take to complete the steps required to perform the test, not including the time you spent waiting for results

**Table 145.** Stats for Q697: “How long did it take to complete the steps required to perform the [CareStart] test?”

| Field | Minimum | Maximum | Mean | Std Deviation | Variance | Count |
| --- | --- | --- | --- | --- | --- | --- |
| How long did it take to complete the steps required to perform the test, not including the time you spent waiting for results. | 1.00 | 2.00 | 1.67 | 0.47 | 0.22 | 3 |

**Table 146.** Responses to Q697: “How long did it take to complete the steps required to perform the [CareStart] test?”

| Answer | % | Count |
| --- | --- | --- |
| Under 5 minutes | 33.33% | 1 |
| 5 - 10 minutes | 66.67% | 2 |
| 10 - 15 minutes | 0.00% | 0 |
| Over 15 minutes | 0.00% | 0 |
| Total | 100% | 3 |

### Q698 - What format of instructions did you use?

**Table 147.** Responses to Q698: “What format of [CareStart] instructions did you use?”

| Answer | % | Count |
| --- | --- | --- |
| Paper | 50.00% | 2 |
| Digital (webpages, downloaded, etc.) | 25.00% | 1 |
| None of the above | 0.00% | 0 |
| Associated Covid Test App (Smartphone) | 25.00% | 1 |
| Total | 100% | 4 |

### Q699 - Were you able to access electronic information via a QR Code?

**Table 148.** Stats for Q699: “Were you able to access electronic information via a [CareStart] QR Code?”

| Field | Minimum | Maximum | Mean | Std Deviation | Variance | Count |
| --- | --- | --- | --- | --- | --- | --- |
| Were you able to access electronic information via a QR Code? | 1.00 | 1.00 | 1.00 | 0.00 | 0.00 | 2 |

**Table 149.** Responses to Q699: “Were you able to access electronic information via a [CareStart] QR Code?”

| Answer | % | Count |
| --- | --- | --- |
| Yes | 100.00% | 2 |
| No | 0.00% | 0 |
| Total | 100% | 2 |

### Q700 - How easy was opening the package of the CareStart test for you? (without assistance)

**Table 150.**
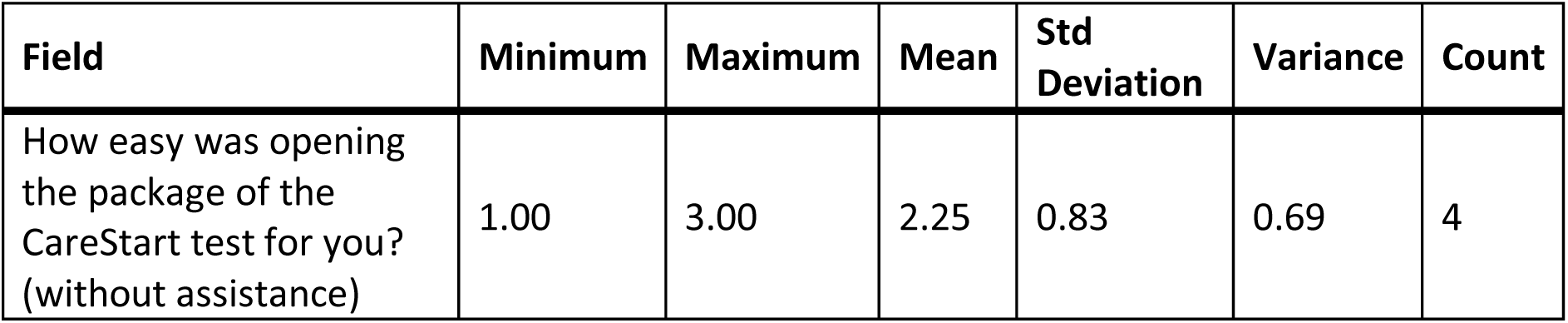
Stats for Q700: “How easy was opening the package of the CareStart Test for you? (without assistance)”

| Field | Minimum | Maximum | Mean | Std Deviation | Variance | Count |
| --- | --- | --- | --- | --- | --- | --- |
| How easy was opening the package of the CareStart test for you? (without assistance) | 1.00 | 3.00 | 2.25 | 0.83 | 0.69 | 4 |

**Table 151.** Responses to Q700: “How easy was opening the package of the CareStart Test for you? (without assistance)”

| Answer | % | Count |
| --- | --- | --- |
| Very difficult | 25.00% | 1 |
| Difficult | 25.00% | 1 |
| Neither easy nor difficult | 50.00% | 2 |
| Easy | 0.00% | 0 |
| Very easy | 0.00% | 0 |
| Total | 100% | 4 |

### Q701 - How easy was completing the test protocol for you? (without assistance)

**Table 152.** Stats for Q701: “How easy was completing the [CareStart] test protocol for you? (without assistance)”

| Field | Minimum | Maximum | Mean | Std Deviation | Variance | Count |
| --- | --- | --- | --- | --- | --- | --- |
| How easy was completing the test protocol for you? (without assistance) | 1.00 | 3.00 | 2.25 | 0.83 | 0.69 | 4 |

**Table 153.**
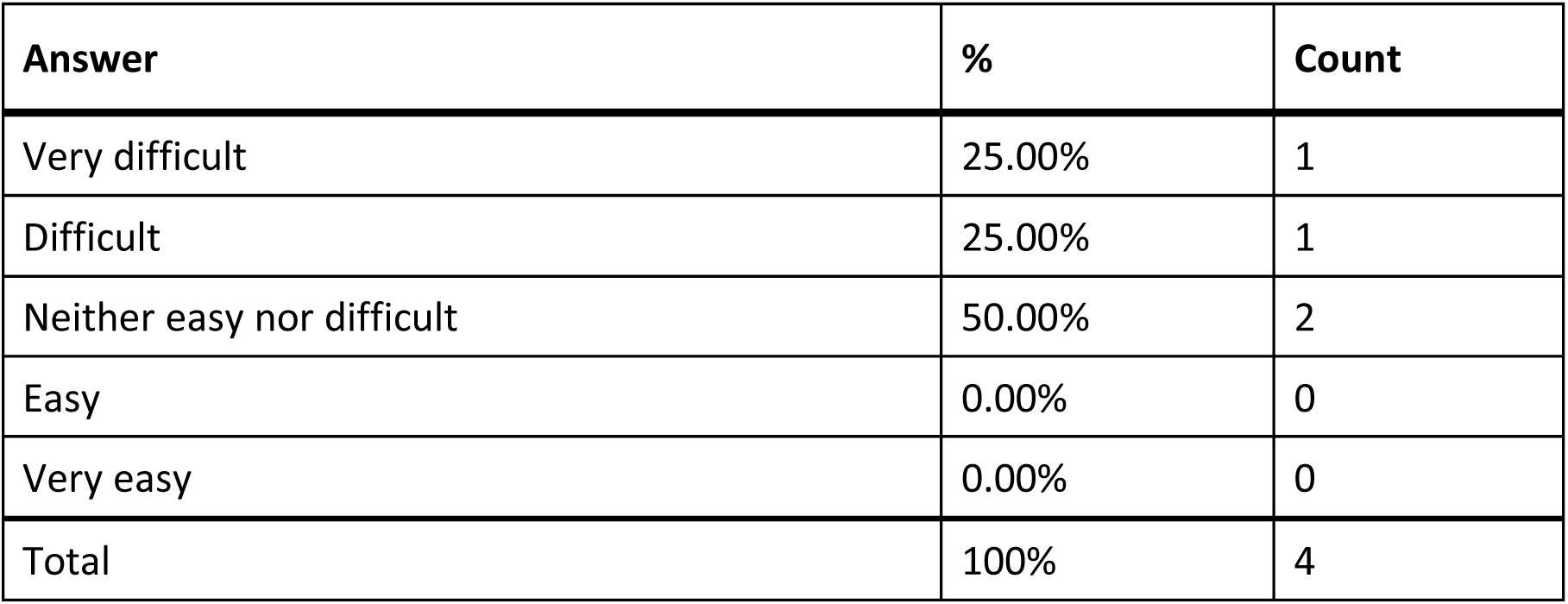
Responses to Q701: “How easy was completing the [CareStart] test protocol for you? (without assistance)”

### Q702 - How easy was interpreting the test results for you? (without assistance)

**Table 154.**
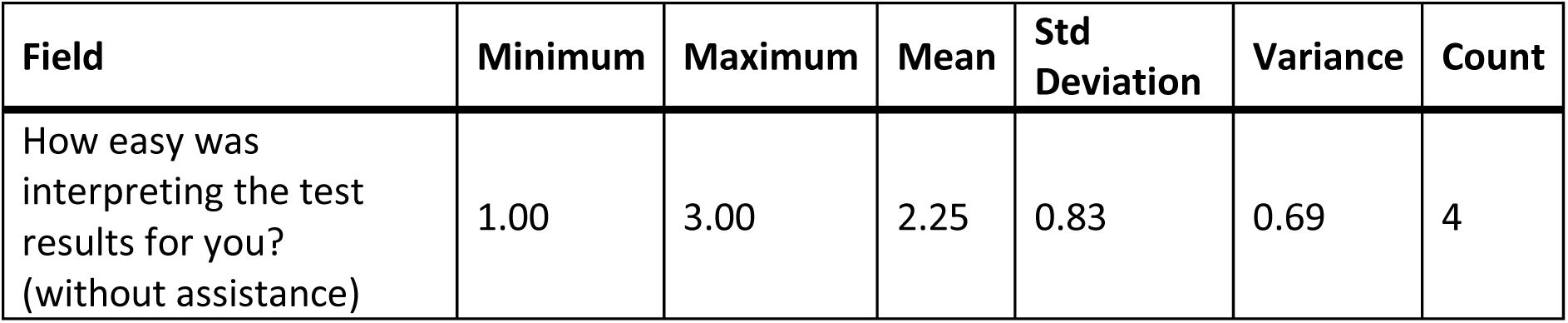
Stats for Q702: “How easy was interpreting the [CareStart] test results for you? (without assistance)”

**Table 155.**
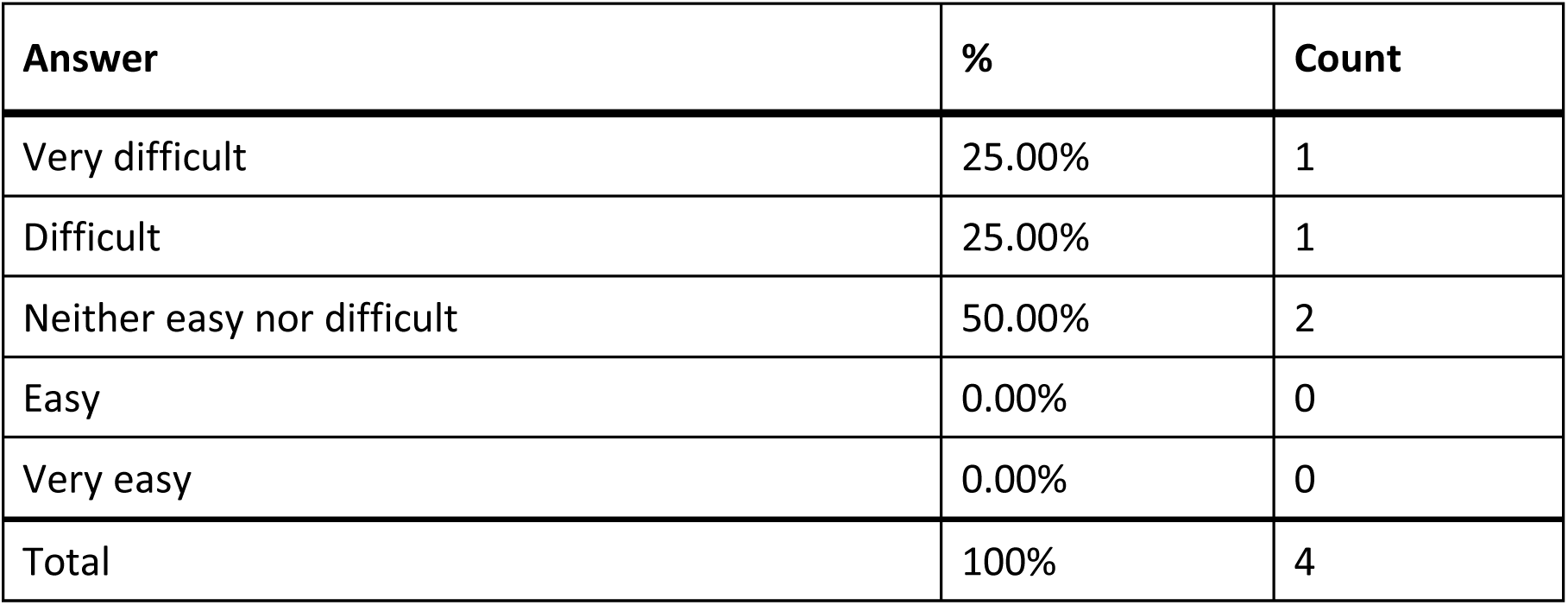
Responses to Q702: “How easy was interpreting the [CareStart] test results for you? (without assistance)”

### Q703 - Were you able to interpret the test results? (without assistance)

**Table 156.** Stats for Q704: “If there was an associated app for the [CareStart] test, how easy was it for you to use? (without assistance)”

| Field | Minimum | Maximum | Mean | Std Deviation | Variance | Count |
| --- | --- | --- | --- | --- | --- | --- |
| Were you able to interpret the test results? (without assistance) | 1.00 | 1.00 | 1.00 | 0.00 | 0.00 | 4 |

**Table 157.** Responses to Q704: “If there was an associated app for the [CareStart] test, how easy was it for you to use? (without assistance)”

| Answer | % | Count |
| --- | --- | --- |
| Very difficult | 33.33% | 1 |
| Difficult | 0.00% | 0 |
| Neither easy nor difficult | 0.00% | 0 |
| Easy | 66.67% | 2 |
| Very easy | 0.00% | 0 |
| There is an app, but I didn't use it | 0.00% | 0 |
| Total | 100% | 3 |

### Q705 - How helpful was the app in assisting you with completing the test?

**Table 158.** Stats for Q705: “How helpful was the [CareStart] app in assisting you with completing the test?”

| Field | Minimum | Maximum | Mean | Std Deviation | Variance | Count |
| --- | --- | --- | --- | --- | --- | --- |
| How helpful was the app in assisting you with completing the test? | 1.00 | 4.00 | 3.00 | 1.41 | 2.00 | 3 |

**Table 159.** Responses to Q705: “How helpful was the [CareStart] app in assisting you with completing the test?”

| Answer | % | Count |
| --- | --- | --- |
| Very unhelpful | 33.33% | 1 |
| Unhelpful | 0.00% | 0 |
| Neither unhelpful nor helpful | 0.00% | 0 |
| Helpful | 66.67% | 2 |
| Very helpful | 0.00% | 0 |
| Total | 100% | 3 |

### Q706 - Did you receive a valid test result with the CareStart test (positive or negative)?

**Table 160.** Stats for Q706: “Did you receive a valid test result with the CareStart test (positive or negative)?”

| Field | Minimum | Maximum | Mean | Std Deviation | Variance | Count |
| --- | --- | --- | --- | --- | --- | --- |
| Did you receive a valid test result with the CareStart test (positive or negative)? | 1.00 | 1.00 | 1.00 | 0.00 | 0.00 | 5 |

**Table 161.** Responses to Q706: “Did you receive a valid test result with the CareStart test (positive or negative)?”

| Answer | % | Count |
| --- | --- | --- |
| Yes | 100.00% | 5 |
| No | 0.00% | 0 |
| Total | 100% | 5 |

### Q707 - Did you attempt to contact CarerStart’s customer support?

**Table 162.**
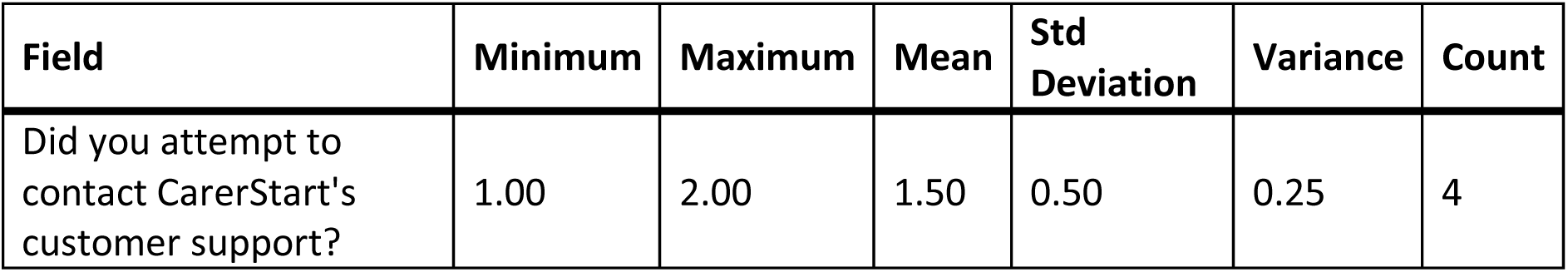
Stats for Q707: “Did you attempt to contact CareStart’s customer support?”

**Table 163.** Responses to Q707: “Did you attempt to contact CareStart’s customer support?”

| Answer | % | Count |
| --- | --- | --- |
| Yes | 50.00% | 2 |
| No | 50.00% | 2 |
| Total | 100% | 4 |

### Q708 - Were you able to talk with customer support?

**Table 164.** Stats for Q708: “Were you able to talk with [CareStart] customer support?”

| Field | Minimum | Maximum | Mean | Std Deviation | Variance | Count |
| --- | --- | --- | --- | --- | --- | --- |
| Were you able to talk with customer support? | 1.00 | 2.00 | 1.50 | 0.50 | 0.25 | 2 |

**Table 165.** Responses to Q708: “Were you able to talk with [CareStart] customer support?”

| Answer | % | Count |
| --- | --- | --- |
| Yes | 50.00% | 1 |
| No | 50.00% | 1 |
| Total | 100% | 2 |

### Q709 - Was customer support helpful in answering your question(s)?

**Table 166.** Stats for Q709: “Was [CareStart] customer support helpful in answering your question(s)?”

| Field | Minimum | Maximum | Mean | Std Deviation | Variance | Count |
| --- | --- | --- | --- | --- | --- | --- |
| Was customer support helpful in answering your question(s)? | 5.00 | 5.00 | 5.00 | 0.00 | 0.00 | 1 |

**Table 167.**
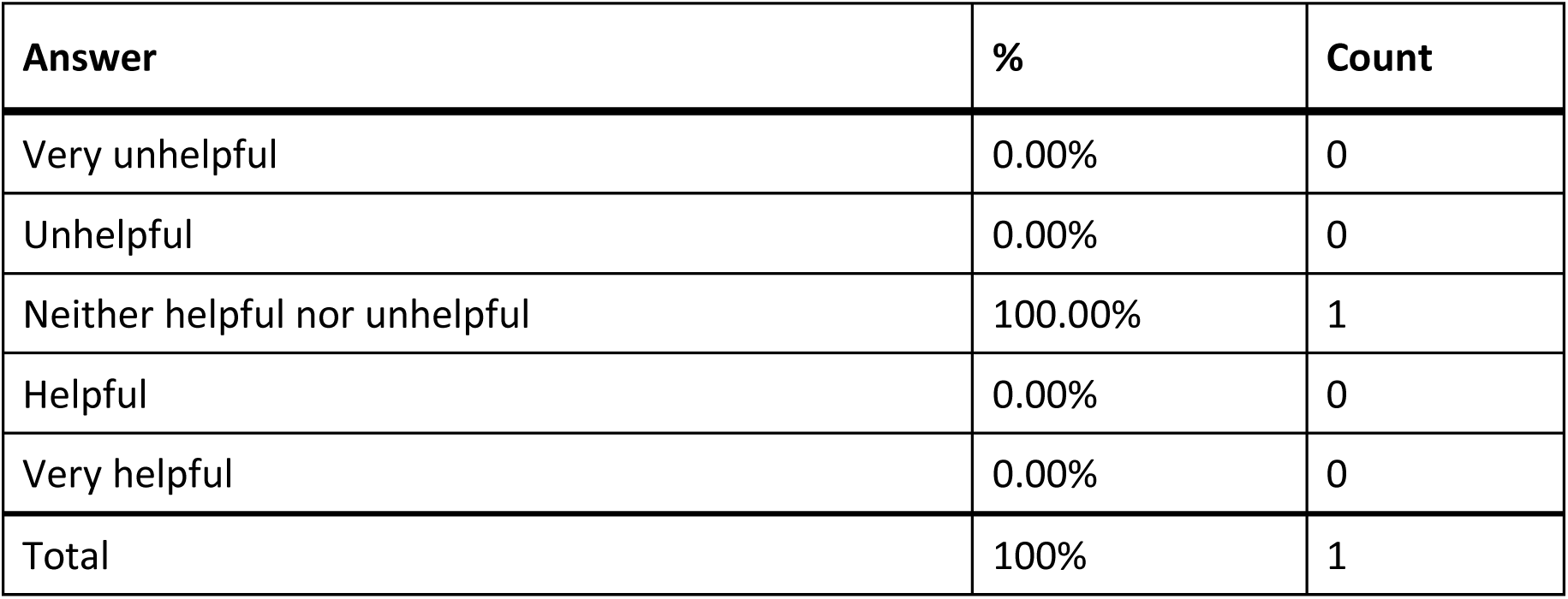
Responses to Q709: “Was [CareStart] customer support helpful in answering your question(s)?”

### Q710 - If you have performed the CareStart test multiple times, how did your experience change?

**Table 168.**
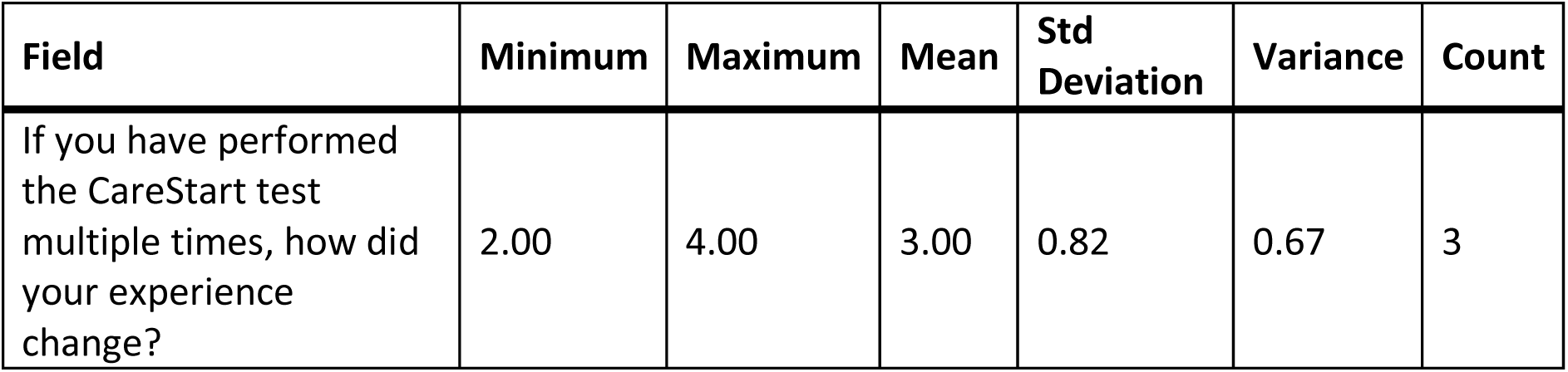
Stats for Q710: “If you have performed the CareStart test multiple times, how did your experience change?”

| Field | Minimum | Maximum | Mean | Std Deviation | Variance | Count |
| --- | --- | --- | --- | --- | --- | --- |
| If you have performed the CareStart test multiple times, how did your experience change? | 2.00 | 4.00 | 3.00 | 0.82 | 0.67 | 3 |

**Table 169.**
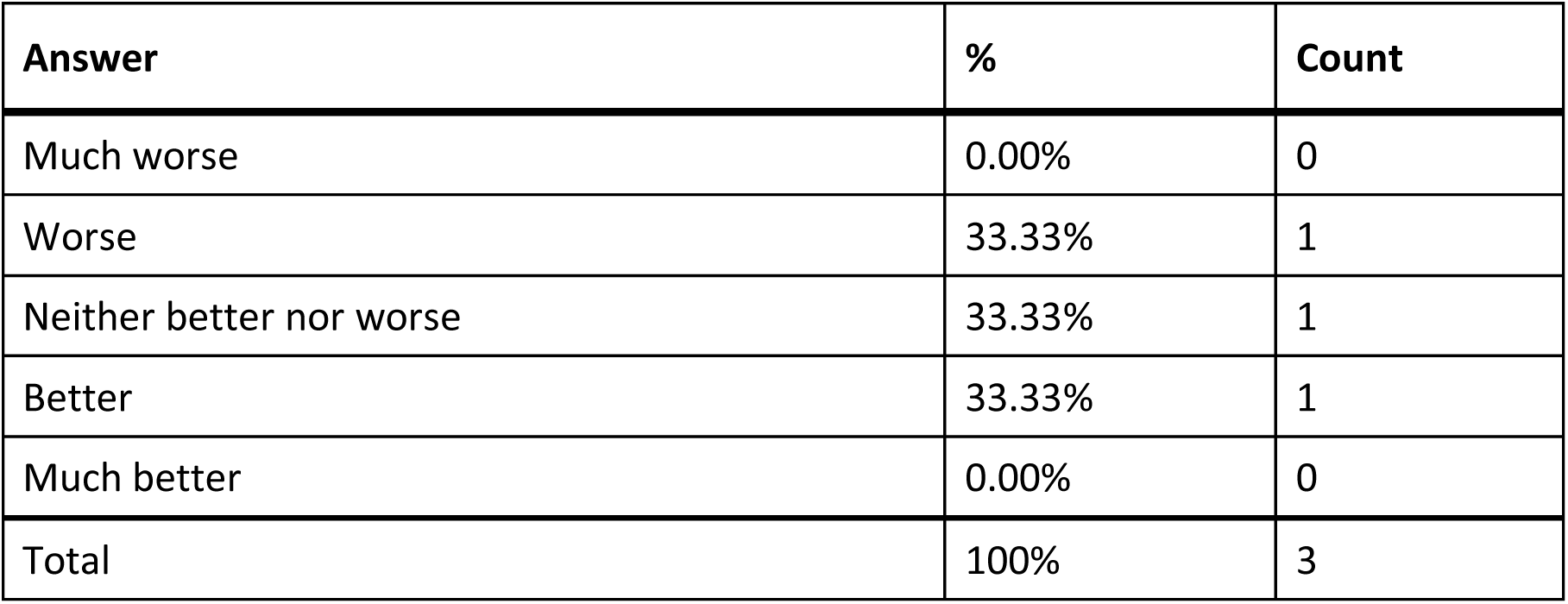
Responses to Q710: “If you have performed the CareStart test multiple times, how did your experience change?”

### Q711 - What did you like about the CareStart test?

**Table 170:**
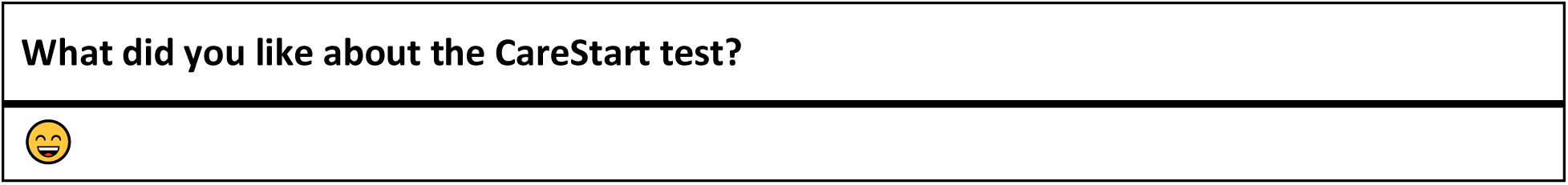
Open responses to Q711: “What did you like about the CareStart test?”

### Q712 - What would you like to see improved for the CareStart test?

**Table 171.**
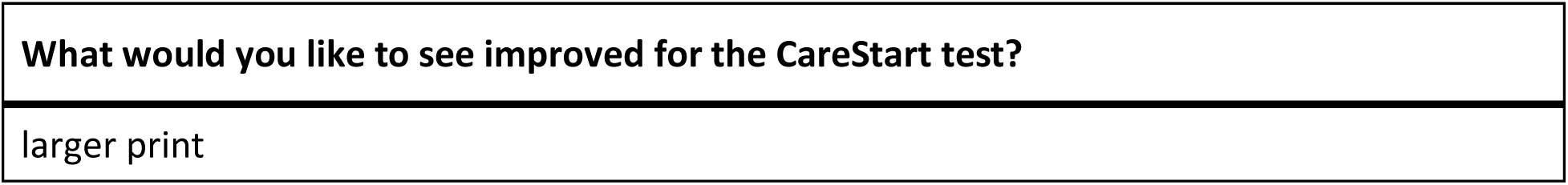
Open response to Q712: “What would you like to see improved for the CareStart test?”

### Q713 - Flowflex Think of the first time you used the Flowflex test. Did you have difficulty completing any of the following tasks? Select all that apply

**Table 172.** Responses to Q713: “Flowflex - Did you have difficulty completing any of the following tasks?”

| Answer | % | Count |
| --- | --- | --- |
| Opening the packaging | 5.45% | 3 |
| Reading the instructions | 23.64% | 13 |
| Identifying test components | 14.55% | 8 |
| Collecting the test sample | 7.27% | 4 |
| Manipulating test components | 16.36% | 9 |
| Interpreting the test results | 20.00% | 11 |
| Other | 0.00% | 0 |
| I did not have difficulty completing the test | 12.73% | 7 |
| Total | 100% | 55 |

#### Q713_7_TEXT - Other

None

### Q714 - What tools or assistance, if any, did you use to perform the test?

**Table 173.** Responses to Q714: “What tools or assistance, if any did you use to perform the [Flowflex] test?”

| Answer | % | Count |
| --- | --- | --- |
| Timer | 21.05% | 8 |
| Scissors | 10.53% | 4 |
| Remote visual assistance (AIRA, Be My Eyes, Facetime, Video-chat, etc.) | 7.89% | 3 |
| Screen Reader | 0.00% | 0 |
| Magnification | 10.53% | 4 |
| Other technology (OCR, Bar Code Readers, Light Detectors, etc.) | 7.89% | 3 |
| In person human assistance | 23.68% | 9 |
| Other | 0.00% | 0 |
| I did not need any tools or assistance | 18.42% | 7 |
| Total | 100% | 38 |

#### Q714_8_TEXT - Other

None

### Q715 - Were you able to conduct this test on your first try?

**Table 174.** Stats for Q715: “Were you able to conduct this [Flowflex] test on your first try?”

| Field | Minimum | Maximum | Mean | Std Deviation | Variance | Count |
| --- | --- | --- | --- | --- | --- | --- |
| Were you able to conduct this test on your first try? | 1.00 | 2.00 | 1.12 | 0.32 | 0.11 | 25 |

**Table 175.** Responses to Q715: “Were you able to conduct this [Flowflex] test on your first try?”

| Answer | % | Count |
| --- | --- | --- |
| Yes | 88.00% | 22 |
| No | 12.00% | 3 |
| Total | 100% | 25 |

### Q716 - How long did it take to complete the steps required to perform the test, not including the time you spent waiting for results

**Table 176.** Stats for Q716: “How long did it take to complete the steps required to perform the [Flowflex] test?”

| Field | Minimum | Maximum | Mean | Std Deviation | Variance | Count |
| --- | --- | --- | --- | --- | --- | --- |
| How long did it take to complete the steps required to perform the test, not including the time you spent waiting for results. | 1.00 | 4.00 | 1.89 | 0.87 | 0.77 | 27 |

**Table 177.** Responses to Q716: “How long did it take to complete the steps required to perform the [Flowflex] test?”

| Answer | % | Count |
| --- | --- | --- |
| Under 5 minutes | 40.74% | 11 |
| 5 - 10 minutes | 33.33% | 9 |
| 10 - 15 minutes | 22.22% | 6 |
| Over 15 minutes | 3.70% | 1 |
| Total | 100% | 27 |

### Q717 - What format of instructions did you use?

**Table 178.** Responses to Q717: “What format of [Flowflex] instructions did you use?”

| Answer | % | Count |
| --- | --- | --- |
| Paper | 61.29% | 19 |
| Digital (webpages, downloaded, etc.) | 19.35% | 6 |
| None of the above | 16.13% | 5 |
| Associated Covid Test App (Smartphone) | 3.23% | 1 |
| Total | 100% | 31 |

### Q718 - Were you able to access electronic information via a QR Code?

**Table 179.** Stats for Q718: “Were you able to access [Flowflex] electronic information via a QR Code?”

| Field | Minimum | Maximum | Mean | Std Deviation | Variance | Count |
| --- | --- | --- | --- | --- | --- | --- |
| Were you able to access electronic information via a QR Code? | 1.00 | 7.00 | 3.40 | 2.94 | 8.64 | 5 |

**Table 180.**
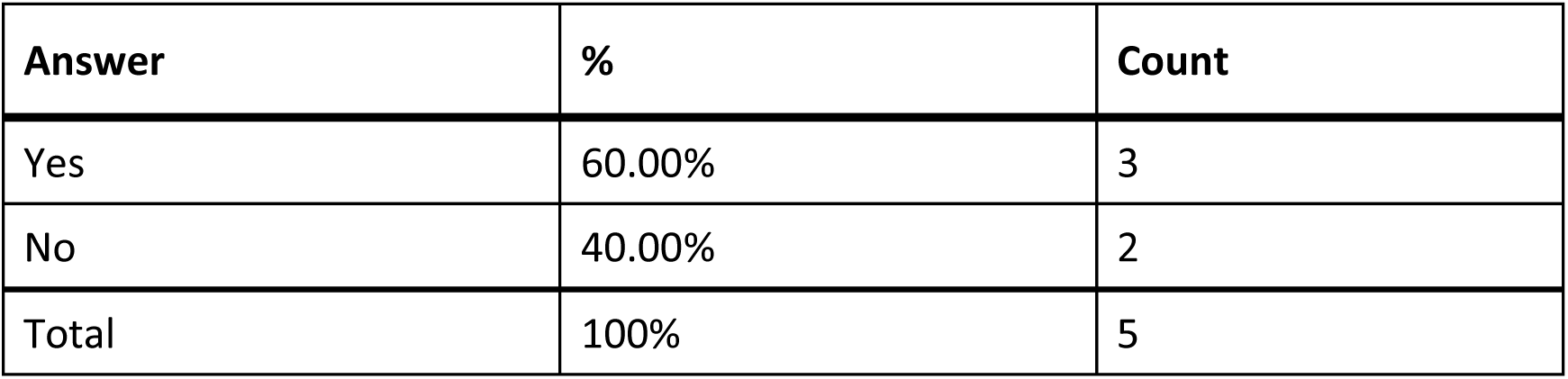
Responses to Q718: “Were you able to access [Flowflex] electronic information via a QR Code?”

### Q719 - How easy was opening the package of the Flowflex test for you? (without assistance)

**Table 181.** Stats for Q719: “How easy was opening the package of the Flowflex test for you? (without assistance)”

| Field | Minimum | Maximum | Mean | Std Deviation | Variance | Count |
| --- | --- | --- | --- | --- | --- | --- |
| How easy was opening the package of the Flowflex test for you? (without assistance) | 1.00 | 5.00 | 3.15 | 1.21 | 1.46 | 27 |

**Table 182.** Responses to Q719: “How easy was opening the package of the Flowflex tests for you? (without assistance)”

| Answer | % | Count |
| --- | --- | --- |
| Very difficult | 11.11% | 3 |
| Difficult | 18.52% | 5 |
| Neither easy nor difficult | 29.63% | 8 |
| Easy | 25.93% | 7 |
| Very easy | 14.81% | 4 |
| Total | 100% | 27 |

### Q720 - How easy was completing the test protocol for you? (without assistance)

**Table 183.** Stats for Q720: “How easy was completing the [Flowflex] test protocol for you? (without assistance)”

| Field | Minimum | Maximum | Mean | Std Deviation | Variance | Count |
| --- | --- | --- | --- | --- | --- | --- |
| How easy was completing the test protocol for you? (without assistance) | 1.00 | 5.00 | 3.00 | 1.20 | 1.44 | 25 |

**Table 184.** Responses to Q720: “How easy was completing the [Flowflex] test protocol for you? (without assistance)”

| Answer | % | Count |
| --- | --- | --- |
| Very difficult | 12.00% | 3 |
| Difficult | 24.00% | 6 |
| Neither easy nor difficult | 28.00% | 7 |
| Easy | 24.00% | 6 |
| Very easy | 12.00% | 3 |
| Total | 100% | 25 |

### Q721 - How easy was interpreting the test results for you? (without assistance)

**Table 185.**
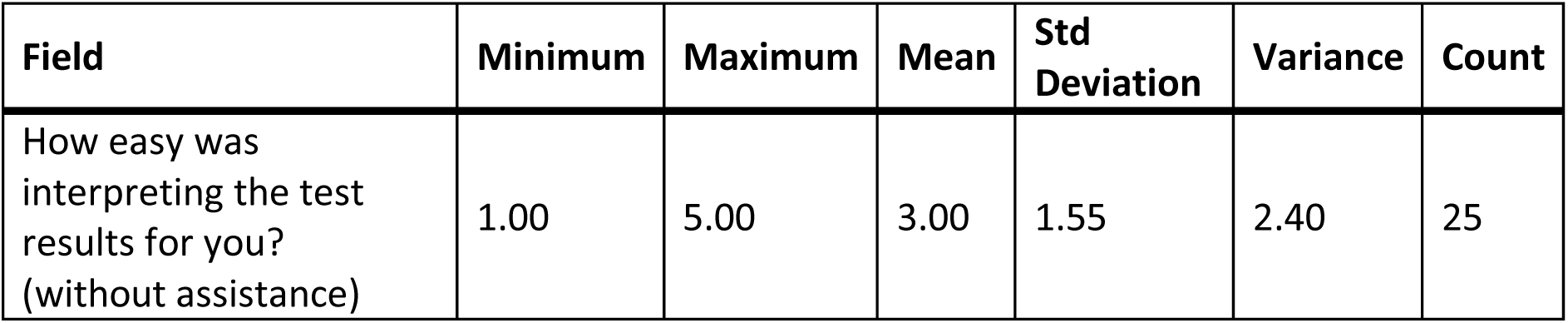
Stats for Q721: “How easy was interpreting the [Flowflex] test results for you? (without assistance)”

| Field | Minimum | Maximum | Mean | Std Deviation | Variance | Count |
| --- | --- | --- | --- | --- | --- | --- |
| How easy was interpreting the test results for you? (without assistance) | 1.00 | 5.00 | 3.00 | 1.55 | 2.40 | 25 |

**Table 186.**
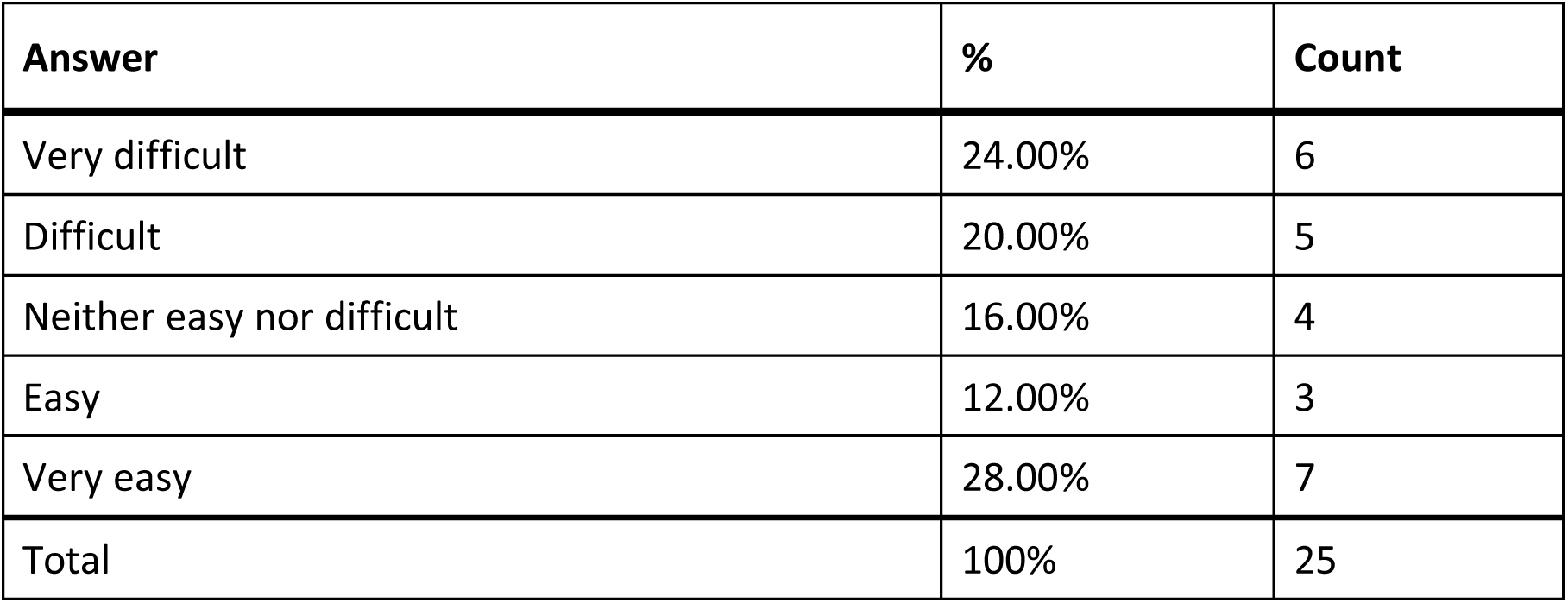
Responses to Q721: “How easy was interpreting the [Flowflex] test results for you? (without assistance)”

### Q722 - Were you able to interpret the test results? (without assistance)

**Table 187.** Stats for Q722: “Were you able to interpret the [Flowflex] test results? (without assistance)”

| Field | Minimum | Maximum | Mean | Std Deviation | Variance | Count |
| --- | --- | --- | --- | --- | --- | --- |
| Were you able to interpret the test results? (without assistance) | 1.00 | 2.00 | 1.35 | 0.48 | 0.23 | 26 |

**Table 188.** Responses to Q722: “Were you able to interpret the [Flowflex] test results? (without assistance)”

| Answer | % | Count |
| --- | --- | --- |
| Yes | 65.38% | 17 |
| No | 34.62% | 9 |
| Total | 100% | 26 |

### Q723 - If there was an associated app for the test, how easy was it for you to use? (without assistance)

**Table 189.**
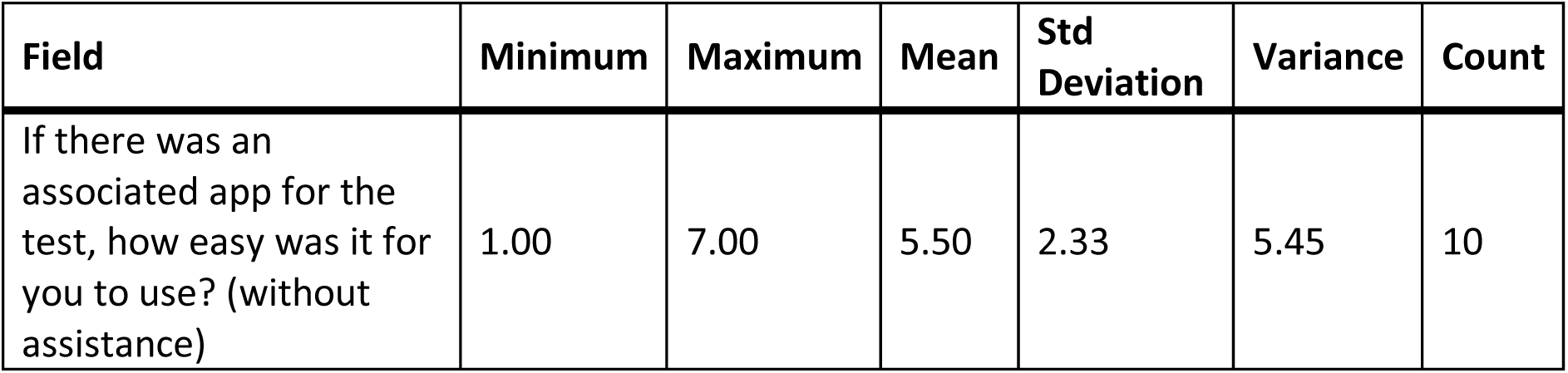
Stats for Q723: “If there was an associated app for the [Flowflex] test, how easy was it for you to use? (without assistance)”

**Table 190.**
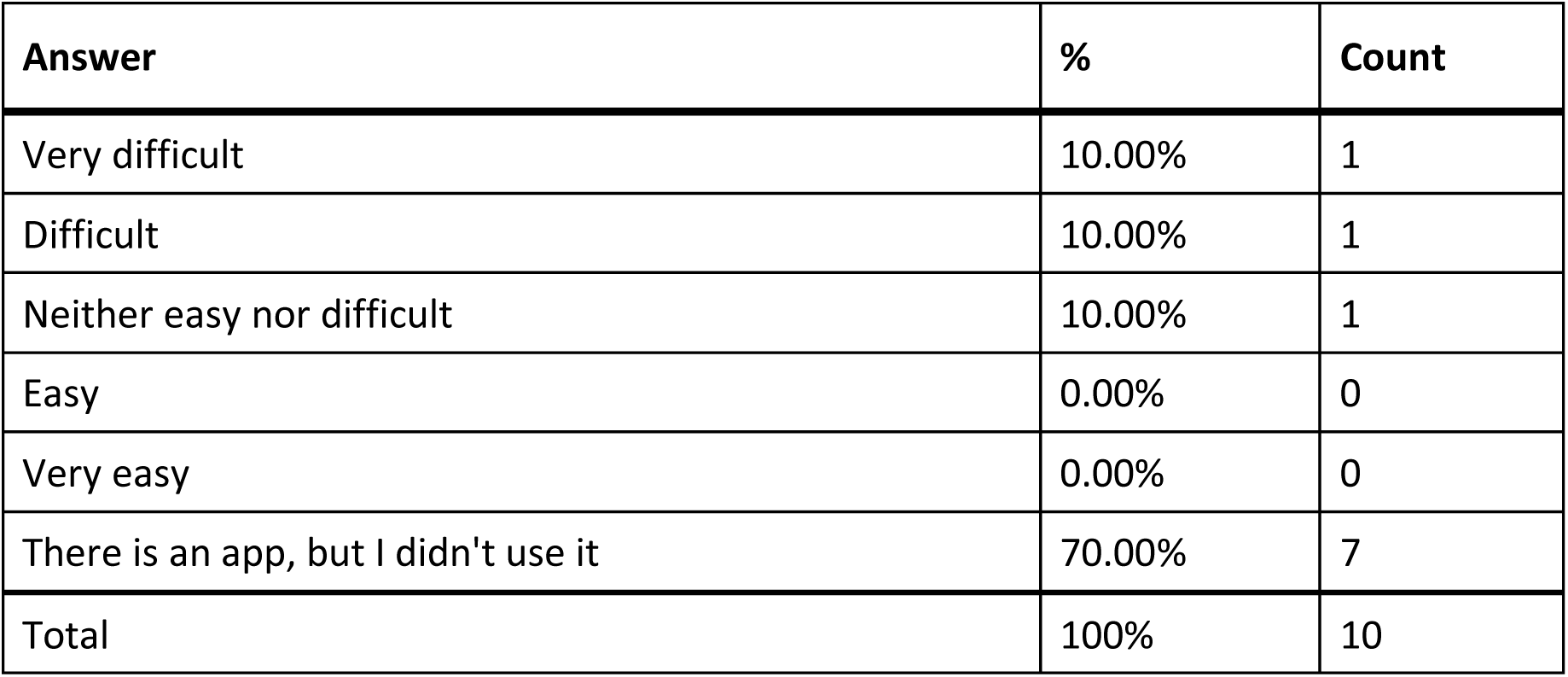
Responses to Q723: “If there was an associated app for the [Flowflex] test, how easy was it for you to use? (without assistance)”

| Answer | % | Count |
| --- | --- | --- |
| Very difficult | 10.00% | 1 |
| Difficult | 10.00% | 1 |
| Neither easy nor difficult | 10.00% | 1 |
| Easy | 0.00% | 0 |
| Very easy | 0.00% | 0 |
| There is an app, but I didn't use it | 70.00% | 7 |
| Total | 100% | 10 |

### Q724 - How helpful was the app in assisting you with completing the test?

**Table 191.** Stats for Q724: “How helpful was the app in assisting you with completing the [Flowflex] test?”

| Field | Minimum | Maximum | Mean | Std Deviation | Variance | Count |
| --- | --- | --- | --- | --- | --- | --- |
| How helpful was the app in assisting you with completing the test? | 1.00 | 4.00 | 2.33 | 1.25 | 1.56 | 3 |

**Table 192.** Responses to Q724: “How helpful was the app in assisting you with completing the [Flowflex] test?”

| Answer | % | Count |
| --- | --- | --- |
| Very unhelpful | 33.33% | 1 |
| Unhelpful | 33.33% | 1 |
| Neither unhelpful nor helpful | 0.00% | 0 |
| Helpful | 33.33% | 1 |
| Very helpful | 0.00% | 0 |
| Total | 100% | 3 |

### Q725 - Did you receive a valid test result with the Flowflex test (positive or negative)?

**Table 193.** Stats for Q725: “Did you receive a valid test result with the Flowflex test (positive or negative)?”

| Field | Minimum | Maximum | Mean | Std Deviation | Variance | Count |
| --- | --- | --- | --- | --- | --- | --- |
| Did you receive a valid test result with the Flowflex test (positive or negative)? | 1.00 | 2.00 | 1.12 | 0.32 | 0.10 | 26 |

**Table 194.**
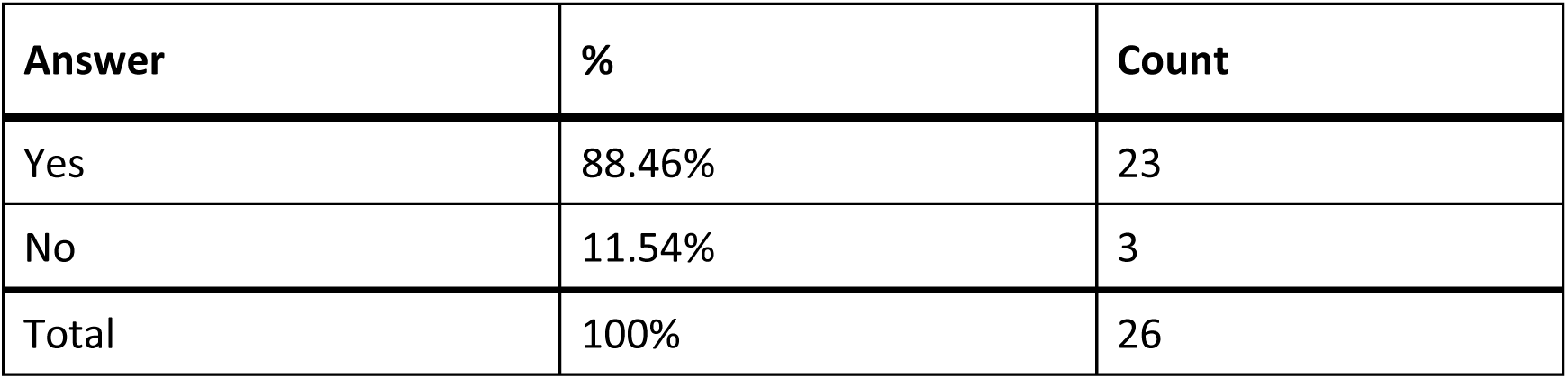
Responses to Q725: “Did you receive a valid test results with the Flowflex test (positive or negative)?”

| Answer | % | Count |
| --- | --- | --- |
| Yes | 88.46% | 23 |
| No | 11.54% | 3 |
| Total | 100% | 26 |

### Q726 - Did you attempt to contact Flowflex’s customer support?

**Table 195.** Stats for Q726: “Did you attempt to contact Flowflex’s customer support?”

| Field | Minimum | Maximum | Mean | Std Deviation | Variance | Count |
| --- | --- | --- | --- | --- | --- | --- |
| Did you attempt to contact Flowflex's customer support? | 1.00 | 2.00 | 1.89 | 0.31 | 0.10 | 27 |

**Table 196.** Responses to Q726: “Did you attempt to contact Flowflex’s customer support?”

| Answer | % | Count |
| --- | --- | --- |
| Yes | 11.11% | 3 |
| No | 88.89% | 24 |
| Total | 100% | 27 |

### Q727 - Were you able to talk with customer support?

**Table 197.** Stats for Q:727: “Were you able to talk with [Flowflex] customer support?”

| Field | Minimum | Maximum | Mean | Std Deviation | Variance | Count |
| --- | --- | --- | --- | --- | --- | --- |
| Were you able to talk with customer support? | 1.00 | 1.00 | 1.00 | 0.00 | 0.00 | 3 |

**Table 198.**
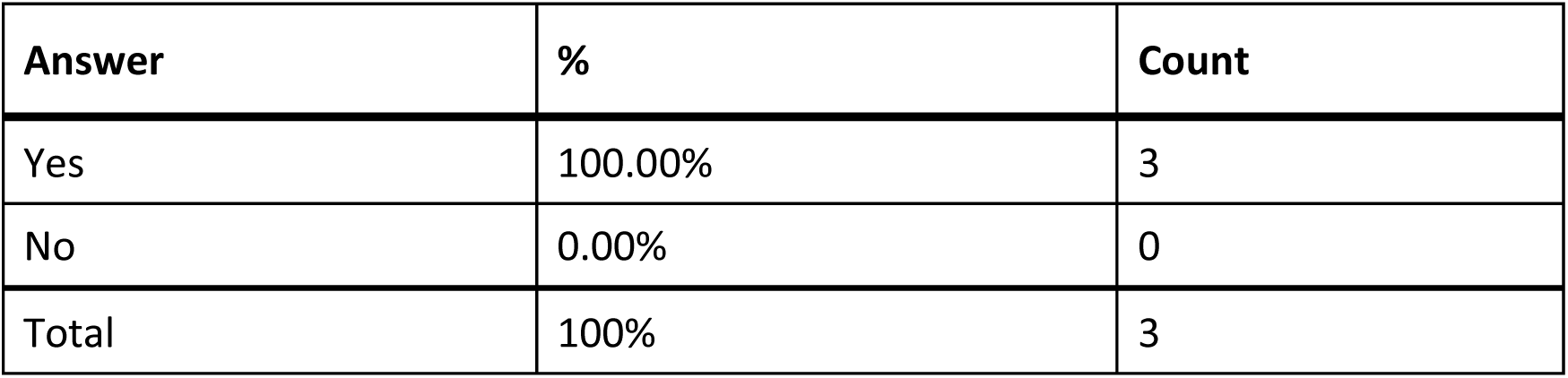
Responses to Q727: “Were you able to talk with customer support?”

### Q728 - Was customer support helpful in answering your question(s)?

**Table 199.** Stats for Q728: “Was [Flowflex] customer support helpful in answering your question(s)?

| Field | Minimum | Maximum | Mean | Std Deviation | Variance | Count |
| --- | --- | --- | --- | --- | --- | --- |
| Was customer support helpful in answering your question(s)? | 5.00 | 6.00 | 5.33 | 0.47 | 0.22 | 3 |

**Table 200.** Responses to Q728: “Was [Flowflex] customer support helpful in answering your question(s)?”

| Answer | % | Count |
| --- | --- | --- |
| Very unhelpful | 0.00% | 0 |
| Unhelpful | 0.00% | 0 |
| Neither helpful nor unhelpful | 66.67% | 2 |
| Helpful | 33.33% | 1 |
| Very helpful | 0.00% | 0 |
| Total | 100% | 3 |

### Q729 - If you have performed the Flowflex test multiple times, how did your experience change?

**Table 201.** Stats for Q729: “If you have performed the Flowflex test multiple times, how did your experience change?”

| Field | Minimum | Maximum | Mean | Std Deviation | Variance | Count |
| --- | --- | --- | --- | --- | --- | --- |
| If you have performed the Flowflex test multiple times, how did your experience change? | 1.00 | 5.00 | 3.10 | 0.77 | 0.59 | 20 |

**Table 202.** Responses to Q729: “If you have performed the Flowflex test multiple times, how did your experience change?”

| Answer | % | Count |
| --- | --- | --- |
| Much worse | 5.00% | 1 |
| Worse | 5.00% | 1 |
| Neither better nor worse | 70.00% | 14 |
| Better | 15.00% | 3 |
| Much better | 5.00% | 1 |
| Total | 100% | 20 |

### Q730 - What did you like about the Flowflex test?

**Table 203.** Open response to Q730: “What did you like about the Flowflex test?”

| **What did you like about the Flowflex test?** |
| --- |
| it is a little easier to use than other tests because I just put the sample right in the liquid |
| n/a |
| Relatively easy to use, didn't have to leave my home to get tested |
| Hi |
| N/A |
| Small package, less waste material than some other tests. |
| It was free and a sighted-supporter could help complete it |
| I neither like nor dislike the test. |
| The components were easy to identify, and the test was easy to use with remote visual assistance. |
| This wasn't accessible at all. And it didn't explain very well. |
| The directions were in a slightly larger print than the previous test I had taken. |
| Fast results |
| The instructions on the Flowflex test were so small in print that my brother could not read it. |
| Very easy to use. |
| Liked the use of the swab in a tube rather than a dropper in the cassette. |
| Placing swab in card easier than droplet tube. |
| Nothing . Confusing. |

### Q731 - What would you like to see improved for the Flowflex test?

**Table 204.** Open response to Q731: “What would you like to see improved for the Flowflex test?”

| What would you like to see improved for the Flowflex test? |
| --- |
| I just needed another set of hands to do it well |
| n/a |
| Charge to purchase the test is consistent and low cost so consumers of all socioeconomic backgrounds can afford |
| N/A |
| A way to provide accessible instructions and results. |
| Same problem with instruction. Lots of warnings. looked identical to the other brand... just a different box. |
| I have no opinion on this. The test would need to be completely redesigned in order to make it an accessible and useful tool. |
| As with any test, I would like to see the test made accessible without the need to use remote visual assistance. |
| Accessibility features. |
| A larger testing surface, so that it's easier to determine a negative vs. positive result. |
| Tactile identification in which end opens for the swab. |
| It would help if the Flowflex test would have large print or audible instructions then I would not have to depend on someone to assist me in performing COVID test. |
| Most of the survey is not applicable because I got in person human assistance for the tests but ideally wouldn't need to pour anything and have a more straight forward process like most of the RAT tests |
| The instructions and drawings for the steps involving swirling the swab are confusing. |
| Read results |
| Make it like Binax. |

### Q732 - Roche Think of the first time you used the Roche test. Did you have difficulty completing any of the following tasks? Select all that apply

**Table 205.** Response to Q732: “Roche - Did you have any difficulty completing any of the following tasks?”

| Answer | % | Count |
| --- | --- | --- |
| Opening the packaging | 10.53% | 4 |
| Reading the instructions | 21.05% | 8 |
| Identifying test components | 10.53% | 4 |
| Collecting the test sample | 7.89% | 3 |
| Manipulating test components | 21.05% | 8 |
| Interpreting the test results | 15.79% | 6 |
| Other | 0.00% | 0 |
| I did not have difficulty completing the test | 13.16% | 5 |
| Total | 100% | 38 |

#### Q732_7_TEXT - Other

None

### Q733 - What tools or assistance, if any, did you use to perform the test?

**Table 206.** Responses to Q733: “What tools or assistance, if any, did you use to perform the [Roche] test?”

| Answer | % | Count |
| --- | --- | --- |
| Timer | 20.59% | 7 |
| Scissors | 23.53% | 8 |
| Remote visual assistance (AIRA, Be My Eyes, Facetime, Video-chat, etc.) | 11.76% | 4 |
| Screen Reader | 5.88% | 2 |
| Magnification | 11.76% | 4 |
| Other technology (OCR, Bar Code Readers, Light Detectors, etc.) | 5.88% | 2 |
| In person human assistance | 14.71% | 5 |
| Other | 2.94% | 1 |
| I did not need any tools or assistance | 2.94% | 1 |
| Total | 100% | 34 |

#### Q733_8_TEXT - Other

None

### Q734 - Were you able to conduct this test on your first try?

**Table 207.** Stats for Q734: “Were you able to conduct this [Roche] test on your first try?”

| Field | Minimum | Maximum | Mean | Std Deviation | Variance | Count |
| --- | --- | --- | --- | --- | --- | --- |
| Were you able to conduct this test on your first try? | 1.00 | 2.00 | 1.19 | 0.39 | 0.15 | 16 |

**Table 208.**
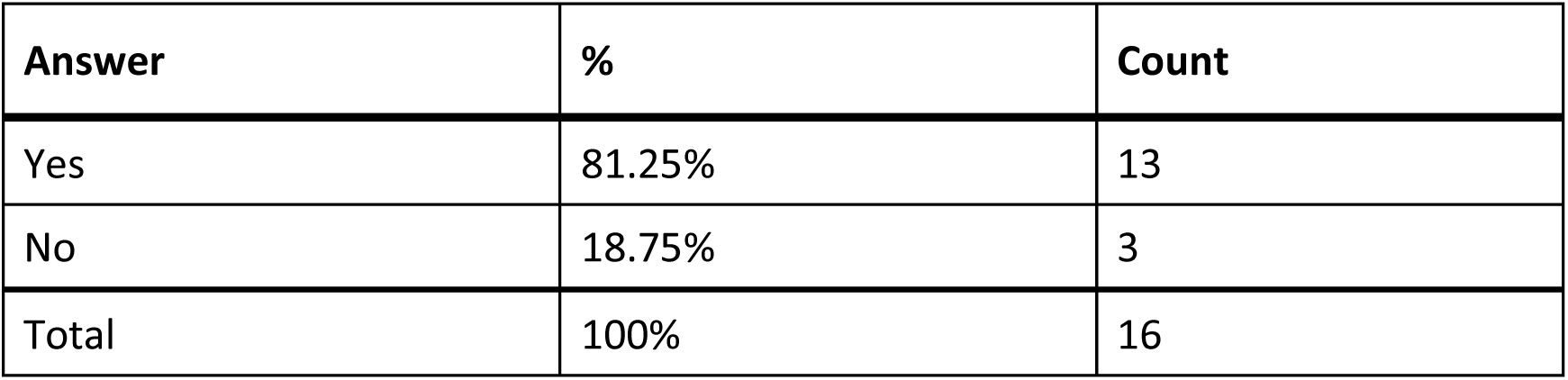
Responses to Q734:“Were you able to conduct this [Roche] test on your first try?”

| Answer | % | Count |
| --- | --- | --- |
| Yes | 81.25% | 13 |
| No | 18.75% | 3 |
| Total | 100% | 16 |

### Q735 - How long did it take to complete the steps required to perform the test, not including the time you spent waiting for results

**Table 209.** Stats for Q735: “How long did it take to complete the steps required to perform the [Roche] test?”

| Field | Minimum | Maximum | Mean | Std Deviation | Variance | Count |
| --- | --- | --- | --- | --- | --- | --- |
| How long did it take to complete the steps required to perform the test, not including the time you spent waiting for results. | 1.00 | 4.00 | 2.50 | 1.12 | 1.25 | 16 |

**Table 210.** Responses to Q735: “How long did it take to complete the steps required to perform the [Roche] test?”

| Answer | % | Count |
| --- | --- | --- |
| Under 5 minutes | 18.75% | 3 |
| 5 - 10 minutes | 43.75% | 7 |
| 10 - 15 minutes | 6.25% | 1 |
| Over 15 minutes | 31.25% | 5 |
| Total | 100% | 16 |

### Q736 - What format of instructions did you use?

**Table 211.** Responses to Q736: “What format of [Roche] instructions did you use?”

| Answer | % | Count |
| --- | --- | --- |
| Paper | 61.11% | 11 |
| Digital (webpages, downloaded, etc.) | 22.22% | 4 |
| None of the above | 11.11% | 2 |
| Associated Covid Test App (Smartphone) | 5.56% | 1 |
| Total | 100% | 18 |

### Q737 - Were you able to access electronic information via a QR Code?

**Table 212.** Stats for Q737: “Were you able to access electronic information via [Roche} QR Code?”

| Field | Minimum | Maximum | Mean | Std Deviation | Variance | Count |
| --- | --- | --- | --- | --- | --- | --- |
| Were you able to access electronic information via a QR Code? | 1.00 | 7.00 | 4.00 | 3.00 | 9.00 | 2 |

**Table 213.**
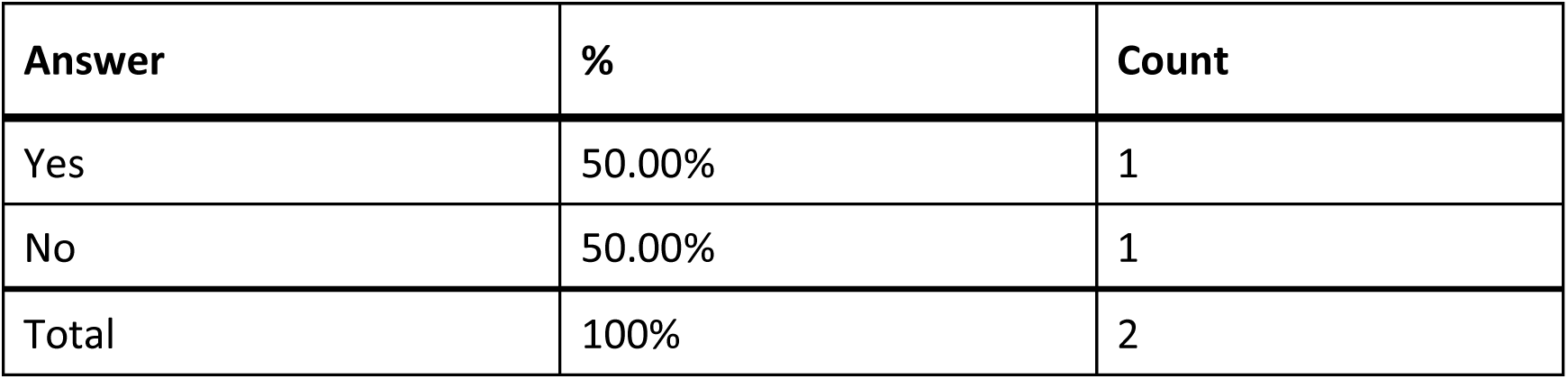
Responses to Q737: “Were you able to access electronic information via [Roche] QR Code?”

### Q738 - How easy was opening the package of the Roche test for you? (without assistance)

**Table 214.** Stats for Q738: “How easy was opening the package of the Roche test for you? (without assistance)”

| Field | Minimum | Maximum | Mean | Std Deviation | Variance | Count |
| --- | --- | --- | --- | --- | --- | --- |
| How easy was opening the package of the Roche test for you? (without assistance) | 1.00 | 5.00 | 3.47 | 1.31 | 1.72 | 15 |

**Table 215.** Responses to Q738: “How easy was opening the package of the Roche test for you? (without assistance)”

| Answer | % | Count |
| --- | --- | --- |
| Very difficult | 6.67% | 1 |
| Difficult | 20.00% | 3 |
| Neither easy nor difficult | 26.67% | 4 |
| Easy | 13.33% | 2 |
| Very easy | 33.33% | 5 |
| Total | 100% | 15 |

### Q739 - How easy was completing the test protocol for you? (without assistance)

**Table 216.** Stats for Q739: “How easy was completing the [Roche] test protocol for you? (without assistance)”

| Field | Minimum | Maximum | Mean | Std Deviation | Variance | Count |
| --- | --- | --- | --- | --- | --- | --- |
| How easy was completing the test protocol for you? (without assistance) | 1.00 | 5.00 | 3.13 | 1.36 | 1.85 | 15 |

**Table 217.** Responses to Q739: “How easy was completing the [Roche] test protocol for you? (without assistance)”

| Answer | % | Count |
| --- | --- | --- |
| Very difficult | 6.67% | 1 |
| Difficult | 40.00% | 6 |
| Neither easy nor difficult | 13.33% | 2 |
| Easy | 13.33% | 2 |
| Very easy | 26.67% | 4 |
| Total | 100% | 15 |

### Q740 - How easy was interpreting the test results for you? (without assistance)

**Table 218.**
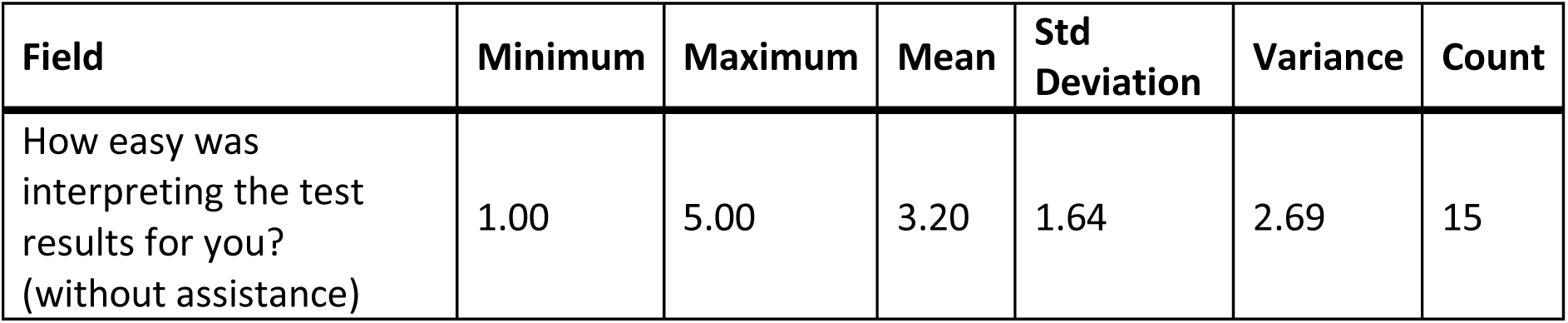
Stats for Q740: “How easy was interpreting the [Roche] test results for you? (without assistance)”

**Table 219.** Responses to Q740: “How easy was interpreting the [Roche] test results for you? (without assistance)”

| Answer | % | Count |
| --- | --- | --- |
| Very difficult | 26.67% | 4 |
| Difficult | 13.33% | 2 |
| Neither easy nor difficult | 6.67% | 1 |
| Easy | 20.00% | 3 |
| Very easy | 33.33% | 5 |
| Total | 100% | 15 |

### Q741 - Were you able to interpret the test results? (without assistance)

**Table 220.**
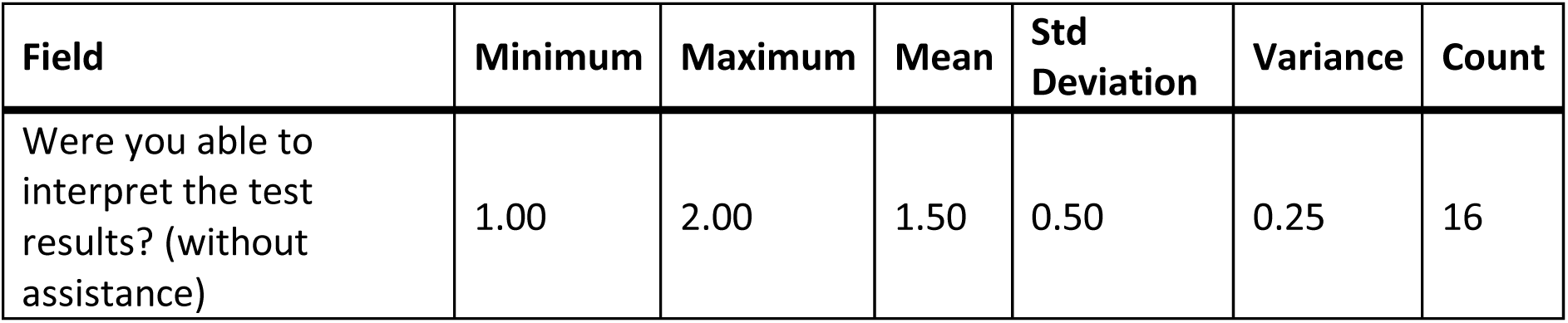
Stats for Q741: “Were you able to interpret the test results? (with assistance)”

**Table 221.** Responses to Q741: “Were you able to interpret the test results? (with assistance)”

| Answer | % | Count |
| --- | --- | --- |
| Yes | 50.00% | 8 |
| No | 50.00% | 8 |
| Total | 100% | 16 |

### Q742 - If there was an associated app for the test, how easy was it for you to use? (without assistance)

**Table 222.**
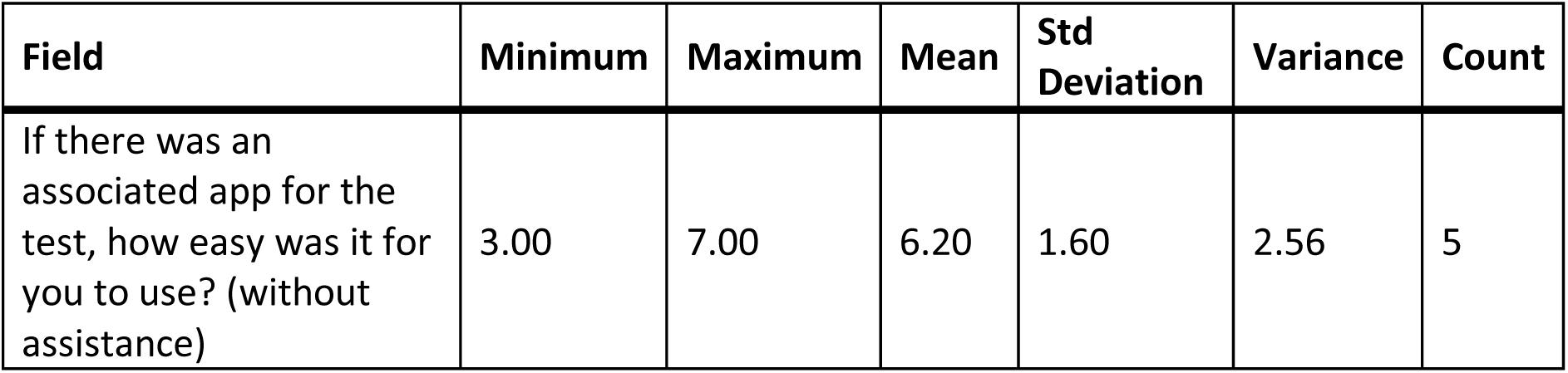
Stats for Q742: “If there was an associated app for the [Roche] test, how easy was it for you to use? (without assistance)”

| Field | Minimum | Maximum | Mean | Std Deviation | Variance | Count |
| --- | --- | --- | --- | --- | --- | --- |
| If there was an associated app for the test, how easy was it for you to use? (without assistance) | 3.00 | 7.00 | 6.20 | 1.60 | 2.56 | 5 |

**Table 223.** Response to Q742: “If there was an associated app for the [Roche] test, how easy was it for you to use? (without assistance)”

| Answer | % | Count |
| --- | --- | --- |
| Very difficult | 0.00% | 0 |
| Difficult | 0.00% | 0 |
| Neither easy nor difficult | 20.00% | 1 |
| Easy | 0.00% | 0 |
| Very easy | 0.00% | 0 |
| There is an app, but I didn't use it | 80.00% | 4 |
| Total | 100% | 5 |

### Q743 - How helpful was the app in assisting you with completing the test?

**Table 224.** Stats for Q743: “How helpful was the app in assisting you with completing the [Roche] test?”

| Field | Minimum | Maximum | Mean | Std Deviation | Variance | Count |
| --- | --- | --- | --- | --- | --- | --- |
| How helpful was the app in assisting you with completing the test? | 3.00 | 3.00 | 3.00 | 0.00 | 0.00 | 1 |

**Table 225.** Responses to Q743: “How helpful was the app in assisting you with completing the [Roche] test?”

| Answer | % | Count |
| --- | --- | --- |
| Very unhelpful | 0.00% | 0 |
| Unhelpful | 0.00% | 0 |
| Neither unhelpful nor helpful | 100.00% | 1 |
| Helpful | 0.00% | 0 |
| Very helpful | 0.00% | 0 |
| Total | 100% | 1 |

### Q744 - Did you receive a valid test result with the Roche test (positive or negative)?

**Table 226.**
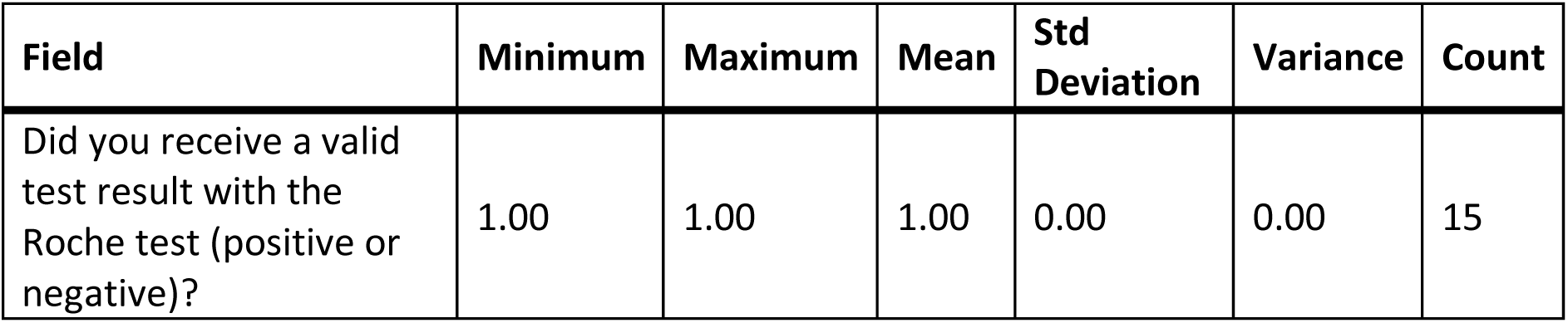
Stats for Q744: “Did you receive a valid test result with the Roche test (positive or negative)?”

| Field | Minimum | Maximum | Mean | Std Deviation | Variance | Count |
| --- | --- | --- | --- | --- | --- | --- |
| Did you receive a valid test result with the Roche test (positive or negative)? | 1.00 | 1.00 | 1.00 | 0.00 | 0.00 | 15 |

**Table 227.** Responses to Q744: “Did you receive a valid test result with the Roche test (positive or negative)?”

| Answer | % | Count |
| --- | --- | --- |
| Yes | 100.00% | 15 |
| No | 0.00% | 0 |
| Total | 100% | 15 |

### Q745 - Did you attempt to contact Roche’s customer support?

**Table 228.** Stats for Q745: “Did you attempt to contact Roche’s customer support?”

| Field | Minimum | Maximum | Mean | Std Deviation | Variance | Count |
| --- | --- | --- | --- | --- | --- | --- |
| Did you attempt to contact Roche's customer support? | 2.00 | 2.00 | 2.00 | 0.00 | 0.00 | 16 |

**Table 229.** Responses to Q745: “Did you attempt to contact Roche’s customer support?”

| Answer | % | Count |
| --- | --- | --- |
| Yes | 0.00% | 0 |
| No | 100.00% | 16 |
| Total | 100% | 16 |

### Q746 - Were you able to talk with customer support?

**Table 230.** Stats for Q746: “Were you able to talk with [Roche] customer support?”

| Field | Minimum | Maximum | Mean | Std Deviation | Variance | Count |
| --- | --- | --- | --- | --- | --- | --- |
| Were you able to talk with customer support? | 0.00 | 0.00 | 0.00 | 0.00 | 0.00 | 0 |

**Table 231.** Responses to Q746: “Were you able to talk with [Roche] customer support?”

| Answer | % | Count |
| --- | --- | --- |
| Yes | 0.00% | 0 |
| No | 0.00% | 0 |
| Total |  | 0 |

### Q747 - Was customer support helpful in answering your question(s)?

**Table 232.**
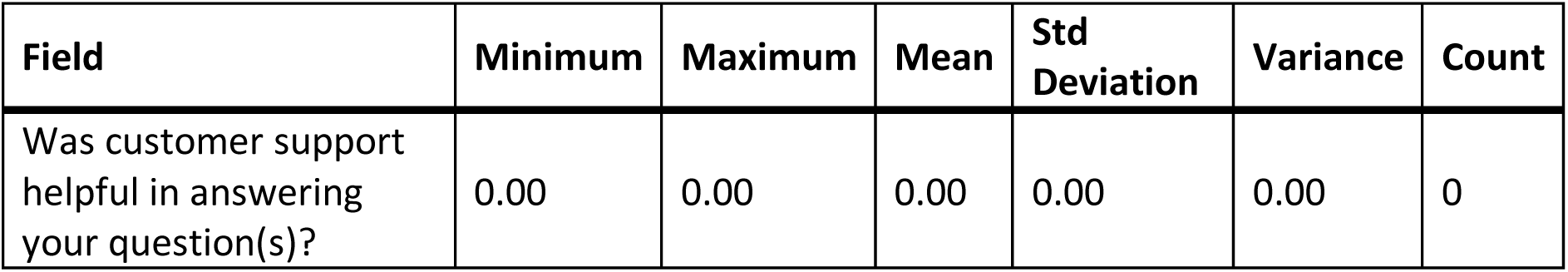
Stats for Q747: “Was [Roche] customer support helpful in answering your question(s)?”

**Table 233.**
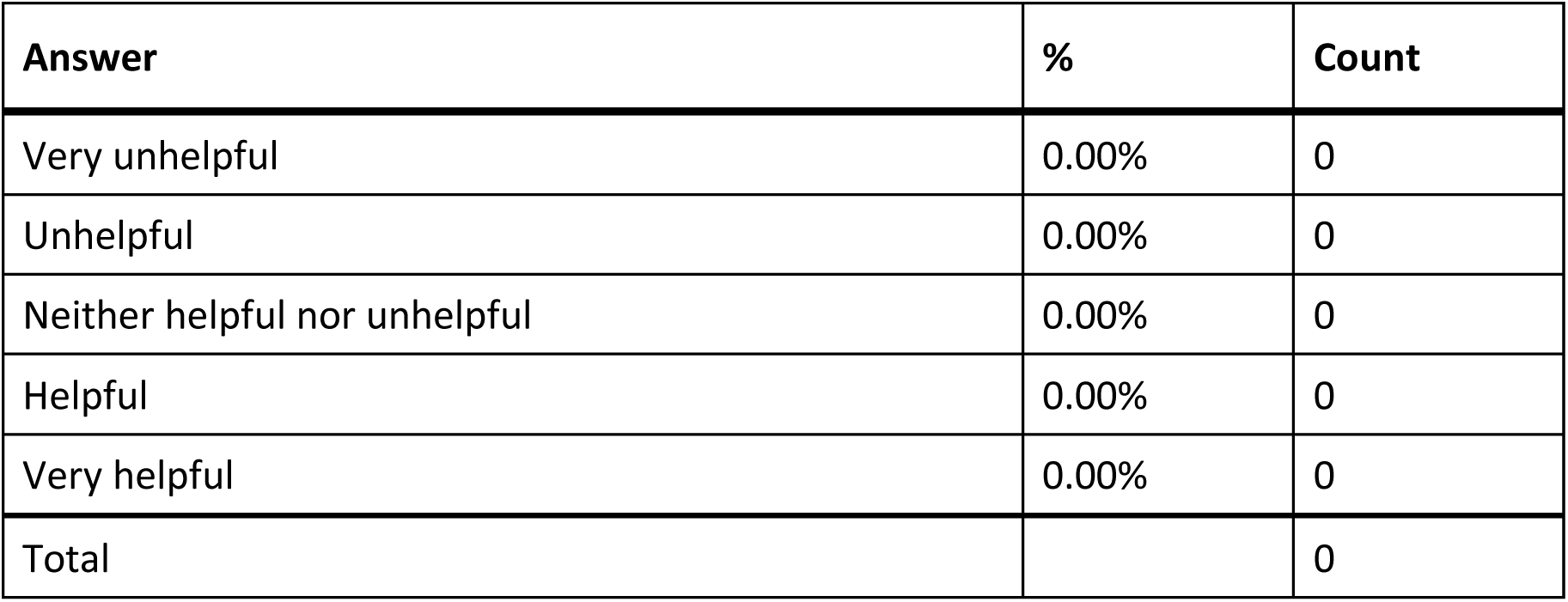
Responses to Q747: “Was [Roche] customer support helpful in answering your question(s)?”

| Answer | % | Count |
| --- | --- | --- |
| Very unhelpful | 0.00% | 0 |
| Unhelpful | 0.00% | 0 |
| Neither helpful nor unhelpful | 0.00% | 0 |
| Helpful | 0.00% | 0 |
| Very helpful | 0.00% | 0 |
| Total |  | 0 |

### Q748 - If you have performed the Roche test multiple times, how did your experience change?

**Table 234.** Stats for Q748: “If you have performed the Roche test multiple times, how did you experience change?”

| Field | Minimum | Maximum | Mean | Std Deviation | Variance | Count |
| --- | --- | --- | --- | --- | --- | --- |
| If you have performed the Roche test multiple times, how did your experience change? | 3.00 | 4.00 | 3.55 | 0.50 | 0.25 | 11 |

**Table 235.** Responses to Q748: “If you have performed the Roche test multiple times, how did you experience change?”

| Answer | % | Count |
| --- | --- | --- |
| Much worse | 0.00% | 0 |
| Worse | 0.00% | 0 |
| Neither better nor worse | 45.45% | 5 |
| Better | 54.55% | 6 |
| Much better | 0.00% | 0 |
| Total | 100% | 11 |

### Q749 - What did you like about the Roche test?

**Table 236.** Open response to Q749: “What did you like about the Roche test?”

| What did you like about the Roche test? |
| --- |
| Easy to administer. |
| N/A |
| Solution already in test tube. |
| ease of use; clarity |
| 4 tests in one pack |
| Nothing. |
| Ability to use in the privacy of my home |
| It worked. |
| Instructions were simple, and the product was easy to use. |
| nothing |

### Q750 - What would you like to see improved for the Roche test?

**Table 237.**
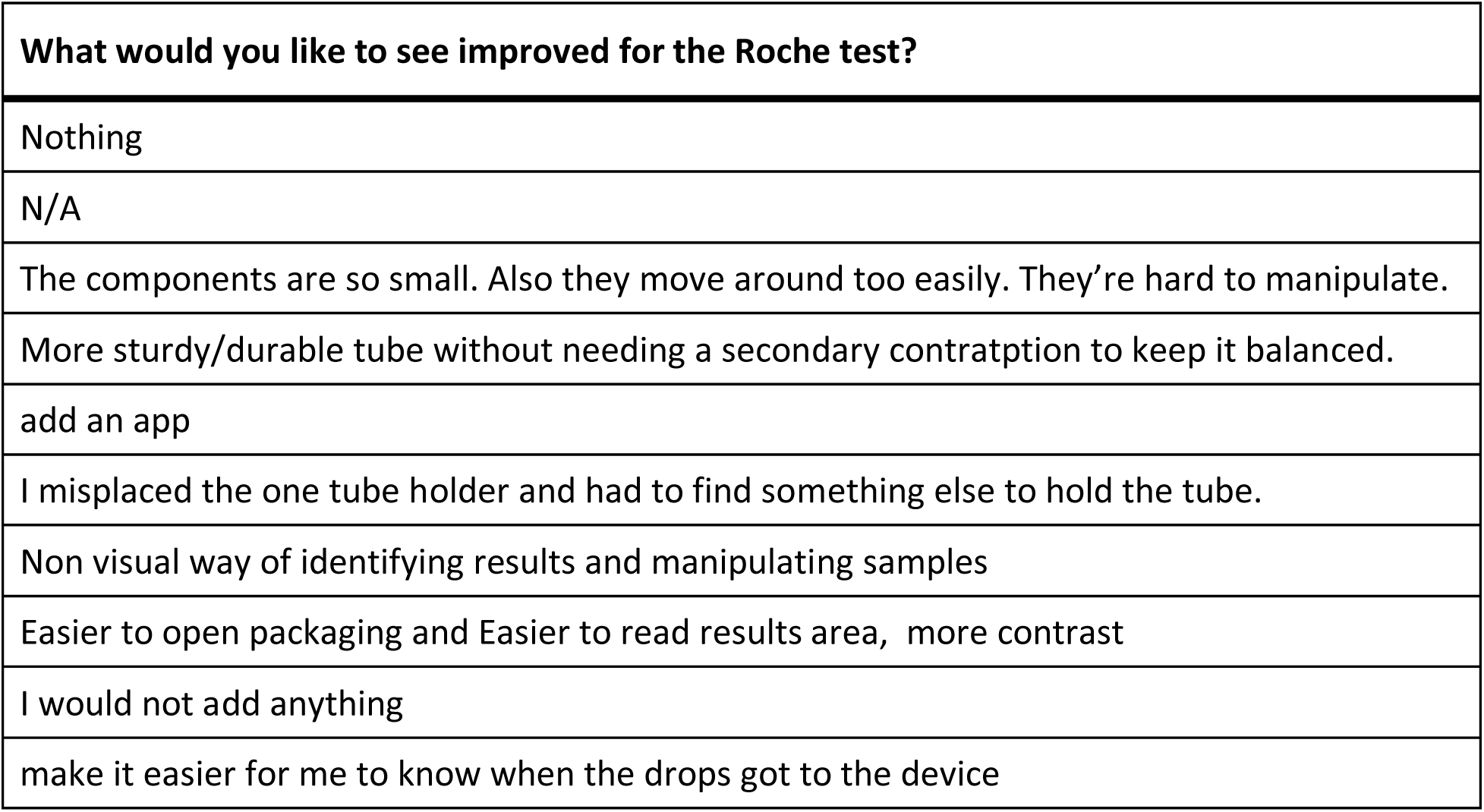
Open response to Q750: “What would you like to see improved for the Roche test?”

### Q770 - InteliSwab Think of the first time you used the InteliSwab test. Did you have difficulty completing any of the following tasks? Select all that apply

**Table 238.**
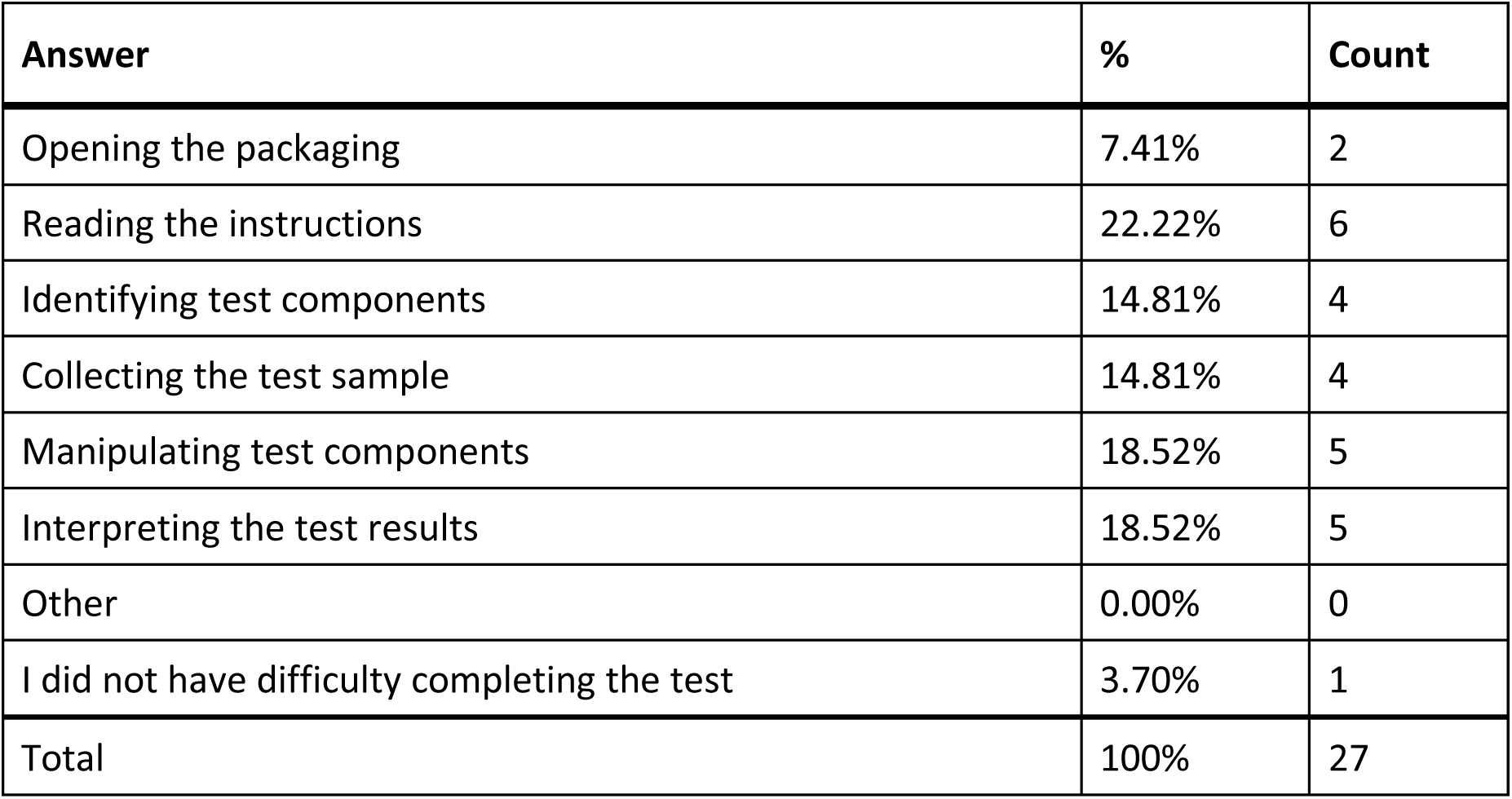
Responses to Q770: “IneliSwab - Did you have difficulty completing any of the following tasks?”

| Answer | % | Count |
| --- | --- | --- |
| Opening the packaging | 7.41% | 2 |
| Reading the instructions | 22.22% | 6 |
| Identifying test components | 14.81% | 4 |
| Collecting the test sample | 14.81% | 4 |
| Manipulating test components | 18.52% | 5 |
| Interpreting the test results | 18.52% | 5 |
| Other | 0.00% | 0 |
| I did not have difficulty completing the test | 3.70% | 1 |
| Total | 100% | 27 |

#### Q770_7_TEXT - Other

None

### Q771 - What tools or assistance, if any, did you use to perform the test?

**Table 239.**
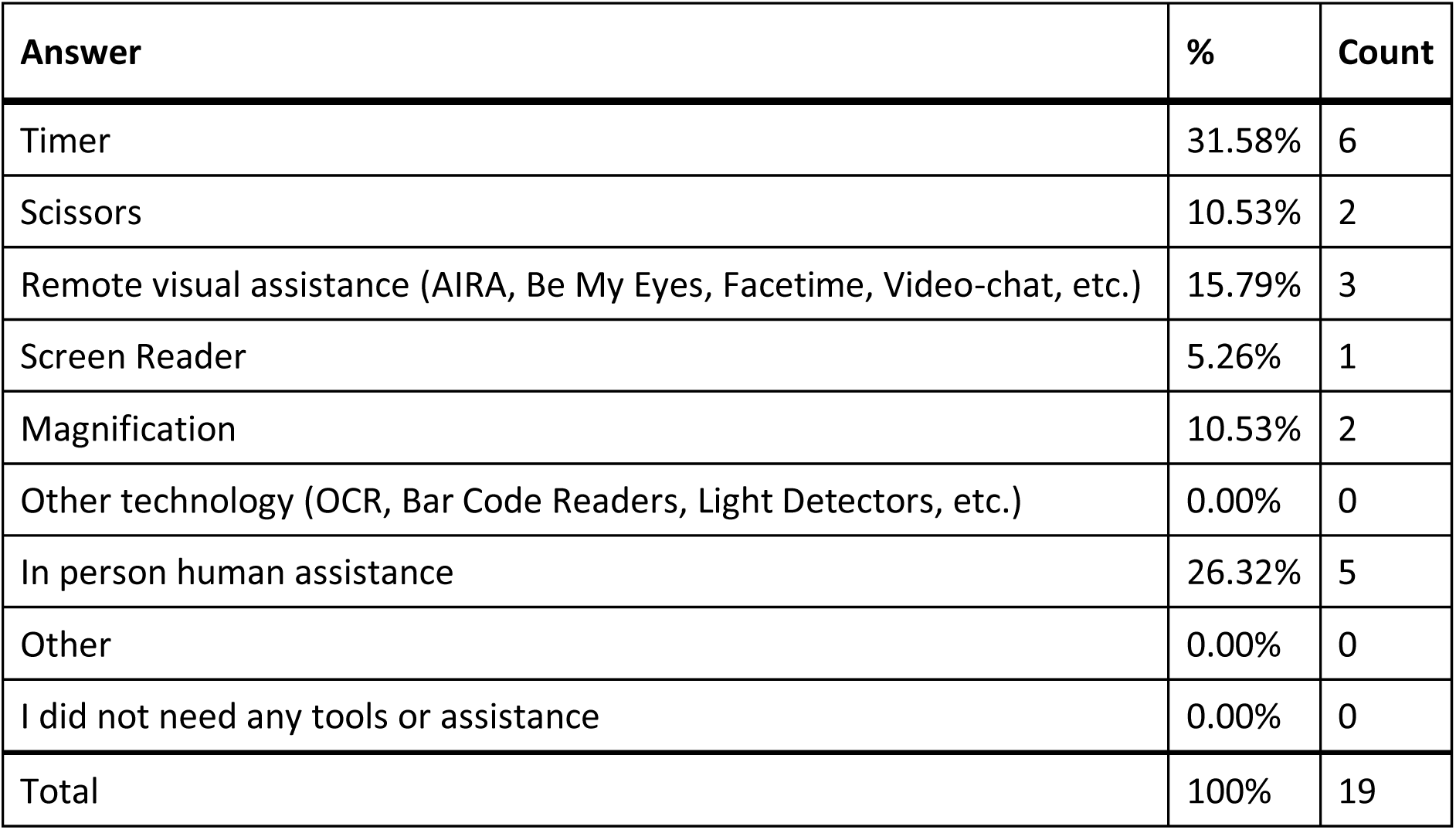
Responses to Q771: “What tools or assistance, if any, did you use to perform the [InteliSwab] test?”

| Answer | % | Count |
| --- | --- | --- |
| Timer | 31.58% | 6 |
| Scissors | 10.53% | 2 |
| Remote visual assistance (AIRA, Be My Eyes, Facetime, Video-chat, etc.) | 15.79% | 3 |
| Screen Reader | 5.26% | 1 |
| Magnification | 10.53% | 2 |
| Other technology (OCR, Bar Code Readers, Light Detectors, etc.) | 0.00% | 0 |
| In person human assistance | 26.32% | 5 |
| Other | 0.00% | 0 |
| I did not need any tools or assistance | 0.00% | 0 |
| Total | 100% | 19 |

#### Q771_8_TEXT - Other

None

### Q772 - Were you able to conduct this test on your first try?

**Table 240.** Stats for Q772: “Were you able to conduct this [InteliSwab] test on your first try?”

| Field | Minimum | Maximum | Mean | Std Deviation | Variance | Count |
| --- | --- | --- | --- | --- | --- | --- |
| Were you able to conduct this test on your first try? | 1.00 | 2.00 | 1.25 | 0.43 | 0.19 | 8 |

**Table 241.** Responses to Q772: “Were you able to conduct this [InteliSwab] test on your first try?”

| Answer | % | Count |
| --- | --- | --- |
| Yes | 75.00% | 6 |
| No | 25.00% | 2 |
| Total | 100% | 8 |

### Q773 - How long did it take to complete the steps required to perform the test, not including the time you spent waiting for results

**Table 242.** Stats for Q773: “How long did it take to complete the steps required to perform the [InteliSwab] test?”

| Field | Minimum | Maximum | Mean | Std Deviation | Variance | Count |
| --- | --- | --- | --- | --- | --- | --- |
| How long did it take to complete the steps required to perform the test, not including the time you spent waiting for results. | 2.00 | 4.00 | 3.13 | 0.78 | 0.61 | 8 |

**Table 243.** Responses to Q773: “How long did it take to complete the steps required to perform the [InteliSwab] test?”

| Answer | % | Count |
| --- | --- | --- |
| Under 5 minutes | 0.00% | 0 |
| 5 - 10 minutes | 25.00% | 2 |
| 10 - 15 minutes | 37.50% | 3 |
| Over 15 minutes | 37.50% | 3 |
| Total | 100% | 8 |

### Q774 - What format of instructions did you use?

**Table 244.** Response to Q774: “What format of [InteliSwab] instructions did you use?”

| Answer | % | Count |
| --- | --- | --- |
| Paper | 45.45% | 5 |
| Digital (webpages, downloaded, etc.) | 18.18% | 2 |
| None of the above | 9.09% | 1 |
| Associated Covid Test App (Smartphone) | 27.27% | 3 |
| Total | 100% | 11 |

### Q775 - Were you able to access electronic information via a QR Code?

**Table 245.** Stats for Q775: “Were you able to access electronic information via a [InteliSwab] QR Code?”

| Field | Minimum | Maximum | Mean | Std Deviation | Variance | Count |
| --- | --- | --- | --- | --- | --- | --- |
| Were you able to access electronic information via a QR Code? | 1.00 | 7.00 | 4.00 | 3.00 | 9.00 | 2 |

**Table 246.**
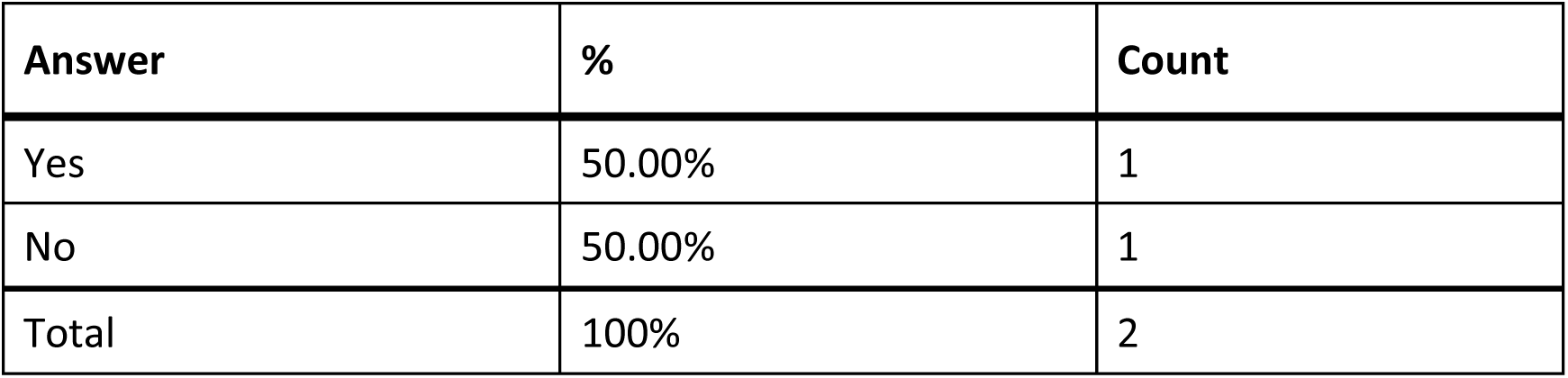
Responses to Q775: “Were you able to access electronic information via a [InteliSwab] QR Code?”

### Q776 - How easy was opening the package of the InteliSwab test for you? (without assistance)

**Table 247.**
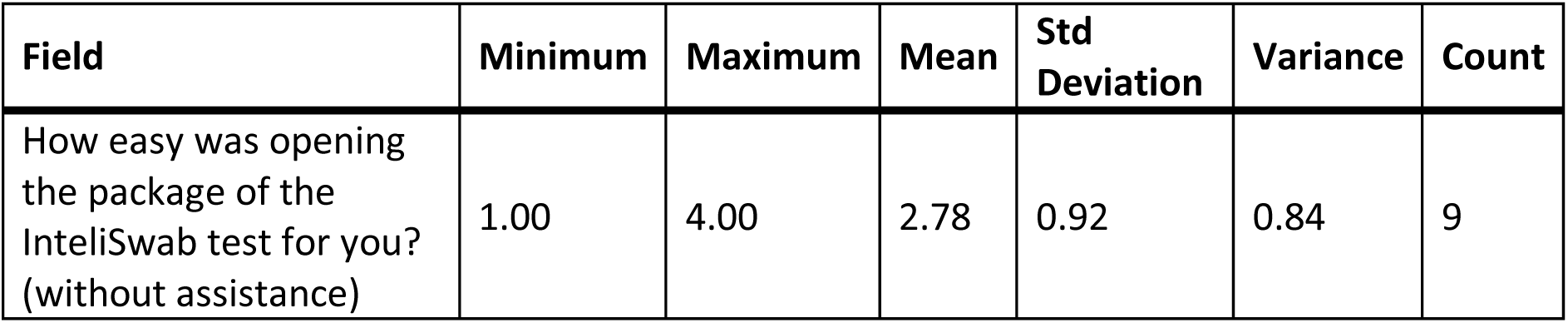
Stats for Q776: “How easy was opening the package of the InteliSwab test for you? (without assistance)”

**Table 248.** Responses to Q776: “How easy was opening the package of the InteliSwab test for you? (without assistance)”

| Answer | % | Count |
| --- | --- | --- |
| Very difficult | 11.11% | 1 |
| Difficult | 22.22% | 2 |
| Neither easy nor difficult | 44.44% | 4 |
| Easy | 22.22% | 2 |
| Very easy | 0.00% | 0 |
| Total | 100% | 9 |

### Q777 - How easy was completing the test protocol for you? (without assistance)

**Table 249.** Stats for Q777: “How easy was completing the [InteliSwab] test protocol for you? (without assistance)”

| Field | Minimum | Maximum | Mean | Std Deviation | Variance | Count |
| --- | --- | --- | --- | --- | --- | --- |
| How easy was completing the test protocol for you? (without assistance) | 1.00 | 3.00 | 1.63 | 0.70 | 0.48 | 8 |

**Table 250.** Responses to Q777: “How easy was completing the [Inteliswab] test protocol for you? (without assistance)”

| Answer | % | Count |
| --- | --- | --- |
| Very difficult | 50.00% | 4 |
| Difficult | 37.50% | 3 |
| Neither easy nor difficult | 12.50% | 1 |
| Easy | 0.00% | 0 |
| Very easy | 0.00% | 0 |
| Total | 100% | 8 |

### Q778 - How easy was interpreting the test results for you? (without assistance)

**Table 251.** Stats for Q778: “How easy was interpreting the [InteliSwab] test results for you? (without assistance)”

| Field | Minimum | Maximum | Mean | Std Deviation | Variance | Count |
| --- | --- | --- | --- | --- | --- | --- |
| How easy was interpreting the test results for you? (without assistance) | 1.00 | 3.00 | 1.75 | 0.83 | 0.69 | 8 |

**Table 252.** Responses to Q778: “How easy was interpreting the [InteliSwab] results for you? (without assistance)”

| Answer | % | Count |
| --- | --- | --- |
| Very difficult | 50.00% | 4 |
| Difficult | 25.00% | 2 |
| Neither easy nor difficult | 25.00% | 2 |
| Easy | 0.00% | 0 |
| Very easy | 0.00% | 0 |
| Total | 100% | 8 |

### Q779 - Were you able to interpret the test results? (without assistance)

**Table 253.** Stats for Q779: “Were you able to interpret the [Inteliswab] test results? (without assistance)”

| Field | Minimum | Maximum | Mean | Std Deviation | Variance | Count |
| --- | --- | --- | --- | --- | --- | --- |
| Were you able to interpret the test results? (without assistance) | 1.00 | 2.00 | 1.63 | 0.48 | 0.23 | 8 |

**Table 254.**
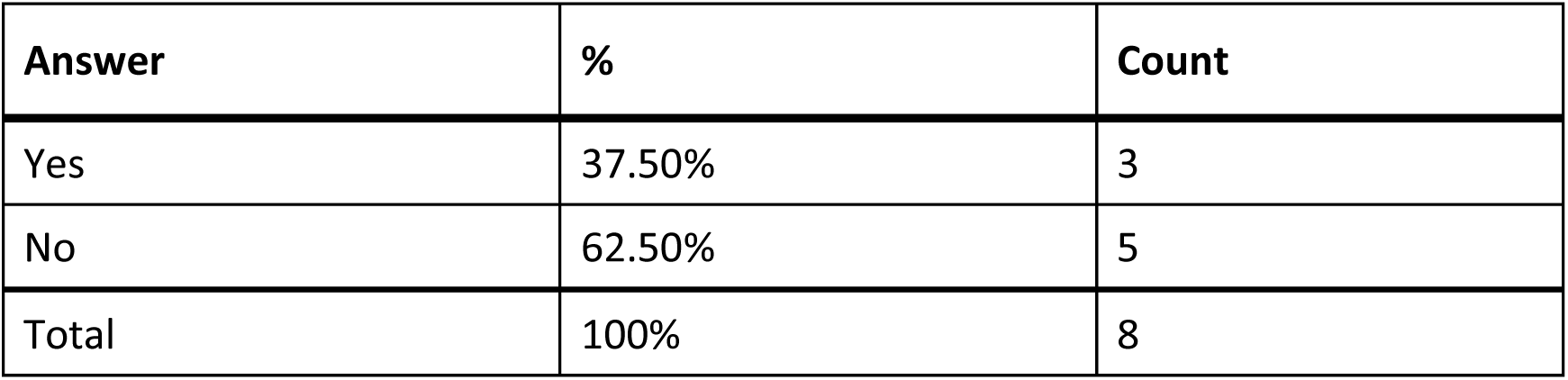
Responses to Q779: “Were you able to interpret the [InteliSwab] test results? (without assistance)”

### Q780 - If there was an associated app for the test, how easy was it for you to use? (without assistance)

**Table 255.** Stats for Q780: “If there was an associated app for the [InteliSwab] test, how easy was it for you to use? (without assistance)”

| Field | Minimum | Maximum | Mean | Std Deviation | Variance | Count |
| --- | --- | --- | --- | --- | --- | --- |
| If there was an associated app for the test, how easy was it for you to use? (without assistance) | 1.00 | 3.00 | 2.00 | 0.82 | 0.67 | 3 |

**Table 256.**
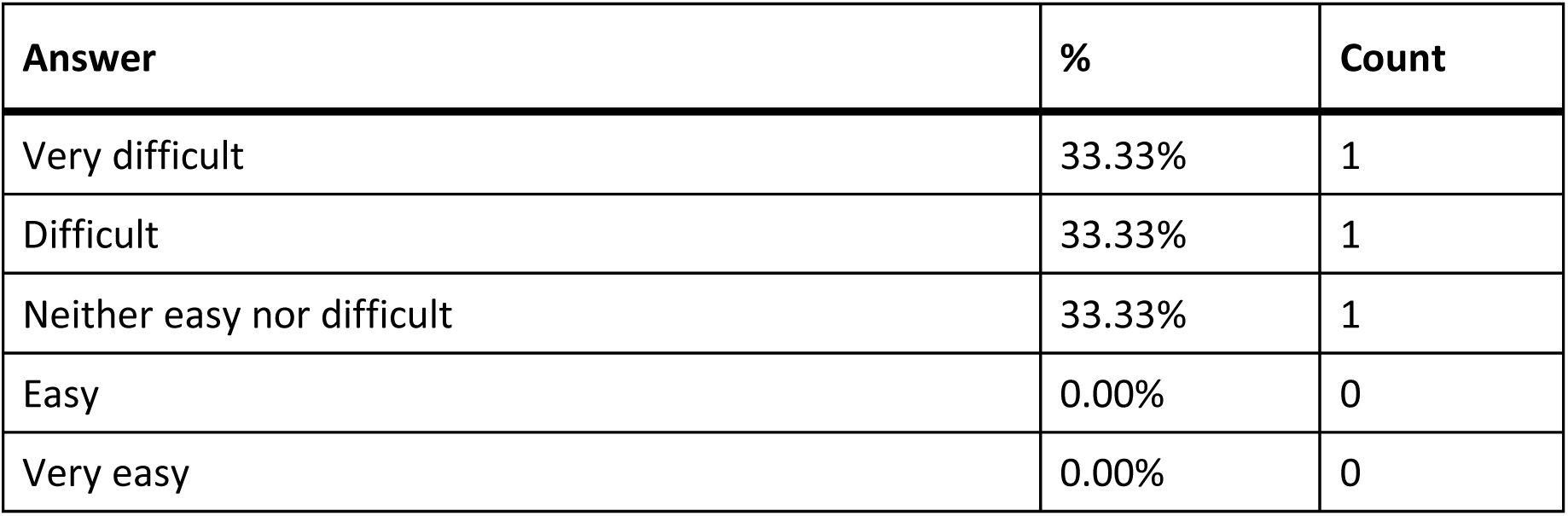
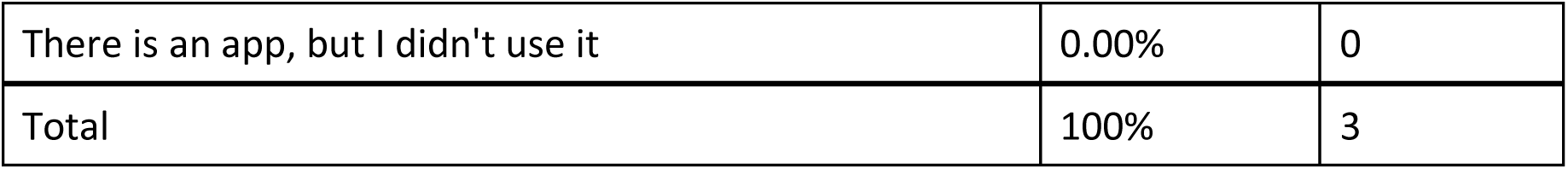
Responses to Q780: “If there was an associated app for the [InteliSwab] test, how easy was it for you to use? (without assistance)?”

| Answer | % | Count |
| --- | --- | --- |
| Very difficult | 33.33% | 1 |
| Difficult | 33.33% | 1 |
| Neither easy nor difficult | 33.33% | 1 |
| Easy | 0.00% | 0 |
| Very easy | 0.00% | 0 |
| There is an app, but I didn't use it | 0.00% | 0 |
| Total | 100% | 3 |

### Q781 - How helpful was the app in assisting you with completing the test?

**Table 257.** Stats for Q781: “How helpful was the app in assisting you with completing the [InteliSwab] test?”

| Field | Minimum | Maximum | Mean | Std Deviation | Variance | Count |
| --- | --- | --- | --- | --- | --- | --- |
| How helpful was the app in assisting you with completing the test? | 2.00 | 4.00 | 3.00 | 0.82 | 0.67 | 3 |

**Table 258.**
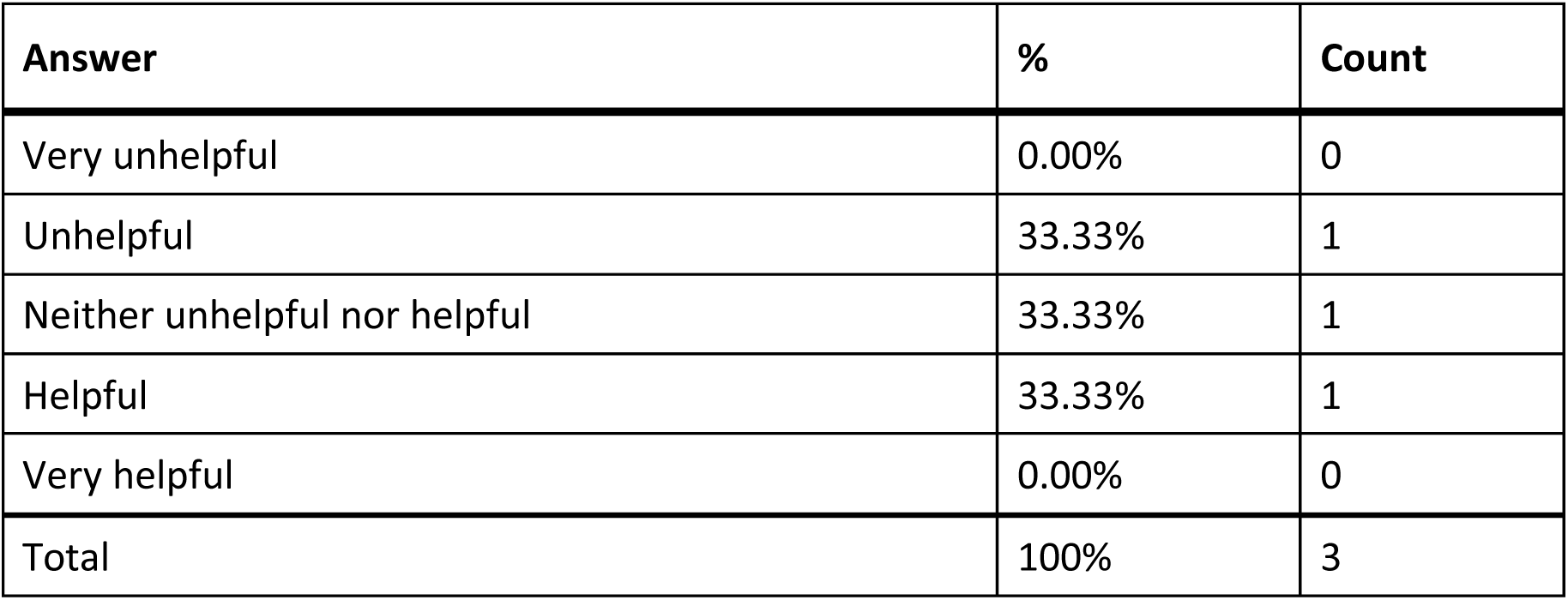
Responses to Q781: “How helpful was the app in assisting you with completing the [InteliSwab] test?”

| Answer | % | Count |
| --- | --- | --- |
| Very unhelpful | 0.00% | 0 |
| Unhelpful | 33.33% | 1 |
| Neither unhelpful nor helpful | 33.33% | 1 |
| Helpful | 33.33% | 1 |
| Very helpful | 0.00% | 0 |
| Total | 100% | 3 |

### Q782 - Did you receive a valid test result with the InteliSwab test (positive or negative)?

**Table 259.**
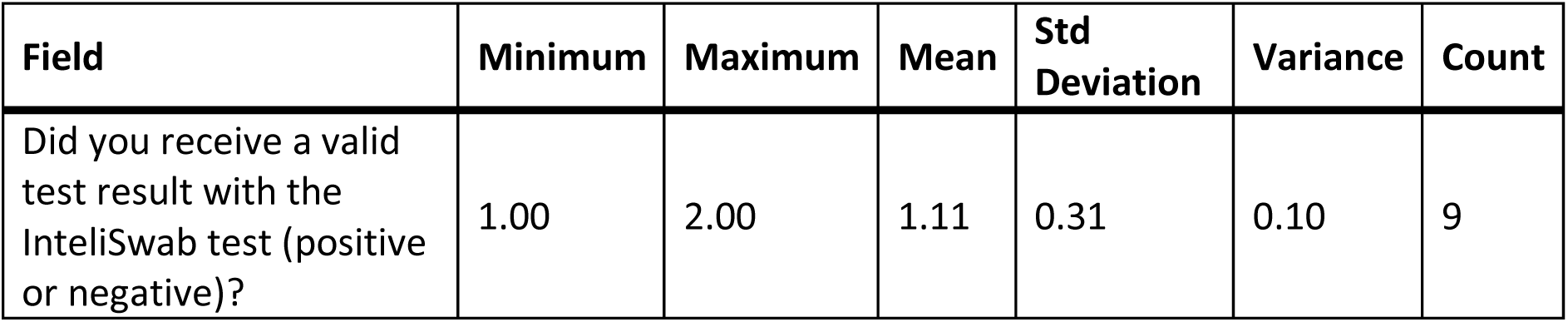
Stats for Q782: “Did you receive a valid test result with the InteliSwab test (positive or negative)?”

| Field | Minimum | Maximum | Mean | Std Deviation | Variance | Count |
| --- | --- | --- | --- | --- | --- | --- |
| Did you receive a valid test result with the InteliSwab test (positive or negative)? | 1.00 | 2.00 | 1.11 | 0.31 | 0.10 | 9 |

**Table 260.** Responses to Q782: “Did you receive a valid test result with the InteliSwab test (positive or negative)?”

| Answer | % | Count |
| --- | --- | --- |
| Yes | 88.89% | 8 |
| No | 11.11% | 1 |
| Total | 100% | 9 |

### Q783 - Did you attempt to contact InteliSwab’s customer support?

**Table 261.**
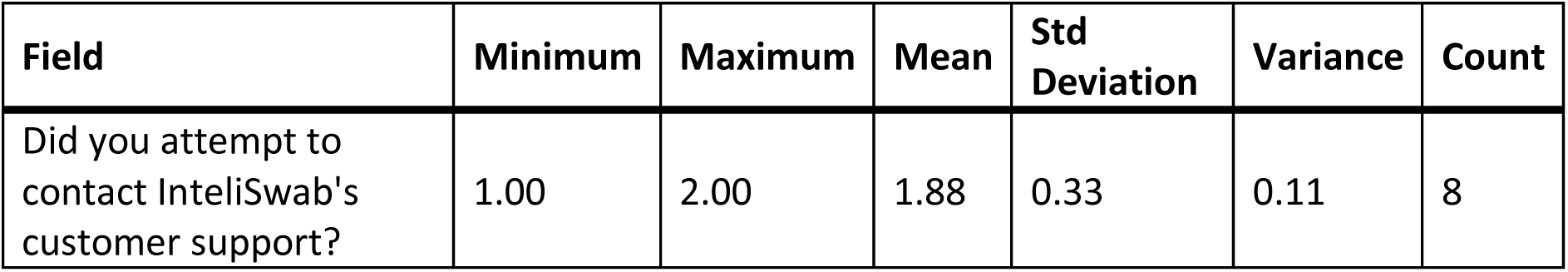
Stats for Q783: “Did you attempt to contact InteliSwab’s customer support?”

**Table 262.**
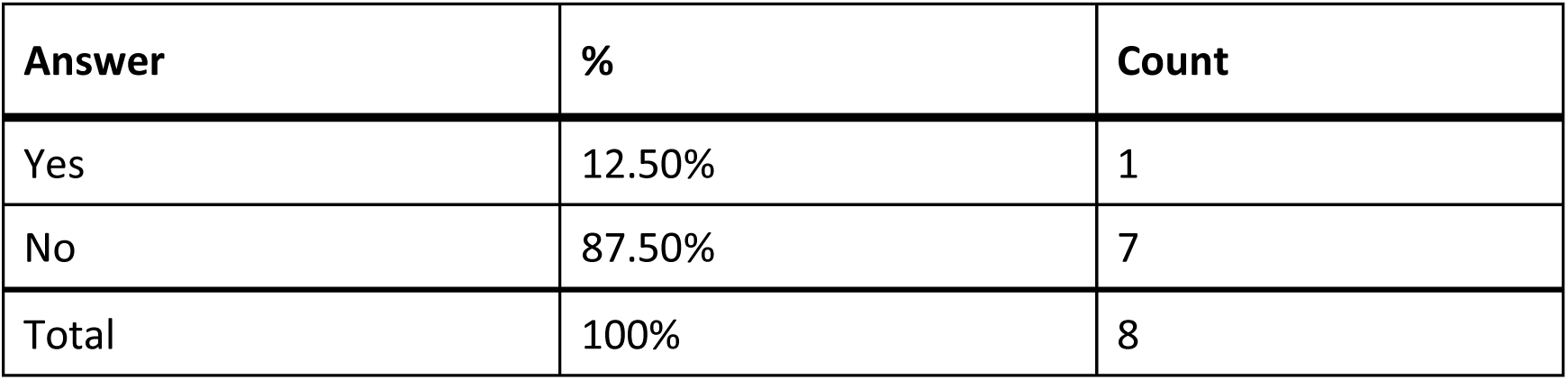
Responses to Q783: “Did you attempt to contact InteliSwab’s customer support?”

| Answer | % | Count |
| --- | --- | --- |
| Yes | 12.50% | 1 |
| No | 87.50% | 7 |
| Total | 100% | 8 |

### Q784 - Were you able to talk with customer support?

**Table 263.**
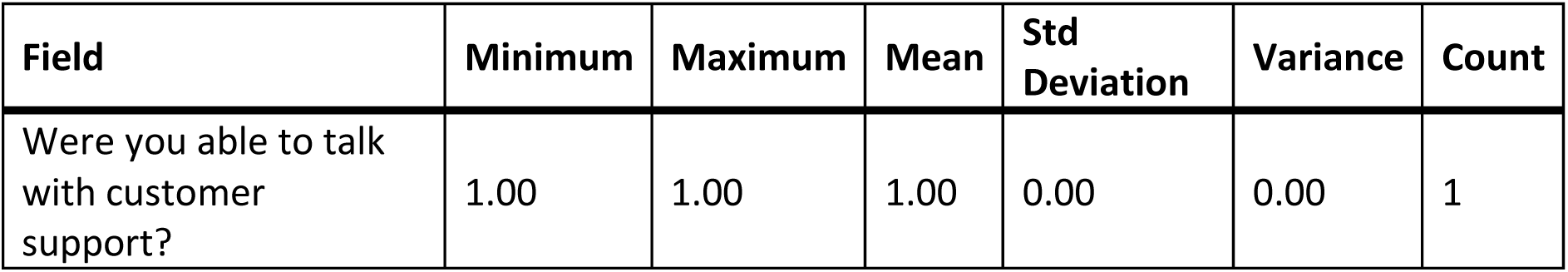
Stats for Q784: “Were you able to talk with [InteliSwab’s] customer support?”

**Table 264.** Responses to Q784: “Were you able to talk with [InteliSwab’s] customer support?”

| Answer | % | Count |
| --- | --- | --- |
| Yes | 100.00% | 1 |
| No | 0.00% | 0 |
| Total | 100% | 1 |

### Q785 - Was customer support helpful in answering your question(s)?

**Table 265.** Stats for Q785: “Was [InteliSwab] customer support helpful in answering your question(s)?”

| Field | Minimum | Maximum | Mean | Std Deviation | Variance | Count |
| --- | --- | --- | --- | --- | --- | --- |
| Was customer support helpful in answering your question(s)? | 4.00 | 4.00 | 4.00 | 0.00 | 0.00 | 1 |

**Table 266.** Responses to Q785: “Was [InteliSwab] customer support helpful in answering your question(s)?”

| Answer | % | Count |
| --- | --- | --- |
| Very unhelpful | 0.00% | 0 |
| Unhelpful | 100.00% | 1 |
| Neither helpful nor unhelpful | 0.00% | 0 |
| Helpful | 0.00% | 0 |
| Very helpful | 0.00% | 0 |
| Total | 100% | 1 |

### Q786 - If you have performed the InteliSwab test multiple times, how did your experience change?

**Table 267.** Stats for Q786: “If you have performed the InteliSwab test multiple times, how did your experience change?”

| Field | Minimum | Maximum | Mean | Std Deviation | Variance | Count |
| --- | --- | --- | --- | --- | --- | --- |
| If you have performed the InteliSwab test multiple times, how did your experience change? | 3.00 | 4.00 | 3.20 | 0.40 | 0.16 | 5 |

**Table 268.** Responses to Q786: “If you have performed the InteliSwab test multiple times, how did your experience change?”

| Answer | % | Count |
| --- | --- | --- |
| Much worse | 0.00% | 0 |
| Worse | 0.00% | 0 |
| Neither better nor worse | 80.00% | 4 |
| Better | 20.00% | 1 |
| Much better | 0.00% | 0 |
| Total | 100% | 5 |

### Q787 - What did you like about the InteliSwab test?

**Table 269.** Open response to Q787: “What did you like about the InteliSwab test?”

| What did you like about the IntelliSwab test? |
| --- |
| N/A |
| With repetition mastery gets accomplished |
| It was easy to use with remote assistance. |
| N/A |
| I don't know. A sighted person did all the work. |

### Q788 - What would you like to see improved for the InteliSwab test?

**Table 270.** Open responses to Q788: “What would you like to see improved for the InteliSwab test?”

| What would you like to see improved for the IntelliSwab test? |
| --- |
| N/A |
| Not rely on services such as AIRA to read out the results etc. |
| It would be interesting to try an app that could read the instructions and results. |
| phone app that is fully accessible using voiceover |

### Q789 - On/Go Think of the first time you used the On/Go test. Did you have difficulty completing any of the following tasks? Select all that apply

**Table 271.** Responses to Q789: “On/Go - Did you have any difficulty completing any of the following tasks?”

| Answer | % | Count |
| --- | --- | --- |
| Opening the packaging | 3.13% | 1 |
| Reading the instructions | 12.50% | 4 |
| Identifying test components | 18.75% | 6 |
| Collecting the test sample | 12.50% | 4 |
| Manipulating test components | 18.75% | 6 |
| Interpreting the test results | 21.88% | 7 |
| Other | 6.25% | 2 |
| I did not have difficulty completing the test | 6.25% | 2 |
| Total | 100% | 32 |

#### Q789_7_TEXT – Other

**Table 272.** Open response to Other - Q789: “On/Go - Did you have any difficulty completing any of the following tasks?”

| Other - Text |
| --- |
| Very little audio. You had to scan the box to start the test. I did not start any test. |
| my roommate did it |

### Q790 - What tools or assistance, if any, did you use to perform the test?

**Table 273.** Responses to Q790: “What tools of assistance, if any, did you use to perform the [On/Go] test?”

| Answer | % | Count |
| --- | --- | --- |
| Timer | 13.64% | 3 |
| Scissors | 4.55% | 1 |
| Remote visual assistance (AIRA, Be My Eyes, Facetime, Video-chat, etc.) | 13.64% | 3 |
| Screen Reader | 13.64% | 3 |
| Magnification | 9.09% | 2 |
| Other technology (OCR, Bar Code Readers, Light Detectors, etc.) | 13.64% | 3 |
| In person human assistance | 27.27% | 6 |
| Other | 0.00% | 0 |
| I did not need any tools or assistance | 4.55% | 1 |
| Total | 100% | 22 |

#### Q790_8_TEXT - Other

None

### Q791 - Were you able to conduct this test on your first try?

**Table 274.** Stats Q791: “Were you able to conduct this [On/Go] test on your first try?”

| Field | Minimum | Maximum | Mean | Std Deviation | Variance | Count |
| --- | --- | --- | --- | --- | --- | --- |
| Were you able to conduct this test on your first try? | 1.00 | 2.00 | 1.44 | 0.50 | 0.25 | 9 |

**Table 275.** Responses to Q791: “Were you able to conduct this [On/Go] test on your first try?”

| Answer | % | Count |
| --- | --- | --- |
| Yes | 55.56% | 5 |
| No | 44.44% | 4 |
| Total | 100% | 9 |

### Q792 - How long did it take to complete the steps required to perform the test, not including the time you spent waiting for results

**Table 276.** Stats for Q792: “How long did it take to complete the steps required to perform the [On/Go] test?”

| Field | Minimum | Maximum | Mean | Std Deviation | Variance | Count |
| --- | --- | --- | --- | --- | --- | --- |
| How long did it take to complete the steps required to perform the test, not including the time you spent waiting for results. | 2.00 | 4.00 | 2.78 | 0.92 | 0.84 | 9 |

**Table 277.** Responses to Q792: “How long did it take to complete the steps required to perform the [On/Go] test?”

| Answer | % | Count |
| --- | --- | --- |
| Under 5 minutes | 0.00% | 0 |
| 5 - 10 minutes | 55.56% | 5 |
| 10 - 15 minutes | 11.11% | 1 |
| Over 15 minutes | 33.33% | 3 |
| Total | 100% | 9 |

### Q793 - What format of instructions did you use?

**Table 278.** Responses to Q793: “What format of [On/Go] instructions did you use?”

| Answer | % | Count |
| --- | --- | --- |
| Paper | 40.00% | 4 |
| Digital (webpages, downloaded, etc.) | 10.00% | 1 |
| None of the above | 30.00% | 3 |
| Associated Covid Test App (Smartphone) | 20.00% | 2 |
| Total | 100% | 10 |

### Q794 - Were you able to access electronic information via a QR Code?

**Table 279.** Stats for Q794: “Were you able to access electronic information via a [On/Go] QR Code?”

| Field | Minimum | Maximum | Mean | Std Deviation | Variance | Count |
| --- | --- | --- | --- | --- | --- | --- |
| Were you able to access electronic information via a QR Code? | 7.00 | 7.00 | 7.00 | 0.00 | 0.00 | 2 |

**Table 280.**
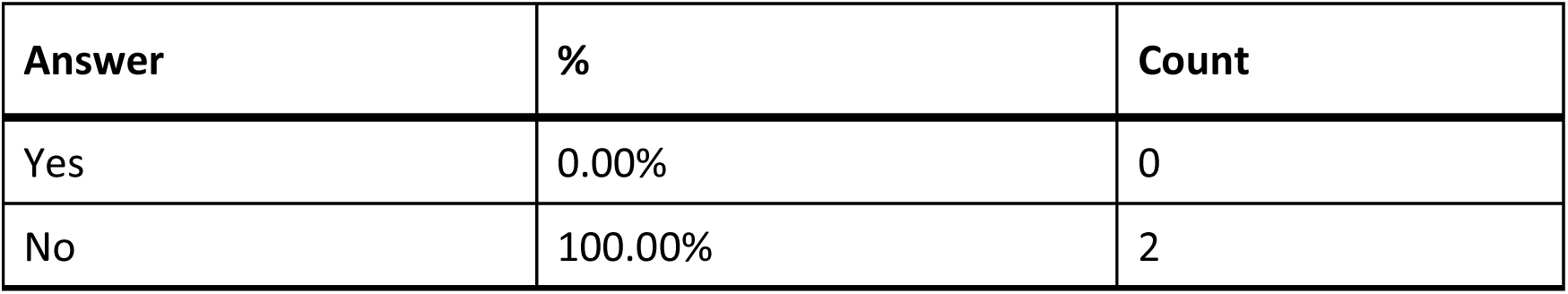
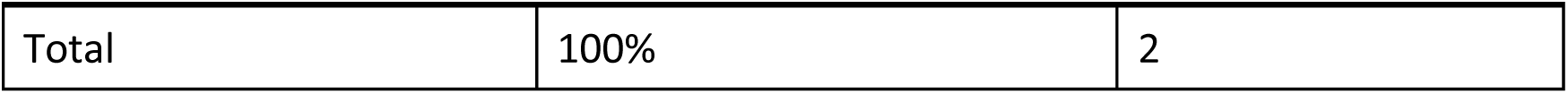
Responses to Q794: “Were you able to access electonic information via a [On/Go] QR Code?”

| Answer | % | Count |
| --- | --- | --- |
| Yes | 0.00% | 0 |
| No | 100.00% | 2 |
| Total | 100% | 2 |

### Q795 - How easy was opening the package of the On/Go test for you? (without assistance)

**Table 281.** Stats for Q795: “How easy was opening the package of the On/Go test for you? (without assistance)”

| Field | Minimum | Maximum | Mean | Std Deviation | Variance | Count |
| --- | --- | --- | --- | --- | --- | --- |
| How easy was opening the package of the On/Go test for you? (without assistance) | 1.00 | 5.00 | 3.22 | 1.40 | 1.95 | 9 |

**Table 282.** Responses to Q795: “How easy was opening the package of the On/Go test for you? (without assistance)”

| Answer | % | Count |
| --- | --- | --- |
| Very difficult | 22.22% | 2 |
| Difficult | 0.00% | 0 |
| Neither easy nor difficult | 33.33% | 3 |
| Easy | 22.22% | 2 |
| Very easy | 22.22% | 2 |
| Total | 100% | 9 |

### Q796 - How easy was completing the test protocol for you? (without assistance)

**Table 283.** Stats for Q796: “How easy was completing the [On/Go] test protocol for you? (without assistance)”

| Field | Minimum | Maximum | Mean | Std Deviation | Variance | Count |
| --- | --- | --- | --- | --- | --- | --- |
| How easy was completing the test protocol for you? (without assistance) | 1.00 | 4.00 | 2.11 | 1.37 | 1.88 | 9 |

**Table 284.** Responses to Q796: “How easy was completing the [On/Go] test protocol for you? (without assistance)”

| Answer | % | Count |
| --- | --- | --- |
| Very difficult | 55.56% | 5 |
| Difficult | 11.11% | 1 |
| Neither easy nor difficult | 0.00% | 0 |
| Easy | 33.33% | 3 |
| Very easy | 0.00% | 0 |
| Total | 100% | 9 |

### Q797 - How easy was interpreting the test results for you? (without assistance)

**Table 285.** Stats for Q797: “How easy was interpreting the [On/Go] test results for you? (without assistance)”

| Field | Minimum | Maximum | Mean | Std Deviation | Variance | Count |
| --- | --- | --- | --- | --- | --- | --- |
| How easy was interpreting the test results for you? (without assistance) | 1.00 | 5.00 | 1.89 | 1.45 | 2.10 | 9 |

**Table 286.** Responses to Q797: “How easy was interpreting the [On/Go] test results for you? (without assistance)”

| Answer | % | Count |
| --- | --- | --- |
| Very difficult | 66.67% | 6 |
| Difficult | 11.11% | 1 |
| Neither easy nor difficult | 0.00% | 0 |
| Easy | 11.11% | 1 |
| Very easy | 11.11% | 1 |
| Total | 100% | 9 |

### Q798 - Were you able to interpret the test results? (without assistance)

**Table 287.** Stats for Q798: “Were you able to interpret the [On/Go] test results? (without assistance)”

| Field | Minimum | Maximum | Mean | Std Deviation | Variance | Count |
| --- | --- | --- | --- | --- | --- | --- |
| Were you able to interpret the test results? (without assistance) | 1.00 | 2.00 | 1.78 | 0.42 | 0.17 | 9 |

**Table 288.** Responses to Q798: “Were you able to interpret the [On/Go] test results? (without assistance)”

| Answer | % | Count |
| --- | --- | --- |
| Yes | 22.22% | 2 |
| No | 77.78% | 7 |
| Total | 100% | 9 |

### Q799 - If there was an associated app for the test, how easy was it for you to use? (without assistance)

**Table 289.** Stats for Q799: “If there was an associated app for the [On/Go] test, how easy was it for you to use? (without assistance)”

| Field | Minimum | Maximum | Mean | Std Deviation | Variance | Count |
| --- | --- | --- | --- | --- | --- | --- |
| If there was an associated app for the test, how easy was it for you to use? (without assistance) | 1.00 | 7.00 | 2.80 | 2.23 | 4.96 | 5 |

**Table 290.** Responses to Q799: “If there was an associated app for the [On/Go] test, how easy was it for you to use? (without assistance)”

| Answer | % | Count |
| --- | --- | --- |
| Very difficult | 40.00% | 2 |
| Difficult | 20.00% | 1 |
| Neither easy nor difficult | 20.00% | 1 |
| Easy | 0.00% | 0 |
| Very easy | 0.00% | 0 |
| There is an app, but I didn't use it | 20.00% | 1 |
| Total | 100% | 5 |

### Q800 - How helpful was the app in assisting you with completing the test?

**Table 291.** Stats for Q800: “How helpful was the [On/Go] app in assisting you with completing the test?”

| Field | Minimum | Maximum | Mean | Std Deviation | Variance | Count |
| --- | --- | --- | --- | --- | --- | --- |
| How helpful was the app in assisting you with completing the test? | 1.00 | 3.00 | 1.50 | 0.87 | 0.75 | 4 |

**Table 292.** Responses to Q800: “How helpful was the [On/Go] app in assisting you with completing the test?”

| Answer | % | Count |
| --- | --- | --- |
| Very unhelpful | 75.00% | 3 |
| Unhelpful | 0.00% | 0 |
| Neither unhelpful nor helpful | 25.00% | 1 |
| Helpful | 0.00% | 0 |
| Very helpful | 0.00% | 0 |
| Total | 100% | 4 |

### Q801 - Did you receive a valid test result with the On/Go test (positive or negative)?

**Table 293.** Stats for Q801: “Did you receive a valid test result with the On/Go test (positive or negative)?”

| Field | Minimum | Maximum | Mean | Std Deviation | Variance | Count |
| --- | --- | --- | --- | --- | --- | --- |
| Did you receive a valid test result with the On/Go test (positive or negative)? | 1.00 | 2.00 | 1.40 | 0.49 | 0.24 | 10 |

**Table 294.** Responses to Q801: “Did you receive a valid test result with the On/Go test (positive or negative)?”

| Answer | % | Count |
| --- | --- | --- |
| Yes | 60.00% | 6 |
| No | 40.00% | 4 |
| Total | 100% | 10 |

### Q802 - Did you attempt to contact On/Go’s customer support?

**Table 295.** Stats for Q802: “Did you attempt to contact On/Go’s customer support?”

| Field | Minimum | Maximum | Mean | Std Deviation | Variance | Count |
| --- | --- | --- | --- | --- | --- | --- |
| Did you attempt to contact On/Go's customer support? | 1.00 | 2.00 | 1.90 | 0.30 | 0.09 | 10 |

**Table 296.** Responses to Q802: “Did you attempt to contact On/Go’s customer support?”

| Answer | % | Count |
| --- | --- | --- |
| Yes | 10.00% | 1 |
| No | 90.00% | 9 |
| Total | 100% | 10 |

### Q803 - Were you able to talk with customer support?

**Table 297.** Stats for Q803: “Were you able to talk with [On/Go] customer support?”

| Field | Minimum | Maximum | Mean | Std Deviation | Variance | Count |
| --- | --- | --- | --- | --- | --- | --- |
| Were you able to talk with customer support? | 2.00 | 2.00 | 2.00 | 0.00 | 0.00 | 1 |

**Table 298.** Responses to Q803: “Were you able to talk with [On/Go] customer support?”

| Answer | % | Count |
| --- | --- | --- |
| Yes | 0.00% | 0 |
| No | 100.00% | 1 |
| Total | 100% | 1 |

### Q804 - Was customer support helpful in answering your question(s)?

**Table 299.** Stats for Q804: “Was [On/Going] customer support helpful in answering your question(s)?”

| Field | Minimum | Maximum | Mean | Std Deviation | Variance | Count |
| --- | --- | --- | --- | --- | --- | --- |
| Was customer support helpful in answering your question(s)? | 0.00 | 0.00 | 0.00 | 0.00 | 0.00 | 0 |

**Table 300.**
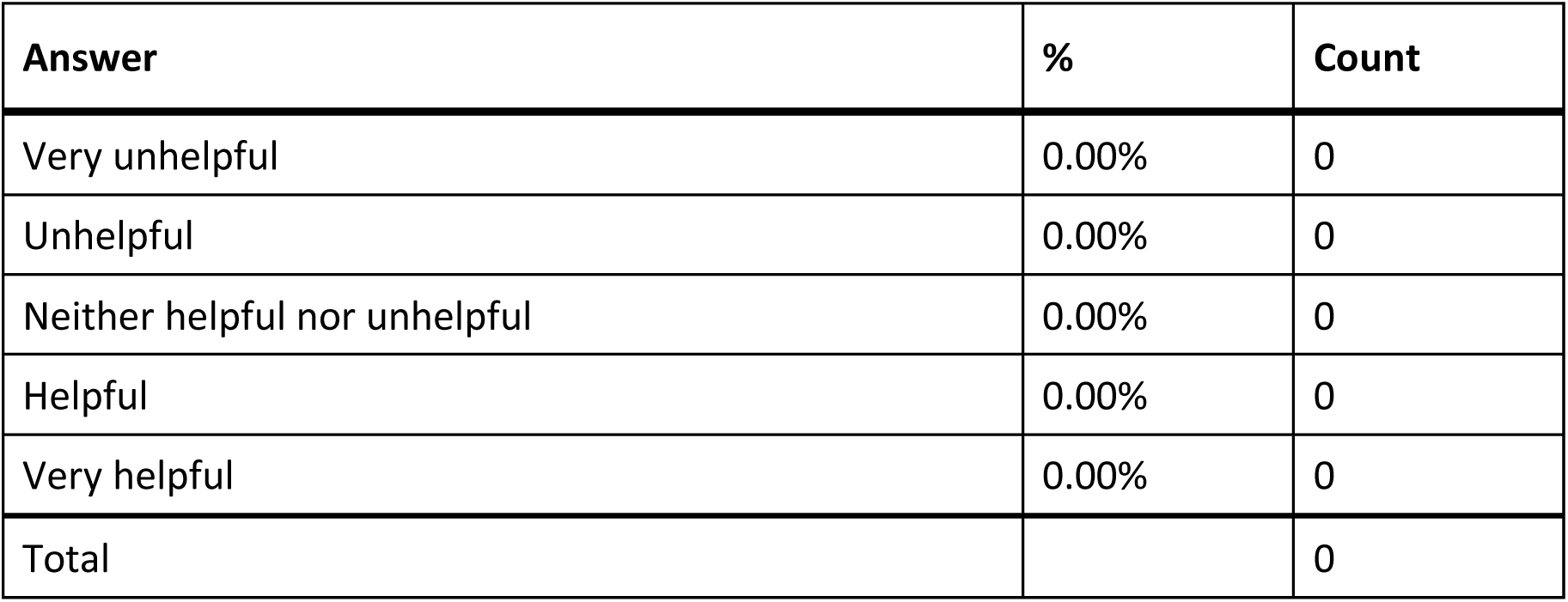
Responses to Q804: “Was [On/Going] customer support helpful in answering your question(s)?”

### Q805 - If you have performed the On/Go test multiple times, how did your experience change?

**Table 301.** Stats for Q805: “If you have performed the On/Go test multiple times, how did your experience change?”

| Field | Minimum | Maximum | Mean | Std Deviation | Variance | Count |
| --- | --- | --- | --- | --- | --- | --- |
| If you have performed the On/Go test multiple times, how did your experience change? | 1.00 | 5.00 | 2.67 | 1.70 | 2.89 | 3 |

**Table 302.**
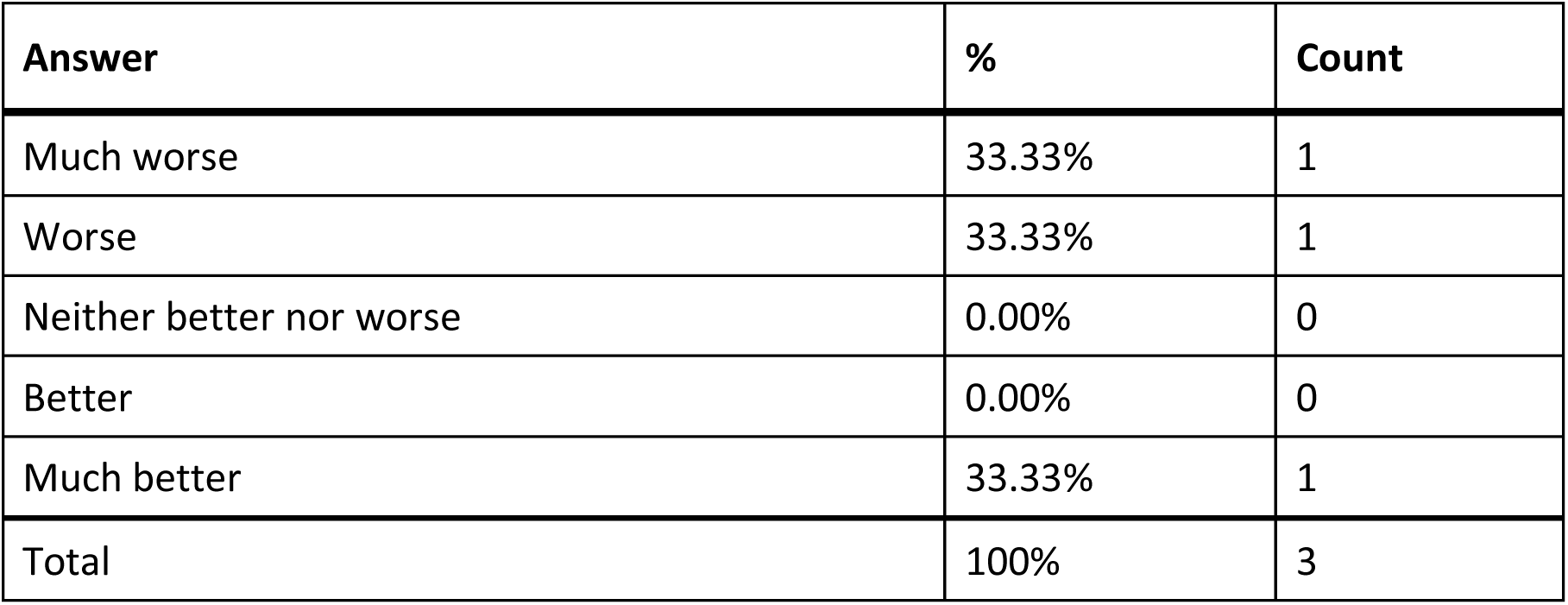
Responses to Q805: “If you have performed the On/Go test multiple times, how did you experience change?”

| Answer | % | Count |
| --- | --- | --- |
| Much worse | 33.33% | 1 |
| Worse | 33.33% | 1 |
| Neither better nor worse | 0.00% | 0 |
| Better | 0.00% | 0 |
| Much better | 33.33% | 1 |
| Total | 100% | 3 |

### Q806 - What did you like about the On/Go test?

**Table 303.** Open responses to Q806: “What did you like about the On/Go test?”

| What did you like about the On/Go test? |
| --- |
| Fast results & easy to read |
| Visually fabrication of test home covid test design |
| Nothing |
| my roommate did it with me |
| I just like the fact that I could do the COVID-19 test on my own |
| Nothing, it was very difficult to use. |
| Nothing |
| Not much |

### Q807 - What would you like to see improved for the On/Go test?

**Table 304.** Open responses to Q807: “What would you like to see improved for the On/Go test?”

| What would you like to see improved for the On/Go test? |
| --- |
| Less steps |
| Accessibility materials and components to understand home COVID-19 test kit and gear belonging to test kit. |
| More accessibility with screen readers |
| Associated app, more straightforward, testing components to be identifiable in a better way |
| The application of the fluid into the tray. I could not get the testing liquid into the slot in order to get an accurate result. Both attempts showed inconclusive results. When I had to ultimately go for an in person test, I did have Covid. |
| It wasn't accessible and I couldn't complete the test. |
| Clearer colors |

### Q808 - Ellume Think of the first time you used the Ellume test. Did you have difficulty completing any of the following tasks? Select all that apply

**Table 305.** Responses to Q808: “Ellume - Did you have difficulty completing any of the following tasks?”

| Answer | % | Count |
| --- | --- | --- |
| Opening the packaging | 6.51% | 19 |
| Reading the instructions | 16.78% | 49 |
| Identifying test components | 21.58% | 63 |
| Collecting the test sample | 9.93% | 29 |
| Manipulating test components | 19.86% | 58 |
| Interpreting the test results | 8.22% | 24 |
| Other | 8.90% | 26 |
| I did not have difficulty completing the test | 8.22% | 24 |
| Total | 100% | 292 |

#### Q808_7_TEXT – Other

**Table 306.** Open response to Other - Q808: “Ellume - Did you have difficulty completing any of the following tasks?”

| Other - Text |
| --- |
| I had help from my sister; the only thing I did independently was to swab my nose |
| The app was not fully accessible. Needed sighted help. |
| Measuring the sample fluid to be analyzed after the nasal swab was screwed into the beaker. |
| Initially, I had some difficulty pairing between the app and the test |
| I had to get a sighted person to tell me what app I needed to download. |
| App had several unlabeled buttons, video was not clear for blind users, app crashed a few times and had to get new tests |
| Connecting it to my bluetooth |
| Having to figure out how to watch the video and when I called the Help Desk, it was closed for the weekend! Highly frustrating. |
| Getting liquid in the right place. |
| Test expired with no results. I did not understand where to put the sample until too late. |
| I thought the fluid goes directly into the analyzer port, which invalidated my test. |
| The IOS App had several problems. For example, some buttons did not have correct labels and VoiceOver did not read the screen on its own. |
| I couldn't get the test analyzer to pair with my iPhone. |
| The app is clunky and isn't designed from an accessibility point of view. I had to seek assistance from a sided friend. It took us more than 20 minutes and I didn't get the result at the end of the test. |
| Blinking light |
| Knowing if the LED light was blinking slow, fast or solid, and adding 5 drops of sample to analyzer |
| Did not succeed in getting test results |
| We did not open the top, nor put the drops in the reader. |
| Using the associated smartphone app and getting the test module to pair with my smartphone. |
| Getting started with the app and setting it up was a bit involved, and it still isn't clear if you have to go through the set-up every time you take an Ellume test. |
| I had difficulty determining how many drops were being squeezed out of the tube onto the testing component. |
| There were no verbal descriptions of the components. Thus you had to guess what the complaints were. |
| Knowing that the test strip was completely connected to the Bluetooth on my phone. It has a flashing light that was very hard for me to see and there was no audio Q when it completely connected. I did eventually get a message that I had to magnify on my phone to be able to read |
| figuring out the unlabelled button in the app and knowing which fields were mandatory |
| the bluetooth receiver did not beep or give any tactile indication like vibration that it was on or connected. |

### Q809 - What tools or assistance, if any, did you use to perform the test?

**Table 307.** Responses to Q809: “What tools or assistance, if any, did you use to perform the [Ellume] test?”

| Answer | % | Count |
| --- | --- | --- |
| Timer | 14.05% | 34 |
| Scissors | 15.29% | 37 |
| Remote visual assistance (AIRA, Be My Eyes, Facetime, Video-chat, etc.) | 10.74% | 26 |
| Screen Reader | 28.51% | 69 |
| Magnification | 5.79% | 14 |
| Other technology (OCR, Bar Code Readers, Light Detectors, etc.) | 2.48% | 6 |
| In person human assistance | 14.46% | 35 |
| Other | 4.13% | 10 |
| I did not need any tools or assistance | 4.55% | 11 |
| Total | 100% | 242 |

#### Q809_8_TEXT – Other

**Table 308.** Open responses to Other - Q809: “What tools or assistance, if any, did you use to perform the [Ellume] test?”

| Other - Text |
| --- |
| Used workarounds |
| phone assistance from a fellow blind person |
| I had to look up the instructions and descriptions of the components to use it. |
| Had to ask a sighted person where the QR code was. |
| I called an 800# only to discover the person was clueless & resorted to going online to learn about accessibility options for using this kit (something I had already done). |
| By word of mouth in the blind community I learned that I needed to download a smartphone app and someone told me how to agree to the terms and conditions before starting the test which was not very non-visually intuitive. If I did not have this knowledge I probably would have called Aira to read the paper instructions. |
| iPhone app |
| Since the instructions were in print, I had difficulty reading them with any of the apps I use, and before receiving my tests I wasn't made aware of which app I needed to download from the app store, I used my laptop to Google more information about who manufactured the tests and find the name of the app. |
| IPHONE |

### Q810 - Were you able to conduct this test on your first try?

**Table 309.** Stats for Q810: “Were you able to conduct this [Ellume] test on your first try?”

| Field | Minimum | Maximum | Mean | Std Deviation | Variance | Count |
| --- | --- | --- | --- | --- | --- | --- |
| Were you able to conduct this test on your first try? | 1.00 | 2.00 | 1.32 | 0.47 | 0.22 | 125 |

**Table 310.** Responses to Q810: “Were you able to conduct this [Ellume] test on your first try?”

| Answer | % | Count |
| --- | --- | --- |
| Yes | 68.00% | 85 |
| No | 32.00% | 40 |
| Total | 100% | 125 |

### Q811 - How long did it take to complete the steps required to perform the test, not including the time you spent waiting for results

**Table 311.** Stats for Q811: “How long did it take to complete the steps required to perform the [Ellume] test?”

| Field | Minimum | Maximum | Mean | Std Deviation | Variance | Count |
| --- | --- | --- | --- | --- | --- | --- |
| How long did it take to complete the steps required to perform the test, not including the time you spent waiting for results. | 1.00 | 4.00 | 2.82 | 1.00 | 1.00 | 124 |

**Table 312.** Responses to Q811: “How long did it take to complete the steps required to perform the [Ellume] test?”

| Answer | % | Count |
| --- | --- | --- |
| Under 5 minutes | 9.68% | 12 |
| 5 - 10 minutes | 31.45% | 39 |
| 10 - 15 minutes | 25.81% | 32 |
| Over 15 minutes | 33.06% | 41 |
| Total | 100% | 124 |

### Q812 - What format of instructions did you use?

**Table 313.** Responses to Q812: “What format of [Ellume] instructions did you use?”

| Answer | % | Count |
| --- | --- | --- |
| Paper | 11.33% | 17 |
| Digital (webpages, downloaded, etc.) | 21.33% | 32 |
| None of the above | 2.67% | 4 |
| Associated Covid Test App (Smartphone) | 64.67% | 97 |
| Total | 100% | 150 |

### Q813 - Were you able to access electronic information via a QR Code?

**Table 314.** Stats for Q813: “Were you able to access electonic information via a [Ellume] QR Code?”

| Field | Minimum | Maximum | Mean | Std Deviation | Variance | Count |
| --- | --- | --- | --- | --- | --- | --- |
| Were you able to access electronic information via a QR Code? | 1.00 | 7.00 | 5.85 | 2.36 | 5.58 | 99 |

**Table 315.** Responses to Q813: “Were you able to access electronic information via a [Ellume] QR Code?”

| Answer | % | Count |
| --- | --- | --- |
| Yes | 19.19% | 19 |
| No | 80.81% | 80 |
| Total | 100% | 99 |

### Q814 - How easy was opening the package of the Ellume test for you? (without assistance)

**Table 316.** Stats for Q814: “How easy was opening the package of the Ellume test for you? (without assistance)”

| Field | Minimum | Maximum | Mean | Std Deviation | Variance | Count |
| --- | --- | --- | --- | --- | --- | --- |
| How easy was opening the package of the Ellume test for you? (without assistance) | 1.00 | 5.00 | 3.50 | 1.08 | 1.16 | 121 |

**Table 317.**
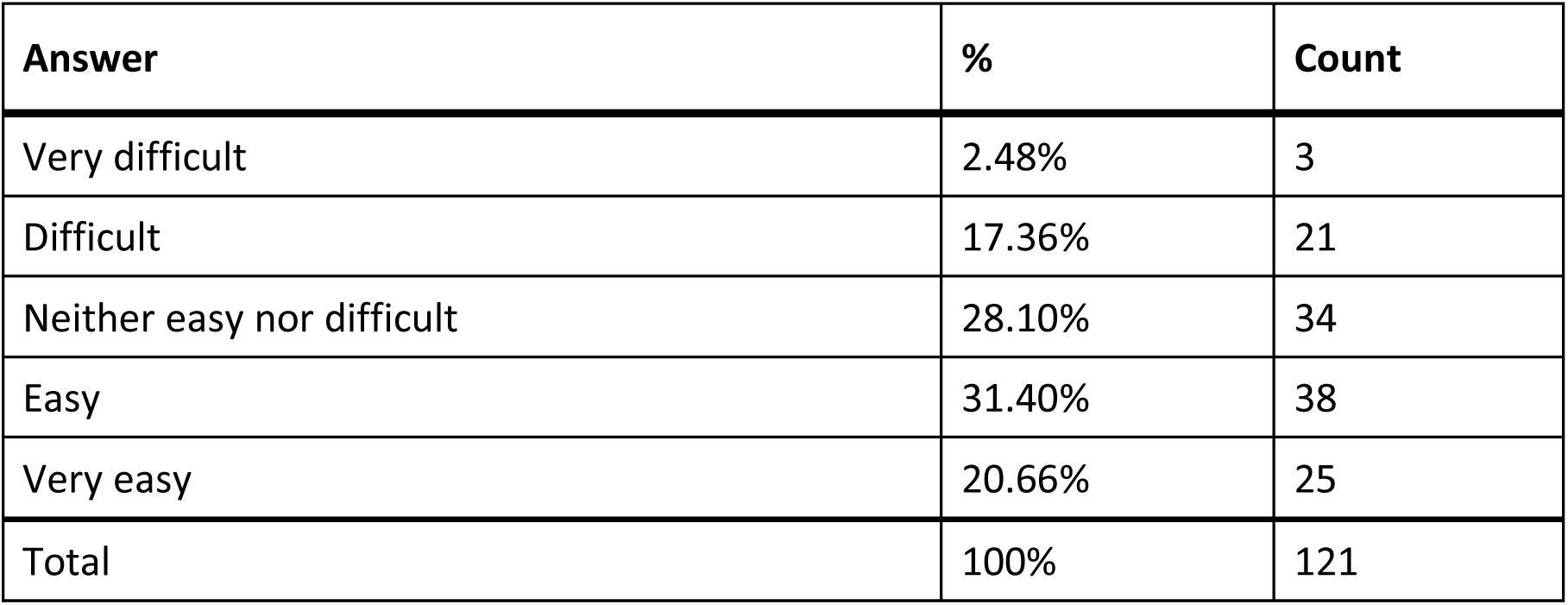
Responses to Q814: “How easy was opening the package of the Ellume test for you? (without assistance)”

### Q815 - How easy was completing the test protocol for you? (without assistance)

**Table 318.**
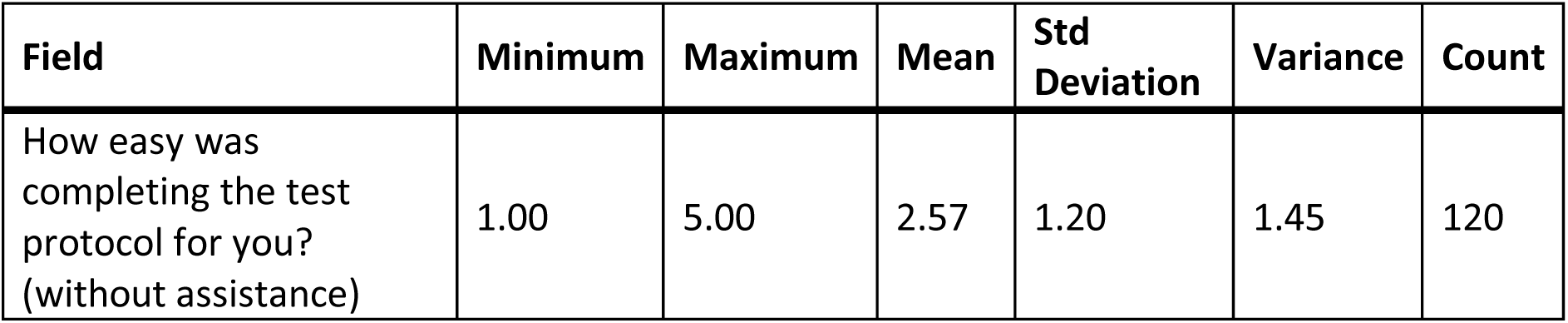
Stats for Q815: “How easy was completing the [Ellume] test protocol for you? (without assistance)”

| Field | Minimum | Maximum | Mean | Std Deviation | Variance | Count |
| --- | --- | --- | --- | --- | --- | --- |
| How easy was completing the test protocol for you? (without assistance) | 1.00 | 5.00 | 2.57 | 1.20 | 1.45 | 120 |

**Table 319.** Responses to Q815: “How easy was completing the [Ellume] test protocol for you? (without assistance)”

| Answer | % | Count |
| --- | --- | --- |
| Very difficult | 23.33% | 28 |
| Difficult | 28.33% | 34 |
| Neither easy nor difficult | 21.67% | 26 |
| Easy | 21.67% | 26 |
| Very easy | 5.00% | 6 |
| Total | 100% | 120 |

### Q816 - How easy was interpreting the test results for you? (without assistance)

**Table 320.**
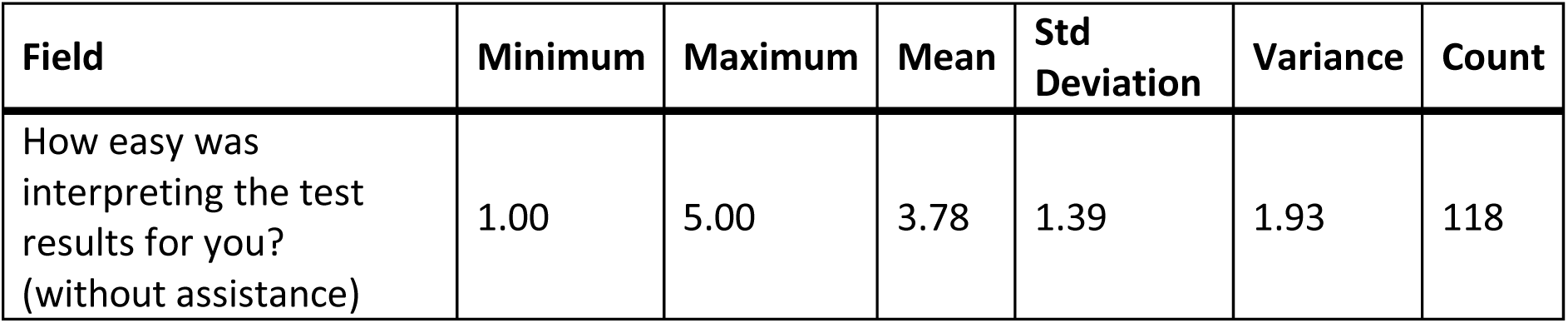
Stats for Q816: “How easy was interpreting the [Ellume] test results for you? (without assistance)”

**Table 321.**
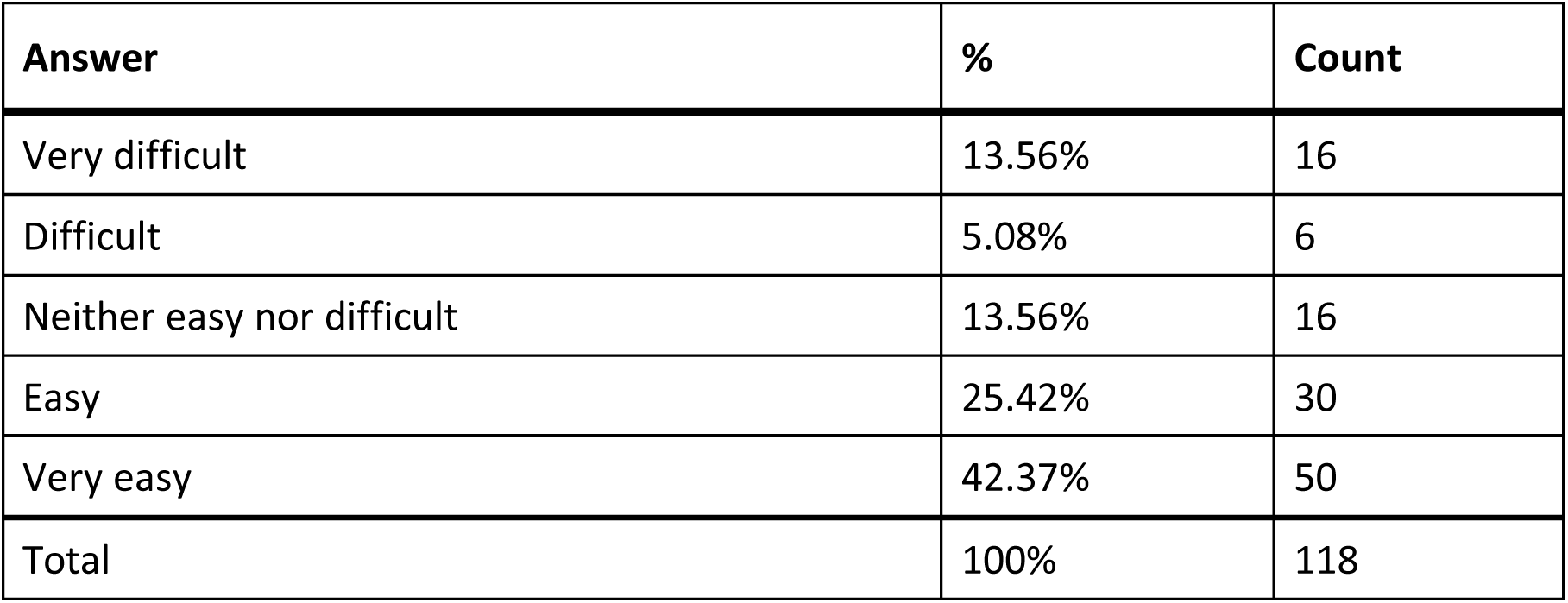
Responses to Q816: “How easy was interpreting the [Ellume] test results for you? (without assistance)”

### Q817 - Were you able to interpret the test results? (without assistance)

**Table 322.** Stats for Q817: “Were you able to interpret the [Ellume] test results (without assistance)”

| Field | Minimum | Maximum | Mean | Std Deviation | Variance | Count |
| --- | --- | --- | --- | --- | --- | --- |
| Were you able to interpret the test results? (without assistance) | 1.00 | 2.00 | 1.24 | 0.43 | 0.18 | 120 |

**Table 323.** Responses to Q817: “Were you able to interpret the [Ellume] test results (without assistance)”

| Answer | % | Count |
| --- | --- | --- |
| Yes | 75.83% | 91 |
| No | 24.17% | 29 |
| Total | 100% | 120 |

### Q818 - If there was an associated app for the test, how easy was it for you to use? (without assistance)

**Table 324.** Stats for Q818: “If there was an associated app for the [Ellume] test, how easy was it for you to use? (without assistance)”

| Field | Minimum | Maximum | Mean | Std Deviation | Variance | Count |
| --- | --- | --- | --- | --- | --- | --- |
| If there was an associated app for the test, how easy was it for you to use? (without assistance) | 1.00 | 7.00 | 3.43 | 1.39 | 1.93 | 115 |

**Table 325.** Responses to Q818: “If there was an associated app for the [Ellume] test, how easy was it for you to use? (without assistance)”

| Answer | % | Count |
| --- | --- | --- |
| Very difficult | 7.83% | 9 |
| Difficult | 22.61% | 26 |
| Neither easy nor difficult | 16.52% | 19 |
| Easy | 32.17% | 37 |
| Very easy | 17.39% | 20 |
| There is an app, but I didn't use it | 3.48% | 4 |
| Total | 100% | 115 |

### Q819 – How helpful was the app in assisting you with completing the test?

**Table 326.** Stats for Q819: “How helpful was the [Ellume] app in assisting you with completing the test?”

| Field | Minimum | Maximum | Mean | Std Deviation | Variance | Count |
| --- | --- | --- | --- | --- | --- | --- |
| How helpful was the app in assisting you with completing the test? | 1.00 | 5.00 | 3.45 | 1.33 | 1.76 | 110 |

**Table 327.** Responses to Q819: “How helpful was the [Ellume] app in assisting you with completing the test?”

| Answer | % | Count |
| --- | --- | --- |
| Very unhelpful | 12.73% | 14 |
| Unhelpful | 15.45% | 17 |
| Neither unhelpful nor helpful | 8.18% | 9 |
| Helpful | 41.82% | 46 |
| Very helpful | 21.82% | 24 |
| Total | 100% | 110 |

### Q820 - Did you receive a valid test result with the Ellume test (positive or negative)?

**Table 328.**
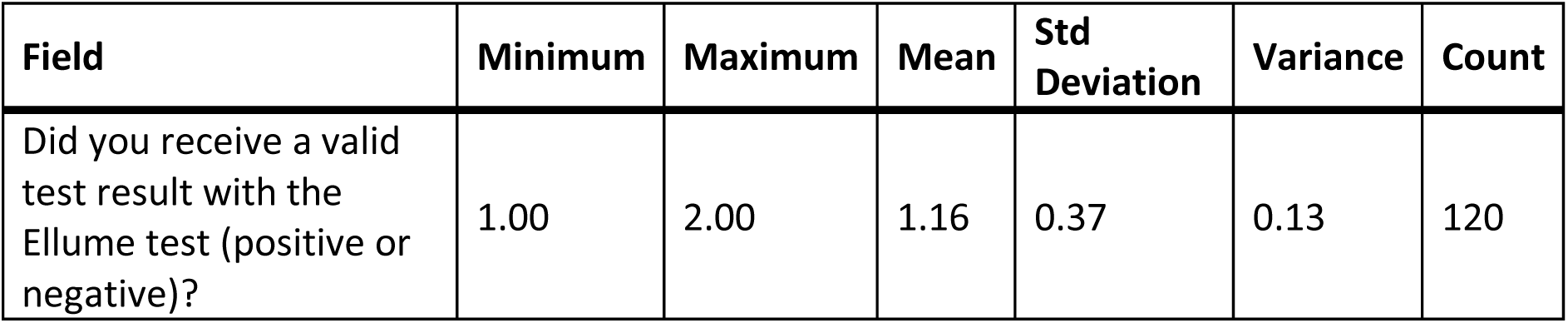
Stats for Q820: “Did you receive a valid result with the Ellume test (positive or negative)?”

| Field | Minimum | Maximum | Mean | Std Deviation | Variance | Count |
| --- | --- | --- | --- | --- | --- | --- |
| Did you receive a valid test result with the Ellume test (positive or negative)? | 1.00 | 2.00 | 1.16 | 0.37 | 0.13 | 120 |

**Table 329.** Responses to Q820: “Did you receive a valid result with the Ellume test (positive or negative)?”

| Answer | % | Count |
| --- | --- | --- |
| Yes | 84.17% | 101 |
| No | 15.83% | 19 |
| Total | 100% | 120 |

### Q821 - Did you attempt to contact Ellume customer support?

**Table 330.** Stats for Q821: “Did you attempt to contact Ellume customer support?”

| Field | Minimum | Maximum | Mean | Std Deviation | Variance | Count |
| --- | --- | --- | --- | --- | --- | --- |
| Did you attempt to contact Ellume customer support? | 1.00 | 2.00 | 1.92 | 0.28 | 0.08 | 119 |

**Table 331.**
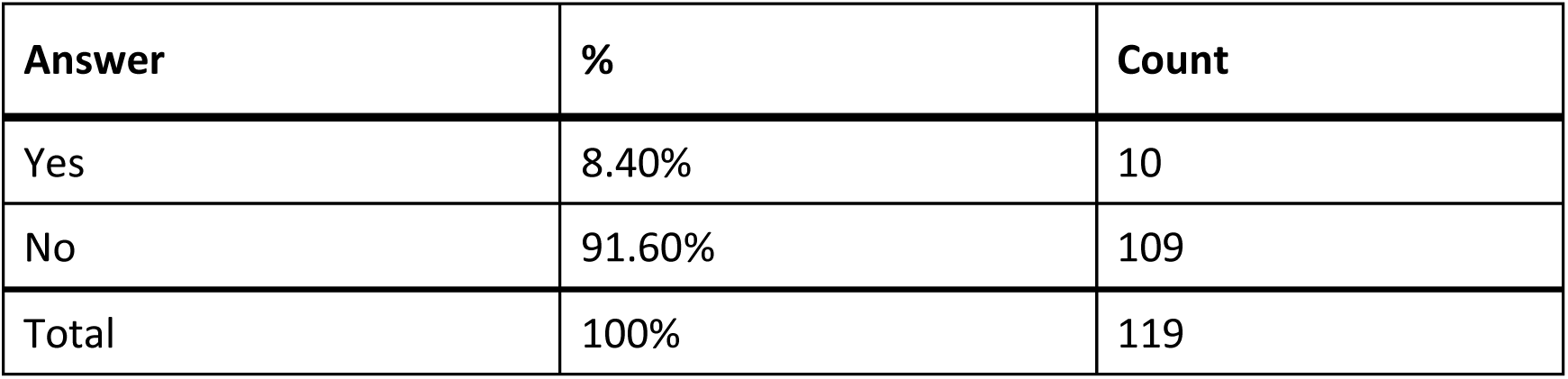
Responses to Q821: “Did you attempt to contact Ellume customer support?”

### Q822 - Were you able to talk with customer support?

**Table 332.**
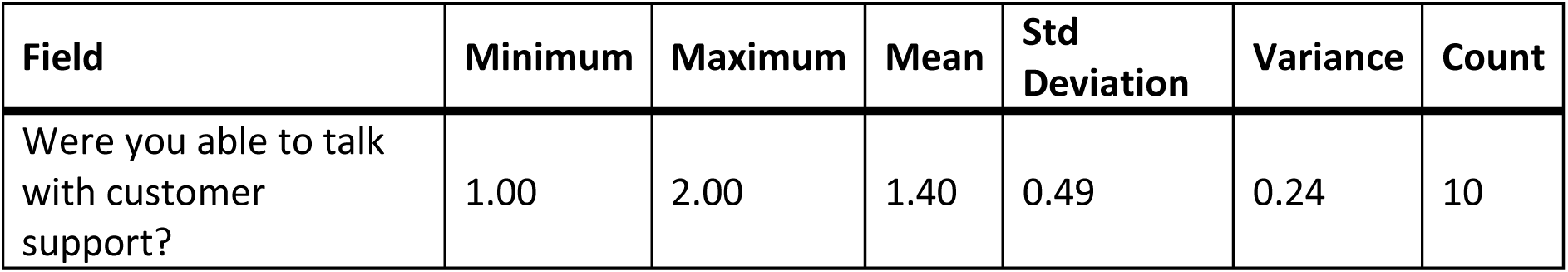
Stats for Q822: “Were you able to talk with [Ellume] customer support?”

| Field | Minimum | Maximum | Mean | Std Deviation | Variance | Count |
| --- | --- | --- | --- | --- | --- | --- |
| Were you able to talk with customer support? | 1.00 | 2.00 | 1.40 | 0.49 | 0.24 | 10 |

**Table 333.**
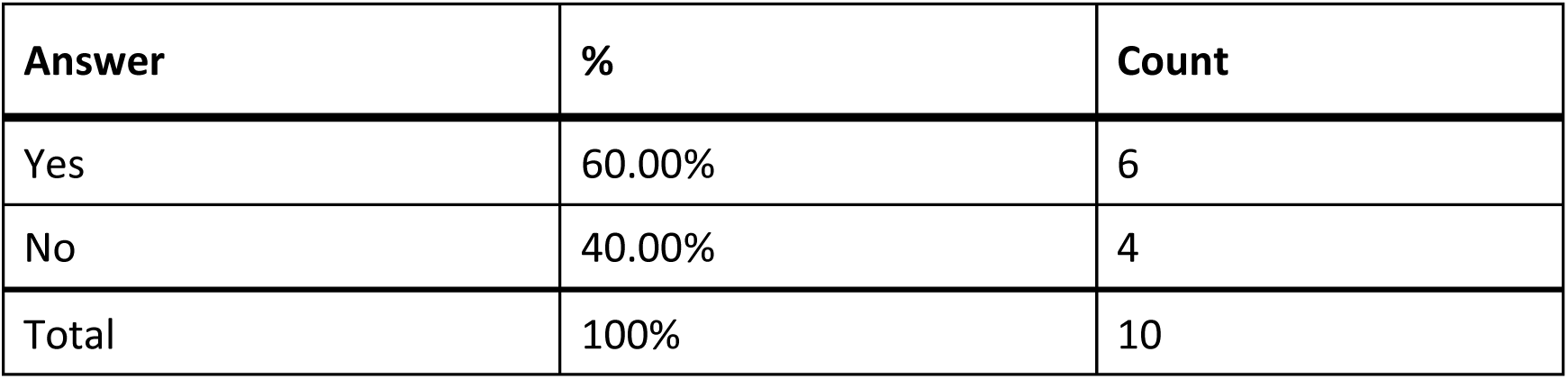
Response to Q822: “Were you able to talk with [Ellume] customer support?”

### Q823 - Was customer support helpful in answering your question(s)?

**Table 334.** Stats for Q823: “Was [Ellume] customer support helpful in answering your question(s)?”

| Field | Minimum | Maximum | Mean | Std Deviation | Variance | Count |
| --- | --- | --- | --- | --- | --- | --- |
| Was customer support helpful in answering your question(s)? | 1.00 | 6.00 | 3.67 | 2.05 | 4.22 | 6 |

**Table 335.**
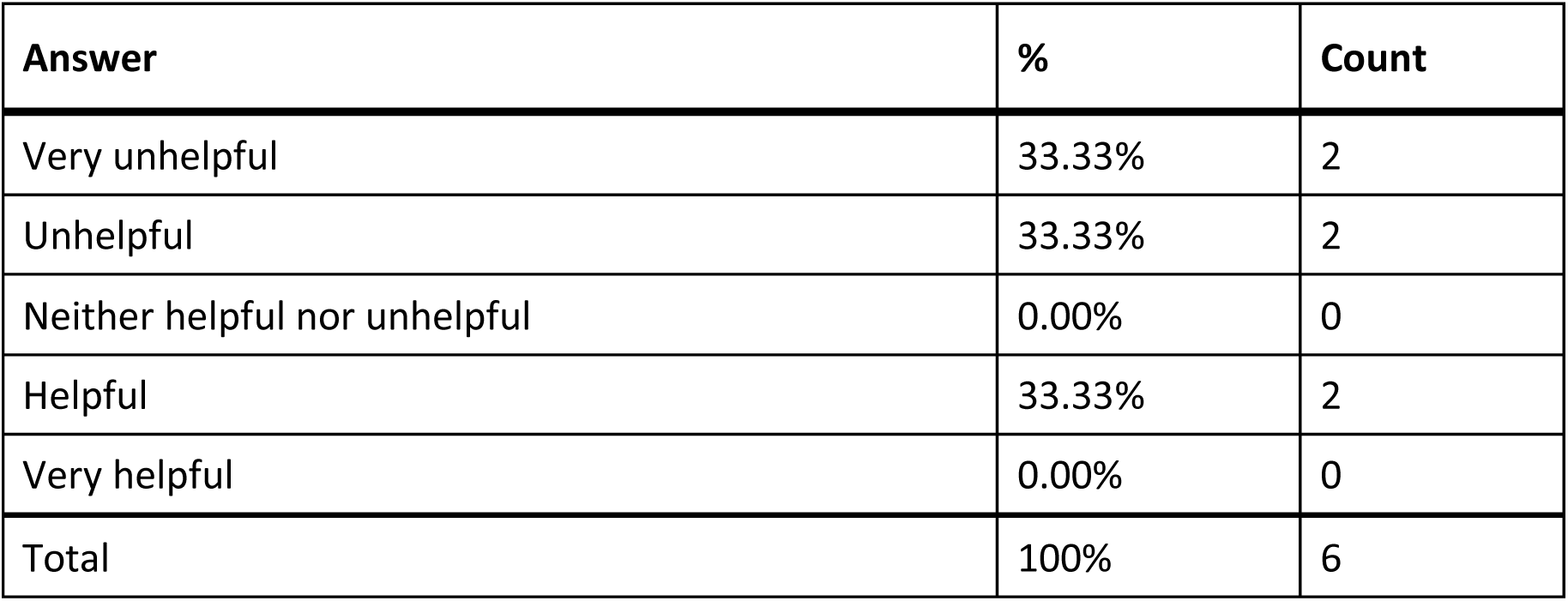
Response to Q823: “Was [Ellume] customer support helpful in answering your question(s)?”

### Q824 - If you have performed the Ellume test multiple times, how did your experience change?

**Table 336.** Stats for Q824: “If you have performed the Ellume test multiple times, how did your experience change?”

| Field | Minimum | Maximum | Mean | Std Deviation | Variance | Count |
| --- | --- | --- | --- | --- | --- | --- |
| If you have performed the Ellume test multiple times, how did your experience change? | 2.00 | 5.00 | 3.73 | 0.78 | 0.61 | 77 |

**Table 337.** Responses to Q824: “If you have performed the Ellume test multiple times, how did your experience change?”

| Answer | % | Count |
| --- | --- | --- |
| Much worse | 0.00% | 0 |
| Worse | 2.60% | 2 |
| Neither better nor worse | 40.26% | 31 |
| Better | 38.96% | 30 |
| Much better | 18.18% | 14 |
| Total | 100% | 77 |

### Q825 - What did you like about the Ellume test?

**Table 338.** Open responses to Q825: “What did you like about the Ellume test?”

| What did you like about the Ellume test? |
| --- |
| it was cheap |
| ease of use |
| Hi |
| N/A |
| N/A |
| N/A |
| App was almost entirely accessible and provided results. Pairing the test device to my phone was easy. |
| I liked that the results of the test were e-mailed to me in an accessible, electronic format that I could then forward to others. |
| It's accessible to me! I can use it! It e-mails me to let me know my result! I love it! |
| I can read my own results and don't have to find assistance to take a covid test |
| I liked that I could complete it and obtain results independently with the app. |
| Easy to use |
| Very accessible with sreen readers. |
| Mostly accessible. |
| For the most part it was pretty accessible and came highly recommended by the Biden Administration, so that was what I had ordered. |
| it was for the most part accessible |
| Sturdy components. Easy, straightforward process once you get past the atypicalities. |
| The instructions (when read by a screen reader) were adequate. |
| I appreciated the attempt to make the test accessible. |
| What's to like or dislike. It worked. |
| It seemed simple to understand once someone explained what all the items in the package were for. |
| nothing |
| The results did not take very long to recieve. Only about 15 minutes. |
| The company ripped off the government by calling it accessible. The app is a joke. |
| The test was easy to take and there was a clear indication of how far to stick the swob in your nostrils. It was easy to put the fluid in the testing card. I didn't have to call Aira to get the results |
| Ease of manipulation of components. |
| Instructions were accessible, using voiceover on the iPhone. The countdown timer and test results were easy to access. Once I learned the various components of the test I was able to independently, use the test and view the results. |
| I greatly appreciate the fact that their app is accessible using VoiceOver. It felt great to be able to conduct the test on my own without having to put anyone else's health at risk to come and assist me with taking a test. I found the entire process from start to finish to be accessible. |
| The test could be completed without vision. |
| I received a definitive result very easily without needing any help. |
| The accessibility! |
| I can view my results with the app. |
| Potential to be accessible |
| Spoken instructions via the app |
| I liked that. I could do it myself independently and I didn't need assistance. |
| It was great to be able to conduct a COVID test myself and read the results independently. The app did a nice job explaining the pieces within the kit and how to set up everything step-by-step. The app gave me the result clearly and I did not have any trouble pairing the test pad to my device. |
| Nothing |
| The app and the accessibility. |
| Results were accessible to a blind person |
| That the results are announced audibly. |
| Nothing |
| Love the fact it has an app and walks you through the process |
| The only comfort was knowing some company was making an attempt at accessibility for me. |
| It was mostly accessible. |
| the app made it easier |
| It has an accessible app |
| I liked that I could use the screen reader on my smart phone to access the app and read the instructions that way instead of having to scan them using an OCR app or ask someone else |
| to read them. also, once I realized that I needed to squeeze the bottle harder to release the testing fluid, I found this test to be entirely accessible to me. |
| The app was accessible. |
| The convenience of the test |
| I don't mind using my phone to privately access test results. |
| The independence of doing my own test |
| It is the only electronic/more accessible test that I know, and it is the one provided free by the US government (though obtaining them is difficult). |
| No interpretation of results needed |
| Being able to email the the the results |
| I like the fact that they pass a video app to show you how to use the product. |
| That the results could be shared electronically. |
| The app gave clear instructions and descriptions of all the parts of the test. using my cell phone and bluetooth was very simple to get my results independently |
| QR code reader instructions in an app and test results in app |
| Nothing. They never tested their test kit or the app with a blind user. |
| The iOS app worked well. The rest was complicated without sighted help. |
| The only thing I liked about the test was that I could read the test results for myself. |
| I liked that there was an app that made it possible to interpret the test results without assistance. |
| Able to 100% independently get result by app without any type of sighted in person or remote interpreter assistance. |
| I could read my test results from my phone. |
| The app. I like technology. Seemed to make the process easier overall and results were definitive and easy to read |
| It is a start in making tests accessible, but only a start. |
| I liked the fact that as a blind person, I could conceivably do it myself. It would have taken me much longer and I didn't feel comfortable dealing with the fluid stuff without sighted assistance. |
| I like that it was accessible. |
| The experience was accessible from start to finish. The smart phone app made it so much easier to follow along instructions with voiceover and tutorial video. All of the pieces were appropriate size and no difficulties collecting sample. |
| The notion or initiative of including blind people |
| Seemed to be simple and straight forward to use. |
| Convenience to use at home & do myself ! |
| Nothing |
| It was free and marketed as accessible to the blind community |
| The instructions are clear. The app is compatible with voiceover on the iPhone. |
| This test at least does make a start at being accessible, however, it does have a long way to go. |
| It did make it possible for me to use the test privately and independently once I learned how. |
| The ONLY thing I liked about the Ellume home test was the fact that I received an email with my test results. |
| Get test results in the app and not the test applicator |
| I like that there is an app which guides users through the steps. The video narration is very helpful, and that is unfortunately very uncommon for instructional videos so I very much appreciated that. I have some feedback about some of the features not being the most accessible but most of it is and there was very clear step-by-step guidance. |
| I like that it was simple and straight forward. Also, I like that every part of it was in its own packaging and easy to identify. |
| I did not like anything about the test. When a company claims that their test is acceptable, I expect it to be usable by all Disabled groups independently. That is certainly not the case for blind and deaf blind individuals. |
| Phone app, |
| I could use my phone to see the results. |
| Designed with low vision and blind folks in mind. |
| The test results were accessible through the mobile app |
| I liked that the unit read me the results when the test was complete. I like the dropper better on this test because it was easier to manage after taking the collection. |
| That' it was free and it arrived in 6 days |
| I liked being able to access the test results independently. |
| It was possible to do the Ellume test independently. |
| I liked that I could potentially perform the test independently, and that the app gave step by step instructions. |
| Bluetooth and that I don't have to attempt to read the results. |
| I was able to do it by myself without assistance from anyone. Privacy was kept. |
| A clear answer to the results was sent to my email it was easy to read |
| the verbal instructions were excellent, both in the video and I appreciated that they were done step by step as I went along with the test.. |
| I was able to process the test by myself. |
| The app made it easy for someone with a visual impairment |
| It was easy to use, and the instructions were given step by step. I was able to perform the test independently. |
| It works with a smart phone. |
| I liked that there was an associated smartphone app. |

### Q826 - What would you like to see improved for the Ellume test?

**Table 339.** Open responses to Q826: “What would you like to see improved for the Ellume test?”

| What would you like to see improved for the Ellume test? |
| --- |
| cost |
| Not having to watch the video each time a test is performed. |
| N/A |
| N/A |
| N/A |
| A few text boxes weren't readable by voiceover. The process took far too long, including forcing the user to watch a video that was less helpful than just reading the instructions. Took too much personal data before letting me use the app |
| The associated app for this test needs to be revised for accessibility with screen reading technology including the navigation of form fields and audio description of test protocol video to accompany image-based content. |
| If you've taken the test before, sitting through the tutorial over and over can be annoying. I'd like the packaging to be openable without scissors. You can't use your phone while you're waiting for the test results. |
| The app. There is no clear indication either in the app or from the test device when bluetooth is connected. A simple tone or text on the screen would be sufficient. Also, there is a strange bug that when my phone is muted, video audio does not play. I had to get sighted assistance to help me understand what was happening. |
| I would like to not have to enter my information in the app. I would also like to skip the intro video in the app. |
| Better identification of the components. |
| Easier way to know if drops go to the right place for testing, not having to watch video once you've seen it before. |
| More properly-labelled buttons in the app and a Braille quick-start guide. |
| Not having to create a new profile each time the same individual takes a test. |
| Better accessibility of form fields and other controls in the app. The ability to create a profile and eliminate the need to enter the same information with every new test. Multiple profiles would also be useful for households with multiple members. The ability to skip the video. |
| larger print, color differences of items (nothing clear) |
| Text in app is small; had to use magnifier and screen reading. Some instructions are incomplete (do not identify all components in the test well). Was a challenge to use the dropper and get the proper number of drops into the analyzer. Finally figured out to rest the dropper in the slot while squeezing. |
| I would like braille instructions at least to be able to get started. |
| If I remember correctly, the test reader has lights on it to signify the level or where it's at in the process. I think as a blind person it would be helpful if there's either a tone with varying pitches to detect the step it's at or vibrations for someone who's deaf-blind. |
| I don't like the fact that I have to download an app to my phone in order to get results |
| I would like to see the tests made easier to access independently. Currently, one has to relay on a third person to tell them whether they are positive or negative. |
| Hire someone who is trained in accessible equipment design. |
| I had a few accessibility issues with the app. Sometimes when I was clicking on an edit field it wouldn't open so I can type. Sometimes it would skip over edit fields too. |
| Audible feedback when pressing power/connect button; ability to create patient profiles so info not needed to be entered each time; ability to skip video after familiarity with test. |
| Not having to re-enter personal information for every test used, and the option to skip the video after it has been viewed the first time. Also having some way for the test to signal when the correct number of drops has been added to the test in order to analyze the sample. it is not possible to independently and non-visually determine the number of drops being added. |
| Ellume covid test 1. I could not tell which app to download from the test kit box. The name Ellume was miss pronounced by the screen reader when I used Seeing AI to OCR the test kit box. 2. Voice over could not swipe through all the elements in the app. Explore by touch was necessary to reach all controls. 3. The birthdate field was visible to Voice over as two edit fields but date pickers for day month and year popped up but were not announced by the screen reader. I found the date pickers only by accident. 4. Some of the edit fields were improperly exposed as two edit fields to voice over. It took 2 swipes right to move to the next edit field. 5. The instructions did not explain in terms that someone who cannot read the packaging which device was which. 6. As a blind person I can't see the blinking light under most lighting. Please add sound. 7. No controls have been provided for voice over to stop the video. 8. Don't ask a blind person to place 5 drops. Instead, design a bladder with the necessary liquid that pops when the swab is secured in the container. Likewise provide a valve or again puncture the test sample when it is inserted into the analyzer. So swab |
| inserts into the bladder of liquid and the entire swab container inserts into the analyzer. 9. The app froze and I had to quit it in order to see my results. 10. It's impossible to keep the phone and the analyzer less than 3 inches apart for the entire test. I use the on screen braille keyboard to fill out the test questions. 11. |
| No improvements needed. |
| All is fine! |
| Don't make people watch that same video every time they test. Give us the option to watch it. That and tell people ahead of time what app to download. |
| The app not requiring so much personal information before you can administer the test and somehow to have the drops pre-counted since blind people can't count drops from a dropper accurately |
| I would like for the spoken directions to better explain the different components of the test, and what they should feel like along with instructions on how to use each component. Also, there was a component that lit up and due to my visual impairment, I could not tell when that component lit up. A better design would be a sound for the visually impaired when it is ready to use that component |
| I would like better identification of the complaint parts, especially when you're doing it the first time I had to figure it out on my own because the descriptions in the app went good enough to identify the individual components. |
| I did not have issues pairing the test to my iPhone, but did hear that some people had trouble and were not able to use it to receive their test results. I also did use Aira assistance to read the name of the test on the box and go through the contents before I installed the app. I wish the kit came with braille instructions somehow or a braille label on the box so I would have known what the brand name was to download the app myself, but it was not a big deal. If there was a QR code that had this information on, I was not aware of it. A QR code may be a good addition if it is not already there. |
| I wish the app was easier to use/see. |
| Having to watch the instructional video . I would prefer to just read the instructions. |
| Clearer instructions from the vendor, more stable app as it crashed several times during testing, shorter/more comfortable swab |
| Better instructions via the app, not just a video. Better packaging to tell what is needed when. Remove the salt preservation packet! |
| It was very inconvenient to have to keep the test flat on the table next to my phone which also had to be flat on the table. This made it impossible to use magnification to count the drops. I had to get a sighted person's help. Counting drops is really next to impossible for me. |
| No app Identify the package size feel to identify what is in it |
| 1. The process to drop the sample in the sample well involves counting 5 drops, yet you have no way of knowing whether a drop dropped, let alone figuring out how many drops you dropped. It would be great if the app can make a sound or some kind of indication that a drop has been added to the sample well. |
| The app said to press the button on the analyzer for bluetooth connection; I found nothing on any of the contents that looked like a button. When I couldn't establish a connection, I abandoned the test. |
| stop requiring video each time and pare down NINETEEN pages of directions. Even though all were not in English, the amount of paperwork in microprint was quite daunting. Especially when the Help Desk was closed for the holiday weekend while NFB members were trying accessible testing prior to convention. |
| Don't make me watch the video every time once I've been through the procedure before; make it optional. |
| mixing liquids needs to be made more simple |
| The way you apply the solution to the test |
| I did not have any issues with the test not starting up because I couldn't see if the light had come on or not, but maybe having a beep or some other audio feedback when the light comes on would be helpful. |
| Better description of parts; easier interpretation of error codes; better accessible instructions; actually tailored for visually impaired people, rather than just saying, the whole thing was accessible. pretty much everything. |
| I did not like that i had to start my identification process over every time i used my same phone to take an ellum test. It should create an profile so you can access the process without the intro once you have used it more than once. Thats must my opinion |
| The second time, it was like the analyzer didn't work. The light just continued to blink after I applied the sample. That time, my husband had helped me perform the test. |
| I would like to see a way of saving your information you enter in the app. I have to repeatedly enter in my name, address, and so on the six times I used this app. There really should be a way to save profiles if multiple people use the app. |
| A step-by-step description of the process (pretend that the viewer cannot see) and a description of the testing components. The app can be improved (the buttons move and scrolling to the end does not work); the opt-out of arbitration provision is inaccessible, requiring a blind person to mail a signed statement without providing an email option, which is inaccessible and perhaps violative of the ADA. |
| Better respect for user privacy |
| Maybe a description of component's with picture |
| I would like to see a better way for a vi person to know when the device has been paired to the phone. Not using scheduled to see if there will be a light is flashing or not. |
| That it be more accessible to the visually impaired person |
| the only thing that slowed the process down was the fact that every test I had to reenter my personal information. that took time that I could have been performing the test. |
| Still cannot count drops of fluid as a blind person, had to put way too much personal information into the app to use the app. |
| I need it to work. |
| They are not qualified to produce an accessible test kit. |
| Better labeling of the components. |
| The smartphone app needs to be radically redesigned. First, I don't want to be required to fill in my personal information EVERY SINGLE TIME I take the test. The data entry fields need to be optimized for screen reader users. Users shouldn't have to watch the video every time we take the test. Blind people can't see that video anyway. E E The video is a complete waste of time. In short, the app is a piece of junk, and it needs a major overhaul. |
| More descriptive details about the test components. |
| App is clunky to use, have to type in patient info does not store it, have to watch video and no way to skip video. Also not as accurate as the iHealth test. Has a lot of promise but the app needs major improvement in terms of ease of use and speed to get it setup. |
| Better instructions tailored to someone who is blind. Also the video needs either captions or you should be able to choose between reading the text of the instructions or playing the video. The video is not accessible to someone who is DeafBlind. |
| Easier to open packaging |
| Large print instructions, that would help everyone. better tactile information. |
| I tried to have the results emailed to me, and I selected that during the process, but it didn't work, and I almost didn't get the result. It came too quickly and vanished. I luckily heard it once. |
| Away for individuals who do not have smart phones to be able to take this test, alternative instructions that can be accessed by individuals who are deaf blind, and an easier way to know exactly how much liquid to put onto the test strip. |
| Absolute overhaul of the end products, and bringing someone who is well-versed with accessibility standards onboard |
| The way the solution and the applicator thing that the swab went into is too hard to squeeze to get the solution into the tester. |
| Not have to enter information and watch video everytime. |
| Your target audience is the blind community When you Box a test it would be very helpful to physically describe what is in the Box example the agitator is no long rectangular Box or package or package there is a file about an inch long take that file find the very tiny tiny hole in the agitator and pour it into it definitely descrdefinitely descriptions could be so much better |
| Add a description of each item in the box to make sure that you're performing the test correctly |
| App developer needs to appropriately code the app |
| The one part of the test that I am not able to complete independently is ensuring that the correct number of drops or put into the test reservoir. The app reiterates that the specified number of drops is crucial for accurate test results. I would potentially feel comfortable trying to complete the test myself if there was some margin of error for number of drops. |
| I would like to see an option for the test that does not rely on a smartphone or tablet as not everyone has one of these devices. Additionally, I would like to see it be easier to connect the receiver with the smartphone. Despite multiple attempts, I could never complete this step. As a result, I could not use the test. Lastly, Braille test instructions would be great. |
| Instructions should not be accessible with a QR code. The app should not collect data from users with visual disabilities. Test components should not use flashing lights or other visual indicators to establish when connections are made. |
| labeling of the components and making it easier to add the drops of solution. |
| Everything else! Although the Ell-me app gave step-by-step instructions about how to administer the test, there is no way at all that a blind person can carry out these steps without sighted assistance. Just because the app can talk you through it doesn't make the test itself in any way accessible! |
| Not have a blinking light |
| There needs to be more advertising about where users can find digital instructions, or a tactile boundary and braille QR code label on the box as it's not discoverable. The terms and conditions agree button needs a better label, feedback I am aware has been passed along but has yet to be remedied. The video sound does not play if an iPhone mute switch is on and this can throw off blind users into thinking the video might not be playing. Since watching the instructional video is required before each test can begin, I appreciate that there is a no sound option as it's annoying to listen to repeatedly. However, there needs to be a screen reader detectable label on the play/pause button to indicated state. Lots of small fixes that would be improved if they complied with WCAG 2.1. I do not have disabilities that impact my dexterity but I found the package very difficult to open, probably for child proofing reasons, but it's not accessible. I have to use scissors. Finally, it would be great if the test strip had a window to display the result on it so using a smartphone would not be required. I understand the visual display would not be accessible but it seems like a missed opportunity for people who would benefit from that. |
| In addition to the LED light, they could add a sound to know status of light. |
| I do not want to watch the video every single time. It is quite honestly a waste of time after watching it once. It would be nice if there was an option to skip it, even if you have to check a checkbox that affirms that you know all of the information contained in the video if you are going to perform a second test. It does not need to be skipped automatically, but there should be a skip button. |
| More descriptive video |
| The test claims to be accessible, but it is not. There is absolutely no way for a totally blind person to use the test independently. And there is no excuse for that. The components could have some sort of raised markers so that blind people could identify the components. There could be braille instructions. And for those with arthritis another hand challenges, the packaging could be easier to open. |
| Clearer instructions for getting fluid into tester. |
| It was hard to see the number of drops applied. If I were totally blind this would have proven to be even more difficult |
| Make the packaging clear to help sort out what was what. Make the video available to play again without having to start the log in all over again. |
| n/a |
| I resented that I had to provide so much personal information on the app plus it made the whole process much more time consuming and complicated. I didn't give my real info because I had no idea why they needed it or what they were going to do with it or my test results. I also didn't like the collection method of sticking the swab so far up my nose. I also had to call Aira because I didn't know which end of the swab to open since we couldn't touch it. |
| Better tube and equipment identifiers ie tactile or collered coordinated because not all can read braille |
| The user interface is poorly designed for screen reader users. Many of the controls are difficult to manipulate. The app should store user information, so one does not have to go through the tedious task of entering user data and going through a video that a blind user can't see anyway. Pairing the test module with the sm smartphone needs to be easier. |
| More description of the component parts, and precise instructions. |
| I would like to have some means for blind people to know how many drops of the testing solution are being squeezed out. The number of times someone squeezes the tube are not necessarily accurate, as how firmly or gently to squeeze factors in. This could cause inaccurate test results. Also, it would be nice if, after the first time a person takes a test, each time one does an additional test, the video demonstration and entering of personal details could be bypassed. |
| Easier packaging. The app needs to store info so you don't have to enter it each time and also record that you've already watched the video. It takes so long to do both of those things each time. |
| They need to have better verbal or written descriptions of the components. And better idea of how they are used. That was the problem I had to figure out what the components did so the first time I used it I got some of the components incorrect. |
| There was no large print on any of the components and it was not immediately apparent which component was which when I first open the box. I had to enter a tremendous amount of personal information into the app before I could move forward with just taking the test. This is a personal invasion of my information and very time consuming while I was quite sick. No indication how all of that information was going to be used. I should be able to use my smart phone and take each component while still in its package and have it read the barcode to tell me what the component really is rather than me having to guess. The one piece of audible instruction was a waste of time because you can't go back to the audible instruction once you proceed forward with the test. I am a former critical care trauma nurse I've had to do very complicated tests on patients for years. I am completely independent and still cook full recipes and work on the computer daily as part of my job as a nurse. The packaging for each component should've been either very clear and easy to read or a barcode should've |
| been easily scanned so I will be able to recognize the component and know which one was which to have it in the proper order. Also there should be an audio Q once you have coded enough solution onto the test strip for the test to begin. |
| I would like a verbal description of the items because it took me a while to figure out which was which; and perhaps directions on how to take them out of the box so I wouldn't touch the end that I shouldn't. |
| I would like to be able to bypass the video in the iPhone app and just use the instructions for the test itself. I believe that there was one unlabelled button that I needed to tap to do something. mandatory fields on the app should be marked |
| Identifying package contents, not having to register for a new profile each time I took a test. |
| I would like to be able to skip the video on the app each time I perform the test. |
| The numbers of drops going into the port is not easy to ID when you are completely blind. So I've no clue how many were actually inserted. |
| increased usability for people with limited motor skills. |

### Q827 - Siemens Clinitest Think of the first time you used the Siemens Clinitest. Did you have difficulty completing any of the following tasks? Select all that apply

**Table 340.** Responses to Q827: “Siemens Clinitest - Did you have difficulty completing any of the following tasks?”

| Answer | % | Count |
| --- | --- | --- |
| Opening the packaging | 9.09% | 2 |
| Reading the instructions | 22.73% | 5 |
| Identifying test components | 9.09% | 2 |
| Collecting the test sample | 13.64% | 3 |
| Manipulating test components | 13.64% | 3 |
| Interpreting the test results | 18.18% | 4 |
| Other | 0.00% | 0 |
| I did not have difficulty completing the test | 13.64% | 3 |
| Total | 100% | 22 |

#### Q827_7_TEXT - Other

None

### Q828 - What tools or assistance, if any, did you use to perform the test?

**Table 341.** Responses to Q828: “What tools or assistance, if any, did you use to perform the [Siemens Clinitest] test?”

| Answer | % | Count |
| --- | --- | --- |
| Timer | 40.00% | 8 |
| Scissors | 5.00% | 1 |
| Remote visual assistance (AIRA, Be My Eyes, Facetime, Video-chat, etc.) | 15.00% | 3 |
| Screen Reader | 5.00% | 1 |
| Magnification | 15.00% | 3 |
| Other technology (OCR, Bar Code Readers, Light Detectors, etc.) | 5.00% | 1 |
| In person human assistance | 10.00% | 2 |
| Other | 0.00% | 0 |
| I did not need any tools or assistance | 5.00% | 1 |
| Total | 100% | 20 |

#### Q828_8_TEXT - Other

None

### Q829 - Were you able to conduct this test on your first try?

**Table 342.** Stats for Q829: “Were you able to conduct this [Siemens Clinitest] test on your first try?”

| Field | Minimum | Maximum | Mean | Std Deviation | Variance | Count |
| --- | --- | --- | --- | --- | --- | --- |
| Were you able to conduct this test on your first try? | 1.00 | 2.00 | 1.09 | 0.29 | 0.08 | 11 |

**Table 343.** Responses to Q829: “Were you able to conduct this [Siemens Clinitest] test on your first try?”

| Answer | % | Count |
| --- | --- | --- |
| Yes | 90.91% | 10 |
| No | 9.09% | 1 |
| Total | 100% | 11 |

### Q830 - How long did it take to complete the steps required to perform the test, not including the time you spent waiting for results

**Table 344.** Stats for Q830: “How long did it take to complete the steps required to perform the [Siemens Clinitest] test?”

| Field | Minimum | Maximum | Mean | Std Deviation | Variance | Count |
| --- | --- | --- | --- | --- | --- | --- |
| How long did it take to complete the steps required to perform the test, not including the time you spent waiting for results. | 1.00 | 4.00 | 2.45 | 0.78 | 0.61 | 11 |

**Table 345.** Responses for Q830: “How long did it take to complete the steps required to perform the [Siemens Clinitest] test?”

| Answer | % | Count |
| --- | --- | --- |
| Under 5 minutes | 9.09% | 1 |
| 5 - 10 minutes | 45.45% | 5 |
| 10 - 15 minutes | 36.36% | 4 |
| Over 15 minutes | 9.09% | 1 |
| Total | 100% | 11 |

### Q831 - What format of instructions did you use?

**Table 346.** Responses to Q831: “What format of [Siemens Clinitest] instructions did you use?”

| Answer | % | Count |
| --- | --- | --- |
| Paper | 72.73% | 8 |
| Digital (webpages, downloaded, etc.) | 9.09% | 1 |
| None of the above | 9.09% | 1 |
| Associated Covid Test App (Smartphone) | 9.09% | 1 |
| Total | 100% | 11 |

### Q832 - Were you able to access electronic information via a QR Code?

**Table 347.** Stats for Q832: “Were you able to access electronic information via a [Siemens Clinitest] QR Code?”

| Field | Minimum | Maximum | Mean | Std Deviation | Variance | Count |
| --- | --- | --- | --- | --- | --- | --- |
| Were you able to access electronic information via a QR Code? | 1.00 | 7.00 | 4.00 | 3.00 | 9.00 | 2 |

**Table 348.**
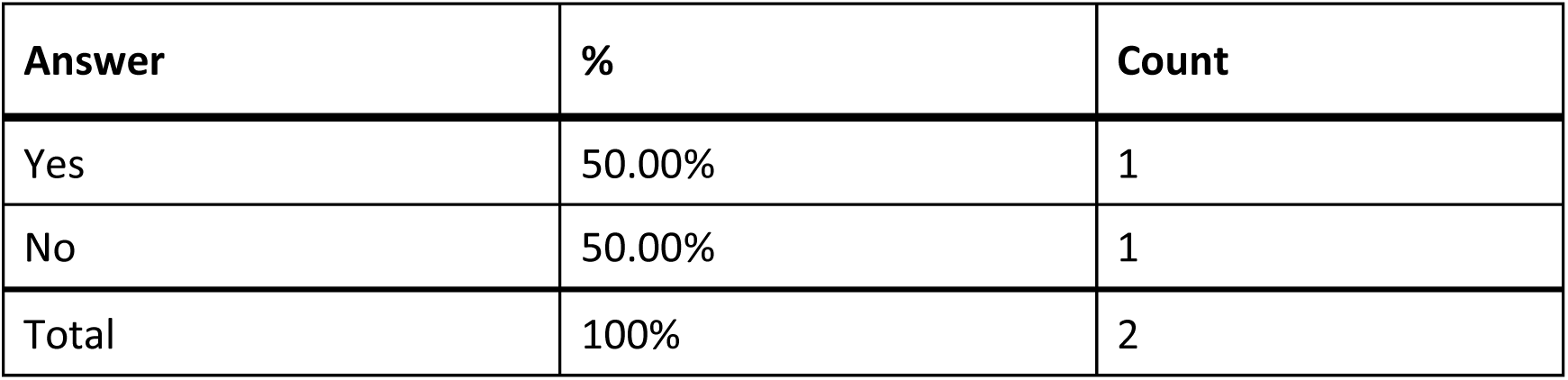
Responses to Q832: “Were you able to access electronic information via a [Siemens Clinitest] QR Code?”

### Q833 - How easy was opening the package of the Siemens Clinitest for you? (without assistance)

**Table 349.**
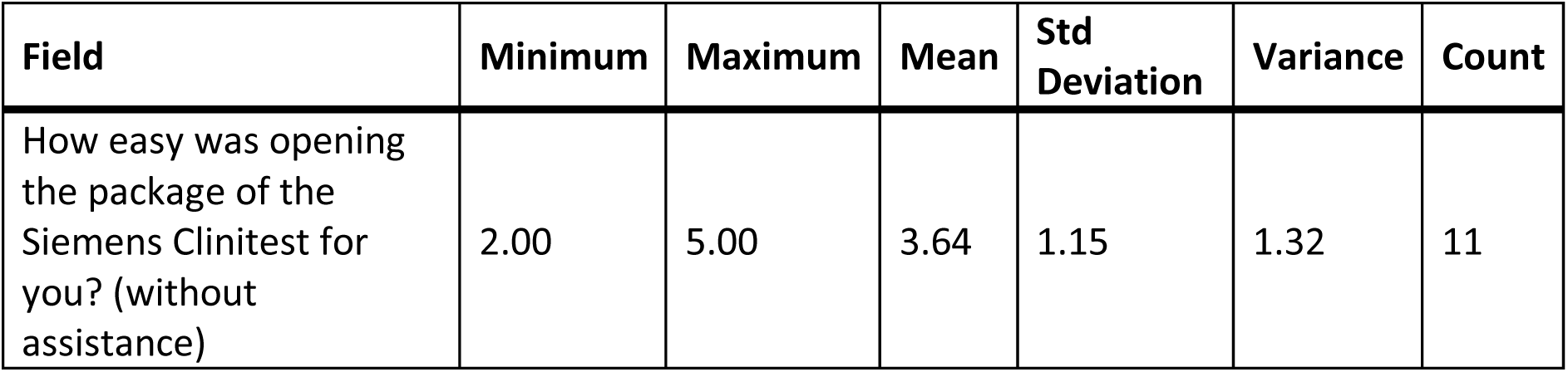
Stats for Q833: “How easy was opening the package of the Siemens Clinitest for you? (without assistance)”

**Table 350.**
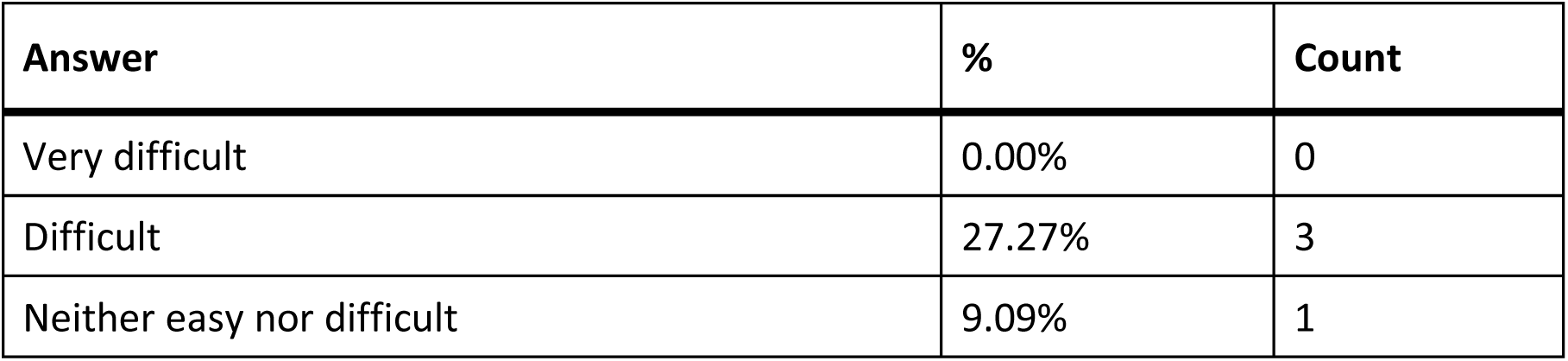
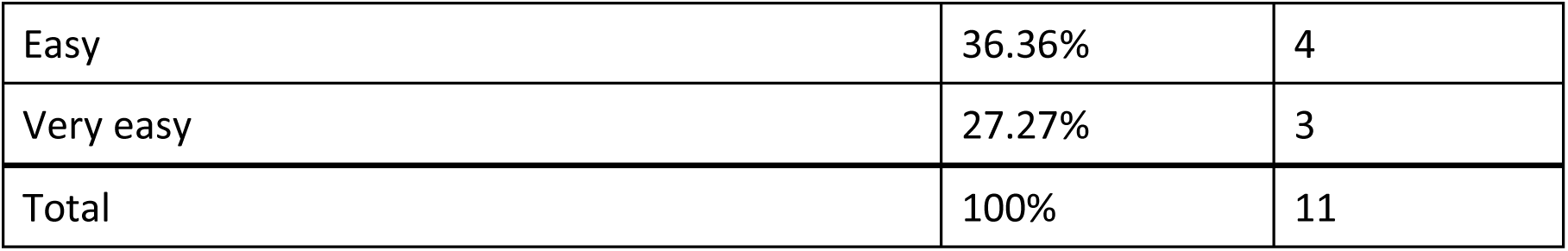
Responses to Q833: “How easy was opening the package of the Siemens Clinitest for you? (without assistance)”

| Answer | % | Count |
| --- | --- | --- |
| Very difficult | 0.00% | 0 |
| Difficult | 27.27% | 3 |
| Neither easy nor difficult | 9.09% | 1 |
| Easy | 36.36% | 4 |
| Very easy | 27.27% | 3 |
| Total | 100% | 11 |

### Q834 - How easy was completing the test protocol for you? (without assistance)

**Table 351.** Stats for Q834: “How easy was completing the [Siemens Clinitest] test protocol for you? (without assistance)?”

| Field | Minimum | Maximum | Mean | Std Deviation | Variance | Count |
| --- | --- | --- | --- | --- | --- | --- |
| How easy was completing the test protocol for you? (without assistance) | 1.00 | 5.00 | 2.45 | 1.23 | 1.52 | 11 |

**Table 352.** Responses to Q834: “How easy was completing the [Siemens Clinitest] test protocol for you? (without assistance)?”

| Answer | % | Count |
| --- | --- | --- |
| Very difficult | 27.27% | 3 |
| Difficult | 27.27% | 3 |
| Neither easy nor difficult | 27.27% | 3 |
| Easy | 9.09% | 1 |
| Very easy | 9.09% | 1 |
| Total | 100% | 11 |

### Q835 - How easy was interpreting the test results for you? (without assistance)

**Table 353.** Stats for Q835: “How easy was interpreting the [Siemens Clinitest] test results for you? (without assistance)”

| Field | Minimum | Maximum | Mean | Std Deviation | Variance | Count |
| --- | --- | --- | --- | --- | --- | --- |
| How easy was interpreting the test results for you? (without assistance) | 1.00 | 5.00 | 3.45 | 1.44 | 2.07 | 11 |

**Table 354.**
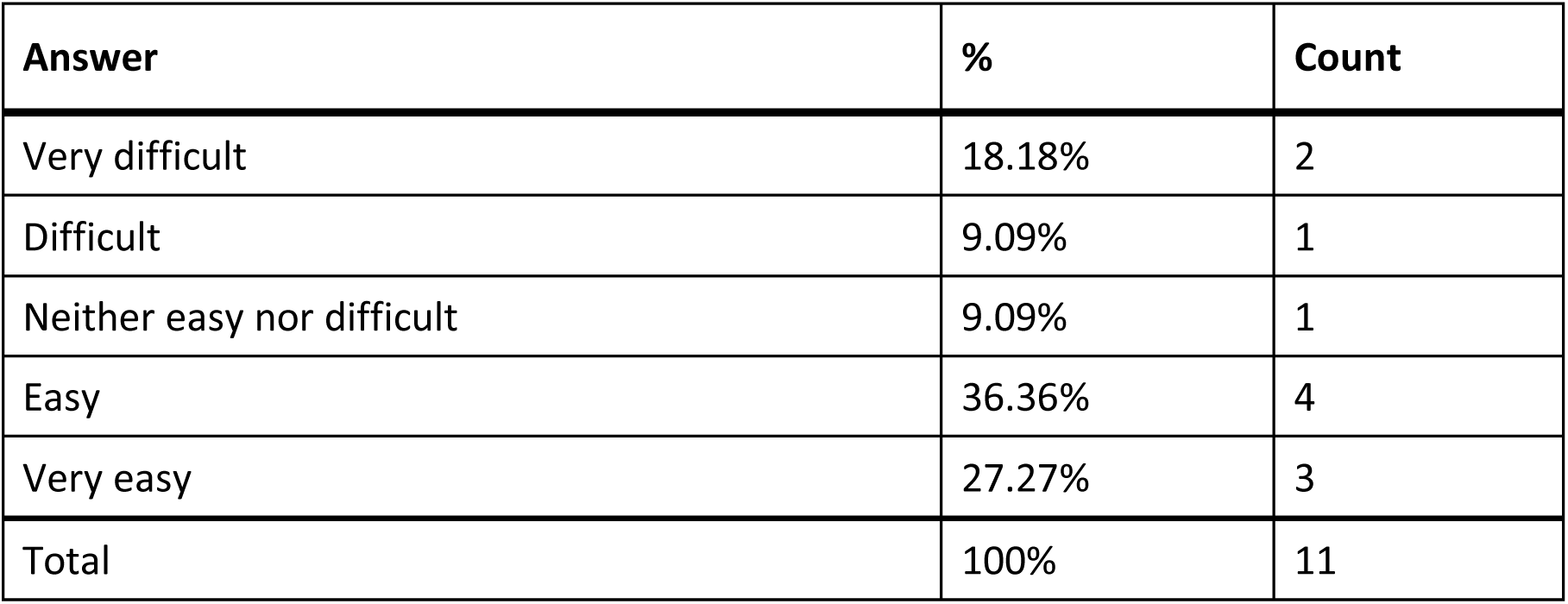
Responses to Q835: “How easy was interpreting the [Siemens Clinitest] test results for you? (without assistance)”

| Answer | % | Count |
| --- | --- | --- |
| Very difficult | 18.18% | 2 |
| Difficult | 9.09% | 1 |
| Neither easy nor difficult | 9.09% | 1 |
| Easy | 36.36% | 4 |
| Very easy | 27.27% | 3 |
| Total | 100% | 11 |

### Q836 - Were you able to interpret the test results? (without assistance)

**Table 355.** Stats for Q836: “Were you able to interpret the [Siemens Clinitest] test results? (without assistance)”

| Field | Minimum | Maximum | Mean | Std Deviation | Variance | Count |
| --- | --- | --- | --- | --- | --- | --- |
| Were you able to interpret the test results? (without assistance) | 1.00 | 2.00 | 1.36 | 0.48 | 0.23 | 11 |

**Table 356.** Responses to Q836: “Were you able to interpret the [Siemens Clinitest] test results? {without assistance)”

| Answer | % | Count |
| --- | --- | --- |
| Yes | 63.64% | 7 |
| No | 36.36% | 4 |
| Total | 100% | 11 |

### Q837 - If there was an associated app for the test, how easy was it for you to use? (without assistance)

**Table 357.**
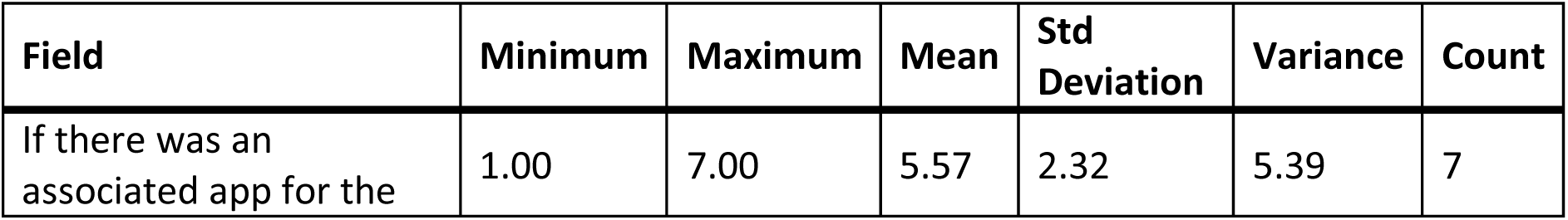
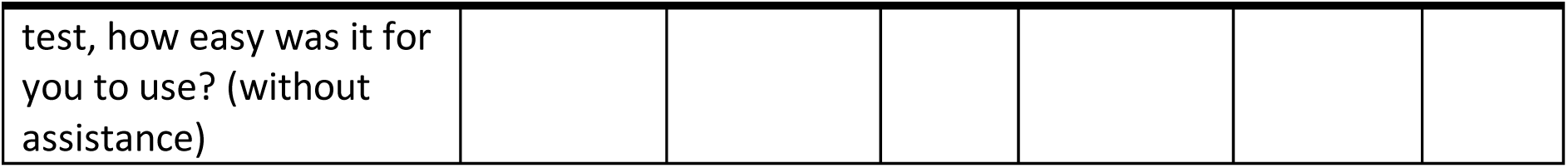
Stats for Q837: “If there was an associated app for the [Siemens Clinitest] test, how easy was it for you to use? (without assistance)”

**Table 358.** Responses to Q837: “If there was an associated app for the [Siemens Clinitest] test, how easy was it for you to use? (without assistance)”

| Answer | % | Count |
| --- | --- | --- |
| Very difficult | 14.29% | 1 |
| Difficult | 0.00% | 0 |
| Neither easy nor difficult | 14.29% | 1 |
| Easy | 0.00% | 0 |
| Very easy | 0.00% | 0 |
| There is an app, but I didn't use it | 71.43% | 5 |
| Total | 100% | 7 |

### Q838 - How helpful was the app in assisting you with completing the test?

**Table 359.** Stats for Q838: “How helpful was the app in assisting you with completing the [Siemens Clinitest] test?”

| Field | Minimum | Maximum | Mean | Std Deviation | Variance | Count |
| --- | --- | --- | --- | --- | --- | --- |
| How helpful was the app in assisting you with completing the test? | 3.00 | 4.00 | 3.50 | 0.50 | 0.25 | 2 |

**Table 360.** Responses to Q838: “How helpful was the app in assisting you with completing the [Siemens Clinitest] test?”

| Answer | % | Count |
| --- | --- | --- |
| Very unhelpful | 0.00% | 0 |
| Unhelpful | 0.00% | 0 |
| Neither unhelpful nor helpful | 50.00% | 1 |
| Helpful | 50.00% | 1 |
| Very helpful | 0.00% | 0 |
| Total | 100% | 2 |

### Q839 - Did you receive a valid test result with the Siemens Clinitest (positive or negative)?

**Table 361.** Stats for Q839: “Did you receive a valid test result with the Siemens Clinitest (positive or negative)?”

| Field | Minimum | Maximum | Mean | Std Deviation | Variance | Count |
| --- | --- | --- | --- | --- | --- | --- |
| Did you receive a valid test result with the Siemens Clinitest (positive or negative)? | 1.00 | 2.00 | 1.09 | 0.29 | 0.08 | 11 |

**Table 362.** Responses to Q839: “Did you receive a valid test result with the Siemens Clinitest (positive or negative)?”

| Answer | % | Count |
| --- | --- | --- |
| Yes | 90.91% | 10 |
| No | 9.09% | 1 |
| Total | 100% | 11 |

### Q840 - Did you attempt to contact Siemens Clinitest customer support?

**Table 363.** Stats for Q840: “Did you attempt to contact Siemens Clinitest customer support?”

| Field | Minimum | Maximum | Mean | Std Deviation | Variance | Count |
| --- | --- | --- | --- | --- | --- | --- |
| Did you attempt to contact Siemens Clinitest customer support? | 1.00 | 2.00 | 1.91 | 0.29 | 0.08 | 11 |

**Table 364.** Responses to Q840: “Did you attempt to contact Siemens Clinitest customer support?”

| Answer | % | Count |
| --- | --- | --- |
| Yes | 9.09% | 1 |
| No | 90.91% | 10 |
| Total | 100% | 11 |

### Q841 - Were you able to talk with customer support?

**Table 365.** Stats for Q841: “Were you able to talk with [Siemens Clinitest] customer support?”

| Field | Minimum | Maximum | Mean | Std Deviation | Variance | Count |
| --- | --- | --- | --- | --- | --- | --- |
| Were you able to talk with customer support? | 2.00 | 2.00 | 2.00 | 0.00 | 0.00 | 1 |

**Table 366.** Responses to Q841: “Were you able to talk with [Siemens Clinitest] customer support?”

| Answer | % | Count |
| --- | --- | --- |
| Yes | 0.00% | 0 |
| No | 100.00% | 1 |
| Total | 100% | 1 |

### Q842 - Was customer support helpful in answering your question(s)?

**Table 367.** Stats for Q842: “Was [Siemens Clinitest] customer support helpful in answering your question(s)?”

| Field | Minimum | Maximum | Mean | Std Deviation | Variance | Count |
| --- | --- | --- | --- | --- | --- | --- |
| Was customer support helpful in answering your question(s)? | 0.00 | 0.00 | 0.00 | 0.00 | 0.00 | 0 |

**Table 368.**
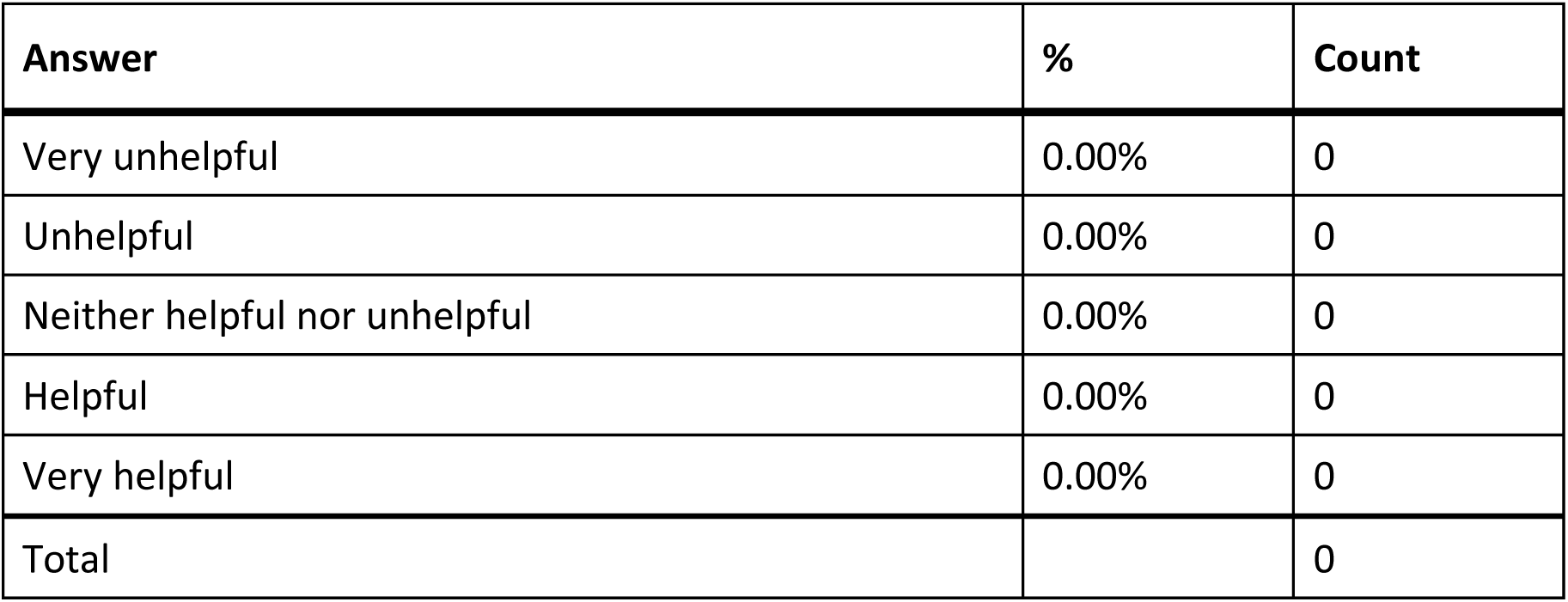
Responses to Q842: “Was [Siemens Clinitest] customer support helpful in answering your question(s)?”

### Q843 - If you have performed the Siemens Clinitest multiple times, how did your experience change?

**Table 369.** Stats for Q843: “If you have performed the Siemens Clinitest multiple times, how did you experience change?”

| Field | Minimum | Maximum | Mean | Std Deviation | Variance | Count |
| --- | --- | --- | --- | --- | --- | --- |
| If you have performed the Siemens Clinitest multiple times, how did your experience change? | 3.00 | 4.00 | 3.29 | 0.45 | 0.20 | 7 |

**Table 370.** Responses to Q843: “If you have performed the Siemens Clinitest multiple times, how did you experience change?”

| Answer | % | Count |
| --- | --- | --- |
| Much worse | 0.00% | 0 |
| Worse | 0.00% | 0 |
| Neither better nor worse | 71.43% | 5 |
| Better | 28.57% | 2 |
| Much better | 0.00% | 0 |
| Total | 100% | 7 |

### Q844 - What did you like about the Siemens Clinitest?

**Table 371.** Open responses to Q844: “What did you like about the Siemens Clinitest?”

| What did you like about the Siemens Clinitest? |
| --- |
| The holder for the test vial built into the box. |
| Hi |
| After I did it the first time, it was easy to do each progressive time. |
| Simple |
| Multiple tests per package. |
| Ease of use |
| I was able to perform the test very easily with the assistance of Aira. |

### Q845 - What would you like to see improved for the Siemens Clinitest?

**Table 372.**
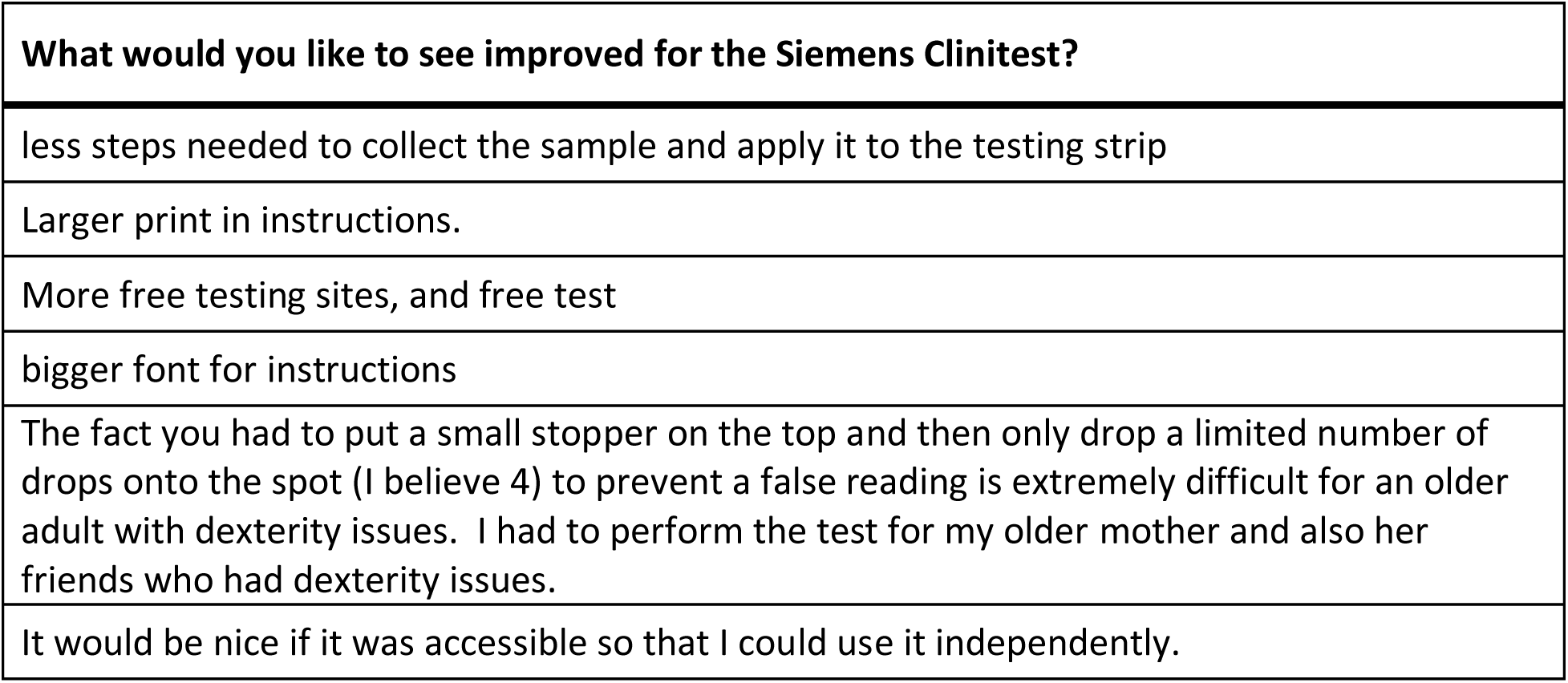
Open responses to Q845: “What would you like to see improved for the Siemens Clinitest?”

| What would you like to see improved for the Siemens Clinitest? |
| --- |
| less steps needed to collect the sample and apply it to the testing strip |
| Larger print in instructions. |
| More free testing sites, and free test |
| bigger font for instructions |
| The fact you had to put a small stopper on the top and then only drop a limited number of drops onto the spot (I believe 4) to prevent a false reading is extremely difficult for an older adult with dexterity issues. I had to perform the test for my older mother and also her friends who had dexterity issues. |
| It would be nice if it was accessible so that I could use it independently. |

### Q867 - [QID4-ChoiceTextEntryValue-10] The following questions ask about the Other test you indicated “[QID4-ChoiceTextEntryValue-10].” Think of the first time you used the [QID4-ChoiceTextEntryValue-10] test. Did you have difficulty completing any of the following tasks? Select all that apply

**Table 373.** Responses to Q867: Other tests - “Did you have difficulty completing any of the following tasks?”

| Answer | % | Count |
| --- | --- | --- |
| Opening the packaging | 8.43% | 7 |
| Reading the instructions | 20.48% | 17 |
| Identifying test components | 14.46% | 12 |
| Collecting the test sample | 9.64% | 8 |
| Manipulating test components | 12.05% | 10 |
| Interpreting the test results | 15.66% | 13 |
| Other | 4.82% | 4 |
| I did not have difficulty completing the test | 14.46% | 12 |
| Total | 100% | 83 |

#### Q867_7_TEXT – Other

**Table 374.**
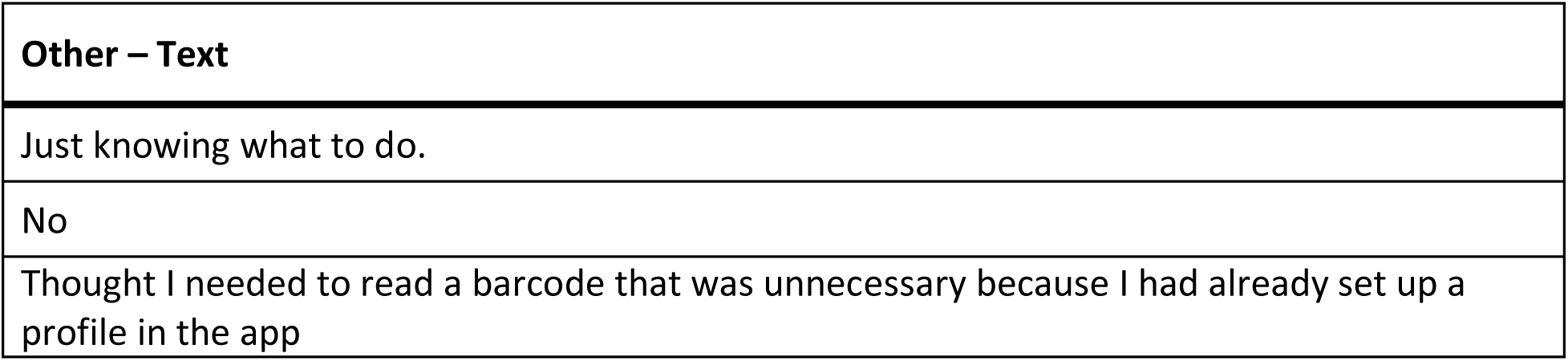
Open responses to Other - Q867: “Other tests - Did you have difficulty completing any of the following tasks?”

|  |
| --- |
| **Other – Text** |
| Just knowing what to do. |
| No |
| Thought I needed to read a barcode that was unnecessary because I had already set up a profile in the app |

### Q868 - What tools or assistance, if any, did you use to perform the test?

**Table 375.**
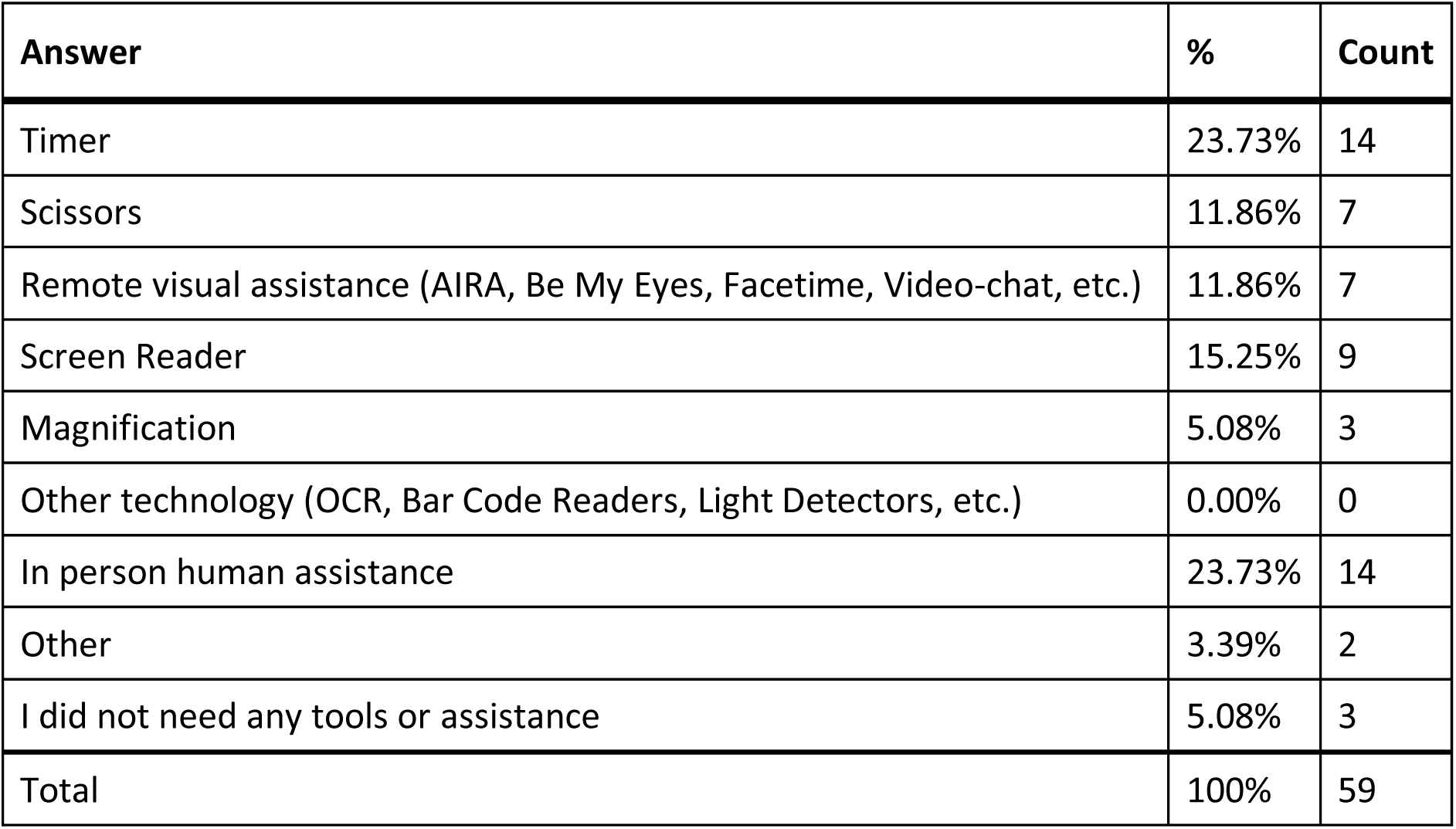
Responses to Q868: “What tools or assistance, if any, did you use to perform the [Other] test?”

#### Q868_8_TEXT – Other

**Table 376.**
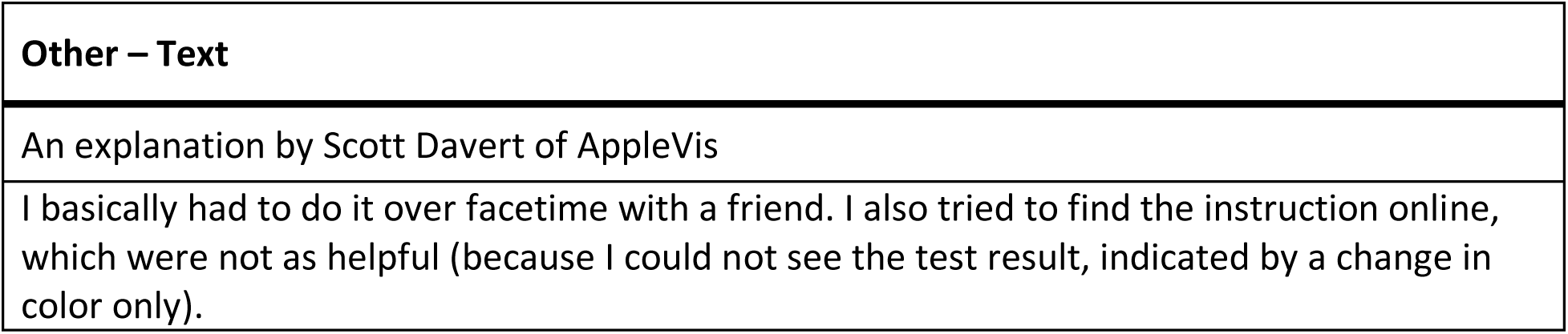
Open responses to Other - Q868: “What tools or assistance, if any, did you use to perform the [Other] test?”

### Q869 - Were you able to conduct this test on your first try?

**Table 377.** Stats for Q869: “Were you able to conduct this [Other] test on your first try?”

| Field | Minimum | Maximum | Mean | Std Deviation | Variance | Count |
| --- | --- | --- | --- | --- | --- | --- |
| Were you able to conduct this test on your first try? | 1.00 | 2.00 | 1.09 | 0.28 | 0.08 | 34 |

**Table 378.**
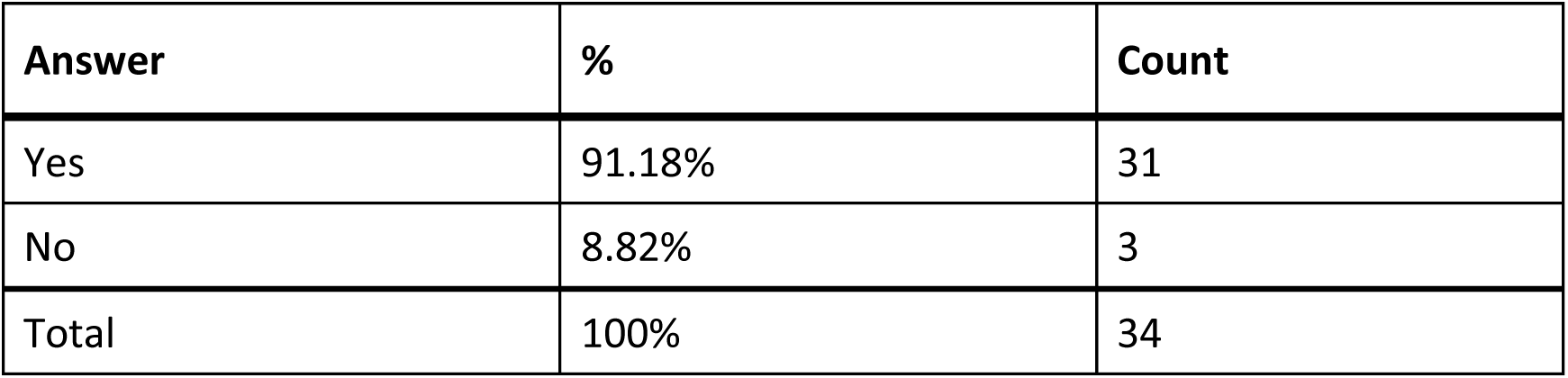
Responses to Q869: “Were you able to conduct this [Other] test on your first try?”

### Q870 - How long did it take to complete the steps required to perform the test, not including the time you spent waiting for results

**Table 379.** Stats for Q870: “How long did it take to complete the steps required to perform the [Other] test?”

| Field | Minimum | Maximum | Mean | Std Deviation | Variance | Count |
| --- | --- | --- | --- | --- | --- | --- |
| How long did it take to complete the steps required to perform the test, not including the time you spent waiting for results. | 1.00 | 4.00 | 2.15 | 1.17 | 1.36 | 34 |

**Table 380.** Responses to Q870: “How long did it take to complete the steps required to perform the [Other] test?”

| Answer | % | Count |
| --- | --- | --- |
| Under 5 minutes | 38.24% | 13 |
| 5 - 10 minutes | 32.35% | 11 |
| 10 - 15 minutes | 5.88% | 2 |
| Over 15 minutes | 23.53% | 8 |
| Total | 100% | 34 |

### Q871 - What format of instructions did you use?

**Table 381.** Responses to Q871: “What format of [Other] instructions did you use?”

| Answer | % | Count |
| --- | --- | --- |
| Paper | 45.00% | 18 |
| Digital (webpages, downloaded, etc.) | 17.50% | 7 |
| None of the above | 15.00% | 6 |
| Associated Covid Test App (Smartphone) | 22.50% | 9 |
| Total | 100% | 40 |

### Q872 - Were you able to access electronic information via a QR Code?

**Table 382.** Stats for Q872: “Were you able to access electronic information via a [Other test] QR Code?”

| Field | Minimum | Maximum | Mean | Std Deviation | Variance | Count |
| --- | --- | --- | --- | --- | --- | --- |
| Were you able to access electronic information via a QR Code? | 1.00 | 7.00 | 3.73 | 2.99 | 8.93 | 11 |

**Table 383.** Responses to Q872: “Were you able to access electronic information via a [Other test] QR Code?”

| Answer | % | Count |
| --- | --- | --- |
| Yes | 54.55% | 6 |
| No | 45.45% | 5 |
| Total | 100% | 11 |

### Q873 - How easy was opening the package of the [QID4-ChoiceTextEntryValue-10] test for you? (without assistance)

**Table 384.** Stats for Q873: “How easy was opening the package of the [Other] test for you? (without assistance)”

| Field | Minimum | Maximum | Mean | Std Deviation | Variance | Count |
| --- | --- | --- | --- | --- | --- | --- |
| How easy was opening the package of the [QID4-ChoiceTextEntryValue-10] test for you? (without assistance) | 1.00 | 5.00 | 3.66 | 1.11 | 1.23 | 32 |

**Table 385.** Responses to Q873: “How easy was opening the package of the [Other] test for you? (without assistance)”

| Answer | % | Count |
| --- | --- | --- |
| Very difficult | 3.13% | 1 |
| Difficult | 15.63% | 5 |
| Neither easy nor difficult | 18.75% | 6 |
| Easy | 37.50% | 12 |
| Very easy | 25.00% | 8 |
| Total | 100% | 32 |

### Q874 - How easy was completing the test protocol for you? (without assistance)

**Table 386.** Stats for Q874: “How easy was completing the [Other] test protocol for you? (without assistance)”

| Field | Minimum | Maximum | Mean | Std Deviation | Variance | Count |
| --- | --- | --- | --- | --- | --- | --- |
| How easy was completing the test protocol for you? (without assistance) | 1.00 | 5.00 | 2.66 | 1.58 | 2.50 | 29 |

**Table 387.** Responses to Q874: “How easy was completing the [Other] test protocol for you? (without assistance)”

| Answer | % | Count |
| --- | --- | --- |
| Very difficult | 44.83% | 13 |
| Difficult | 0.00% | 0 |
| Neither easy nor difficult | 13.79% | 4 |
| Easy | 27.59% | 8 |
| Very easy | 13.79% | 4 |
| Total | 100% | 29 |

### Q875 - How easy was interpreting the test results for you? (without assistance)

**Table 388.** Stats for Q875: “How easy was interpreting the [Other] test results for you? (without assistance)”

| Field | Minimum | Maximum | Mean | Std Deviation | Variance | Count |
| --- | --- | --- | --- | --- | --- | --- |
| How easy was interpreting the test results for you? (without assistance) | 1.00 | 5.00 | 2.97 | 1.62 | 2.63 | 30 |

**Table 389.** Responses to Q875: “How easy was interpreting the [Other] test results for you? (without assistance)”

| Answer | % | Count |
| --- | --- | --- |
| Very difficult | 36.67% | 11 |
| Difficult | 0.00% | 0 |
| Neither easy nor difficult | 16.67% | 5 |
| Easy | 23.33% | 7 |
| Very easy | 23.33% | 7 |
| Total | 100% | 30 |

### Q876 - Were you able to interpret the test results? (without assistance)

**Table 390.** Stats for Q876: “Were you able to interpret the [Other] test results? (without assistance)”

| Field | Minimum | Maximum | Mean | Std Deviation | Variance | Count |
| --- | --- | --- | --- | --- | --- | --- |
| Were you able to interpret the test results? (without assistance) | 1.00 | 2.00 | 1.45 | 0.50 | 0.25 | 33 |

**Table 391.** Responses to Q876: “Were you able to interpret the [Other] test results? (without assistance)”

| Answer | % | Count |
| --- | --- | --- |
| Yes | 54.55% | 18 |
| No | 45.45% | 15 |
| Total | 100% | 33 |

### Q877 - If there was an associated app for the test, how easy was it for you to use? (without assistance)

**Table 392.** Stats for Q877: “If there was an associated app for the [Other] test, how easy was it for you to use? (without assistance)”

| Field | Minimum | Maximum | Mean | Std Deviation | Variance | Count |
| --- | --- | --- | --- | --- | --- | --- |
| If there was an associated app for the test, how easy was it for you to use? (without assistance) | 3.00 | 7.00 | 4.67 | 1.25 | 1.56 | 12 |

**Table 393.** Responses to Q877: “If there was an associated app for the [Other] test, how easy was it for you to use? (without assistance)”

| Answer | % | Count |
| --- | --- | --- |
| Very difficult | 0.00% | 0 |
| Difficult | 0.00% | 0 |
| Neither easy nor difficult | 16.67% | 2 |
| Easy | 33.33% | 4 |
| Very easy | 33.33% | 4 |
| There is an app, but I didn't use it | 16.67% | 2 |
| Total | 100% | 12 |

### Q878 - How helpful was the app in assisting you with completing the test?

**Table 394.** Stats for Q878: “How helpful was the app in assisting you with completing the [Other] test?”

| Field | Minimum | Maximum | Mean | Std Deviation | Variance | Count |
| --- | --- | --- | --- | --- | --- | --- |
| How helpful was the app in assisting you with completing the test? | 1.00 | 5.00 | 4.11 | 1.20 | 1.43 | 9 |

**Table 395.** Responses to Q878: “How helpful was the app in assisting you with completing the [Other] test?”

| Answer | % | Count |
| --- | --- | --- |
| Very unhelpful | 11.11% | 1 |
| Unhelpful | 0.00% | 0 |
| Neither unhelpful nor helpful | 0.00% | 0 |
| Helpful | 44.44% | 4 |
| Very helpful | 44.44% | 4 |
| Total | 100% | 9 |

### Q879 - Did you receive a valid test result with the [QID4-ChoiceTextEntryValue-10] test (positive or negative)?

**Table 396.** Stats for Q879: “Did you receive a valid test result with the [Other] test (positive or negative)?”

| Field | Minimum | Maximum | Mean | Std Deviation | Variance | Count |
| --- | --- | --- | --- | --- | --- | --- |
| Did you receive a valid test result with the [QID4-ChoiceTextEntryValue-10] test (positive or negative)? | 1.00 | 1.00 | 1.00 | 0.00 | 0.00 | 32 |

**Table 397.**
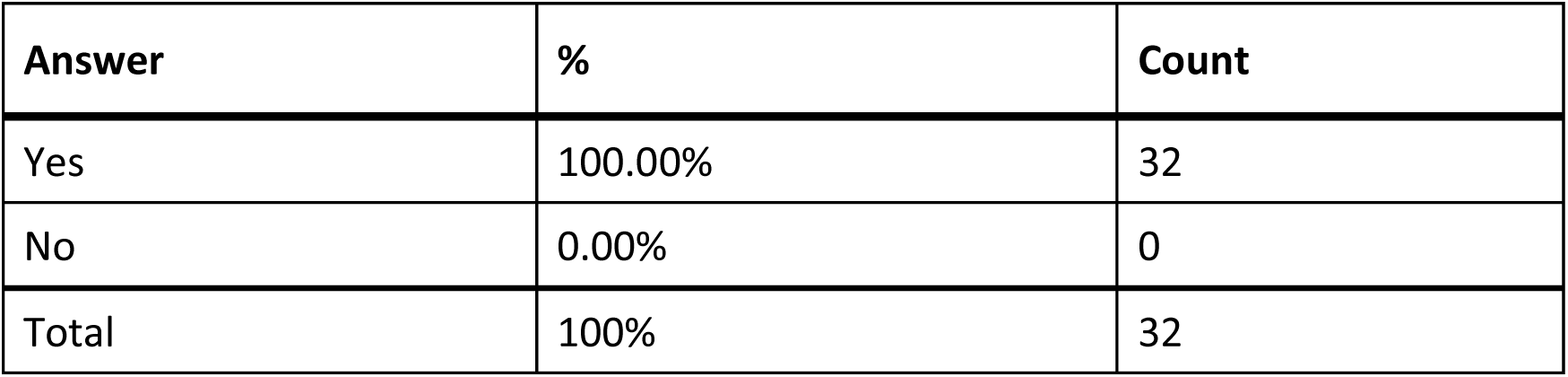
Responses to Q879: “Did you receive a valid test result with the [Other] test (positive or negative)?”

| Answer | % | Count |
| --- | --- | --- |
| Yes | 100.00% | 32 |
| No | 0.00% | 0 |
| Total | 100% | 32 |

### Q880 - Did you attempt to contact [QID4-ChoiceTextEntryValue-10] test customer support?

**Table 398.**
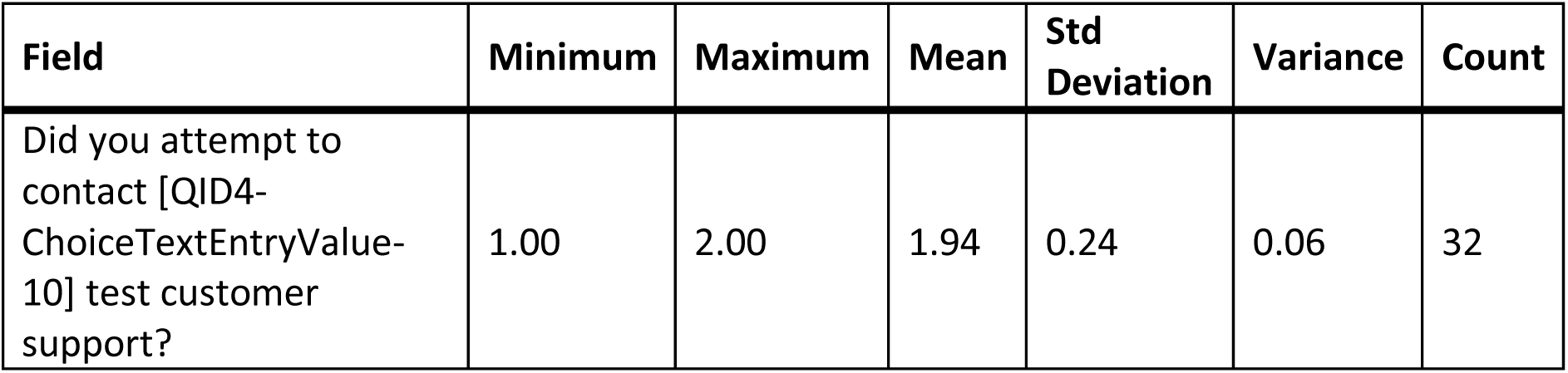
Stats for Q880: “Did you attempt to contact [Other] test customer support?”

**Table 399.** Responses to Q880: “Did you attempt to contact [Other] test customer support?”

| Answer | % | Count |
| --- | --- | --- |
| Yes | 6.25% | 2 |
| No | 93.75% | 30 |
| Total | 100% | 32 |

### Q881 - Were you able to talk with customer support?

**Table 400.** Stats for Q881: “Were you able to talk with [Other] customer support?”

| Field | Minimum | Maximum | Mean | Std Deviation | Variance | Count |
| --- | --- | --- | --- | --- | --- | --- |
| Were you able to talk with customer support? | 1.00 | 2.00 | 1.50 | 0.50 | 0.25 | 2 |

**Table 401.**
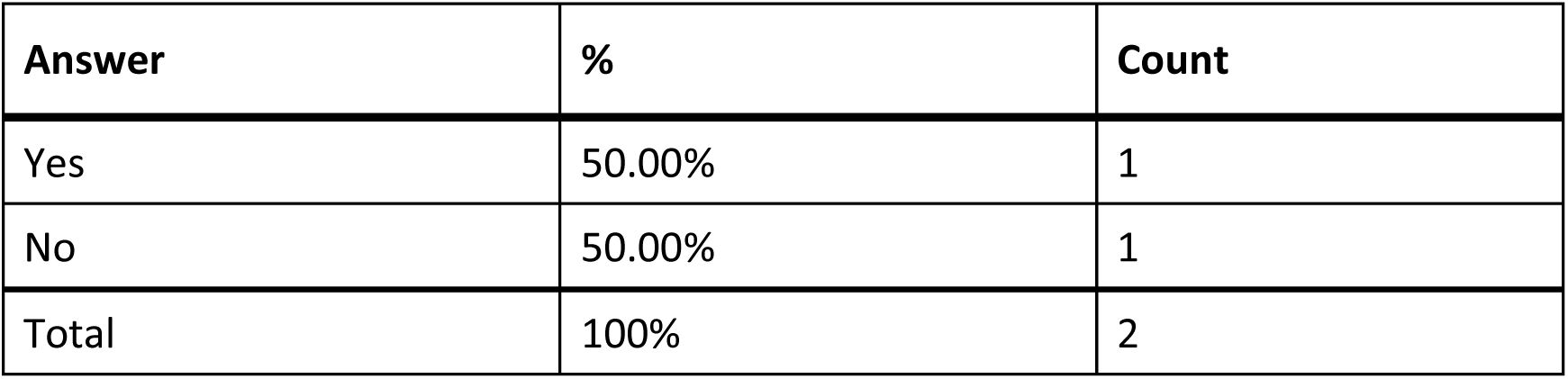
Responses to Q881: “Were you able to talk with [Other] customer support?”

### Q882 - Was customer support helpful in answering your question(s)?

**Table 402.** Stats for Q882: “Was [Other] customer support helpful in answering your question(s)?”

| Field | Minimum | Maximum | Mean | Std Deviation | Variance | Count |
| --- | --- | --- | --- | --- | --- | --- |
| Was customer support helpful in answering your question(s)? | 6.00 | 6.00 | 6.00 | 0.00 | 0.00 | 1 |

**Table 403.** Response to Q882: “Was [Other] customer support helpful in answering your question(s)?”

| Answer | % | Count |
| --- | --- | --- |
| Very unhelpful | 0.00% | 0 |
| Unhelpful | 0.00% | 0 |
| Neither helpful nor unhelpful | 0.00% | 0 |
| Helpful | 100.00% | 1 |
| Very helpful | 0.00% | 0 |
| Total | 100% | 1 |

### Q883 - If you have performed the [QID4-ChoiceTextEntryValue-10] test multiple times, how did your experience change?

**Table 404.**
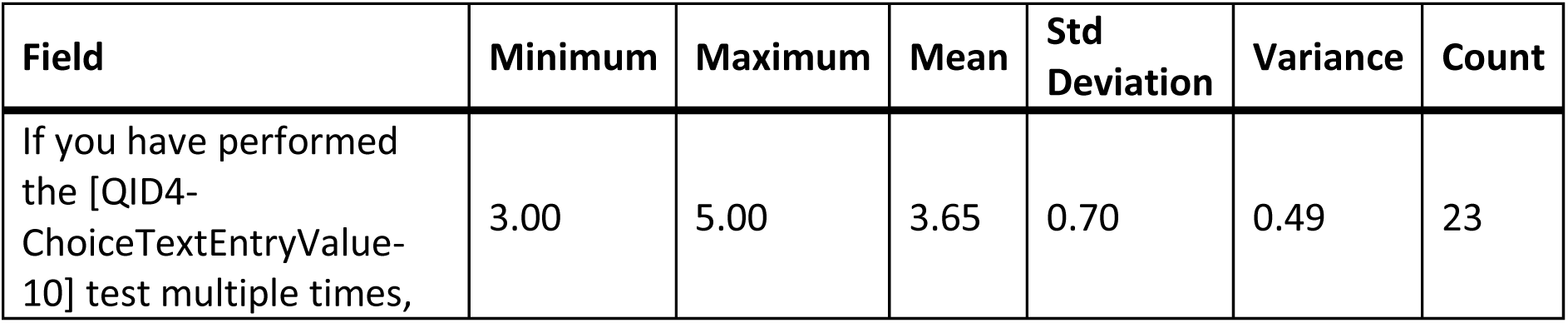
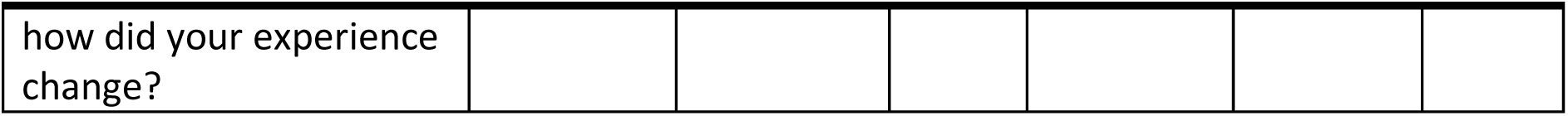
Stat for Q883: “If you have performed the [Other] test multiple times, how did your experience change?”

**Table 405.** Responses to Q883: “If you have performed the [Other] test multiple times, how did you experience change?”

| Answer | % | Count |
| --- | --- | --- |
| Much worse | 0.00% | 0 |
| Worse | 0.00% | 0 |
| Neither better nor worse | 47.83% | 11 |
| Better | 39.13% | 9 |
| Much better | 13.04% | 3 |
| Total | 100% | 23 |

### Q884 - What did you like about the [QID4-ChoiceTextEntryValue-10] test?

**Table 406.** Open responses to Q884: “What did you like about the [Other] test?”

| What did you like about the [QID4-ChoiceTextEntryValue-10] test? |
| --- |
| N/A |
| N/A |
| There wasn't anything to like about the test, it was just to verify results of the test. |
| That I could use at home and it was relatively fast |
| Ease of use |
| Free |
| It's super accessible and simple to use. |
| You could perform them right from your home |
| I liked it that my sighted friend could help me with them.u |
| The Cue Health NAAT test is the only test system that I know of that is fully accessible to blind people. It is easy and straightforward to use. The app is accessible and compatible with the VoiceOver iPhone screen reader. |
| I didn't have to go to a clinic and have it done. I could do it easily at home. |
| That it was quick |
| quick results |
| It works for those who do not wish to disclose any information or use technology. It is inaccessible. |
| Compact, streamlined use |
| easy to use integrated well with smart phone, app is accessible. |
| Only have to read one code number to register kit and later kits have code number pre-populated in online registration form. |
| Not having to add solution in the cartage |
| As a clarification, I listed multiple types of tests in this answer. Cue Health tests are very accessible and I did not answer the questions about those tests. I answer the questions about tests I bought at my local pharmacy and got from the US government. I like that despite some brand differences, the components tended to be similar so once I learned how to conduct one inaccessible test brand on my own I could pretty well figure out how to conduct other brands' tests on my own and I only needed to do a video call to look at the results. I like that the tube of liquid requires squeezing before it pours into the test strip. I had been nervous that I would spill it in the wrong place and this feature prevented me from doing that. |
| The app simplifies the entire process, and the results can be shared through the app, by email or other means. |
| Both seem to somewhat the same fuction |
| I didn't like much about it, since I had to have someone sighted available to assist. |
| seemed to be pretty straight forward in terms of the instructions |
| I liked the test I took because everything was packaged separately and I used "be my eyes" to administer the test. |

### Q885 - What would you like to see improved for the [QID4- ChoiceTextEntryValue-10] test?

**Table 407.** Open responses to Q885: “What would you like to see improved for the [Other] test?”

| What would you like to see improved for the [QID4-ChoiceTextEntryValue-10] test? |
| --- |
| N/A |
| N/A |
| less packing - WiFi capability through an app. |
| Some instructions, and perhaps being able to use it individually without personal assistant |
| Less product/packaging waste |
| better accuracy |
| The only difficulty I had with the Cue Health test was connecting the reader to the app. I think it would be helpful if it had tactile feedback for finding the QR Code. |
| Compatible results if there are new strains of Covid |
| My sighted friend who was helping me is dyslexic. I had to magnify the instructions and read them to him. This was very hard to do |
| The only downside of the Cue Health test system is the cost. It is much more expensive than other at-home COVID tests. The reader device costs \$250, and each test kit is \$75. |
| Have a chip in it so it makes a sound if negative and a different sound if it is positive |
| I would like to be able to do it on my own without assistance. |
| I can pour solution into something but cannot determine if it is at a designated line and have no idea how to measure five drops and get that solution where it needs to be without contaminating the sample. |
| ???? |
| Detector of results |
| Na |
| Make it more accessible for visually impaired individuals |
| Nonvisual way to register kit. Faster processing of PCR test once received by LabCorp lab. An app to get results rather than just a web site. |
| Not lose accessibility of the app |
| Well advertised digital instructions and a way to read the result by touch. |
| n/a |
| Maybe less materials if possible |
| I don't think there was an app. I totally got help w/the test. |
| I would like to see some tactile indication and perhaps a phone number I could call that would read the directions to me. Just test took entirely too long to administer so I am very glad that be my eyes was there. I would also like for the description by phone to describe each piece of the test components described specifically so I would know exactly what I had and what I needed to do with it. I would also like to see something tactile or maybe a heat sensor of some kind to tell me whether the test was positive or end or negative so I could administer it independently |

### Q846 - For the unknown brand of COVID-19 test you’ve used, did you have difficulty completing any of the following tasks? Select all that apply

**Table 408.**
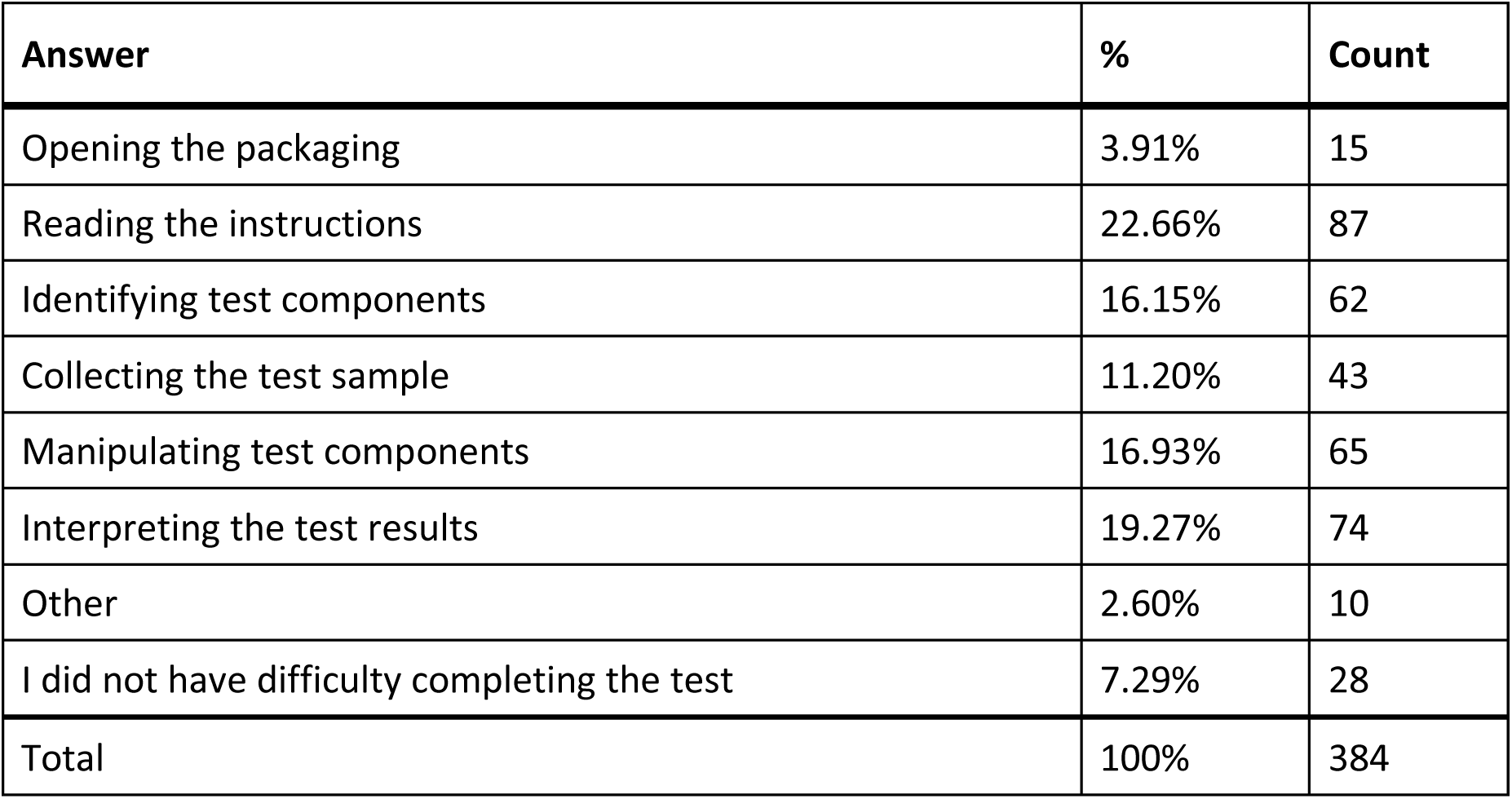
Responses to Q846: “For the unknown brand of COVID-19 test you’ve used, did you have any difficulty completing the following tasks?”

#### Q846_7_TEXT – Other

**Table 409.**
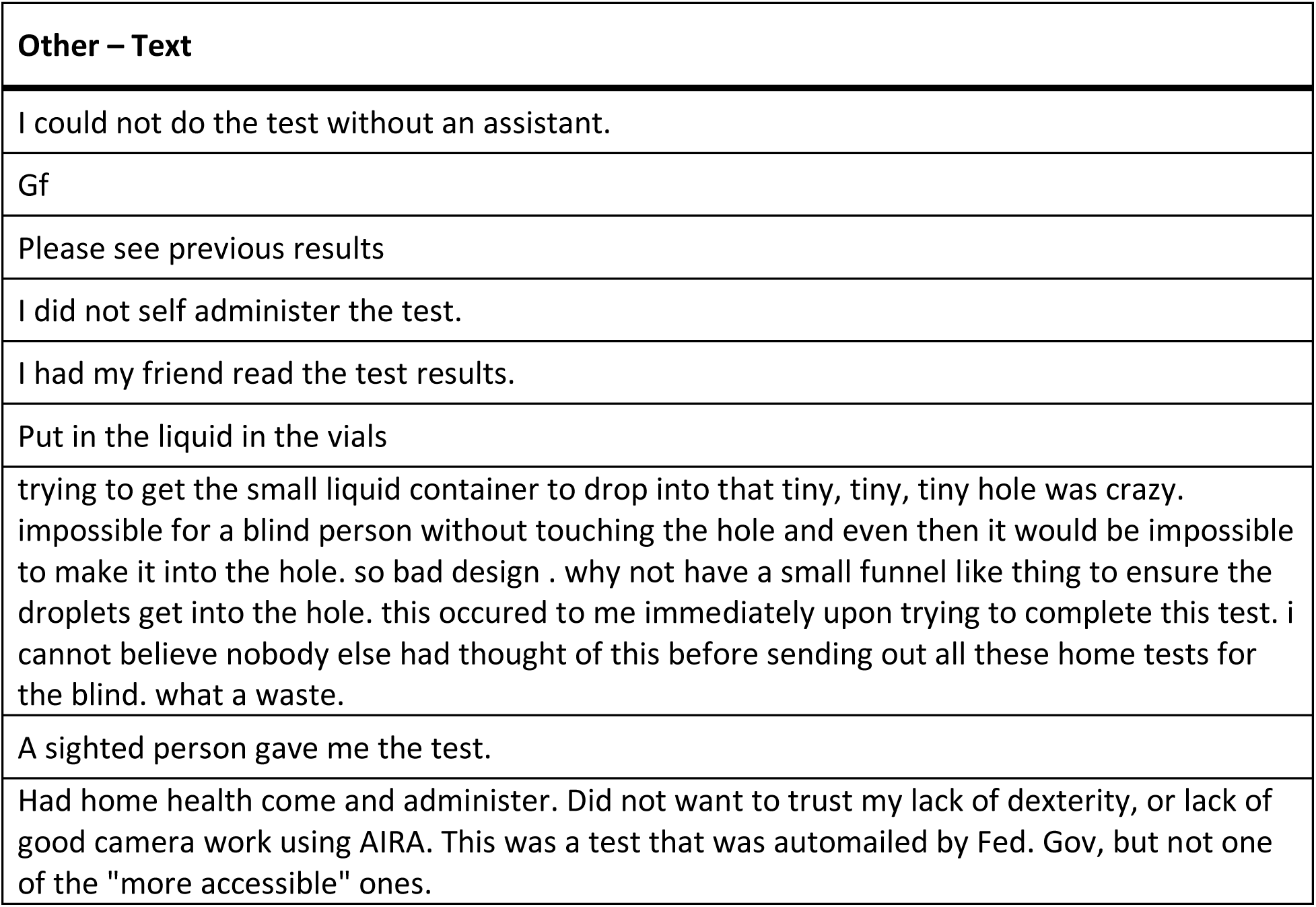
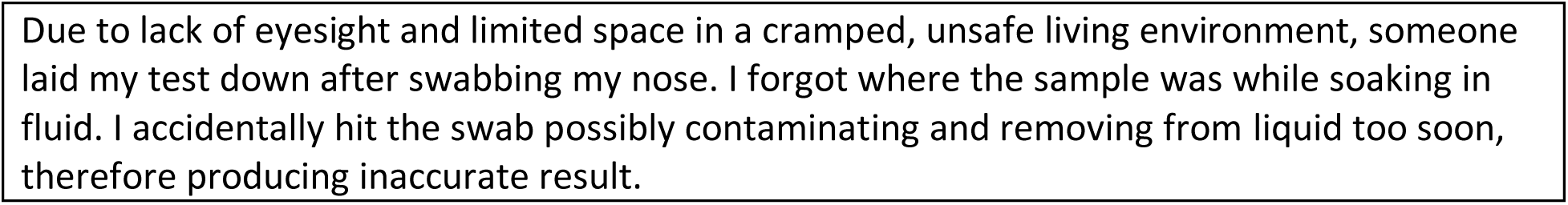
Open response for Other - Q846: “Unknown test - Did you have any difficulty completing the following tasks?”

### Q847 - What tools or assistance, if any, did you use to perform the test?

**Table 410.** Responses to Q847: “What tools or assistance, if any, did you use to perform the [unknown] test?”

| Answer | % | Count |
| --- | --- | --- |
| Timer | 20.95% | 53 |
| Scissors | 15.81% | 40 |
| Remote visual assistance (AIRA, Be My Eyes, Facetime, Video-chat, etc.) | 11.86% | 30 |
| Screen Reader | 8.70% | 22 |
| Magnification | 5.14% | 13 |
| Other technology (OCR, Bar Code Readers, Light Detectors, etc.) | 1.19% | 3 |
| In person human assistance | 30.04% | 76 |
| Other | 1.98% | 5 |
| I did not need any tools or assistance | 4.35% | 11 |
| Total | 100% | 253 |

#### Q847_8_TEXT – Other

**Table 411.** Open responses to Other - Q847: “What tools or other assistance, if any, did you use to perform the [unknown] test?”

| Other – Text |
| --- |
| My husband assisted. |
| FaceTime friend or family. |
| My cell phone camera to enlarge the instructions in order to be able to read them. |
| Used my phone to take picture. |
| Would use timer, remote assistance and screen reader rather than directt human assistance, if successful interpretation of result could be achieved. Would use scissors if needed. |

### Q848 - Were you able to conduct this test on your first try?

**Table 412.** Stats for Q848: “Were you able to conduct this [unknown] test on your first try?”

| Field | Minimum | Maximum | Mean | Std Deviation | Variance | Count |
| --- | --- | --- | --- | --- | --- | --- |
| Were you able to conduct this test on your first try? | 1.00 | 2.00 | 1.26 | 0.44 | 0.19 | 130 |

**Table 413:** Responses to Q848: “Were you able to conduct this [unknown] test on your first try?”

| Answer | % | Count |
| --- | --- | --- |
| Yes | 73.85% | 96 |
| No | 26.15% | 34 |
| Total | 100% | 130 |

### Q849 - How long did it take to complete the steps required to perform the test, not including the time you spent waiting for results

**Table 414:** Stats for Q849: “How long did it take to complete the steps required to perform the [unknown] test?”

| Field | Minimum | Maximum | Mean | Std Deviation | Variance | Count |
| --- | --- | --- | --- | --- | --- | --- |
| How long did it take to complete the steps required to perform the test, not including the time you spent waiting for results. | 1.00 | 4.00 | 2.53 | 1.08 | 1.17 | 130 |

**Table 415.** Responses to Q849: “How long did it take to complete the steps required to perform the [unknown] test?”

| Answer | % | Count |
| --- | --- | --- |
| Under 5 minutes | 18.46% | 24 |
| 5 - 10 minutes | 37.69% | 49 |
| 10 - 15 minutes | 16.15% | 21 |
| Over 15 minutes | 27.69% | 36 |
| Total | 100% | 130 |

### Q850 - What format of instructions did you use?

**Table 416.**
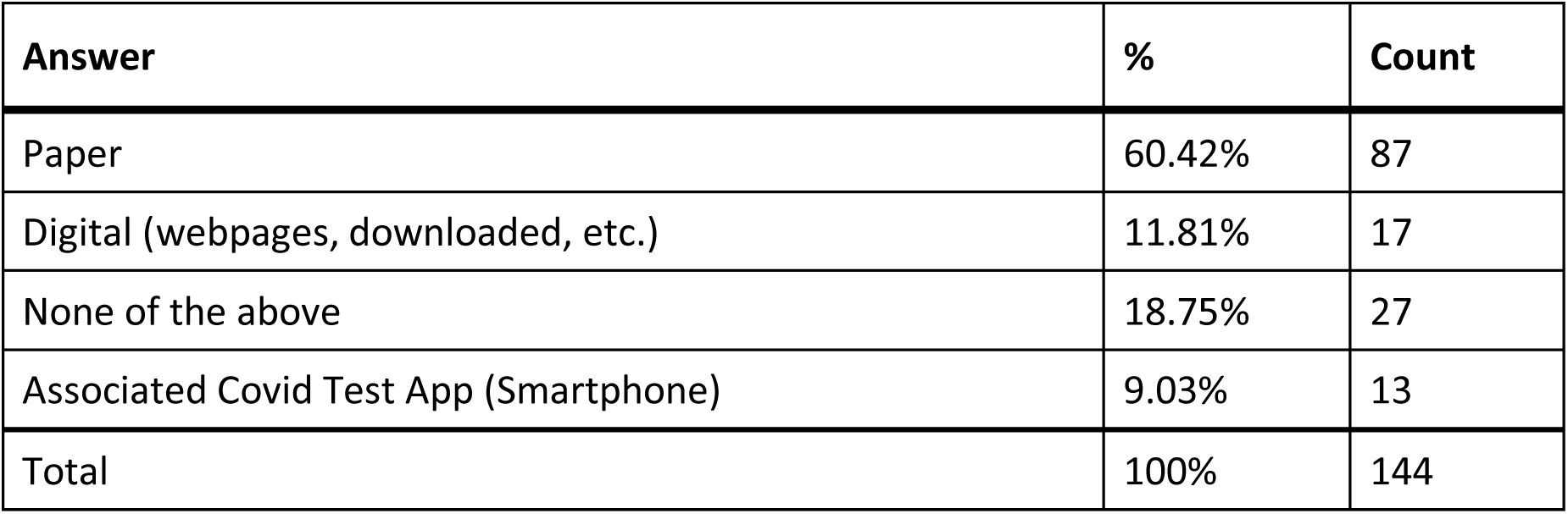
Responses to Q850: “What format of [unknown test] instructions did you use?”

### Q851 - Were you able to access electronic information via a QR Code?

**Table 417.** Stats for Q851: “Were you able to access electronic information via a [unknown test] QR Code?”

| Field | Minimum | Maximum | Mean | Std Deviation | Variance | Count |
| --- | --- | --- | --- | --- | --- | --- |
| Were you able to access electronic information via a QR Code? | 1.00 | 7.00 | 4.53 | 2.95 | 8.72 | 17 |

**Table 418.** Responses to Q851: “Were you able to access electronic information via a [unknown test] QR Code?”

| Answer | % | Count |
| --- | --- | --- |
| Yes | 41.18% | 7 |
| No | 58.82% | 10 |
| Total | 100% | 17 |

### Q852 - How easy was opening the package of your COVID-19 home test for you? (without assistance)

**Table 419.**
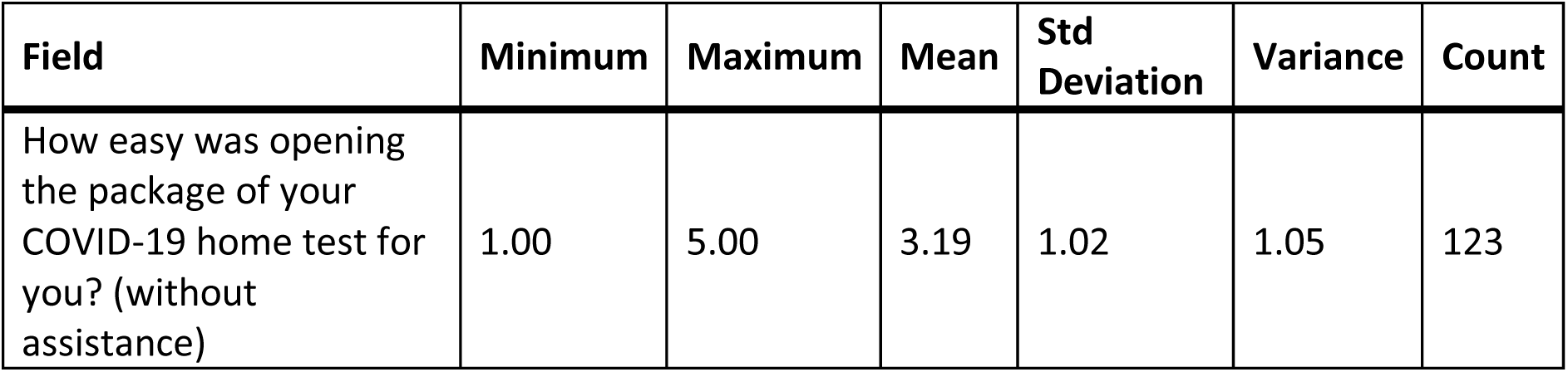
Stats for Q852: “How easy was opening the package of your [unknown] COVID-19 home test for you? (without assistance)”

| Field | Minimum | Maximum | Mean | Std Deviation | Variance | Count |
| --- | --- | --- | --- | --- | --- | --- |
| How easy was opening the package of your COVID-19 home test for you? (without assistance) | 1.00 | 5.00 | 3.19 | 1.02 | 1.05 | 123 |

**Table 420.** Responses to Q852: “How easy was opening the package of your [unknown] COVID- 19 home test for you? (without assistance)”

| Answer | % | Count |
| --- | --- | --- |
| Very difficult | 4.07% | 5 |
| Difficult | 20.33% | 25 |
| Neither easy nor difficult | 40.65% | 50 |
| Easy | 22.76% | 28 |
| Very easy | 12.20% | 15 |
| Total | 100% | 123 |

### Q853 - How easy was completing the test protocol for you? (without assistance)

**Table 421.** Stats for Q853: “How easy was completing the [unknown] test protocol for you? (without assistance)”

| Field | Minimum | Maximum | Mean | Std Deviation | Variance | Count |
| --- | --- | --- | --- | --- | --- | --- |
| How easy was completing the test protocol for you? (without assistance) | 1.00 | 5.00 | 2.05 | 1.22 | 1.50 | 124 |

**Table 422.** Responses to Q853: “How easy was completing the [unknown] test protocol for you? (without assistance)”

| Answer | % | Count |
| --- | --- | --- |
| Very difficult | 46.77% | 58 |
| Difficult | 22.58% | 28 |
| Neither easy nor difficult | 14.52% | 18 |
| Easy | 11.29% | 14 |
| Very easy | 4.84% | 6 |
| Total | 100% | 124 |

### Q854 - How easy was interpreting the test results for you? (without assistance)

**Table 423.** Stats for Q854: “How easy was interpreting the [unknown] test results for you? (without assistance)”

| Field | Minimum | Maximum | Mean | Std Deviation | Variance | Count |
| --- | --- | --- | --- | --- | --- | --- |
| How easy was interpreting the test results for you? (without assistance) | 1.00 | 5.00 | 2.19 | 1.44 | 2.09 | 124 |

**Table 424.** Responses to Q854: “How easy was interpreting the [unknown] test results for you? (without assistance)”

| Answer | % | Count |
| --- | --- | --- |
| Very difficult | 52.42% | 65 |
| Difficult | 11.29% | 14 |
| Neither easy nor difficult | 11.29% | 14 |
| Easy | 15.32% | 19 |
| Very easy | 9.68% | 12 |
| Total | 100% | 124 |

### Q855 - Were you able to interpret the test results? (without assistance)

**Table 425.**
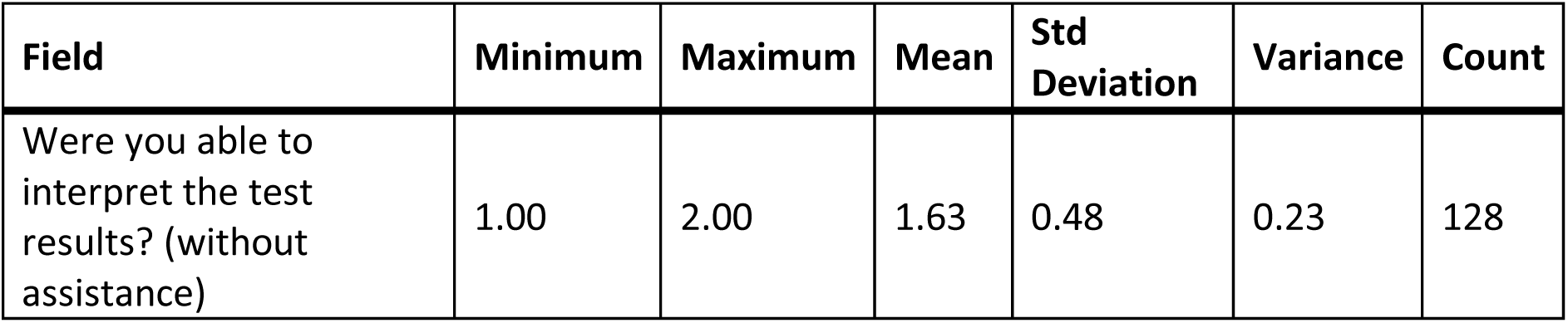
Stats for Q855: “Were you able to interpret the [unknown] test results? (without assistance)”

| Field | Minimum | Maximum | Mean | Std Deviation | Variance | Count |
| --- | --- | --- | --- | --- | --- | --- |
| Were you able to interpret the test results? (without assistance) | 1.00 | 2.00 | 1.63 | 0.48 | 0.23 | 128 |

**Table 426.** Responses to Q855: “Were you able to interpret the [unknown] test results? (without assistance)”

| Answer | % | Count |
| --- | --- | --- |
| Yes | 37.50% | 48 |
| No | 62.50% | 80 |
| Total | 100% | 128 |

### Q856 - If there was an associated app for the test, how easy was it for you to use? (without assistance)

**Table 427.**
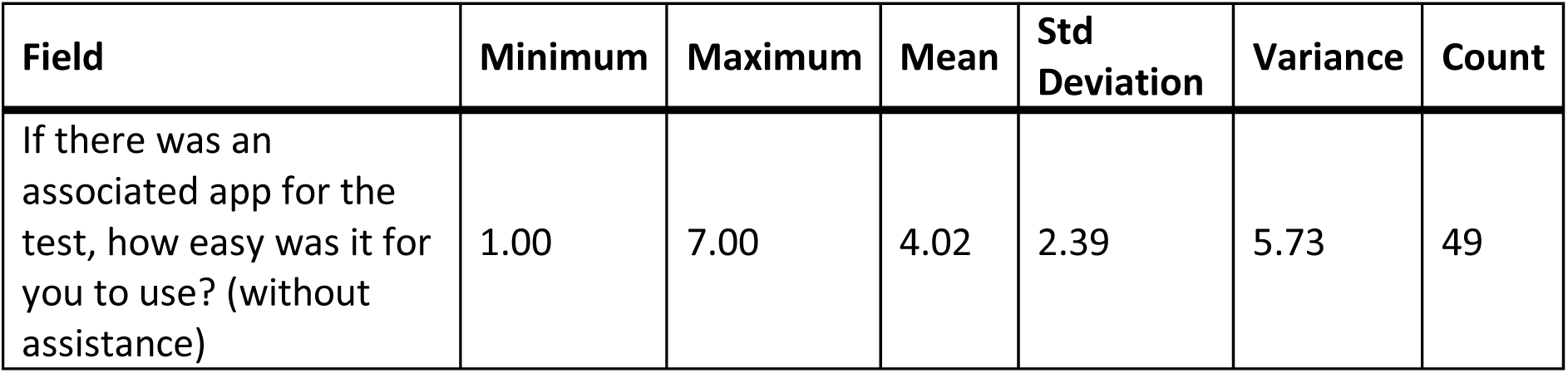
Stats for Q856: “If there was an associated app for the [unknown] test, how easy was it for you to use? (without assistance)”

| Field | Minimum | Maximum | Mean | Std Deviation | Variance | Count |
| --- | --- | --- | --- | --- | --- | --- |
| If there was an associated app for the test, how easy was it for you to use? (without assistance) | 1.00 | 7.00 | 4.02 | 2.39 | 5.73 | 49 |

**Table 428.** Responses to Q856: “If there was an associated app for the [unknown] test, how easy was it for you to use? (without assistance)”

| Answer | % | Count |
| --- | --- | --- |
| Very difficult | 20.41% | 10 |
| Difficult | 14.29% | 7 |
| Neither easy nor difficult | 16.33% | 8 |
| Easy | 10.20% | 5 |
| Very easy | 4.08% | 2 |
| There is an app, but I didn't use it | 34.69% | 17 |
| Total | 100% | 49 |

### Q857 - How helpful was the app in assisting you with completing the test?

**Table 429.** Stats for Q857: “How helpful was the app in assisting you with completing the [unknown] test?”

| Field | Minimum | Maximum | Mean | Std Deviation | Variance | Count |
| --- | --- | --- | --- | --- | --- | --- |
| How helpful was the app in assisting you with completing the test? | 1.00 | 5.00 | 2.50 | 1.17 | 1.38 | 32 |

**Table 430.**
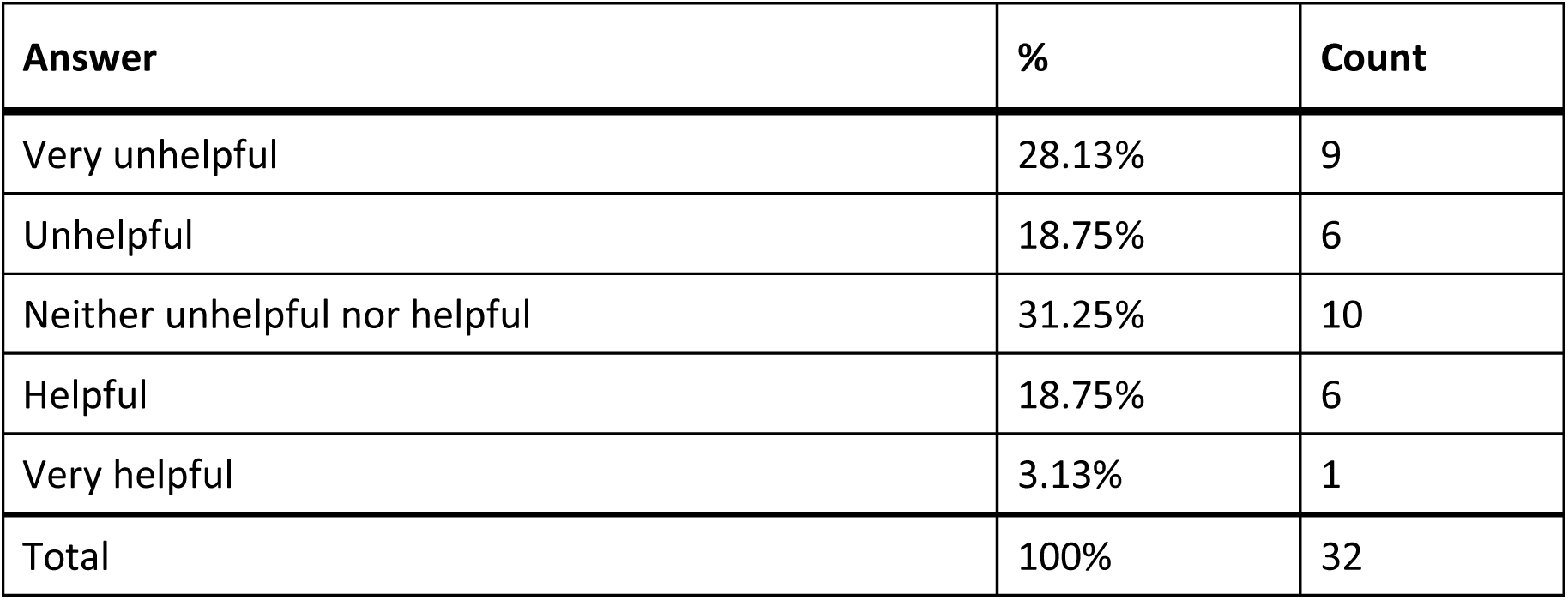
Responses to Q857: “How helpful was the app in assisting you with completing the [unknown] test?”

| Answer | % | Count |
| --- | --- | --- |
| Very unhelpful | 28.13% | 9 |
| Unhelpful | 18.75% | 6 |
| Neither unhelpful nor helpful | 31.25% | 10 |
| Helpful | 18.75% | 6 |
| Very helpful | 3.13% | 1 |
| Total | 100% | 32 |

### Q858 - Did you receive a valid test result with your COVID-19 home test (positive or negative)?

**Table 431.** Stats for Q858: “Did you receive a valid test result with your [unknown] COVID-19 home test (positive or negative)?”

| Field | Minimum | Maximum | Mean | Std Deviation | Variance | Count |
| --- | --- | --- | --- | --- | --- | --- |
| Did you receive a valid test result with your COVID-19 home test (positive or negative)? | 1.00 | 2.00 | 1.12 | 0.33 | 0.11 | 129 |

**Table 432.** Responses to Q858: “Did you receive a valid test result with your [unknown] COVID- 19 home test (positive or negative)?”

| Answer | % | Count |
| --- | --- | --- |
| Yes | 87.60% | 113 |
| No | 12.40% | 16 |
| Total | 100% | 129 |

### Q859 - Did you attempt to contact customer support?

**Table 433.** Stats for Q859: “Did you attempt to contact [unknown test] customer support?”

| Field | Minimum | Maximum | Mean | Std Deviation | Variance | Count |
| --- | --- | --- | --- | --- | --- | --- |
| Did you attempt to contact customer support? | 1.00 | 2.00 | 1.91 | 0.29 | 0.08 | 129 |

**Table 434.** Responses to Q859: “Did you attempt to contact [unknown test] customer support?”

| Answer | % | Count |
| --- | --- | --- |
| Yes | 9.30% | 12 |
| No | 90.70% | 117 |
| Total | 100% | 129 |

### Q860 - Were you able to talk with customer support?

**Table 435.**
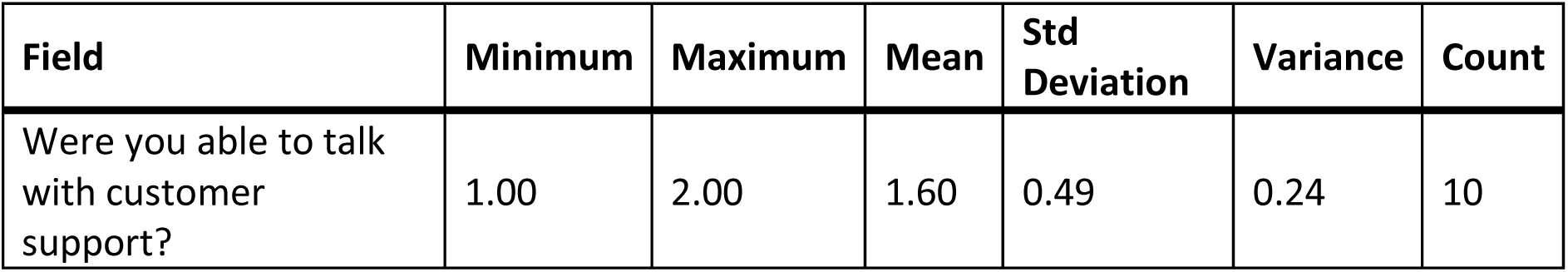
Stats for Q860: “Were you able to talk with [unknown test] customer support?”

| Field | Minimum | Maximum | Mean | Std Deviation | Variance | Count |
| --- | --- | --- | --- | --- | --- | --- |
| Were you able to talk with customer support? | 1.00 | 2.00 | 1.60 | 0.49 | 0.24 | 10 |

**Table 436.**
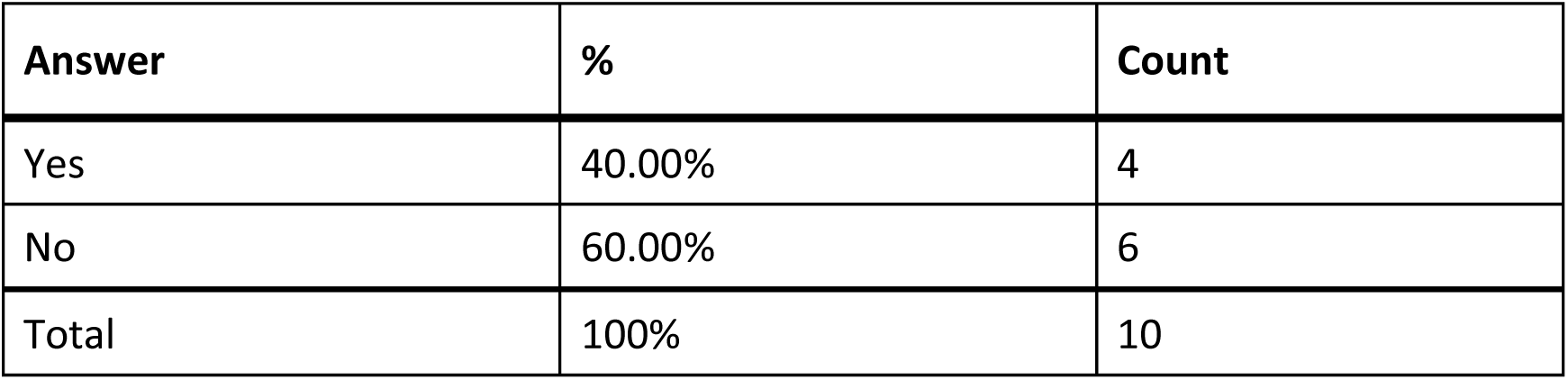
Responses to Q 866: “Were you able to talk with [unknown test] customer support?”

### Q861 - Was customer support helpful in answering your question(s)?

**Table 437.** Stats for Q861: “Was [unknown test] customer support helpful in answering your question(s)?”

| Field | Minimum | Maximum | Mean | Std Deviation | Variance | Count |
| --- | --- | --- | --- | --- | --- | --- |
| Was customer support helpful in answering your question(s)? | 1.00 | 6.00 | 3.75 | 1.79 | 3.19 | 4 |

**Table 438.** Responses to Q861: “Was [unknown test] customer support helpful in answering your question(s)?”

| Answer | % | Count |
| --- | --- | --- |
| Very unhelpful | 25.00% | 1 |
| Unhelpful | 50.00% | 2 |
| Neither helpful nor unhelpful | 0.00% | 0 |
| Helpful | 25.00% | 1 |
| Very helpful | 0.00% | 0 |
| Total | 100% | 4 |

### Q862 - If you have performed your COVID-19 home test multiple times, how did your experience change?

**Table 439.** Stats for Q862: “If you have performed you [unknown] COVID-19 home test multiple times, how did your experience change?”

| Field | Minimum | Maximum | Mean | Std Deviation | Variance | Count |
| --- | --- | --- | --- | --- | --- | --- |
| If you have performed your COVID-19 home test multiple times, how did your experience change? | 2.00 | 5.00 | 3.32 | 0.66 | 0.44 | 81 |

**Table 440.** Responses to Q862: “If you have performed you [unknown] COVID-19 home test multiple times, how did your experience change?”

| Answer | % | Count |
| --- | --- | --- |
| Much worse | 0.00% | 0 |
| Worse | 1.23% | 1 |
| Neither better nor worse | 75.31% | 61 |
| Better | 13.58% | 11 |
| Much better | 9.88% | 8 |
| Total | 100% | 81 |

### Q863 - What did you like about the test?

**Table 441.** Open response to Q863: “What did you like about the [unknown] test?”

| **What did you like about the test?** |
| --- |
| convenience |
| At home convenience |
| I took it at home |
| N/A |
| N/A |
| N/A |
| N/A |
| N/A |
| N/A |
| That it can be performed at home without having to go to a clinic. |
| Nothing. As a blind individual I was unable to take the test without assistance. |
| Honestly, there was really nothing. I liked about the test, except that it was available to take at home. Which actually didn't help me all that much because I needed lots of assistance to complete the task. |
| I like that I can do it from home. |
| Results in 15 minutes |
| Very easy to do and I was glad to do it in my home. |
| It seemed accurate and was easy for my husband to administer as he does not have a vision impairment. |
| Confort to do it at home, and wasn't difficult |
| Nothing |
| it said negative |
| It was available. |
| It was a mechanism to identify any present Covid19 |
| I liked that it gave me results about whether I have the virus or not. |
| not sure |
| My friend was able to clearly read and see the test results. |
| I could do it at home. |
| I liked being able to take the test at home. |
| I liked it it did not take as long as I thought it would. |
| Accuracy |
| It was available. |
| They were able to take it from the comfort of your home |
| It could be done at home, but in my case only with sighted assistance from my son |
| Finding out i had no covid. Seeing only the c line was fun. |
| Test components were easy to identify & set up. |
| I like that the test with the app is a little more accessible than the one without for me. |
| Compact |
| The idea that I could do it on my own was alluring but when it failed because I didn't correctly collect enough sample was disappointing. |
| Not much it was totally in accessible |
| Clear instructions, clear results. |
| The instructions for assembling & performing the test were in an audio & visual video format. |
| That I could do it at home. But I could not do it on my own, I had to have a sighted person help me |
| Free |
| That it was helpful |
| the app available for smart devices |
| nothing |
| It was at home. |
| Not really anything. I couldn't do it by myself! I am maybe if I was better at being blind I could do it but I've had seven years of experience I can see but I can't! Thank goodness my husband |
| was home. We did one test and it was positive and five minutes later we did another test and it was negative! I don't know thanks |
| I liked the attempt to be more accessible. The concept was great, but the execution could have been better. |
| that it did not take very long to get the results |
| Could do at home |
| nothing because it was impossible to make sure the small liquid contents made into the small hole to complete the test. after trying twice, i gave up. again, a small funnel piece would have made sure the liquid contents made into the small hole. very, very bad design. come on...surely you can do better. having to ask a sighted person to help defeats the purpose of having a home test for the visually impaired. i did not want to risk any of my friends or family getting sick to assist me completing the test. think about those of us who are blind and live alone.t |
| I could do it at home |
| i'm totally blind and i could do the test myself without asking someone else |
| not much. |
| i did not like the test |
| Nothing, it is way to hard, and I can't read the print instructions without help. I can't see the indicator lights as they flash. The packaging should be labled in braille. I would preferr a standalone test. I am not using the rest of my tests. |
| There was an app that had testing instruction video. |
| nothing other than a friend was able to provide phone support to my family member (sighted) and me as support number was closed for the day. |
| I liked the ability to know the result myself. |
| I liked that therw as an app availabl. |
| it could be done at home as needed (if I thought Id been exposed) |
| As a clarification, I have used multiple unknown brands of inaccessible Covid tests. However I based my responses to the 'other' category on the same experiences that I based my responses to this category on. |
| The only thing I liked about that test was that it was conveniently there at a time when I needed to take a test. |
| I appreciated being able to test when needed with the assistance of an Aira agent and without adding an app component. Aira allows 30 minutes per day for assistance with testing and other Covid related needs without using one's own minutes. That helped a lot. The hardest part for me was opening the vial; it would not be as hard if had to do the test again. |
| A sighted human did everything, but there is no way to indicate that in the radio buttons. Have taken several rapid tests when going to the dentist with no symptoms using same scenario. It was quicker, though more expensive, than going to my med provider and forcing |
| their system to give test results to the dentist. Missed a couple of appointments because results did not arrive fast enough. |
| I did enjoy having the test at home. I do not drive so I have issues with transporting myself to doctors offices when have symptoms. |
| Nothing as it was not fully accessible. The directions were in print. I had no idea that the boxes were tests since there was no accessible labeling. |
| Quick results |
| I didn't like the test because it wasn't accessible. |
| convenience of getting test results without going to a lab |
| Nothing |
| I didn't have to leave home to take it. |
| it was clean |
| fast results |

### Q864 - What would you like to see improved for your COVID Home test?

**Table 442.** Open responses to Q864: “What would you like to see improved for your [unknown] COVID Home test?”

| What would you like to see improved for your COVID Home test? |
| --- |
| Simpler instructions I have brain injury |
| undecided |
| N/A |
| N/A |
| N/A |
| N/A |
| N/A |
| N/A |
| N/A |
| The test was difficult to manipulate so I needed sighted assistance. It required putting the test fluid on a particular part of the test but not touching it. There was no way for me to read the results by myself. |
| The package could contain a disc or thumb (USB) drive with instructions in an accessible format. If USB was considered costly a return postage paid envelope could be sent to return the documents. When ordering the test a person could be asked if they saved the instructions for future use. |
| Instructions made available via braille and/or audio via an app or an actual braille. Component of the test, more easily manipulatable so parts that are larger and require less pouring of liquid and screwing on of lids in on Waze. Access to test results with a screen reader, without having to use site assistance. |
| Make the restrictions on eating and drinking beforehand clearer |
| 1. Larger font used for directions. 2. pre-filled vial/test tube. 3.perforated opening in package to hold tube upright while unwrapping swab. |
| Nothing |
| Nothing, I fortunately have help via my spouse. |
| Not sure if it was this one or the others, but directions were for 2 types of test kits. I had to first figure out which kit I had. |
| I would like to see a test that will actually give me the results without assistance |
| accessible information on package |
| clearer directions or a video |
| Everything. I would like to make sure that people with disabilities are involved in devising the test, testing the accessibility of the test, and that a wide range of people with disabilities are consulted regarding its accessibility. Having to receive help to take the test automatically means that it is not accessible. |
| Accessibility for those of us who do not own a smartphone or tablet. |
| I would like to have a way that I can do the test on my own. Since I am blind, I would like to have the test so that I can feel everything and when the results appear maybe there could be a sound. |
| I would probably need training how to use the appropriate app. |
| Easier use for totally blind |
| accessibility for those that don't have vision or the motor skills to complete the test independently |
| I would like to be able to take the test independently; possibly by the utilization of an app. Maybe, the test could have a gritty feel when positive, and no change when negative. |
| I would like it to be accessible for people who are totally blind. |
| Accessibility |
| Everything. |
| Audio descriptive by step by step |
| I think it would be much better to already have the solution in the vials, the swabbing part was easy but I totally am an unable to put the solution in the vials, and I am completely unable to see the results, if it said something like OK or Covid if it had some speech output that would help me. But I know that's impossible. And we tried using the app but that was even more complicated so that could not be used. But if the solution was already in the files at least I could swap it and put it in the vile I just couldn't complete the rest myself . |
| Oh wait for me to read it myself since I live by myself and have to know what is white and if it is or isn't and I can't read it in any form and he has no sounds to tell you if it's positive or negative |
| More accessible for blind |
| Large print instructions. |
| Need a non-visual way to interpret & validate results. |
| It's a bit difficult to pore liquid for the final test. Not sure how it can be improved |
| I really don't like that the test with the app requires you to put in your personal information, when other Covid tests don't require me to give up my privacy. |
| Larger lettering for the instructions. |
| Clearer instructions and an app that was really made for those of us who can't see rather than an app that was retrofitted. |
| I don't know how much it would take to improve it. The way that we could insert the Q-tip I guess to get the results. It just seems like the whole test was geared for a cited person. I don't know what to do to make it better for us but it's your needs to be. |
| Accessible tests that do not require a smart phone or other electronics to interpret the test results. |
| Clear language, accessible formatting, and corresponding app for viewing results with screen reader |
| It is very hard for someone with low vision to get the liquid into the tes tube, and to add the drops onto the test strip. It is almost impossible to get just four drops or to count the drops. |
| The ability to be able to rewind or fast forward through the instruction video.. And that each component of the test kit be described in broader detail, what each piece looks like, how each piece will be used or the function of each piece and how all the test kit pieces fit together in order to be able to start the test to get accurate results. |
| To be able to manipulate and see the results without using assistance |
| Somehow making it possible for a Visually Impaired person to test themselves without assistance. |
| Braille instructions |
| Audible way of stating whether it's positive or negative. |
| no videos included in the app that had no visual descriptions or captions. |
| tactile markings for the lines. scanning ease for bluetooth pairing |
| A better way to inform me there was a website or on-line app to help. I didn't and still don't know if my test had/has these options. |
| More accuracy |
| Your test wouldn't let me talk to it. I needed my husband and he's gone seven months here! He was home at this time. I absolutely could not do this test by myself Blind! |
| Continued accessibility |
| I woul like to hear more description of what the test components look and feel ike. That would have been helpful for me to identify things better. I am totally blind and some of the things went right over my head. |
| to provide accessible instructions for those who are not able to read the booklet provided |
| Instructions need to be in braille, results need to be spoken out loud and for older people they should not be required to use an app. These test should be fully accessible to the blind out of the box and not needing any additional assistive technology. |
| The instructions indicated that the line indicating a positive test might be very faint. A much clearer indication of a positive test is needed. |
| Easier to understand directions |
| include a small funnel device to make sure the liquid makes it in to the small hole without having to touch the hole and the opening of the liquid container to try to line them up to get the liquid in there. |
| larger print |
| more tactile marks on test stick so we know how far to stick it in our nose. Tactile or sound indication of the results. Audio or braille directions. |
| i don't want to take any more test |
| A stand alone test with speech. |
| The video that displays the results needs to be more descriptive when reviewing the test kit components. And more descriptive when setting up to start the final result process |
| make it less cumbersome, no mandatory video to watch each time and do away with voluminous instructions foldout in microprint guaranteed to create stress in a person with low vision and even for sighted assistant. |
| I was sent this during the national Federation for the blind conference. I had no idea who it was from as far as what app to get from the App Store. There may have been a QR code on the box but I could not feel any raised indicator or any information on knowing what to do. I had to call a visual interpreting service for help finding the instructions which sort of defeats the purpose in my opinion. The app had several and labeled buttons and even with my experience of using a smart phone was a tad bit clunky. The other thing I didn't like is the fact that I had to give them tons of information where as someone who is just taking a at home Covid test doesn't have to do. Just because I want an accessible way to know if I'm positive or negative I shouldn't have to disclose my birthday etc. to a company I don't even know. |
| Larger print. Plain language. Sustainable testing materials. Easier to grip test bits. |
| Braille instructions. I didn't *know* there was an app available until someone on Twitter told me. |
| The elements clearly identifiable and the process more easily able to perform without sight |
| clearer directions and more accessibility |
| I would like for there to have been an app for this particular test, or a way to complete it independently. |
| I couldn't find the barcode or QR code that I needed to scan also, I didn't know there was one until my sighted spouse told me. Then when I tried to scan it with my phone it didn't work. I was never really able to find the site that I needed to give me the instructions and ended up just having my spouse do it for me. I was super frustrated and gave up quickly. |
| Lack of reliance on an app. |
| I NEED a FULLY accessible, reliable test that I can administer myself. I don't want to have to give info to the testing company that a sighted person would not have to give in order to gain access. I can use an app on my smart phone, IF it FULLY complies with web accessibility guidelines. I DO NOT WANT my (lack of) ability to manage a camera while managing a test to force me to go somewhere and get a PCR test. I don't really want a human to come to my house due to test inaccessibility. If one has symptoms, one isn't supposed to use public |
| transportation, but I have no other kind, so had to risk getting others sick to determine my status. Can't wear masks, only shields, so, potentially more dangerous, if sick. |
| The paper instructions were incredibly small. I was not able to read any of what was written. I am fairly intelligent but was unable to complete my tests at home. |
| Braille and large print label on the box itself, braille, talking and large print instructions. |
| Should not be required to provide personal information in order to use the app. |
| Make one that speaks instructions and results. One that can be used repeatedly with different cartridges |
| Larger print |
| Markings placed on the items in the test kit to identify which parts of the test go together. I also didn't know how to insert the swab into the test tube to verify if I was positive. |
| I'd get a home test that is accessible to blind and low-vision people. |
| ability to perform test and interpret results independently |
| A test that is not so intollerant of components being touch as it is impossible to guide the swob into the test tube without touching it.A dispensing container for the test agent that is pre-measured as the number of drops is impossible to know (including whether or not you got a drop into the test tube).A telephone number with an experienced professional who could help you do the test (featuring the above improvements) over facetime or zoom. |
| audible instructions and results without having to use an app. |
| Better instructions |
| Better instructions that clearly and thoroughly take you through the testing process from beginning to end. These instructions should be in Braille or in an easily-accessible audio format. |
| Needs to be accessible without other human intervention |
| I need instructions and results I can read nonvisually. |
| All home Covid tests should have accessible smart device apps to accompany them which read instructions and results clearly as well as send to healthcare providers and patient portals. Tests should also have raised, tactile dots, bumps, lines, or other indicators identifying positive and negative results for those who do not have access to smart technology, cannot afford, or have skills to use these devices. Tactile identifiers are an additional confirmation for those who do use applications with smart devices. This would be helpful for those living with visual impairment and multiple disabilities such as hearing impairment, mobility, dexterity, cognitive, intellectual, processing, neurological, and sensory disorders. |
| Have accessibility built in from the ground up. For example, it would have been very easy to write the instructions using large print. That would not only help people who use large print, but probably also senior citizens who do not identify as being blind but who have difficulty seeing. |
| accessible instructions in Braille recommendations on which home tests are accessible with an app or by other means |
| nothing |

### Q206 - How would you feel if you were provided with only digital instructions (no printed instructions)?

**Table 443.** Stats for Q206: “How would you feel if you were provided with only digital instructions (no printed instructions)?”

| Field | Minimum | Maximum | Mean | Std Deviation | Variance | Count |
| --- | --- | --- | --- | --- | --- | --- |
| How would you feel if you were provided with only digital instructions (no printed instructions)? | 1.00 | 5.00 | 3.26 | 1.32 | 1.75 | 500 |

**Table 444.** Responses to Q206: “How would you feel if you were provided with only digital instructions (no printed instructions)?”

| Answer | % | Count |
| --- | --- | --- |
| Extremely negatively | 12.80% | 64 |
| Somewhat negatively | 16.60% | 83 |
| Neither positively nor negatively | 24.80% | 124 |
| Somewhat positively | 23.00% | 115 |
| Extremely positively | 22.80% | 114 |
| Total | 100% | 500 |

### Q191 - This is the last survey question. Do you have any feedback regarding COVID-19 home tests in general?

**Table 445.** Open responses to Q191: “Do you have any feedback regarding COVID-19 home tests in general?”

|  |
| --- |
| **This is the last survey question. Do you have any feedback regarding COVID-19 home tests in general?** |
| They need to be easy to obtain and inexpensive to be effective for general population |
| Love it. It's easy. |
| No |
| It wasn't terrible just a bit difficult |
| Simple and easy to understand. Stop sending reagent vials that you have to put together to actually use the test. |
| The expiration date should be easier to find, and more visible w/larger font |
| Make them easier to use for all. |
| As a person with limited fine motor skills, it is extremely difficult for me to hold the small vials, screw on a small lid, and precisely squeeze four drops of the liquid into a very small area. I may accidentally put more or less drops into the sample well or I may miss the sample well entirely. |
| No. |
| They should still be provided for free as it may be cost prohibitive for some people to purchase the tests, thus they will not test and will remain in the community while contagious. |
| no |
| Question accuracy of some tests |
| no |
| They are convenient and private if they come out negative you don't have to take any other steps |
| na |
| The less steps and less possible contamination, the better. The Ellume test seemed to provide more false-positives than the Bianax test. |
| :D Hi Amanda! |
| I would like them to be easier to use for someone with little motor control or coordination |
| Happy they are available |
| No |
| None |
| Easy and available |
| Glad they are available. |
| No |
| No |
| The only test I have used is the is IHEALTH and I have a positive opinion. |
| N/A |
| N/A |
| N/A |
| N/A |
| N/A |
| N/A |
| N/A |
| N/A |
| N/A |
| N/A |
| N/A |
| N/A |
| N/A |
| N/A |
| N/A |
| N/A |
| N/A |
| N/A |
| N/A |
| N/A |
| Who lives in a pineapple under the sea? |
| N/A |
| N/A |
| It would be helpful if there were fewer steps and if one did not have to measure out a certain numbe of drops into the test analyzer. The instructions need to be fully accessible. This applies to any digital ones as well. |
| Test |
| hi |
| Because of the length of time and information required by the Ellume app, I might just use inaccessible tests again in the future. |
| Please understand that just because instructions are provided in a digital format does not make them accessible to people who use screen reading technology for accessing digital content. |
| I have no feedback. |
| Yes. Why is there only one that's accessible to me, and I had to order it from the government to have access to it? What do people without smartphones do? |
| I want a paper set of instructions because it's easier for me to read and reference back to |
| Positive accessibility service for individuals who need COVID-19 home test kits. |
| I wish there were more accessible options, but I am greatly appreciative for the accessible option. |
| No |
| I was advised that there are accessible tests. But also requested that only a person who could not get assistance should order them. So a higher quantity should have been available. |
| The test results need to be made more reliable and accurate. It would also be a good idea to make it possible to activate a checkbox to have the information automatically transmitted to your state reporting service or the CDC if your test is positive. |
| Thanks for opportunity to do it at home, just need improvement in instructions- made it less complicated, use simple words, need bigger size of letters and make parts in the kit colored, otherwise it is almost imposible to do test if you have low aor very low vision. |
| Glad to have home accesss |
| No |
| make the material larger and, even though I had not trouble with the directions, they could be clearer and larger |
| No |
| So much is unclear with at home tests - are they PCR? Are they the same as the rapid tests at the Health Department? Once I test positive, to whom do I turn? |
| A nice tool to have |
| more free tests, more often |
| None |
| I'm very thankful for these at home tests! I wish there was an at home test for the flu and Strep! It would save a lot of infected people from infecting others! |
| I wish they were more accessible for those of us who have severe vision impairments but who don't read Braille. |
| No |
| I don't have enough experience but I heard the Walgreens pharmacy kit are more difficult |
| I would prefer American made COVID-19 home tests. I will not buy the ones made outside of USA. |
| Using the dropper and transferring the liquid holes could be made larter. |
| Some of the packages have a number of tests in them and it's very hard to tell which one is which |
| NO |
| They are not independently accessible |
| a video showing how to do would be great but it needs to have cart and asl |
| Although test have final result reading (+/-) the instructions should match all test kits regardless of the vendor licensing. |
| I hate that you cannot get the accessible ones any longer. |
| I also use a commercial quick test for my work. Three simple steps..if I mess up I have to test again. The home test were just too complecated. |
| Wondering if it was tested on people who are blind... Doesn't seem so or they will know if these problems exist |
| None at this time, although having a QR code to download all the necessary things would be very useful! |
| no |
| They are very handy. I wonder about if virus #'s are useful since many may not have reported positive results. |
| No |
| I thiNK Covid was just a buNch of B.S.! |
| no |
| They are a good thing. |
| There's been a lot of trash/waste generated from these - it would be great to consider enhanced sustainability measures with future tests |
| No |
| none |
| I dont think they are very accurate |
| Uncertainty about their reliability |
| no |
| pricing is varied, the cost should be inexpensive so people will test more regularly |
| The paper option needs to remain to ensure equitable access to testing. Most older adults struggle with the technology/app approach. I had to go and help not only my older adult parent due to dexterity but also her friends who struggled as well with dexterity issues and then also trying to follow the videos online. They were developed for tech able individuals and not for older adult that were at highest risk, infection and critical for monitring. The challenges of using certain tests almost led to older adults not testing themselves in my parent's friend group- many of which had high risk conditions. I instead went and purchased binax and other simpler tests so they would be able to use them. |
| I just tested myself this morning, and after having done this several times, I still needed to ask my husband a few questions just to be sure. It would be nice to have had a simple illustration showing steps one by one (in larger print!) |
| Easier opening needed for most all packaging |
| unreliable |
| As I said, before, I hope that one can find a way to do the test that blind people can feel everything, and when the results appear, there could perhaps be one sound for negative and another sound for positive. |
| It would be great if any home test (covid, pregnancy, etc) was more accessible (getting results in an app). |
| Even with good vision, the tests can be problematic (expired tests sent, tedious manipulations, small print instructions) |
| I don't feel competent to do the test myself. I also feel uncomfortable asking friends to help when I could have covid. Braille instructions, at least to get started could help. I could try to use IRA or Be My Eyes, but I don't know how helpful that would be. |
| Thank you that they are free. |
| I don't have any. My sister says I would need to read directions but I have low vision |
| No |
| no |
| Not really. |
| No feedback at present. |
| They need to make a test that is truly accessible for people who can't see: place raised circle or square around hole for drops; make container for liquid able to drop only one drop at a time so that squeezing it six times will be an accurate indicator that I have put six drops in the hole; making results accessible. |
| They need to be made so blind people can independently take them with little to no interaction with sighted people. Many blind people successfully live alone so don't have a sighted person to read the instructions or help with assembling the test kit in order to put it in the tube or assemble the portion that gets shoved up your nose. |
| No. |
| Even with instructions on administering the test independently, I would still need to depend on a sighted person to read the results. |
| No |
| They need to have more accessibility features for blind users |
| These tests must be made accessible to all people with disabilities. This is a public health matter. It does not matter what the disability is, all at-home tests must be made in such a way that they can be universally accessed by anyone. |
| No, I do not have any questions about home COVID-19 tests. |
| THE instructions make it sound far too complicated. I have had two tests but I went to a testing location to have it done. I have 12 tests that I will not use. |
| You're wasting blind folks' time. The government does not provide these tests any more and I can't afford to buy them. |
| No |
| I am totally blind and I teach technology, these tests no matter what you do to them are not accessible to me without site assistance. Like I said before, if the solution was in the vials I could certainly suave my nose and put the cotton suave in the vile Not a problem but I could not complete the rest myself I could not see the bar graph turn colors I can't even see the bar graph. We tried using the phone app and that was totally useless it was more confusing than helpful. Even if I called AIRA I wouldn't be able to put the one or two drops in the vial without making a mess. |
| Yes, with the state of technology today it is somewhat disappointing that more tests are not accessible for blind and visually impaired users. It is virtually impossible for a blind person to not have to share their test results with someone else. |
| Accessibility features is the only thing that I have to add |
| By creating COVID-19 home tests that are accessible to blind people, it allows us to be independent in administering the test to ourself or family without having to put someone else's health at risk to come and assist with the administration and interpretation of the test results. |
| 1. The app should be screen reader accessible without jumping or getting stuck . The user should be able to left and right swipe to every control on the screen in reading order. 2. Don't ask a blind person to drip a specific number of drops into a container. Instead design the equipment to puncture the liquid as the two sides snap into place. 3. describe the parts of the kit so I can figure out what I am touching and how many pieces to search for in the box. |
| A blind person is unable to do this themselves everything is print based so there's no way for a blind person to tell what the test results are or what they say |
| Why did we have to wait for the Ellume? This should have been the first to be deployed, not the last. |
| No |
| The COVID-19 home tests are NOT ACCESSIBLE!! I cannot read the tiny directions, nor do I have the necessary app. So the tests are packed in a box and totally forgotten. When I needed a test we automatically went to a hospital to get it. The home tests in my garage are a complete waste of tax dollars! You said they would be accessible - what a joke / lie! |
| Easy to use. |
| They are easy because i have dome vision still and i could rnlarge the instrucyikns eith my phone camera. |
| Being legally blind, and unable to use them, I gave my home tests to my brother and sister. |
| None of them is accessible or reliable or even necessary with the endemic state of this virus now. |
| If a test is not accessible, cited assistance, always helps. to me that's another form of accessibility if I can't use apps or access technology. |
| Old at home Cove test should be fully accessible to the blind and visually impaired and be accessible so that we can take the test without any assistance from anyone in the privacy of our own homes |
| Come up with a more accessible option that works for people who are visually impaired but don't have smart phones. Add something tactile, braille or something else, to the boxes so people know what they are. Don't require blind people to give up their privacy to take a test. |
| Not really. |
| Tried to use but instructions almost impossible to follow for |
| I was very happy to hear that there were blind accessible tests. I requested and received a whole box of them. I was 100% not able to use them as gaining access to the digital instructions proved to be a huge barrier. There was some kind of form into which I had to enter information in order to gain access to the instructions. I was unable to do this as a blind person. My fully cited but dyslexic friend was also unable to help me. There was no one else around to help us. This friend is no longer here. So I take the bus and go to test sites and get PCR Covid test there. I wish I had accessible home testing but I don't. |
| Very helpful and needed. |
| I opted in gong to Urgent Care to be tested because I could not read the instructions on the box. |
| Because there are so many parts to this test, then needs to be a better identification of the individual components. And how they all relate to each other. The film that was shown in the app is not descriptive enough. This is especially the case for the vial of Tesl liquid. I wasn't quite sure what it was for and what it should look like. I had to kind to. I had to almost do a dry run of the components to see how it all worked. |
| it would be helpful if the instructions were ditgal |
| I also used a previous test (can't remember the name) with an app. This test was more accessible only because my phone is magnified for my use. But the app was timed & I continually ran out of time performing each step. Reading larger print takes more time because less text will fit on the same screen. So following directions may take longer & the timing will be off. |
| Haven't found one that worked. |
| I hope the home COVID tests will continue to improve and become more accessible. I worry about those who do not have smart phones or other devices to interpret the results. I also hope that more types of tests, like flu or monkey pocks, will become available and be accessible as home test kits. |
| I think instead of self administering the drops, there should be some type of internal capsule that can be punctured by applying pressure, that will release the proper amount of drops |
| directly into the strip. It would leave little human error in measuring for those who have difficulty remembering how many drops entered or visually impaired who may have difficulty seeing where to apply the drops, or even those who suffer mobile control such as shaking that would cause overage in the amount of drops onto the strip. |
| Not at the moment. |
| I wish more apps were more accessible. |
| None |
| Digital |
| It would be great if all blind people could access Cue Health's NAAT system. It is highly accessible, easy to use, and more accurate than rapid antigen tests. |
| Clearly, no one bothered to ask us what we think until now so I have no confidence that anything I say here will be taken seriously. |
| Is there a way to avoid needing to put drops of solution on a teeny tiny contact point? If so, that would help tremendously. |
| I wish they were still free through the government. I am happy I can get reimbursed, but it was easier to just order the tests and have them shipped for free. |
| Having access to health/medical testing such as this is critical for me to be able to monitor my own health and the health of my children. |
| I think this is a fine program. |
| They should all be free and it is a difficult process to get reimbursed. |
| I'd like to be able to determine the expiration date on test packages. It seems to be difficult to figure out, even with assistance. |
| Yes |
| I would like to know how to find an accessible home test. |
| digital instructions would be grate but that doesn't help with the other test items. |
| They should not require the transfer of liquid or counting drops or anything else that requires precision. |
| The test was even difficult for the cited person assisting me to use. The print could be larger and easier to read. |
| No |
| I think the point of transferring the vile to the sample well is the common denominator that is hard for all tests I have seen and presents the most challenge for someone who is blind. |
| HAVE THE TESTS BUT HAVE NOT USED |
| Tactile indicators on package contents wouldl be helpful. I am totally blind so references to flashing light are meaningless; however, if a time lapse is in seconds or minutes, that's helpful. I thought the Elume video was very clear but had to discard the test because I had touched all components. Test kit expiration dates would also be good to know. |
| No |
| I think at home COVID-19 test have gotten much better, I totally prefer a digital way of accessing the instructions on how to do the at home COVID-19 test. I believe that the testing components could have a better way to be identified, and the results should be super quick with a text message |
| I & my family don't use hometests in general as it is not needed. We live our lives with or without Covid. We don't live in fear. When we got sick, we recuperate fast because we eat healthy food, exercise. We don't wear mask. We breath freely . Going to the beach & doing activities outdoors should be encouraged to the blind community. Thank you |
| Nope |
| I do not take one because I cannot see the results |
| Test needs to be as non-visual non-hearing simple as it can be made. Instructions were confusing and the one I used had a video that had no sound or captions. Tests need to be made for even those who are deafblind who have no hearing and no vision. |
| I haven't bothered picking up a home test as I know they are not accessible. When I got sick with what might have been COVID, I just isolated for a week. |
| If I had to perform the test independently, I feel confident that it would be nearly impossible to achieve. |
| It is nice that we have them. But, where is the best place to store them? |
| pages and pages of instruction are daunting. Most tests for use at home are pretty consistent in process. Keep it simple! |
| Nfb funding Aira calls for covid tests has been a life saver. I couldn't read any brand of test without Aira or a human assistant. |
| Make it easier to drop the liquid into the test strip. |
| please consider making tests available from the beginning of an outbreak and not monthes later as an afterthought when people complain. |
| N/a |
| No |
| is it possible to make a general profile since you created the app to stay on the phone. You can swab others by clicking other on the app but the primary covid account should be set up so you do not have to listen to the instructions every time you start a test |
| Or someone like me, cited assistance is necessary. Any home test should offer, virtual, cited assistance, and a process that is possible for a totally blind person to carry out. |
| Thanks fir trying |
| They have improved, but have a long way to go. |
| The app on a smart phone needs to provide feedback as the test is performed, such as "sample applied" so the user knows it is being done correctly. |
| I would not mind trying other manufacturers to see how they stack up against Ellume. |
| These tests need to be fully accessible to the blind out of the box and until they are we should not release any new tests on the market. I am happy to help test for Accessibility in future tests if you wish to contact me. |
| They are difficult to use. The government does not know how to help, only directing me to contact the postal service, which also does not know. The program, I think, is discontinued, but people could not request more-accessible tests online; they, including me, needed to call to request them by phone, and even then there were difficulties. |
| no |
| The digital ones seemed easier & being able to have the results sent saved time and questions |
| I would like to see the test get better for the blind. |
| Please contact me for assistance in developing a tin a test for visually impaired individuals |
| I am just happy that there are test out there that I as a blind person can perform independently |
| They all suck, none of them is very accessible and they are basically not even necessary anymore so a waste of money. |
| The designer of this survey does not have a clue how blind people use smart phone apps. They did not ask even half the important questions about this poorly designed test. |
| The test I tried to take were hard to figure out. It was unclear what part of the test search which purpose. |
| I have always taken tests without symptoms. If I were actually sick, I would not want the test to be as complicated as it is. |
| I didn't understand the question about printed instructions. The test comes with those in the box. If I could access digital instructions without downloading them myself, that would be great. About the packaging - I don't have any trouble with the tape, but I can see that it might be an issue for someone who doesn't have good manual dexterity. I also think that having the expiration date in a more obvious place would make it easier when using Aira or BME to access the expiration date. |
| I wasted 2 test kits because of accessibility issues. |
| My friends had to struggle with inaccessible tests and get help on a video call. We ordered the accessible ones but they did not arrive in time. Now I'm concerned there will be no options when these expire. |
| These tests are helpful for quick results. |
| Accessible instructions would allow easier use. Don't even know how to get started and what is needed. |
| It would be helpful if there was a high level description of what's involved in completing the test. |
| All home medical tests need to be designed to be performed by blind people independently. |
| No |
| It would be helpful if one could take the test without the use of a smart phone. I am going to see if my color detector will be able to pick up the color of the line on the test strip. |
| Don't assume everyone has access to an app or smartphone when designing the test |
| please refer to my previous comments. very bad design for the blind. |
| I am grateful for home Covid tests. I think they are very important, and I can understand why it would be necessary to obtain a pair of eyes to look at the results. I have one comment on your survey: you asked if I was able to get a positive or a negative result. I do not know if that result was accurate, but I was able to get a result. Since these tests are not perfectly accurate, I cannot tell if it was a false negative. |
| They have along way to go to fully incorporate nonvisual information both in terms of documentation describing the parts, and then like the Ellume being able to fully independently interpret the results. The testing did get easier because I have done about 20 of these tests. |
| I will need to take a home COVID test next week in order to attend a group convention. So far, I have been unable to even enter my patient information into the Ellume COVID Home Test. Good accessibility is absolutely critical for any test intended or marketed for blind and low vision customers. |
| I would want the test kits to be accessible with LARGE print instructions with easy-to-read diagrams on how to perform the tests |
| More at home tests like pregnancy tests need to be accessible and there needs to be a solution for people who do not have smart phones |
| All tests should be designed for accessibility from the beginning so that accessibility will not only be easier to achieve, but cheaper, as well. |
| Digital directions depend on if they are accessible with my screen reading software. Or if they are in non-accessible pdf format. |
| Ellum should be used as the blueprint for all other at home Covid test companies to follow along. |
| no |
| I repeat, I would like to see a standalone test that one doesn't need to use technology for. |
| There needs to be more full accessible ones. the one that I had, was not known to be the most accurate. |
| it doesn't have to be as complicated and off-putting as the so-called "accessible" test currently is. The MO seems pretty much the same for these tests: swab nose, put swab in fluid for x time then drop proscribed amt of fluid into test cartridge. Easy Peasy. Who needs 19 pages of instructions? And the darned video! aaaargh! We all seem to want it fast, easy and accessible. Maybe there could be a design with less measuring of the fluid into the test cartridge, or perhaps the fluid would stop measuring at a certain point. But for the sake of allthings holy, stop with the mandatory video!!! |
| I think I spoke my mind on the last question or we could write in our answer. I would just like to see more thought of the totally blind experience. As I open the package for my at home Covid test I had no idea who the company was I still really dumb what app to get from the |
| App Store etc. When I ordered it through the government there was no instructions on that page they just wanted my address and name. Maybe when shipping it out they could've emailed me information on what to do once receiving the kit. |
| The test also shouldn't require you to have to download an app cause remember we have to find the app and then put your information in put your information in and that's how you get your test results salt it was not an easy process to do the test and why it's great that they send you 12 tests |
| No |
| I believe there is more room for error with at-home tests. |
| I do not trust results of home tests at this time unless I am symptomatic |
| I wish they were more freely available, so that testing could become mandatory in public settings (e.g., church, public transportation, school events, workplaces). |
| pass out more free ones |
| I like the idea of home testing, but it must be made easier to take the sample and put it into the maching without invalidating the test. |
| While I appreciate the effort in creating the "more accessible" at home COVID test, from what I've read and heard, it sounds very complicated to use, requires visual assistance which defeats the purpose and requires the user to enter personal information that someone who is able to use a standard nonaccessible at home test doesn't have to provide. Why can a test with tactile information not be created so that it would be similar to the standard at home test? |
| Have instructions in advance of testing so i know what i need to do. |
| Customer support for the accessibility of COVID home tests needs more effective training. App developers need to work with and hire qualified blind app developers. |
| They need to be able to be completed 100% independently by individuals with disabilities. |
| More consistency of how the tests are done would be great, and no pouring drops. |
| glad they were available through the government |
| Instructions on how to conduct the test should be available also in braille for those who use braille. |
| Positive feedback. Outcome For. Everyone. Involved |
| The steps necessary to administer one of these supposedly accessible home tests are not possible for a blind person to carry out. For one thing, we are advised not to touch certain pieces of the test equipment. But a blind person cannot find out which piece of equipment to use for which step, let alone use it for the purpose for which it is intended, without touching it. A blind person can't measure out a certain number of drops of something to be poured into a very small receptacle. A blind person cannot see a light, flashing or otherwise, when asked to ascertain something by a light. Not every blind person has a scanner to read printed instructions. When the test arrives in the mail, there are no audio instructions in the package |
| to even let you know which app you need to download, and no audio description of what piece of equipment is what, what it is called, or what its purpose might be. Yes, you can successfully open the box, but after that you have no clue what to do next. This whole home test process has to be totally re-evaluated! Should I need to do another home test, I will get a regular one used by sighted people and have a person with sight help me to administer it, as that is what I had to do with the home test that was falsely advertised as accessible, but definitely was not! |
| Yes. I heard that the package the test is in is difficult to open when you're visually impaired. The instructions must be screenreader friendly to iPhone users. |
| Like the elume better than the iHealth. |
| No |
| I am glad the home tests are available as getting a test through urgent care or other medical provider would have taken much longer and would have delayed contact with my doctor and getting appropriate medication. |
| Cue health is the most accessible test that I've used. Even though it's expensive, it's worth for the investment. |
| Why were the accessible tests released so much later than the others??? |
| We need PCR home test kits that will allow throat swabs, and some kind of means by which to report results. I have only one fully functional hand, so need something I can successfully manipulate. A bluetooth dongle will not work for folks who aren't adept with their devices. If a dongle MUST be present, it must beep and vibrate to give signals as to connection status, start of test, and completion of test. My tech skills are adequate, but I worry there are not enough places for blind folks to get tested if they cannot borrow a sighted human safely, nor ways to get to such places, if all they have is public transportation options. Uber and Lyft drivers often have air fresheners in their vehicles from which I can get migraines and Asthma symptoms, and local Paratransit will not allow shields only, unless with a medical exemption. I have not yet been successful in procuring one. |
| It is a good idea to have these at home. If a person is sick they should have the ability to test themselves without endangering the public at large. Providing a larger interface for those who are low vision would be very helpful in the future. |
| It would be nice to have one that truly is fully accessible. To date, I can't take a covid test without sighted assistance. The test I used I had to be able to see lights to tell when the lights stopped blinking. I live by my self and I am totally blind. |
| I think the test should be simplified for everybody. I don't think they need to be as complicated as they are. I know they don't need to be as poorly labeled as they are. And who can read the small print on the instructions? My visual helpers had difficulty. I deserve the same access to information as sighted people have, and I deserve to have it with the same amount of effort that sighted people have to make. I should not have to do 10 things when they only have to do one. And if there had not been free available visual interpreting service, I could not have done the test at home. Not being able to do the test at home would've put others at risk. |
| The ones sent out that were supposed to be accessible were not very accessible. A totally person or someone with extremely low vision would not be able to use them |
| It would be great if one could just swab the nose, insert the swab into a vial of fluid and then hear test results. |
| We were forgotten as usual, I hope the feds learn from this and require these companies to conduct responsible test design especially since our tax money was spent developing them just like everyone else. Very poor performance! |
| No e at this time |
| I have no feedback |
| I would like audio and large print instructions on how to properly use COVID-19 Test Kit. |
| No |
| They are helpful. I use VoiceOver to read some of the text. The Video was good but we were unable to play it over again before starting. My wife was looking at the papers inside and I missed some of the final steps in the video so it was failure as we did not know know to do next ie, open the cover , turn it over and put the drops in the sensor. We had paired the unit via Bluetooth on my phone but that was all... |
| A vast majority of covid tests require visual detection of test line(s) which require a blind person to seek sighted assistance, and compromise health and privacy. Additionally, many such tests requiring pouring and mixing chemical solutions in a very small space which is difficult to do nonvisually without risk of invalidating the test due to contamination of test components. |
| I do not feel the current testing is accessible for someone with sight loss. |
| I had to read the instructions several times to be sure I was doing it correctly. I could not load the app referenced. |
| No |
| I'm extremely grateful for home testing, esepcially because I don't drive and if I think I have Covid, I don't want to ask someone for a ride or get a cab. |
| Maybe an individual can have an option of getting the Covid test via email or paper. Large print braille and a audio described YouTube channel |
| There need to be other ways of getting the instructions, such as Braille for those who do not have internet, nor do they use email. There also needs to be an acessible means of accessing these tests for those who do not own a smartphone. |
| All home tests need to be accessible to users who are blind. Using a smartphone app to read the test results and provide instructions is a decent work-around, but in the long-term, test results should be fully accessible out of the box. |
| N/A |
| No |
| Believe the tests using swabs in a tube instead of a dropper are much better. |
| They do need to be more accessible for people who are blind or have low vision. I would like to see separate directions for people who are blind or have low vision in large print or braille to follow when taking the test. The app is helpful, but it makes taking the test much more complicated when trying to read the directions and then conduct the test. If accessible written instructions were available, it would be easier to refer to them as proceeding through the steps of the test. |
| Was unable to use it. Needed help from sighted person |
| no |
| I wish these tests were easy & accessible to use myself so I do not have to rely on another & worry about infecting that person. |
| COVID-19 and all diagnostic tests need to be made independently accessible, not relying on smart phone apps. |
| I was not able to use this test at all. |
| no |
| I'm thankful some efforts are being made to produce accessible Covid tests, despite the difficulties we have encountered so far. I hope further improvements will be made to make the experience even more positive. |
| any accessible instructions are of very questionable value given the difficulties in completing the mechanics of the test |
| There needs to be more accessible tests available and available locally. I can't get them through USPS now and I can't get them at my local pharmacy. I did find them online but there are two versions of the Ellume test one not being accessible and the one that is doesn't say it's accessible you have to interpret the product name. |
| They are too different from each other and you can't pick and choose the one you get so you start from scratch each time you get a new brand. Counting drops is impossible. I could have used AIRA to read the results, but I had on site sighted assistance. A Braille strip inside each box giving the web address would be helpful. |
| No |
| the test that claims to be accessible is not. First, some of the instructions are not clear and it requires an app to perform |
| Glad that there is a way to take a home test and get results but very difficult for blind people without a sighted person to assist and then readers are concerned that they might be infected with the virus if my test results should indicate that I am positive. |
| I think they should be available to those who need them, however, I did not take a home test nor do I have any plans on taking one in the future. |
| None |
| I have received the Ellume accessible COVID-19 home tests. I have not actually taken a test. But I find the app not very easy to use. There are unlabeled buttons, and I have not yet been able to complete entering my personal information. I cannot get the picker items such as my birth date successfully saved in the app. |
| Wish they had been available sooner |
| The home test worked out well after I learned what the components are. If I had had a way of better understanding what the different components look like and what they were used for it would've worked out better there needs to be better description of the components especially for the first time user. Perhaps you need a starter kit that allowed you to touch and play with their computers so you could learn how they fit it together |
| I am grateful they are available but wish they were more shelf stable. The use period is very short (roughly six months). |
| I don't think they are reliable enough to give a certain result. Now that COVID is in it's 2nd year and we know it'll be with us forever I don't think testing at home is necessary. A positive result has the tendency to worry people when all they need to do is treat the symptoms no matter what the virus. |
| It was misleading to call the ELLUME test Accessible. The only thing that was truly very easy was being able to read the result in my email and having a copy of that result. Now that I've done the test twice I can do it but as a nurse I should've been able to do it right out of the package very quickly and easily because I've done that type of complicated testing before. I showed my daughter who is completely cited all of the components and she couldn't believe how many pieces I had to figure out before I could do the test. and let me state again why does the company need so much information about me before I can start the test? they not only wanted my name they wanted my address my phone number my email and my age. If this was a pregnancy test I would be highly suspicious that the information was going to be shared with some state or federal government agency. And there are indications that this company is going to start releasing test for pregnancy that they consider Accessible. |
| I never got an opportunity to try the accessible version. I would be hesitant to do any of them alone. The results are pretty important, and I would want o as sure as possible for getting it right. |
| I would like some verbal description of how long they are good for and how I should store them. I'm delighted that we, as totally blind people, are able to use them--that the government took our needs seriously. |
| I can't tell when the expiration date is for any of the tests even the more accessible one. |
| My personal concerns are about visual access, but it should be noted that you also need excellent hand coordination, the ability to follow rather complex instructions, and after all that there's no real assurance that it's accurate. |
| If a test has accessible digital instructions, a customer service line that can assist, or other access features, there needs to be some way to indicate that on the box nonvisually so I would know about them. For example, a print box with a printed customer service number, or a printed qr code, or printed website or app name, would be about as unhelpful to me as not having any access features at all. In other words, accessible tests need to also have ways for the access features to be discoverable in an accessible way. |
| All tests no matter the brand should be accessible. |
| It would be great if they were all accessible. |
| Please see question on previous page pertaining to improvement of particular test brand. |
| no |
| n/a |
| Binex was easier to use. Like a pregnancy test. |
| important to have them free of charge for everyone to use |
| There needs to be a way for physically disabled individuals to receive human assistance if self-testing is not possible. |
| No |
| N/a |
| I just wish they were more accessible financially speaking: maybe a discount offered for those who purchase in bulk due to large families, or repeated use needed due to health conditions. |
| THE TESTS ARE TOO HARD TO USE, I FIND ANYONE AT HOME MEDICAL TEST HARD, I OFTEN NEED CONSTANT STEP BY STEP SUPPORT TO ENSURE ITS DONE CORRECTLY, MEDICAL PLACES SHOULD NOT CHARGE HIGH FEES TO GET COVID TEST FOR THOSE OF US UNSABLE TO DO AT HOME TEST |
| The complicated instructions are even more difficult when you HAVE covid and your brain function is impaired. |

